# Genomic epidemiology offers high resolution estimates of serial intervals for COVID-19

**DOI:** 10.1101/2022.02.23.22271355

**Authors:** Jessica E. Stockdale, Kurnia Susvitasari, Paul Tupper, Benjamin Sobkowiak, Nicola Mulberry, Anders Gonçalves da Silva, Anne E. Watt, Norelle Sherry, Corinna Minko, Benjamin P. Howden, Courtney R. Lane, Caroline Colijn

**Affiliations:** Department of Mathematics, Simon Fraser University, Canada; Microbiological Diagnostic Unit Public Health Laboratory, Department of Microbiology & Immunology, University of Melbourne at the Peter Doherty Institute for Infection & Immunity, Melbourne, Victoria, Australia; Victorian Department of Health, Melbourne, Victoria, Australia

**Keywords:** COVID-19, Serial interval, Genomic epidemiology, Whole genome sequencing, Statistical model, Reproduction number

## Abstract

Estimating key aspects of transmission is crucial in infectious disease control. Serial intervals – the time between symptom onset in an infector and infectee – are fundamental, and help to define rates of transmission, estimates of reproductive numbers, and vaccination levels needed to prevent transmission. However, estimating the serial interval requires knowledge of individuals’ contacts and exposures (who infected whom), which is typically obtained through resource-intensive contact tracing efforts. We develop an alternate framework that uses virus sequences to inform who infected whom and thereby estimate serial intervals. The advantages are many-fold: virus sequences are often routinely collected to support epidemiological investigations and to monitor viral evolution. The genomic approach offers high resolution and cluster-specific estimates of the serial interval that are comparable with those obtained from contact tracing data. Our approach does not require contact tracing data, and can be used in large populations and over a range of time periods. We apply our techniques to SARS-CoV-2 sequence data from the first two waves of COVID-19 in Victoria, Australia. We find that serial interval estimates vary between clusters, supporting the need to monitor this key parameter and use updated estimates in onward applications. Compared to an early published serial interval estimate, using cluster-specific serial intervals can cause estimates of the effective reproduction number *R*_*t*_ to vary by a factor of up to 2–3. We also find that serial intervals estimated in settings such as schools and meat processing/packing plants tend to be shorter than those estimated in healthcare facilities.

## 1 Introduction

Whole genome sequence (WGS) data is rapidly becoming a fundamental tool in public health laboratories (PHL) around the world [1–3]. WGS data carry enormous benefits for outbreak investigations, providing finer discriminatory power than traditional sub-typing [4], and facilitating surveillance of AMR [5] as well as easy sharing across jurisdictional boundaries [6]. However, the information content of genomic data alone can be limited under certain conditions, as experienced during the SARS-CoV-2 pandemic [7, 8]. Such situations highlight the limited ability to effectively integrate genomic and epidemiological data in public health. Often, genomic data is combined with epidemiological data in an *ad hoc* fashion by plotting epidemiological data on the tips of phylogenetic trees (derived from genomic data). The results are then interpreted by PHL genomic epidemiology personnel. This approach relies heavily on the interpretation experience of the person generating the report, and thus does not lend itself readily to the desired reproducibility and repeatability standards under PHLs standard operations. In addition, the lack of a more systematic framework for integrating genomic and epidemiological data is leading to significant under-utilization of the information available to PHLs to help inform public health action.

In this work, we develop a framework for the integration of genomic data, in the form of whole-genome virus sequences, into epidemiological investigations, particularly when detailed epidemiological data from contact tracing is unavailable. Pathogen sequence data collected from infected individuals do not directly reveal who infected whom, but nonetheless can offer a high-resolution view of transmission. We focus on cluster-specific estimation of the serial interval, a key measure describing the spread of an infectious disease, which is defined as the length of time between the onset of symptoms in a primary and secondary case. This is informative of both the speed of transmission, as well as when in the infection process transmission is likely to occur.

Serial intervals are typically inferred from small clusters of individuals with known contact and times of symptom onset [9, 10], but collection of such data can be resource intensive, and privacy and reporting considerations limit wide reporting and utilization. As a result, estimates of the serial interval applied in practice (underpinning estimation of other epidemiological quantities such as the time-dependent reproduction number *R*_*t*_) are often taken from small studies, not necessarily from the same population or time as the population in question. Methods that do not require knowledge of who infected whom have been developed, but these assume that the population is fully sampled [11, 12]. When contact tracing data is available, common approaches are to consider the distribution of observed serial intervals between contact-traced pairs assumed to represent direct transmission [13], or to monitor the population for index cases who are infected with the pathogen of interest, and then follow up with close contacts such as members of their household to find secondary cases [14]. Such approaches were extended by Vink et al. [9] to allow for unsampled intermediate cases, using ‘index case-to-case (ICC) intervals’, defined as the lengths of time between symptom onset of all secondary cases and the index case in a small population such as a household, boarding school or closed workplace. By allowing for up to two unsampled cases between the index case and a secondary case, Vink et al. take potential under-reporting into account. However, the limitation on the number of unsampled intermediates and the onus on identification of the index case mean that this approach is most suited to small and closed populations. In this work we present a framework that uses virus sequences in place of direct knowledge of infection pairs. It does not restrict the number of unsampled intermediate cases and it incorporates uncertainty in who infected whom. We explore how estimates of the serial interval can vary between outbreak clusters, even within the same population, and demonstrate that virus sequences offer a practical approach for inference of cluster-specific estimates, even where detailed contact tracing data are not available and whilst taking under-reporting into account.

We investigate the use of virus sequences for estimation of cluster-specific serial intervals using SARS-CoV-2 whole-genome sequences and recorded symptom onset times from Victoria, Australia. We identify a number of genomically- and epidemiologically-defined SARS-CoV-2 clusters from the first and second ‘waves’ of the COVID-19 pandemic in Victoria: with samples collected 6 January–14 April 2020 and 1 June–28 October 2020. We estimate the serial interval in each cluster, allowing for comparison both within and between waves. We additionally compare estimates of the serial interval arising from different types of clusters: healthcare facilities, workplaces and so on. Our approach incorporates uncertainty in who infected whom by sampling a set of feasible transmission networks given the virus sequences and the known times of symptom onset, but requires no knowledge of contact between individuals in the clusters. We then use a mixture model for estimation of the serial interval, which takes into account that the outbreak may not have been fully sampled and so inferred transmission pairs may not represent direct transmission. We explore the impact that using these cluster-specific serial interval estimates has on downstream estimates of the time-dependent reproduction number *R*_*t*_. We compare our sequence-based approach with an equivalent method which uses the same model but detailed contact-tracing information in place of virus sequences.

## 2 Materials and Methods

### 2.1 SARS-CoV-2 whole genome sequencing and clustering, Victoria, Australia

For full details on the sequencing procedure, see [7]. In short, whole RNA was extracted from samples obtained from nasopharyngeal swabs, and positive samples for SARS-CoV-2 were identified by RT-qPCR. Postive samples underwent tiled amplicon sequencing using ARTIC primers [15,16]. Amplicons were prepared for sequencing on an Illumina sequencer using the NexteraXT library prep protocol following the manufacturer’s instructions. Sequencing reads were mapped to the Wuhan-Hu-1 reference sequence (Genbank MN908947.3) and consensus sequences generated using the iVar pipeline [17]. Consensus sequences were kept for downstream analyses if they met the following criteria: ≥ 95% genome recovered, ≤ 25 SNPs from the reference genome, and ≤ 300 ambiguous bases.

#### Wave 1 clustering

1242 samples from 1075 patients were sequenced during the wave 1 study period: 6 January–14 April 2020. This corresponds to 80.7% of identified COVID-19 cases in Victoria during that time period. The data comprise genomic sequence, sequence sampling date and symptom onset date for each sampled case.

The wave 1 genomic clustering procedure is as described in [7]. Of 903/1242 samples passing initial quality control and de-duplication, 737 were identified as belonging to a genomic cluster, for a total of 76 clusters with at least 2 cases. Clustering of the samples was based on a maximum likelihood phylogenetic tree containing a single sequence per patient built using IQtree [18] applying a GTR+Γ4 substitution model. Using the ClusterPicker tool [19], clusters were defined as having at least two samples with the inferred ancestral node having at least 95% bootstrap support and the maximum distance within the cluster of 0.0004 expected substitutions/site. We focus on all genomic clusters with at least 15 cases for the serial interval analysis. After removing sequences with <95% genome recovered or with missing symptom onset time (3 cases), the wave 1 data comprise a total of 10 wave 1 clusters containing a total 312 cases. These clusters are summarized in Table 1.

**Table 1:** Details of the primary Victoria SARS-CoV-2 clusters. *Note: wave 2 cases may appear in multiple clusters.

| **Wave 1** | **Cases** | **Onset date range** |
| --- | --- | --- |
| Cluster A1 | 29 | 2020/02/18 – 2020/04/01 |
| Cluster A2 | 61 | 2020/02/28 – 2020/04/02 |
| Cluster A3 | 21 | 2020/02/28 – 2020/04/05 |
| Cluster A4 | 28 | 2020/02/29 – 2020/03/29 |
| Cluster A5 | 19 | 2020/03/08 – 2020/03/31 |
| Cluster A6 | 24 | 2020/03/10 – 2020/04/09 |
| Cluster A7 | 59 | 2020/03/10 – 2020/04/06 |
| Cluster A8 | 29 | 2020/03/10 – 2020/03/28 |
| Cluster A9 | 30 | 2020/03/10 – 2020/04/06 |
| Cluster A10 | 18 | 2020/03/15 – 2020/04/05 |
| Total | 318 | 2020/02/18 – 2020/04/09 |
| **Wave 2** | **Cases** | **Dates** |
| Cluster B6 | 20 | 2020/06/14 - 2020/07/26 |
| Cluster B7 | 59 | 2020/06/14 - 2020/07/28 |
| Cluster B13 | 115 | 2020/06/18 - 2020/08/27 |
| Cluster B17 | 142 | 2020/06/27 - 2020/08/19 |
| Cluster B24 | 59 | 2020/07/04 - 2020/08/20 |
| Cluster B33 | 117 | 2020/07/07 - 2020/08/19 |
| Cluster B43 | 111 | 2020/07/12 - 2020/08/02 |
| Cluster B47 | 157 | 2020/07/13 - 2020/09/26 |
| Cluster B53 | 19 | 2020/07/15 - 2020/07/28 |
| Cluster B54 | 44 | 2020/07/15 - 2020/08/29 |
| Total | 818* | 2020/06/14 - 2020/09/26 |

#### Wave 2 clustering

Even when pathogen samples are routinely collected and sequenced, genomic clustering can have computational challenges and lead to large, uncertain clusters, particularly when transmission is ongoing and widespread in a community and many cases are sampled. In this context, the concept of what constitutes a cluster is less clearly defined, and tree- or sequence-based clustering can lead to infeasibly large clusters. Public health experts may also be interested in transmission not simply among genetically linked isolates, but within particular locations or groups of people, for example schools and hospitals. Driven by these factors, for the wave 2 analysis we derive clusters epidemiologically rather than genomically: choosing all cases associated with particular exposure sites of interest to public health.

15665 samples from 14075 patients were sequenced during the wave 2 study period: 1 June– 28 October 2020. This corresponds to 83.9% of identified COVID-19 cases in Victoria during that time period. As in wave 1, the data consist of the virus sequence, sequence sampling date and symptom onset date for each sampled case. Epidemiological data, including detailed demographic, risk factor, and contact tracing data were additionally collected for each case through an interview conducted by the Victorian Department of Health, and these data were used to determine locations of possible transmission, known as exposure sites. In our primary analyses, we use only the exposure site information (which cases attended which sites) to define the wave 2 clusters, we do not use any demographic or contact data. As a validation of our overall approach, we perform a supplementary analysis in which we re-estimate the serial intervals without the viral sequences but given full knowledge of the contact data.

Of the 10642/15665 samples passing quality control and de-duplication, 7116 were identified as associated with at least one exposure site. Of these samples, we remove 1371 cases with missing symptom onset time. This provides a total 623 clusters with at least 2 cases. Note that cases may be associated with more than one exposure site, in which case they will be included in more than one cluster. For our primary analysis we focus on 10 key exposure site clusters - we select the largest 2 clusters associated with each of: aged care facilities, healthcare facilities, housing, schools, and meat packing/meat processing plants. These clusters are summarized in Table 1. We perform additional analysis on all wave 2 clusters with at least 15 cases. We remove a further 2 clusters which contain more than 15 cases but have limited signal of transmission (5 or fewer inferred plausible infector-infectee pairs, see Section 2.2). This results in a total of 94 wave 2 clusters comprised of a total 3875 cases.

Phylogenetic trees for both wave 1 and wave 2 data are shown in Figure 1. Phylogenies were produced from the entire set of sequences in each wave, using a custom workflow for building fast SARS-CoV-2 trees available at github.com/MDU-PHL/kovid-trees-nf. Briefly, sequences were cleaned to remove sites with > 5% missing calls and de-duplicated with GOALIGN [20]. An approximate maximum-likelihood tree was built using FastTree [21] and the branch lengths optimised with RAxML-NG [22]. Finally, GOTREE [20] was used to repopulate the tree with duplicate sequences. Symptom onset curves for the 20 primary clusters are shown in Figure 2.

**Figure 1:**
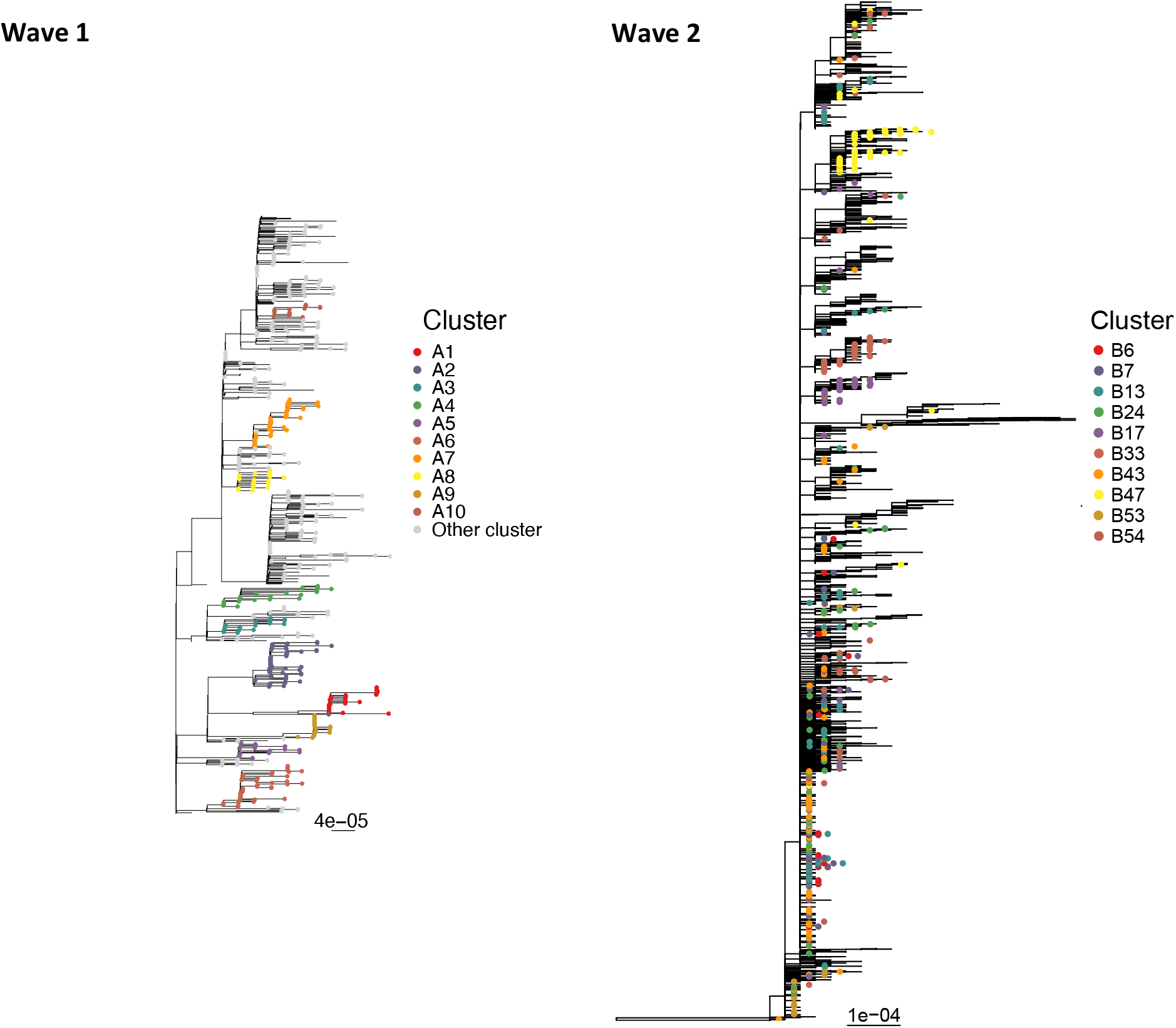
Phylogeny of wave 1 (left) and wave 2 (right) Victorian SARS-CoV-2 data. Primary clusters selected for serial interval analysis are coloured and labelled.

**Figure 2:**
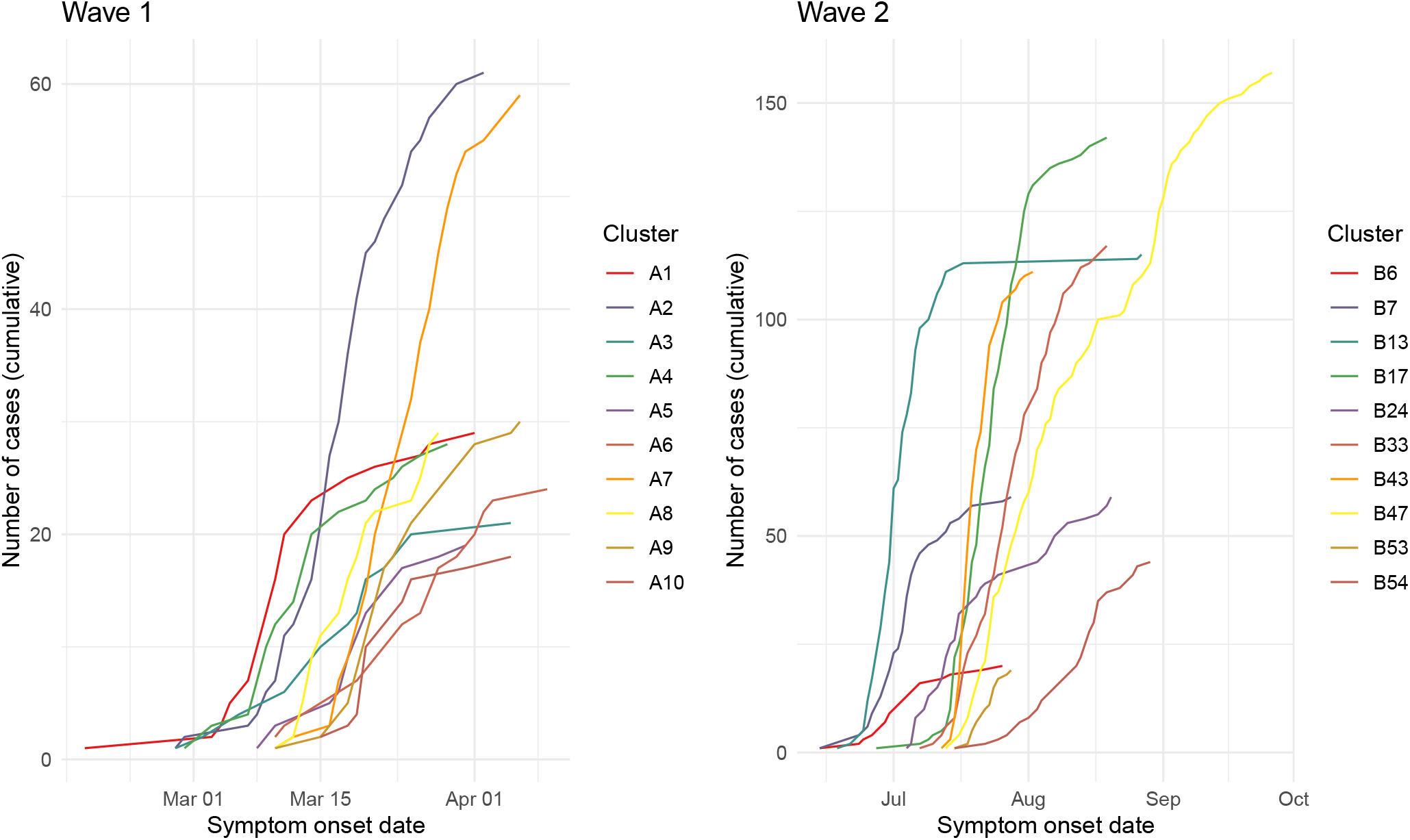
Symptom onset curves of primary wave 1 (left) and wave 2 (right) clusters.

### 2.2 Sampling transmission networks

In order to estimate cluster-specific serial intervals from viral sequences and symptom onset times in a cluster (i.e. in the absence of contact tracing data), we require an approach for obtaining putative transmission pairs. Given that there will usually be considerable uncertainty in who infected whom, we sample a set of plausible transmission networks, accounting for indirect transmission, and perform parameter estimation across the entire set.

We build a pairwise genetic distance matrix between all aligned sequences in a cluster using the *ape* package in *R* and the TN93 model of evolution. We then create a ‘transmission cloud’, that is, a set of all plausible transmission pairs in the cluster. We define a plausible (possibly indirect) transmission pair as 2 cases in the same cluster who:

1. have observed interval between symptom onsets 0 < *T* ≤ 35 days
2. have pairwise genomic distance *G* < 1.1*/*29903 (where 29903 is the alignment length).

Note that this approach may result in a case having multiple plausible infectors, or no plausible infectors: in the second scenario the case would be considered as an importation to the cluster from an unsampled case.

From the transmission cloud, we sample a plausible transmission network by sampling an infector for each infectee from among their list of plausible infectors. For infectee *j*, a plausible infector *i* is selected with higher probability if the genomic distance and/or the difference in symptom onset time between *i* and *j* is lower. The sample weighting *s*(*i, j*) is given by

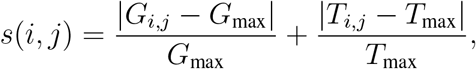

where *G*_*i,j*_ is the genomic distance between *i* and *j*, and *T*_*i,j*_ is the difference in symptom onset time between *i* and *j. G*_max_ and *T*_max_ are the maximum genomic and time distance, respectively, among all plausible pairs in the cluster; their inclusion causes distances *G*_*i,j*_ and *T*_*i,j*_ close to zero to result in smaller sampling probability *s*_*i,j*_. The weighting is normalized to sum to 1 for each infectee *j*. We repeat this process *N* times to obtain a set of *N* plausible transmission networks per cluster. When *i* is a “plausible infector” of *j*, we incorporate the possibility that infection was indirect, through an unknown number of unsampled intermediate cases, or that *i* and *j* were both infected by the same unsampled individual (see below).

As we do not assume that all cases have a sampled ancestor within their cluster, a sampled transmission network is not required to be comprised of a single transmission tree; there can be several distinct sub-trees each spawned by an unsampled case. The result is that we only perform serial interval estimation on those inferred transmission pairs with sufficient confidence.

### 2.3 Serial interval estimation

After sampling a set of plausible transmission networks per cluster, we proceed to estimate the serial interval in each network, and finally combine these for a single cluster estimate. Our approach seeks to estimate the parameters of the true underlying serial interval distribution, that is, the serial interval arising from direct transmission between a pair of individuals. However, we account for the fact that observed serial intervals in data with incomplete sampling may comprise a mixture of direct transmission (*i* → *j*), indirect transmission (*i* → *x* → *j*, for any number of unsampled *x*) and coprimary transmission (*x* → *i, x* → *j*, for unsampled *x*).

We assume that the serial interval follows a Gamma distribution with mean *μ* and standard deviation *σ*. We further assume that any sampled infector *i* and sampled infectee *j* will be separated by some *m* unsampled individuals, 0 ≤ *m* < ∞, where *m* follows a geometric distribution with parameter *π*. Then, *π* can be thought of as a sampling probability within an identified transmission chain, *m* = 0 corresponds to direct transmission and *m* > 0 corresponds to indirect transmission. The distribution of the observed time interval between *i* and *j* in the case of direct or indirect transmission therefore follows a *Compound Geometric Gamma* distribution, CGG(*μ, σ, π*): it is the sum of *m* + 1 i.i.d. Γ(*μ, σ*) random variables. We assume that a proportion *w* of observed serials are of this type.

We assume that the remaining proportion (1 − *w*) of the observed time intervals represent “coprimary” transmission: *i* and *j* were infected by the same unsampled individual. Here, the observed time interval between *i* and *j* can be thought of as the (strictly non-negative) difference between two i.i.d. Γ(*μ, σ*) random variables. It therefore follows a *Folded Gamma Difference* distribution, FGD(*μ, σ*).

For a sampled transmission network *τ* in cluster *c* with *n* infector-infectee pairs, the log likelihood of the model parameters given the set of observed time intervals in the network, 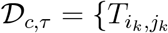, for *k* = 1, 3, …, *n*}, can be calculated as a sum over all infector-infectee pairs independently, and is given by,

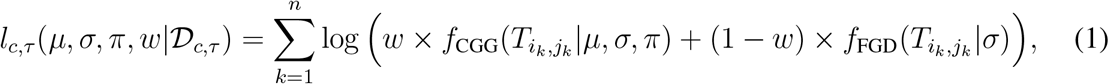

where *f*_CGG_ and *f*_FGD_ are the PDFs of the Compound Geometric Gamma and Folded Gamma Difference distributions, respectively.

We combine the likelihood in Equation (1) with log prior distributions for *π* and *w*, and then find *maximum a posteriori* (MAP) estimates of the model parameters (*μ, σ, π, w*). The MAP approach allows us to refine the estimation using prior knowledge of the fraction of cases with isolates that are sequenced. We assume a Beta distributed prior for both *π* and *w*, and calculate the associated log posterior distribution. The log posterior is then numerically maximized, using the *optim* function in *R*, to obtain MAP estimates of the parameters for each sampled transmission network. We lastly combine the MAPs across all sampled transmission networks *τ* in each cluster *c*, to obtain a single estimate of the model parameters per cluster. Corresponding 95% confidence intervals are calculated, given both the uncertainty within each sampled network MAP and the variation in MAPs between sampled networks.

A full description of the model, likelihood and estimation approach is provided in the Supplemental Materials S1. A schematic diagram of the method is shown in Figure 3. We apply this methodology to the Victorian SARS-CoV-2 data by estimating the serial interval in each of the 10 primary wave 1 and wave 2 clusters shown in Table 1, as well as the additional 84 wave 2 clusters. We sample 100 transmission networks per cluster, and perform an additional analysis in which we pool the sampled networks in each wave, to obtain aggregated whole-wave serial interval estimates. Using the available exposure site data for the wave 2 clusters, we compare the cluster-specific serial interval estimates across exposure site types. We assume a Beta distributed prior Beta(12, 11) for parameters *π* and *w* in all analyses, with mean 0.52 and standard deviation 0.1. This was informed by the proportion of identified Victorian COVID-19 cases during the study period that were sampled and sequenced with sufficient quality (57%), combined with a prior belief of a high case finding rate. Sensitivity to the choice of prior distribution is explored in the Supplemental Materials, Section S3.

**Figure 3:**
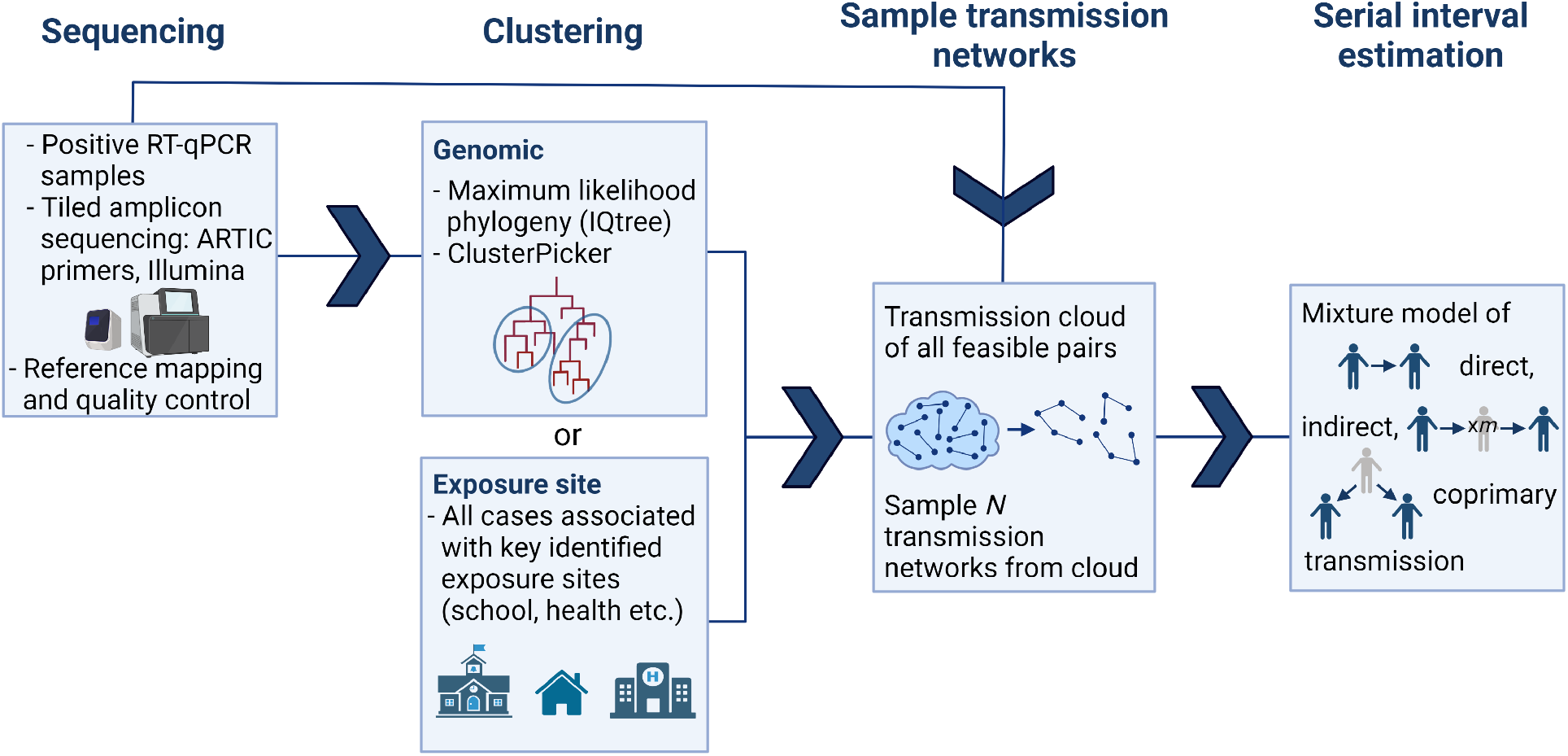
Model schematic.

The above described model is not limited to analyses using genomic data. We validate our genomic approach with a supplemental analysis that applies the same serial interval estimation procedure to epidemiologically-sampled transmission networks from epidemiologically-defined clusters. Here, we define case clusters as the connected components of a network created from contact tracing data. Rather than sampling transmission trees using viral sequences, we preferentially sample infector-infectee pairs with known direct contact as indicated by contact tracing. Full details and results are included in the Supplemental Materials, Section S4. We also compare our cluster-specific serial intervals for COVID-19 to several published estimates.

### 2.4 Estimation of effective reproduction number *R*_*t*_

To contextualize the effects of using cluster-specific serial intervals in downstream analyses, we compare estimates of the effective reproduction number *R*_*t*_ (also known as the time-varying re-production number) using cluster-specific serial intervals and literature-based serial intervals. *R*_*t*_ can be interpreted as the expected number of secondary cases caused by an index case at time *t*. Calculating it relies on the assumption of an underlying serial interval distribution. We use the *R* package *EpiEstim* to estimate *R*_*t*_ on a weekly sliding window in each cluster, using both the cluster-specific serial interval distribution estimated in this work and a Γ(*μ* = 6.3, *σ* = 4.2) distribution as estimated for SARS-CoV-2 by Bi et al. [23]. We compare the estimated *R*_*t*_ values.

### 2.5 Code availability

Analyses in sections 2.2–2.4 are performed using *R* version 4.1.0. Code is available at github.com/jessicastockdale/genomicSIs.

## 3 Results

We estimate the parameters of the serial interval distribution independently for each cluster identified in Table 1, as well as across two combined sets of all wave 1 and all wave 2 clusters. Figure 4 shows the cluster-specific mean and 95% confidence interval estimates of the serial interval mean *μ* and standard deviation *σ*, the sampling probability within identified transmission chains *π*, and the proportion of non-coprimary transmission *w*. We find varying estimates of the serial interval by cluster, with means ranging from 2.64 days (B47) to 6.74 (B43) in the primary clusters. The overall mean across all wave 1 clusters is 4.65 days, and across all wave 2 clusters is 5.17 days, though there is considerable variation between clusters, causing considerable uncertainty when combining across them. Although most of the cluster confidence intervals overlap, we do find some clusters with significantly different means or standard deviations. For example, the confidence intervals in the aforementioned clusters B47 and B43 are completely disjoint. Figure 5 shows the full serial interval distribution estimated in each cluster: in wave 2 we find a larger difference between clusters, with some clusters (B47 in particular) showing strong signal of a shorter serial interval. Full results are included in Table S3 in the Supplemental Materials, presented alongside several published estimates of the COVID-19 serial interval in Table S2 for comparison. Our estimates are in general agreement with others from wild-type SARS-CoV-2, though with wider uncertainty (Figure S7). However, the majority of these published estimates did not take uncertainty in who infected whom, co-primary or indirect transmission into account. Estimates of the parameters concerning sampling rate, *π* and *w*, are controlled relatively strongly by their prior distributions (prior mean 0.52, overall posterior mean 0.59 for *π* and 0.55 for *w*, Figure 4), but we do observe some clusters pushing against this, most notably cluster B53, in which the data suggest a lower sampling rate (*π* = 0.47). A sensitivity analysis to these assumed prior distributions, included in the supplemental materials Section S3, reveals that our results are robust to moderate changes in the sampling rate parameter (*π* and *w*) priors.

**Figure 4:**
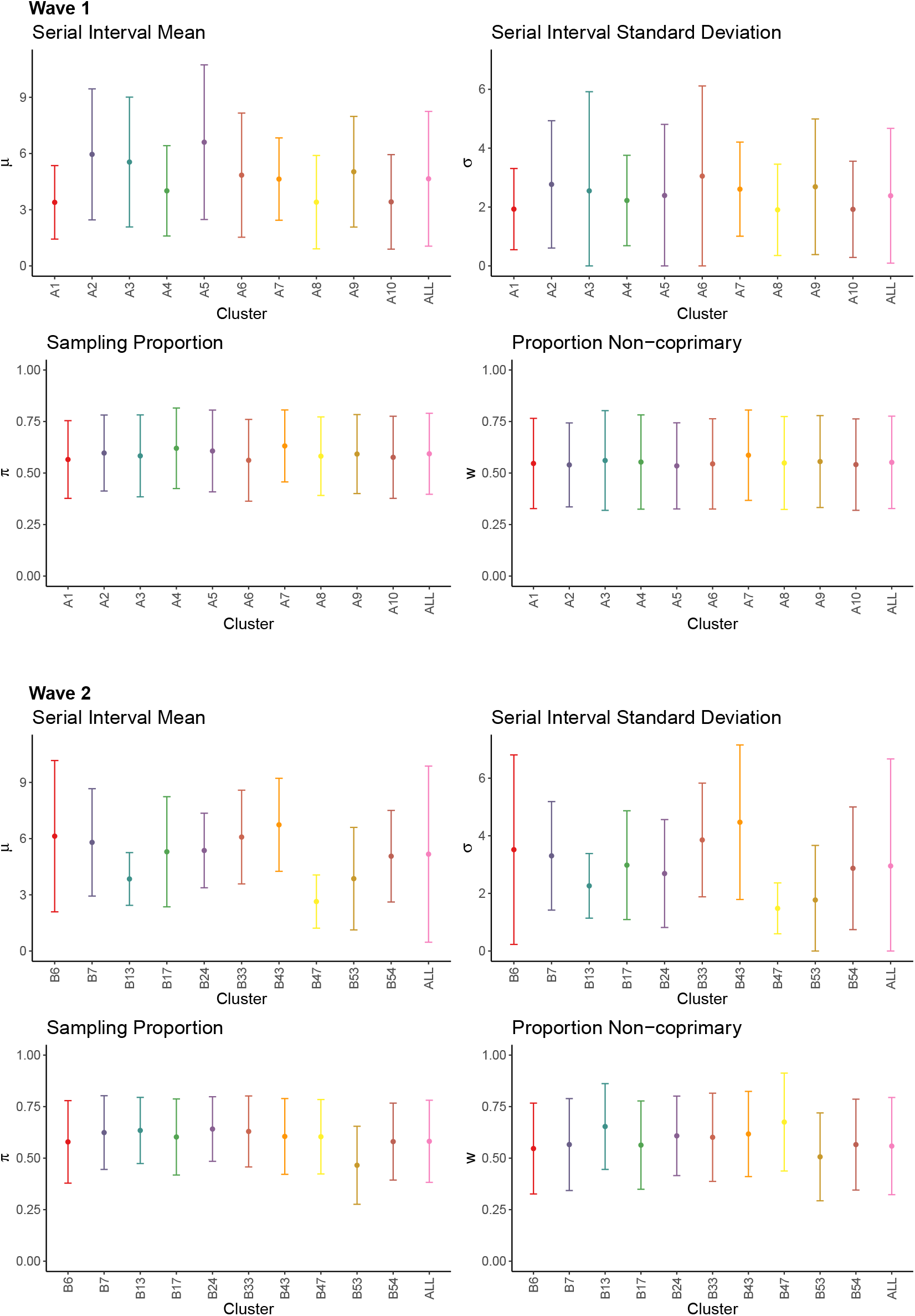
Cluster-specific estimates of model parameters. Wave 1 (top) and wave 2 (bottom). Mean estimates shown as points and 95% confidence intervals as bars. Clusters are labelled chronologically by date of earliest symptom onset. ‘All’ result in wave 2 is calculated from the full set of 94 clusters.

**Figure 5:**
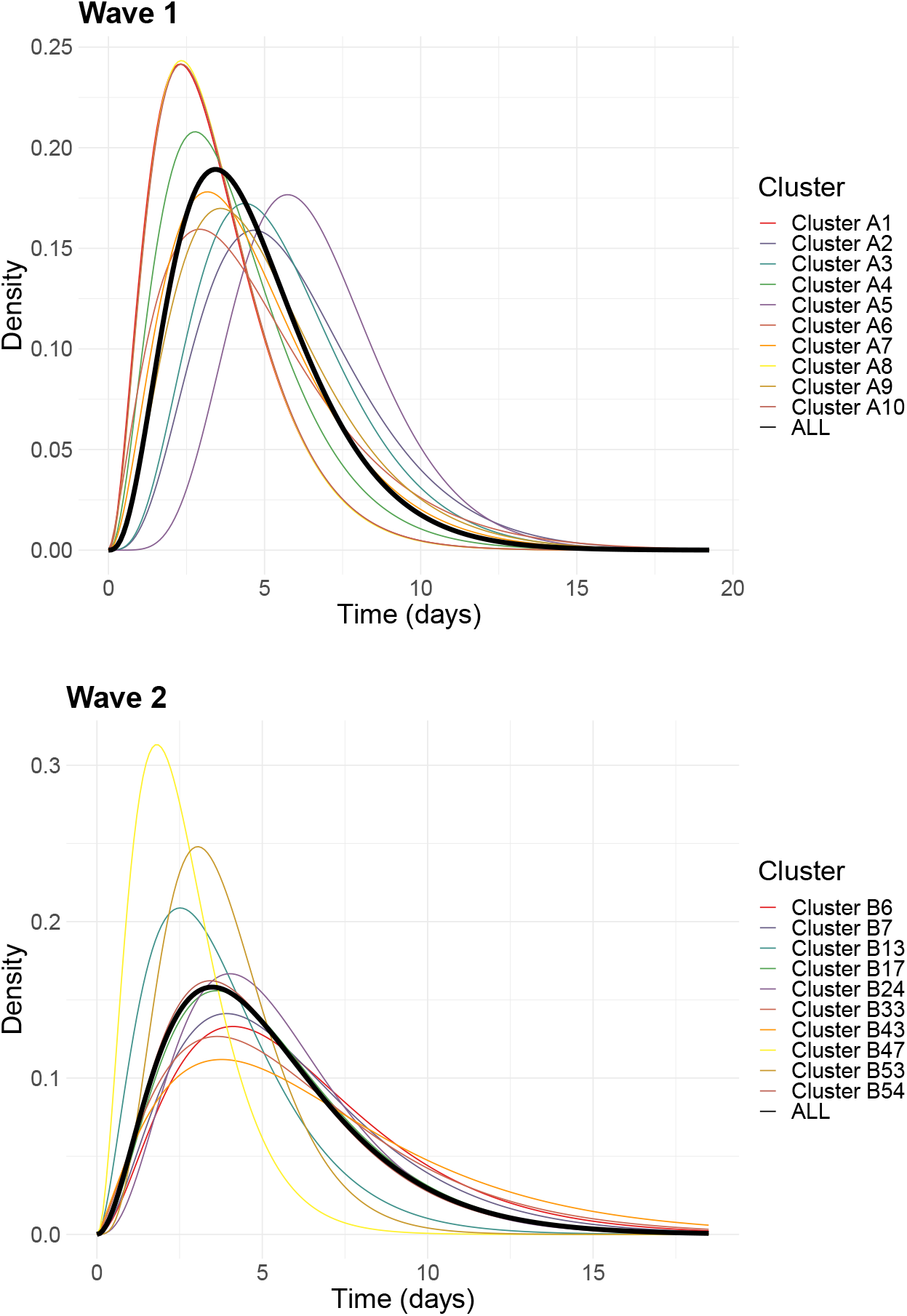
Cluster-specific serial interval distributions. Wave 1 (top) and wave 2 (bottom). Gamma distributed, calculated from mean estimates of *μ* and *σ*. ’All’ result in wave 2 is calculated from the full set of 94 clusters.

We compare estimates of the effective reproduction number *R*_*t*_ in which the underlying serial interval distribution is (i) our cluster-specific estimate and (ii) a published estimate for COVID-19, Γ(*μ* = 6.3, *σ* = 4.2), from Bi et al. [23], in Figure 6. Any difference in *R*_*t*_ in these figures arises solely from difference in the underlying serial interval distribution. Although some clusters remain largely unaffected, for some clusters the estimate of *R*_*t*_ can differ by up to a factor of 2 or 3. This occurs primarily in early cluster estimates of *R*_*t*_, highlighting how even small amounts of uncertainty in the underlying transmission model should be treated carefully when obtaining initial estimates. But even towards the middle or end of cluster outbreaks, the choice of serial interval can be the difference between an estimate of *R*_*t*_ < 1 and *R*_*t*_ > 1 (e.g. A10), which has clear implications for epidemic control.

**Figure 6:**
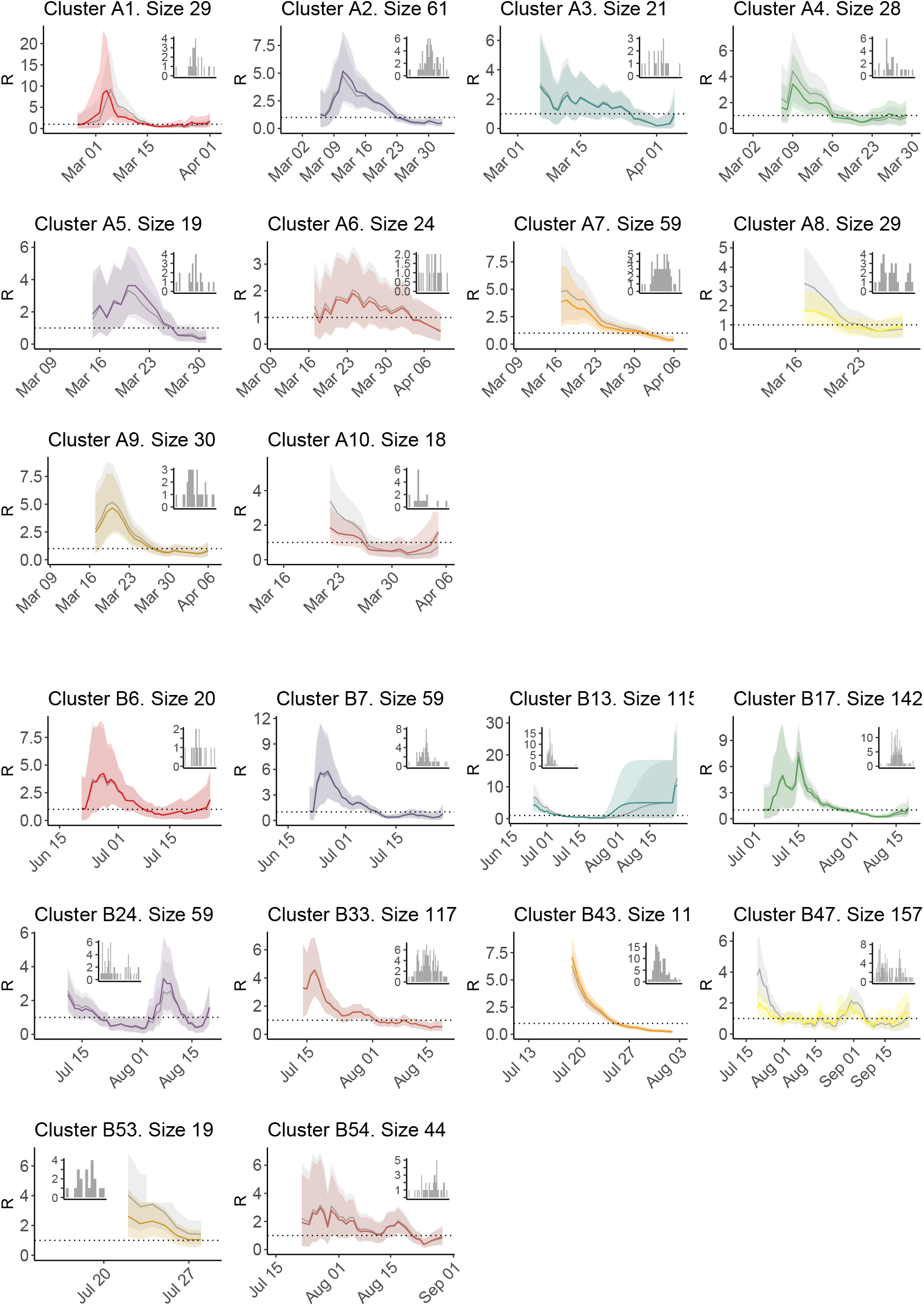
Estimates of *R*_*t*_ using cluster-specific serial intervals (in colour), for wave 1 (top) and wave 2 (bottom). In grey, we compare *R*_*t*_ as estimated using a fixed serial interval distribution, Γ(*μ* = 6.3, *σ* = 4.2), from Bi et al. [23]. Insets show daily incident case counts in each cluster.

We perform additional analysis on the entire set of 94 wave 2 clusters categorized by exposure site type, shown in Figure 7. Estimates of the mean serial interval range from 1.97 to 9.54, though many of the largest estimates have considerable uncertainty. The results suggest some patterns by exposure type: with meat packing/meat processing plants and schools among the shorter serial intervals and healthcare facilities and housing among the longer. As in the primary clusters, sampling proportion *π* and proportion non-coprimary *w* do not greatly diverge from their prior distributions. There are several exceptions, such as clusters B53, B71, and B82, all with *π* ≤ 0.5, suggesting a lower rate of case acquisition. Among the set of all wave 2 clusters, we find significant correlation (PCC (Pearson correlation coefficient) 0.35, p-value 0.0007) between sampling proportion *π* and cluster size, suggesting that larger clusters had higher inferred sampling rates (or conversely that better-sampled clusters had more cases; we note the correlation but do not speculate on the cause). We also find a significant negative correlation (PCC -0.28, p-value 0.006) between mean serial interval *μ* and the initial symptom onset date in the cluster, suggesting that later clusters were associated with shorter serial intervals (Supplementary Figure S1). We do not find any significant association between mean serial interval and cluster size or sampling rate and initial onset date, and our conclusions are the same when calculating Spearman’s correlation on the ranks.

**Figure 7:**
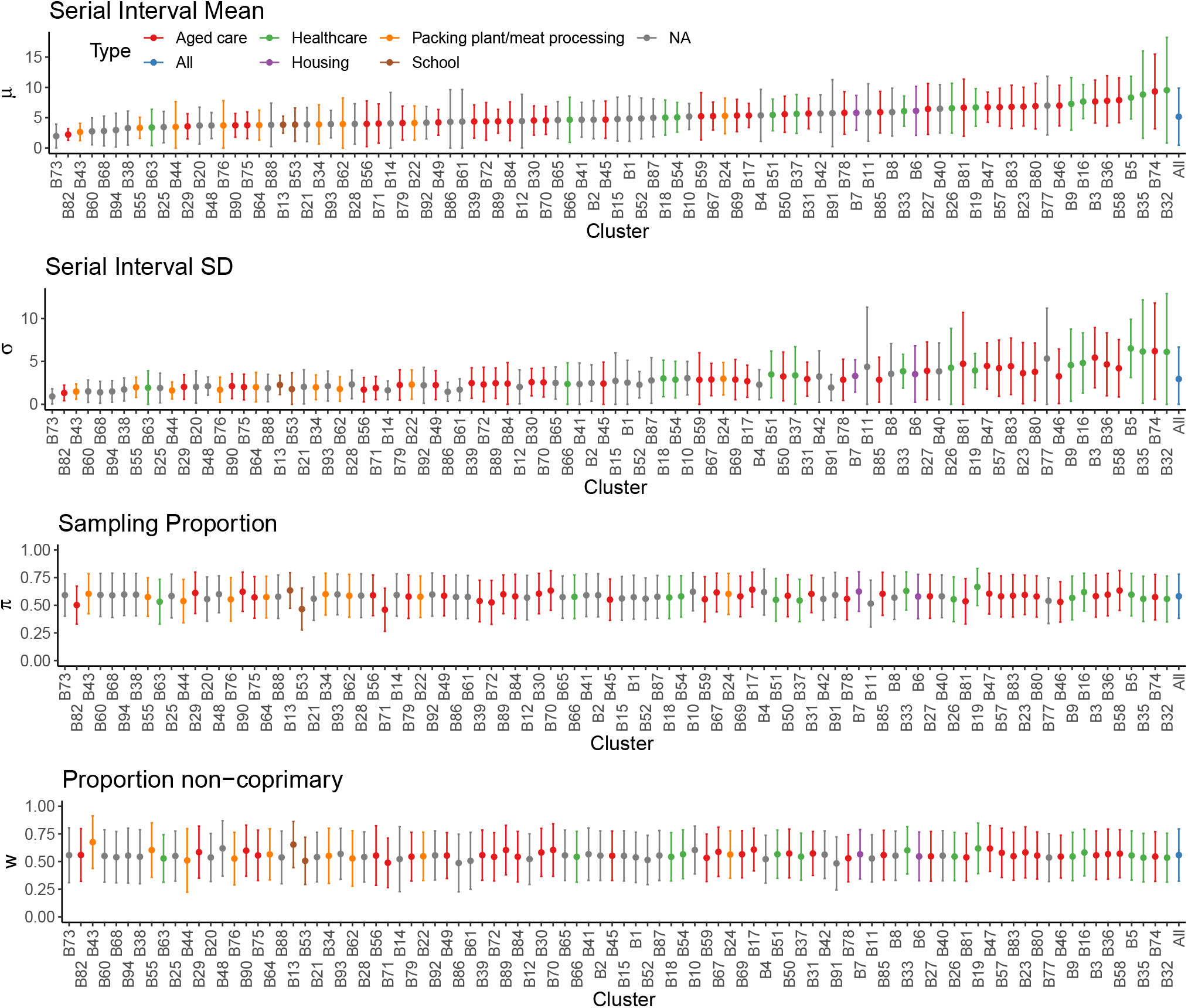
Model parameter estimates for all wave 2 clusters, categorized by exposure site type. Mean estimates shown as points and 95% confidence intervals as bars. Clusters are ordered by increasing mean serial interval.

In order to validate our genomic approach, we perform a supplementary analysis in which we use the same serial interval model, but define the clusters and transmission networks epidemiologically (from contact-tracing data) instead of genomically. The 13 epidemiologically-defined clusters for wave 1 are shown in Figure S4; they are of a comparative size to the genomic clusters. Cluster-specific estimates of the mean serial interval range from 3.02 days (cluster C4) to 7.71 days (cluster C40), with an ‘all clusters’ estimate of 5.04 days (Figure S5). These estimates are similar to those obtained from using viral sequences.

## 4 Discussion

Estimates of the serial interval are key to understanding disease spread, and underlie estimation of other quantities such as the time-dependent reproduction number. Current methods for serial interval estimation are best suited to small, contained populations with high sampling, and require detailed contact studies which can be resource intensive. However, there is growing demand for wide-scale epidemiological analyses across populations, and as such these are often undertaken using estimates of the serial interval from small, early outbreaks in a different location or time than the population under study. Serial intervals are known to contract during disease outbreaks [24], but they may also be changed under different pathogen strains, population mixing or control strategies. In this work, we sought to explore cluster-specific estimates of the serial interval within two waves of the COVID-19 pandemic in Victoria, Australia, through the introduction of a new approach using viral sequences in place of detailed contact data.

We found modest differences between clusters, despite all cases occurring within one population over a 7 month period. Within the second wave, there was an overall signal of serial intervals shortening over time. We also found that clusters occurring in sites associated with longer-term contact, such as healthcare centers and housing, tended to have longer serial intervals than sites attended for shorter lengths of time, such as meat packing or meat processing plants and schools. Although the parameters concerning sampling rate were relatively strongly controlled by a prior distribution, some clusters suggested higher/lower amounts of sampling: such findings could be used to monitor developing outbreaks, especially if the approach was integrated in routine genomic surveillance in real time. We found that using cluster-specific serial intervals in estimation of the time-dependent reproduction number as compared to a literature-based estimate changed *R*_*t*_ by up to a factor of 2-3, particularly early in outbreaks. This highlights how variable estimates of the reproduction number can be, particularly when calculated from small outbreaks in specific settings, and suggests caution should be taken when applying existing parameter estimates to analyses of new outbreaks.

There are several limitations of our methodology. Due to the assumption of gamma distributed serial intervals (required for the construction of the mixture model), we assume serial intervals are strictly positive. There is evidence of negative serial intervals for COVID-19 [13] and in other diseases, and an extension of this model could allow for this. We perform transmission tree sampling from a cloud of plausible infector-infectee pairs, allowing for indirect and coprimary transmission, in order to take into account uncertainty in who infected whom from the genomic data. Although we do this in a probabilistic way which aims to approximate the judgement applied in public health (that cases closer in pathogen sequence and in time are more likely to infect one another), the tree sampling could be improved by incorporating additional epidemiological data e.g. known pairs from contact tracing or relative Ct values, if this were available. It may also be possible to incorporate existing methodologies for sampling of transmission trees, such as the *outbreaker* or *TransPhylo* platforms [25, 26], but these are not well positioned to estimate serial intervals. Lastly, our approach requires prior knowledge of the population sampling rate. This is really an innate identifiability problem with estimation of serial intervals using any model, in that short serial intervals with low sampling would be indistinguishable from long serial intervals with high sampling. However, the sensitivity analysis of our prior assumptions revealed that our results are robust to moderate deviations from the sampling rate priors.

Our genomic approach was able to obtain estimates of the model parameters with relatively high certainty; confidence intervals were comparable to those from the supplemental analysis using contact data. Our results show wider uncertainty than many published estimates, but the majority of these estimates focused on small populations with known contact pairs and did not take potential under-reporting into account. This further highlights a need for repeated estimation of parameters concerning infection and transmission, both to obtain a consensus and to track whether values are changing in time.

A major benefit of the methodology we have presented here is that genomic data can offer a high resolution view of transmission at a large scale, where collection of contact-tracing data may be expensive or infeasible. Particularly during the COVID-19 pandemic, many public health labs have begun routine sample collection and sequencing from a high proportion of identified COVID-19 cases, whereas contact tracing teams can be overwhelmed by high case loads, and data collection and sharing is challenging. Our approach could be used beyond serial interval estimation, for example to compare differences in transmission of COVID-19 Variants of Concern (VOC), between settings, or between times under particular non-pharmaceutical interventions (NPI). If used in real time, it could suggest clusters to focus resources upon, for example those which suggest a lower sampling rate, more rapid transmission, or substantially different/uncertain serial intervals. Although reconstruction of the true transmission chain is not our primary aim, rather we consider the space of plausible chains, there is also opportunity to further explore the genomically-defined sampled transmission networks: for example, to consider patterns of transmission by age, vaccine status or other factors, and whether these change over time. As indicated by the supplementary contact-based analysis, the estimation model presented here, which is novel in and of itself, is not limited to genomic analyses. We explored how either genomic or contact data alone can teach us about transmission, but in practice a combination of data sources may result in the best estimates.

More widely, this research enhances the contributions that virus sequences can make to understanding transmission dynamics. To date, phylodynamics has typically operated at the large scale of global phylogeography [27, 28] and estimation of the past population dynamics of pathogens at the whole-population scale [29, 30]. Genomic epidemiology has, in contrast, had a high level of focus on establishing who infected whom or otherwise analyzing person-to-person transmission [25,26]. Our work establishes an intermediate regime for genomic epidemiology: transmission analysis at the level of small to intermediate groups, in settings where there is insufficient information to identify individual transmission events with high confidence. In order to fully harness this information, linkage to epidemiological data is helpful – here, times of symptom onset make the link to serial intervals, and exposure sites help to refine clusters. Particularly when this linkage is done at early stages rather than in a *post hoc* manner, analysis can be automated, relies less on human interpretation, and therefore can be incorporated into routine public health monitoring, in real-time if desired.

## Supporting information

GISAID accession numbers and laboratory acknowledgements

## Data Availability

GISAID accession numbers, originating/submitting laboratories and onset dates for all samples used in this study are included in the supplemental information.

https://github.com/jessicastockdale/genomicSIs

## 5 Acknowledgements

We thank the public health, clinical, and microbiology staff across Victoria who have been involved in the testing, clinical care and public health responses to COVID-19. We gratefully acknowledge the originating and submitting laboratories of sequences deposited in the GISAID Database that were used in this study. References to these laboratories are available as a supplementary file.

This work is funded by the Victorian Government, National Health and Medical Research Council Australia (APP1149991, APP1196103, APP1174555), Medical Research Future Fund (MRF9200006), Natural Science and Engineering Research Council (Canada) Discovery Grants (RGPIN-2019-06911, RGPIN-2019-06624), Michael Smith Health Research BC (COV-2020-1010) and the Federal Government of Canada’s Canada 150 Research Chair program. Figure 3 was created with BioRender.com.

