## Supplemental Materials for "Genomic epidemiology offers high resolution estimates of serial intervals for COVID-19"

### S1 Serial interval estimation

In this section we fully define our statistical model for estimation of the parameters of the serial interval distribution. This model takes into account that data may be under-reported, and so observed intervals between symptom onsets may be a result of direct, indirect or coprimary transmission. In this description, we assume that a plausible transmission network has already been sampled from the data (as in Section 2.2), and so transmission between a sampled infector-infectee pair $(i, j)$ is considered certain (even if that transmission may be direct, indirect or coprimary). Uncertainty in who infected whom is taken into account with repeated sampling of the transmission network, applying the methodology described in this section independently for each sample.

#### S1.1 Model

Let $T'$ be the true serial interval distribution of the disease under consideration, so that $T'_{ij}$ is the serial interval arising from *direct* transmission between any case $i$ and case $j$. We assume that $T'_{ij} \sim \Gamma(\mu, \sigma)$, for mean $\mu$ and standard deviation $\sigma$. Note that we assume, therefore, that the serial interval is strictly non-negative. This is the quantity we are interested to make inference about, by estimation of $\mu$ and $\sigma$.

Now, let $T_{ij}$ denote the *observed* time interval between the symptom onsets of a particular case $i$ and case $j$. If there is direct transmission from $i$ to $j$, denoted $i \rightarrow j$, then $T_{ij} = T'_{ij}$. However, if we allow for under-reporting in the data, then there may have been unsampled intermediate cases between $i$ and $j$, denoted $i \dashrightarrow j$, in which case $T_{ij}$ is the convolution of multiple serial intervals. It is also possible that both $i$ and $j$ were infected by the same unsampled host, which we call coprimary transmission and denote $i \hookleftarrow\!\!\hookrightarrow j$. Although we allow for a theoretically infinite number of unsampled intermediate cases in $i \dashrightarrow j$, we assume that, in the coprimary case $i \hookleftarrow\!\!\hookrightarrow j$, a single unsampled host must have infected both $i$ and $j$. This greatly simplifies the calculations, and seems reasonable so long as sampling is relatively high and/or the rules for selecting plausible infectors in the transmission cloud are relatively stringent.

More formally, we assume that the serial interval distribution is a mixture of two transmission paths: coprimary and non-coprimary (direct or with unsampled intermediates):

**Non-coprimary transmission**

Sampled infector $i$ and infectee $j$ are separated by $m$ unsampled intermediate hosts, $0 \leq m < \infty$ ($m = 0$ corresponds to direct transmission). Then, the observed interval $T_{ij}$ is the sum of $m + 1$ i.i.d. gamma distributed intervals, $T_{ij} \sim \Gamma((m+1)\mu, \sqrt{(m+1)}\sigma)$. The value of $m$ is unknown but can be considered to follow a Geometric distribution with success probability $\pi$, where 'success' corresponds to sampling an infected individual and $m$ is the number of failures. The parameter $\pi$ then represents a pseudo-sampling probability, for hosts descending from a sampled infector only. Taking this into account, the marginal distribution of $T_{ij}$ will be a *Compound Geometric Gamma* distribution, CGG$(\mu, \sigma, \pi)$.

**Coprimary transmission**

Sampled 'infector' $i$ and 'infectee' $j$ were in actuality both infected by the same unsampled case $x$. Since the serial intervals $T'_{xi}$ and $T'_{xj}$ are i.i.d. $\Gamma(\mu, \sigma)$, the observed interval $T_{ij}$ is the strictly non-negative difference of two i.i.d. gamma distributions, $T_{ij} \sim \left|T'_{xj} - T'_{xi}\right|$. We refer to this as a *Folded Gamma Difference* distribution FGD$(\mu, \sigma)$. This is a folded version of the Gamma Difference distribution introduced in [1, 2], with the simplification that both gamma distributions have the same parameters.

Let $w$ be the unknown proportion of pairs $(i, j)$ who fall into the non-coprimary transmission path, then $1 - w$ is the proportion in the coprimary transmission path.

#### S1.2 Likelihood

We wish to write down the likelihood of the model described above, given a dataset $\mathcal{D}_{c,\tau} = \{T_{i_1,j_1}, T_{i_2,j_2}, \ldots, T_{i_n,j_n}\}_{c,\tau}$ which contains the observed time intervals for all $n$ sampled infector-infectee pairs in network $\tau$ for cluster $c$. Note that any given host can be an infector of multiple cases and an infectee as well, so some $i$ and $j$ may refer to the same individuals.

The log-likelihood of the model parameters given dataset $\mathcal{D}_{c,\tau}$ is given by:

$$l_{c,\tau}(\mu, \sigma, \pi, w|\mathcal{D}_{c,\tau}) = \sum_{k=1}^{n} \log \Big( w \times f_{\text{CGG}}(T_{i_k,j_k}|\mu, \sigma, \pi) + (1 - w) \times f_{\text{FGD}}(T_{i_k,j_k}|\sigma) \Big), \quad \text{(S1)}$$

where $f_{\text{CGG}}$ and $f_{\text{FGD}}$ are PDFs of the Compound Geometric Gamma and Folded Gamma Difference distributions, respectively.

#### S1.3 Maximum a posteriori (MAP) estimation

Rather than maximising the likelihood expression directly to obtain estimates of the model parameters, we take a Bayesian approach to incorporate prior information on parameters $\pi$ and $w$, and hence find maximum a posteriori (MAP) estimates. Given that there may be considerable uncertainty on the true transmission network as obtained from viral sequences and symptom onset times alone, but we often have good prior knowledge of the level of sampling in the population, the use of priors can help to avoid issues of identifiability in the model. (We can imagine that, without restriction of such priors, extremely short serial intervals with very low levels of sampling would provide a good fit to any data, even if we think this is impossible in practice).

We assume a beta distributed prior for both $\pi$ and $w$, desirable as it is restricted to the range [0,1].

The log prior distributions for $\pi$ and $w$ are given by:

$$q(\pi) \sim \log(\text{Beta}(\alpha_\pi, \beta_\pi))$$
$$q(w) \sim \log(\text{Beta}(\alpha_w, \beta_w)).$$

Given these prior distributions and the log-likelihood expression introduced in Equation S1, the log posterior distribution is given by:

$$p_{c,\tau}(\mu, \sigma, \pi, w|\mathcal{D}_{c,\tau}) = l_{c,\tau}(\mu, \sigma, \pi, w|\mathcal{D}_c) + q(\pi) + q(w). \tag{S2}$$

MAP estimates $(\hat{\mu}, \hat{\sigma}, \hat{\pi}, \hat{w})_{c,\tau}$, conditional on a sampled transmission network $\tau$ for cluster $c$, are then found by maximising the log posterior density in Equation S2. In practice, we numerically optimize the log posterior density using the *optim* function in *R*.

What remains is to incorporate uncertainty in the transmission network $\tau$. Rather than sampling a single transmission network $\tau$ from the transmission cloud of potential infector-infectee pairs for cluster $c$, we sample a set of networks $\tau_1, \tau_2, \ldots, \tau_N$. MAPs $(\hat{\mu}, \hat{\sigma}, \hat{\pi}, \hat{w})_{c,\tau_i}$ are obtained for each network $\tau_i$ independently. When fewer than $N$ MAP estimates are returned from the optimization, as can occur when networks are randomly sampled which are not concordant with the assumed priors and so cause the numerical optimization to fail, we sample additional networks until $N$ MAPs are obtained.

The overall MAP for each parameter in cluster $c$ is then calculated as the mean across all sampled transmission networks, for example:

$$\hat{\mu}_c = \frac{1}{N} \sum_{k=1}^{N} \hat{\mu}_{c,\tau_k},$$

and similarly for $\hat{\sigma}_c$, $\hat{\pi}_c$ and $\hat{w}_c$.

#### S1.4 Confidence Intervals

We obtain estimates of the standard error ($\hat{\text{se}}$) of MAPs $(\hat{\mu}, \hat{\sigma}, \hat{\pi}, \hat{w})_{c,\tau}$, for a sampled network $\tau$, using the inverse negative Hessian evaluated at the MAPs, as obtained from the numerical optimization procedure. At confidence level $\alpha$, this provides an approximate confidence interval for $\hat{\mu}_{c,\tau_k}$ of

$$\hat{\mu}_{c,\tau_k} \pm z_{\alpha/2}\hat{\text{se}}(\hat{\mu}_{c,\tau_k}),$$

and similarly for $\hat{\sigma}_{c,\tau_k}$, $\hat{\pi}_{c,\tau_k}$ and $\hat{w}_{c,\tau_k}$.

In order to obtain confidence intervals for the overall cluster estimates $\hat{\mu}_c, \hat{\sigma}_c, \hat{\pi}_c, \hat{w}_c$ i.e. over the space of all sampled transmission networks, we must take into account variation both within and between estimates. The variance of estimator $\hat{\mu}_c$ (and equivalently $\hat{\sigma}_c$, $\hat{\pi}_c$, and $\hat{w}_c$) is derived with the law of total variance:

$$\hat{\text{Var}}(\hat{\mu}_c) = \mathbb{E}_\tau\left(\hat{\text{se}}(\hat{\mu}_{c,\tau_k})^2\right) + \text{Var}_\tau\left(\hat{\mu}_{c,\tau_k}\right).$$

So, the first term incorporates the average uncertainty in each estimate of the MAP (for each sampled transmission network), and the second incorporates the estimate's variability between sampled transmission networks.

This induces a confidence interval for the cluster-level estimates of

$$\hat{\mu}_c \pm z_{\alpha/2}\sqrt{\hat{\text{Var}}(\hat{\mu}_c)}.$$

### S2 Additional information and results

Table S1: **Correspondence of wave 1 clusters with cluster labels in Seemann et al. [3]**

| Cluster | Seemann et al. [3] cluster |
|---|---|
| A1 | 4 |
| A2 | 70 |
| A3 | 74 |
| A4 | 73 |
| A5 | 7 |
| A6 | 9 |
| A7 | 19 |
| A8 | 24 |
| A9 | 76 |
| A10 | 67 |

Table S2: **Published estimates of the COVID-19 serial interval, in days.**

| Mean SI (95% CI) | Data, Country, Dates | Source |
|---|---|---|
| 7.5 (5.3–19) | 425 cases, Wuhan China, Dec 2019–Jan 2020 | Li et al. 2020 [4] |
| 4.0 (3.1–4.9) | 28 pairs, World, Dec 2019–Feb 2020 | Nishiura et al. 2020 [5] |
| 5.8 (4.8–6.8) | 77 pairs, China, Dec 2019–Mar 2020 | He et al. 2020 [6] |
| 6·3 (5·2–7·6) | 48 pairs, Shenzhen China, Jan–Feb 2020 | Bi et al. 2020 [7] |
| 4.9 (3.6–6.2) | 21 pairs, Hong Kong, Jan–Feb 2020 | Zhao et al. 2020 [8] |
| 4.17 (2.44–5.89) | 93 cases, Singapore, Jan–Feb 2020 | Tindale et al. [9] |
| 4.31 (2.91–5.72) | 135 cases, Tianjin China, Jan–Feb 2020 | Tindale et al. [9] |
| 5.54 (4.08–7.01) | 28 pairs, Busan Korea, Jan–Mar 2020 | Son et al. 2020 [10] |
| 4.5 (3.1–5.5) | 37 pairs, Iran, Feb–May 2020 | Zare et al. 2021 [11] |
| 4.0 (3.7–4.3) | 471 pairs, Ireland, Apr-Dec 2020 | McAloon et al. 2021 [12] |
| 3.18 (2.55–3.81) | 186 pairs (significant Alpha), UK, Sep 2020–Feb 2021 | Geismar et al. 2021 [13] |
| 2.3 (1.4–3.3) | 68 pairs (Delta), Guangdong China, May–June 2021 | Zhang et al. 2021 [14] |

Table S3: **Full results table.** Mean parameter estimates with 95% confidence intervals.

| **Wave 1 clusters** | **Serial interval mean** $\mu$ | **Serial interval SD** $\sigma$ | **Sampling rate** $\pi$ | **Proportion non-coprimary** $w$ |
|---|---|---|---|---|
| A1 | 3.39 (1.43, 5.36) | 1.93 (0.55, 3.31) | 0.57 (0.38, 0.75) | 0.55 (0.33, 0.77) |
| A2 | 5.95 (2.46, 9.45) | 2.77 (0.61, 4.93) | 0.60 (0.41, 0.78) | 0.54 (0.34, 0.74) |
| A3 | 5.54 (2.08, 9.01) | 2.55 (0.0, 5.92) | 0.58 (0.38, 0.78) | 0.56 (0.32, 0.80) |
| A4 | 4.01 (1.60, 6.42) | 2.22 (0.69, 3.76) | 0.62 (0.42, 0.82) | 0.55 (0.32, 0.78) |
| A5 | 6.60 (2.48, 10.73) | 2.39 (0.0, 4.81) | 0.61 (0.41, 0.81) | 0.53 (0.33, 0.74) |
| A6 | 4.85 (1.53, 8.16) | 3.05 (0.0, 6.11) | 0.56 (0.36, 0.76) | 0.54 (0.33, 0.76) |
| A7 | 4.64 (2.44, 6.83) | 2.61 (1.01, 4.21) | 0.63 (0.46, 0.81) | 0.59 (0.37, 0.81) |
| A8 | 3.40 (0.91, 5.90) | 1.91 (0.35, 3.46) | 0.58 (0.39, 0.77) | 0.55 (0.32, 0.77) |
| A9 | 5.03 (2.07, 7.98) | 2.69 (0.38, 4.99) | 0.59 (0.40, 0.78) | 0.56 (0.33, 0.78) |
| A10 | 3.42 (0.90, 5.94) | 1.92 (0.29, 3.56) | 0.58 (0.38, 0.78) | 0.54 (0.32, 0.76) |
| **Total** | 4.65 (1.06, 8.25) | 2.38 (0.09, 4.67) | 0.59 (0.40, 0.79) | 0.55 (0.33, 0.78) |
| **Wave 2 clusters** | **Serial interval mean** $\mu$ | **Serial interval SD** $\sigma$ | **Sampling rate** $\pi$ | **Proportion non-coprimary** $w$ |
| B1 | 4.85 (1.12, 8.58) | 2.53 (0, 5.13) | 0.57 (0.37, 0.77) | 0.54 (0.31, 0.77) |
| B2 | 4.66 (1.52, 7.8) | 2.49 (0.36, 4.62) | 0.59 (0.4, 0.79) | 0.55 (0.33, 0.78) |
| B3 | 7.67 (4.13, 11.21) | 5.44 (1.94, 8.94) | 0.58 (0.39, 0.77) | 0.56 (0.34, 0.78) |
| B4 | 5.38 (1.1, 9.67) | 2.28 (0.54, 4.02) | 0.62 (0.41, 0.83) | 0.52 (0.31, 0.74) |
| B5 | 8.31 (4.78, 11.85) | 6.52 (3.12, 9.92) | 0.6 (0.4, 0.79) | 0.56 (0.33, 0.78) |
| B6 | 6.13 (2.09, 10.17) | 3.52 (0.23, 6.81) | 0.58 (0.38, 0.78) | 0.55 (0.33, 0.77) |
| B7 | 5.8 (2.93, 8.66) | 3.3 (1.42, 5.19) | 0.62 (0.45, 0.8) | 0.57 (0.34, 0.79) |
| B8 | 5.92 (1.97, 9.87) | 3.55 (0.03, 7.08) | 0.57 (0.37, 0.77) | 0.55 (0.33, 0.78) |
| B9 | 7.29 (2.95, 11.63) | 4.56 (0.36, 8.76) | 0.57 (0.37, 0.76) | 0.55 (0.33, 0.76) |
| B10 | 5.2 (3.06, 7.34) | 3.05 (1.1, 5) | 0.62 (0.45, 0.79) | 0.6 (0.39, 0.82) |
| B11 | 5.87 (1.13, 10.6) | 4.37 (0, 11.32) | 0.52 (0.3, 0.73) | 0.53 (0.31, 0.74) |
| B12 | 4.44 (0.02, 8.86) | 2.02 (0.04, 4) | 0.57 (0.37, 0.77) | 0.52 (0.29, 0.75) |
| B13 | 3.85 (2.44, 5.25) | 2.26 (1.14, 3.38) | 0.63 (0.47, 0.79) | 0.65 (0.45, 0.86) |
| B14 | 4.07 (0, 9.17) | 1.64 (0.54, 2.73) | 0.59 (0.4, 0.78) | 0.52 (0.23, 0.82) |
| B15 | 4.78 (1.16, 8.39) | 2.73 (0, 5.99) | 0.56 (0.36, 0.76) | 0.55 (0.32, 0.78) |
| B16 | 7.66 (4.86, 10.47) | 4.82 (1.32, 8.32) | 0.62 (0.45, 0.79) | 0.58 (0.37, 0.79) |
| B17 | 5.37 (3.38, 7.36) | 2.69 (0.82, 4.56) | 0.64 (0.48, 0.8) | 0.61 (0.42, 0.8) |
| B18 | 5.02 (2.13, 7.91) | 3.02 (0.89, 5.15) | 0.57 (0.38, 0.76) | 0.54 (0.32, 0.76) |
| B19 | 6.7 (3.59, 9.8) | 3.94 (1.95, 5.92) | 0.67 (0.5, 0.83) | 0.62 (0.39, 0.85) |
| B20 | 3.71 (0.68, 6.74) | 2.02 (0.14, 3.9) | 0.56 (0.36, 0.76) | 0.54 (0.32, 0.75) |
| B21 | 3.88 (1.06, 6.7) | 2.03 (0.03, 4.03) | 0.56 (0.36, 0.76) | 0.54 (0.32, 0.76) |
| B22 | 4.13 (1.31, 6.95) | 2.3 (0.6, 4) | 0.58 (0.39, 0.76) | 0.55 (0.33, 0.77) |
| B23 | 6.84 (3.65, 10.03) | 3.63 (0.06, 7.19) | 0.59 (0.42, 0.77) | 0.58 (0.35, 0.81) |
| B24 | 5.3 (2.36, 8.24) | 2.98 (1.09, 4.87) | 0.6 (0.42, 0.79) | 0.56 (0.35, 0.78) |
| B25 | 3.45 (0.87, 6.04) | 1.9 (0.17, 3.63) | 0.58 (0.39, 0.78) | 0.55 (0.32, 0.78) |
| B26 | 6.57 (2.46, 10.68) | 4.26 (0, 8.85) | 0.55 (0.35, 0.76) | 0.55 (0.33, 0.76) |

| Wave 2 clusters | Serial interval mean $\mu$ | Serial interval SD $\sigma$ | Sampling rate $\pi$ | Proportion non-coprimary $w$ |
|---|---|---|---|---|
| B27 | 6.44 (2.25, 10.64) | 3.9 (0.53, 7.27) | 0.58 (0.38, 0.78) | 0.55 (0.32, 0.77) |
| B28 | 3.98 (0.65, 7.3) | 2.32 (0.65, 3.99) | 0.59 (0.39, 0.78) | 0.54 (0.31, 0.77) |
| B29 | 3.58 (1.5, 5.65) | 2.03 (0.59, 3.46) | 0.61 (0.42, 0.8) | 0.58 (0.35, 0.82) |
| B30 | 4.55 (2.14, 6.96) | 2.58 (0.99, 4.18) | 0.61 (0.42, 0.79) | 0.58 (0.36, 0.8) |
| B31 | 5.7 (3.17, 8.22) | 2.95 (0.97, 4.92) | 0.6 (0.43, 0.77) | 0.57 (0.37, 0.77) |
| B32 | 9.54 (0.82, 18.25) | 6.11 (0, 12.9) | 0.56 (0.35, 0.76) | 0.53 (0.31, 0.76) |
| B33 | 6.08 (3.58, 8.58) | 3.85 (1.88, 5.83) | 0.63 (0.46, 0.8) | 0.6 (0.39, 0.82) |
| B34 | 3.9 (0.66, 7.13) | 1.98 (0.55, 3.42) | 0.6 (0.41, 0.79) | 0.55 (0.3, 0.8) |
| B35 | 8.82 (1.62, 16.02) | 6.15 (0.13, 12.17) | 0.56 (0.35, 0.76) | 0.53 (0.32, 0.75) |
| B36 | 7.78 (3.63, 11.94) | 4.66 (0.99, 8.32) | 0.6 (0.4, 0.79) | 0.57 (0.34, 0.79) |
| B37 | 5.63 (2.42, 8.85) | 3.37 (0.02, 6.73) | 0.54 (0.35, 0.73) | 0.54 (0.33, 0.76) |
| B38 | 3.29 (0.49, 6.1) | 1.73 (0.38, 3.08) | 0.6 (0.4, 0.79) | 0.54 (0.3, 0.79) |
| B39 | 4.38 (1.65, 7.11) | 2.48 (0.7, 4.26) | 0.54 (0.35, 0.73) | 0.56 (0.33, 0.78) |
| B40 | 6.48 (2.5, 10.46) | 3.84 (0.54, 7.14) | 0.58 (0.39, 0.77) | 0.55 (0.33, 0.78) |
| B41 | 4.66 (1.79, 7.52) | 2.36 (0, 4.81) | 0.59 (0.4, 0.78) | 0.57 (0.33, 0.8) |
| B42 | 5.7 (2.6, 8.81) | 3.23 (0.22, 6.24) | 0.56 (0.36, 0.75) | 0.56 (0.35, 0.78) |
| B43 | 2.64 (1.22, 4.06) | 1.48 (0.6, 2.36) | 0.6 (0.42, 0.78) | 0.68 (0.44, 0.91) |
| B44 | 3.48 (0, 7.66) | 1.61 (0.62, 2.61) | 0.54 (0.34, 0.73) | 0.51 (0.22, 0.8) |
| B45 | 4.68 (1.61, 7.74) | 2.41 (0, 4.92) | 0.55 (0.36, 0.74) | 0.55 (0.33, 0.78) |
| B46 | 7 (3.66, 10.34) | 3.26 (0.09, 6.44) | 0.53 (0.35, 0.71) | 0.54 (0.35, 0.74) |
| B47 | 6.74 (4.25, 9.22) | 4.47 (1.79, 7.15) | 0.61 (0.42, 0.79) | 0.62 (0.41, 0.82) |
| B48 | 3.71 (1.57, 5.86) | 2.11 (1.06, 3.17) | 0.6 (0.43, 0.77) | 0.62 (0.37, 0.87) |
| B49 | 4.23 (2.13, 6.34) | 2.24 (0.55, 3.92) | 0.59 (0.4, 0.77) | 0.55 (0.35, 0.76) |
| B50 | 5.57 (2.6, 8.55) | 3.25 (0.4, 6.09) | 0.59 (0.4, 0.77) | 0.57 (0.35, 0.79) |
| B51 | 5.46 (2.85, 8.07) | 3.48 (0.76, 6.21) | 0.55 (0.36, 0.74) | 0.57 (0.35, 0.78) |
| B52 | 4.87 (1.5, 8.24) | 2.28 (0.74, 3.82) | 0.56 (0.37, 0.75) | 0.51 (0.29, 0.74) |
| B53 | 3.86 (1.13, 6.6) | 1.77 (0, 3.67) | 0.47 (0.28, 0.65) | 0.51 (0.29, 0.72) |
| B54 | 5.06 (2.62, 7.51) | 2.87 (0.74, 5) | 0.58 (0.39, 0.77) | 0.57 (0.35, 0.79) |
| B55 | 3.33 (1.59, 5.07) | 1.99 (0.81, 3.17) | 0.57 (0.4, 0.75) | 0.6 (0.36, 0.85) |
| B56 | 3.99 (0.24, 7.74) | 1.71 (0.29, 3.13) | 0.59 (0.41, 0.77) | 0.55 (0.29, 0.82) |
| B57 | 6.74 (3.62, 9.86) | 4.21 (0.93, 7.48) | 0.58 (0.4, 0.77) | 0.58 (0.36, 0.8) |
| B58 | 7.89 (4.18, 11.6) | 4.2 (0.85, 7.55) | 0.63 (0.45, 0.81) | 0.57 (0.35, 0.79) |
| B59 | 5.24 (1.34, 9.13) | 2.85 (0, 6.01) | 0.55 (0.35, 0.76) | 0.53 (0.32, 0.75) |
| B60 | 2.75 (0.53, 4.97) | 1.51 (0.2, 2.82) | 0.59 (0.4, 0.79) | 0.55 (0.31, 0.79) |
| B61 | 4.35 (0, 9.66) | 1.71 (0.45, 2.97) | 0.58 (0.38, 0.77) | 0.51 (0.25, 0.76) |
| B62 | 3.96 (0, 8.26) | 1.78 (0.36, 3.2) | 0.59 (0.39, 0.78) | 0.53 (0.28, 0.78) |
| B63 | 3.39 (0.4, 6.38) | 1.93 (0, 3.92) | 0.53 (0.33, 0.73) | 0.53 (0.31, 0.74) |
| B64 | 3.78 (1.31, 6.25) | 2 (0.29, 3.71) | 0.57 (0.39, 0.76) | 0.56 (0.33, 0.8) |
| B65 | 4.61 (1.56, 7.66) | 2.49 (0.62, 4.36) | 0.57 (0.38, 0.76) | 0.56 (0.33, 0.78) |
| B66 | 4.66 (0.93, 8.38) | 2.37 (0, 4.82) | 0.58 (0.38, 0.77) | 0.54 (0.31, 0.77) |
| B67 | 5.26 (2.96, 7.57) | 2.89 (1, 4.78) | 0.62 (0.44, 0.79) | 0.59 (0.36, 0.81) |
| B68 | 2.8 (0.34, 5.27) | 1.4 (0.14, 2.67) | 0.59 (0.39, 0.79) | 0.54 (0.31, 0.77) |

| Wave 2 clusters | Serial interval mean $\mu$ | Serial interval SD $\sigma$ | Sampling rate $\pi$ | Proportion non-coprimary $w$ |
|---|---|---|---|---|
| B69 | 5.36 (2.7, 8.02) | 2.88 (0.54, 5.22) | 0.58 (0.4, 0.76) | 0.57 (0.35, 0.78) |
| B70 | 4.56 (2.25, 6.87) | 2.56 (0.85, 4.26) | 0.63 (0.45, 0.81) | 0.6 (0.37, 0.84) |
| B71 | 4.04 (0.8, 7.27) | 1.9 (0.55, 3.25) | 0.46 (0.26, 0.65) | 0.49 (0.27, 0.71) |
| B72 | 4.39 (1.29, 7.5) | 2.31 (0.31, 4.32) | 0.53 (0.33, 0.72) | 0.54 (0.32, 0.76) |
| B73 | 1.97 (0, 3.95) | 0.92 (0.03, 1.8) | 0.59 (0.4, 0.78) | 0.56 (0.31, 0.81) |
| B74 | 9.33 (3.17, 15.48) | 6.2 (0.59, 11.82) | 0.57 (0.37, 0.78) | 0.55 (0.32, 0.77) |
| B75 | 3.76 (1.56, 5.96) | 2.01 (0.51, 3.51) | 0.57 (0.38, 0.76) | 0.56 (0.33, 0.78) |
| B76 | 3.73 (0, 7.81) | 1.69 (0.22, 3.17) | 0.55 (0.36, 0.75) | 0.53 (0.29, 0.76) |
| B77 | 7 (2.16, 11.84) | 5.33 (0, 11.21) | 0.54 (0.33, 0.75) | 0.54 (0.32, 0.75) |
| B78 | 5.8 (2.29, 9.31) | 2.86 (0.46, 5.26) | 0.56 (0.37, 0.75) | 0.53 (0.32, 0.74) |
| B79 | 4.12 (1.32, 6.91) | 2.25 (0.49, 4.01) | 0.58 (0.38, 0.78) | 0.54 (0.32, 0.76) |
| B80 | 6.91 (3.15, 10.66) | 3.78 (0.43, 7.13) | 0.58 (0.39, 0.77) | 0.55 (0.34, 0.77) |
| B81 | 6.66 (1.94, 11.37) | 4.72 (0, 10.71) | 0.54 (0.33, 0.74) | 0.54 (0.32, 0.76) |
| B82 | 2.22 (1.26, 3.19) | 1.32 (0.42, 2.23) | 0.5 (0.33, 0.67) | 0.56 (0.32, 0.8) |
| B83 | 6.79 (3.23, 10.35) | 4.44 (1.14, 7.73) | 0.59 (0.39, 0.78) | 0.55 (0.33, 0.76) |
| B84 | 4.42 (1.22, 7.62) | 2.41 (0, 4.85) | 0.58 (0.38, 0.78) | 0.54 (0.32, 0.77) |
| B85 | 5.91 (2.49, 9.34) | 2.86 (0.24, 5.49) | 0.6 (0.41, 0.8) | 0.56 (0.33, 0.78) |
| B86 | 4.31 (0, 9.63) | 1.46 (0.35, 2.56) | 0.57 (0.38, 0.77) | 0.49 (0.23, 0.75) |
| B87 | 4.99 (1.85, 8.12) | 2.78 (0.12, 5.44) | 0.58 (0.38, 0.77) | 0.55 (0.33, 0.78) |
| B88 | 3.82 (0.23, 7.41) | 1.9 (0.3, 3.49) | 0.58 (0.38, 0.77) | 0.54 (0.3, 0.78) |
| B89 | 4.4 (2.44, 6.37) | 2.46 (0.68, 4.24) | 0.6 (0.42, 0.77) | 0.6 (0.38, 0.83) |
| B90 | 3.74 (1.79, 5.69) | 2.11 (0.66, 3.57) | 0.62 (0.44, 0.8) | 0.6 (0.37, 0.83) |
| B91 | 5.75 (0.23, 11.28) | 1.95 (0.43, 3.48) | 0.59 (0.39, 0.8) | 0.48 (0.24, 0.72) |
| B92 | 4.18 (1.5, 6.86) | 2.22 (0.13, 4.3) | 0.6 (0.4, 0.79) | 0.56 (0.33, 0.78) |
| B93 | 3.94 (1.68, 6.2) | 2.12 (0.38, 3.86) | 0.6 (0.41, 0.78) | 0.57 (0.34, 0.8) |
| B94 | 2.96 (0.17, 5.74) | 1.5 (0.23, 2.78) | 0.6 (0.41, 0.79) | 0.55 (0.31, 0.8) |
| **Total** | 5.17 (0.47, 9.87) | 2.95 (0, 6.67) | 0.58 (0.38, 0.78) | 0.56 (0.32, 0.79) |

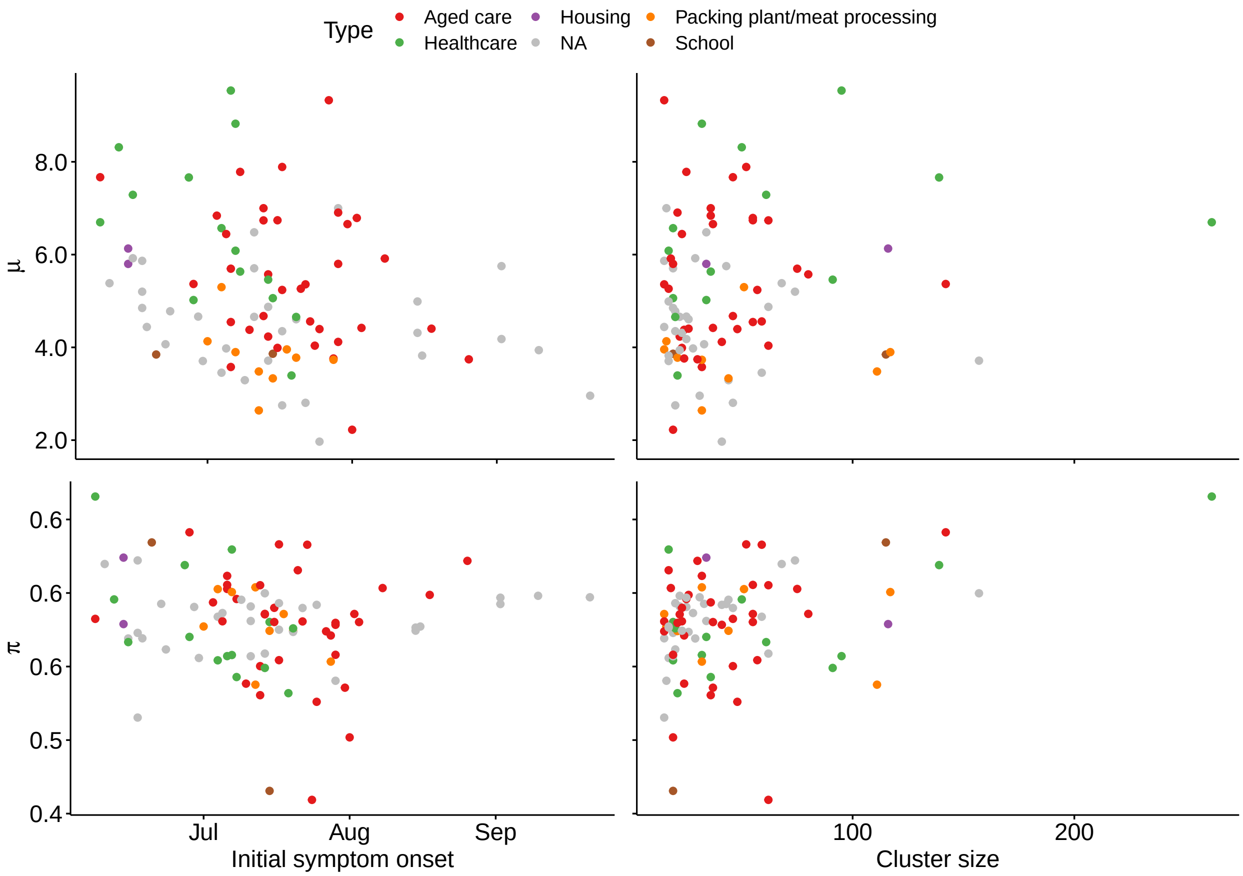


Figure S1: **Relationship between estimates of the mean serial interval ($\mu$) or sampling rate ($\pi$) and the initial symptom onset date or size of each cluster, for wave 2 data.** Colours show exposure site types.

### S3 Sensitivity to prior distribution

In this section, we explore the model sensitivity to the assumed prior distributions for sampling rate $\pi$ and proportion non-coprimary $w$. In the main text, we use a $\text{Beta}(12, 11)$ prior distribution for both parameters. Here, we repeat the analysis of wave 1 clusters under 4 different prior distribution scenarios: increased/decreased mean and increased/decreased standard deviation. These prior distribution scenarios are plotted in Figure S2. For each scenario we sample 100 transmission networks in each of the 10 wave 1 clusters, as before.

Results of the sensitivity analysis are shown in Figure S3. We find that the estimates of serial interval mean $\mu$ and standard deviation $\sigma$ are relatively robust to changes in the sampling priors (that is, the prior distributions for parameters $\pi$ and $w$). The relative ordering of clusters is largely preserved. The estimates of $\pi$ and $w$ are changed somewhat in line with their prior. These results suggest that the data are sufficiently informative of the underlying serial interval distribution, so long as we have some minimal understanding of the case ascertainment rate.

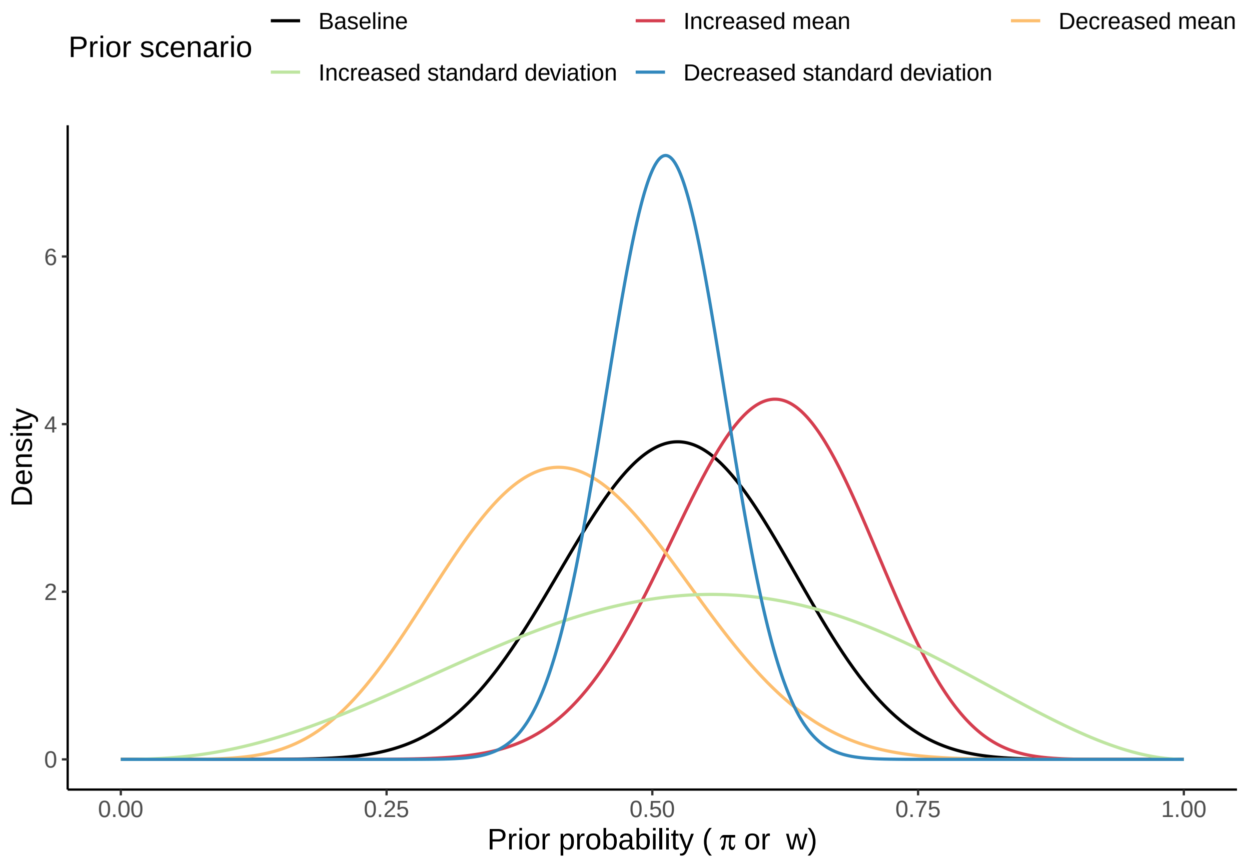


Figure S2: **Four scenarios for exploring sensitivity to prior distribution of $\pi$ and $w$, compared to baseline Beta(12,11) prior from main analysis.** Increased mean: Beta(17,11), decreased mean: Beta(8,11), increased standard deviation: Beta(3.5,3), decreased standard deviation: Beta(42,40).

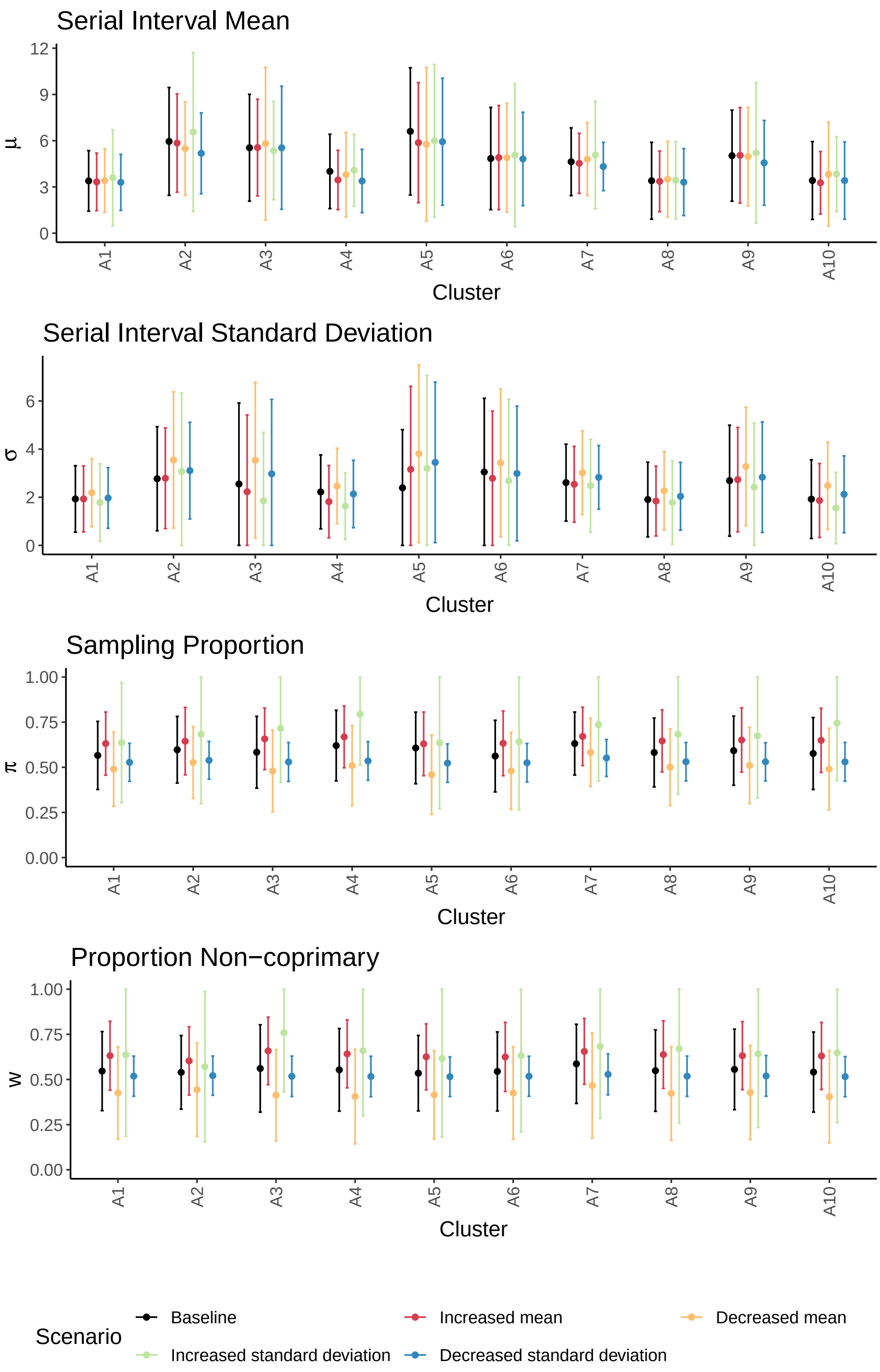


Figure S3: **Results of sensitivity analysis.** Estimates of model parameters $(\mu, \sigma, \pi, w)$ under 4 alternate prior distributions for $\pi$ and $w$, compared to baseline Beta(12,11) prior from main analysis.

### S4 Validation: epidemiologically-defined clusters

To validate our genomic approach, we apply the same estimation procedure defined in Section S1 to epidemiologically-defined clusters and transmission networks. These epidemiological clusters are defined as the connected components of a contact network created from contact tracing data, where an edge in the contact network indicates either a known direct contact or a shared exposure site. We again exclude any clusters with fewer than 15 cases. We focus on only the wave 1 data for this analysis, as the contact tracing data showed higher resolution with less uncertainty. The resulting clusters are shown in Figure S4.

We then define a transmission cloud as the set of all plausible transmission pairs, again using contact data in place of viral sequences. Here, a plausible transmission pair is two cases such that the time between symptom onset dates of the putative infector and infectee pair is positive and less than 35 days (as before, but with no genomic element). As before, this means that each infectee may have more than one plausible infector. Transmission networks are sampled from the transmission cloud by preferentially sampling from edges which represent confirmed contacts (as opposed to shared exposure locations). We sample 100 transmission networks per cluster. We estimate the serial interval in each cluster, using a beta-distributed prior for $\pi$ and $w$. We use a Beta$(9, 2.5)$ prior distribution (mean 0.78) to reflect that a larger proportion of Victorian COVID-19 cases were contact traced than genomically sequenced.

The cluster-specific estimates for the serial interval parameters are shown in Figures S5, with the resulting distributions shown in S6. Figure S7 shows the comparison of these results with the published estimates shown in table S2.

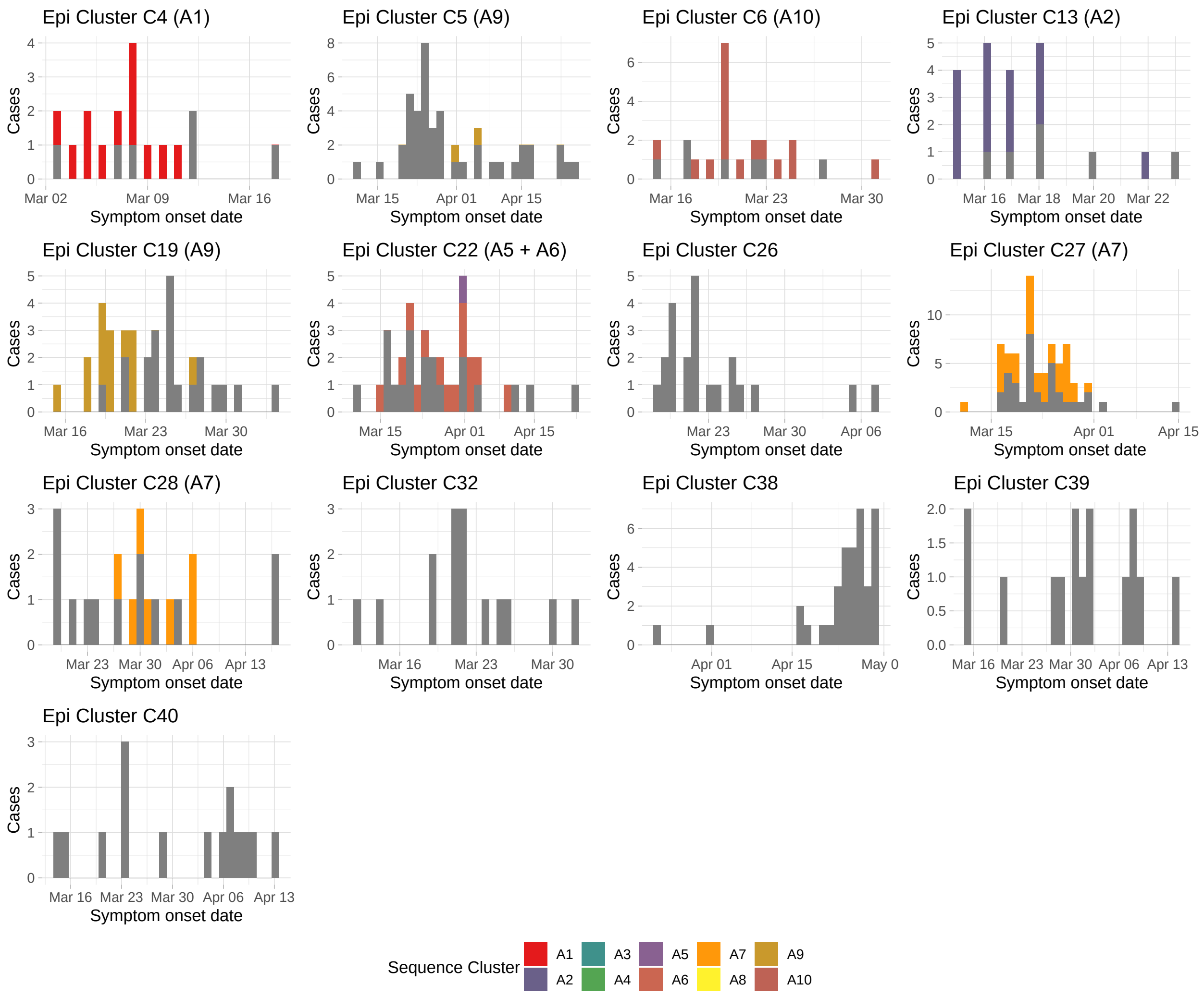


Figure S4: **Summary of epidemiologically defined clusters for wave 1 data**. Cases plotted by reported symptom onset date, for all identified epidemiological clusters with at least 15 cases. Cases are coloured by their cluster membership in the genomically defined clusters A1–A10 from the main text, with grey indicating a case was not included in any of A1–A10. Brackets in plot titles also indicate which epidemiological clusters have cases from which genomic clusters.

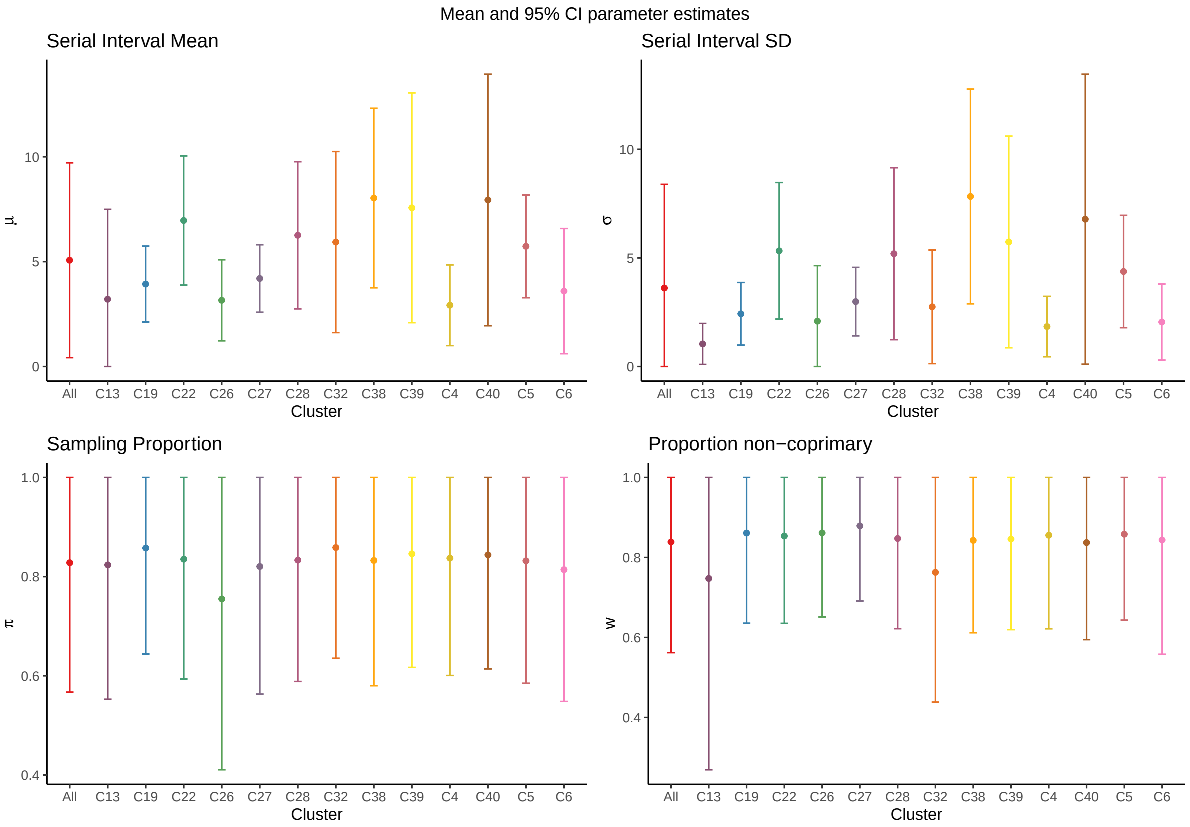


Figure S5: **Estimates of model parameters per epidemiologically defined cluster.** Mean estimates shown as points and 95% confidence intervals as bars.

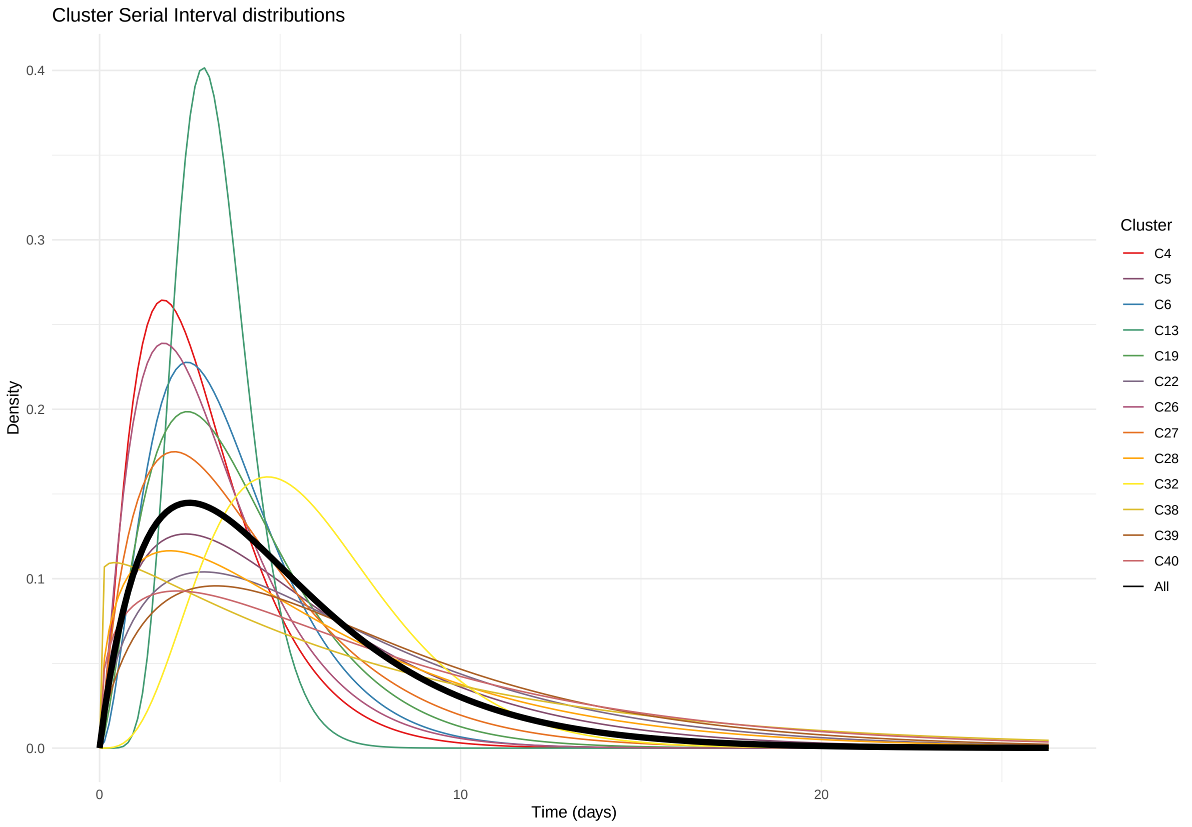


Figure S6: **Estimate of serial interval distribution per epidemiologically defined cluster.** Black (bold) curve indicates the pooled estimate across all clusters in the analysis.

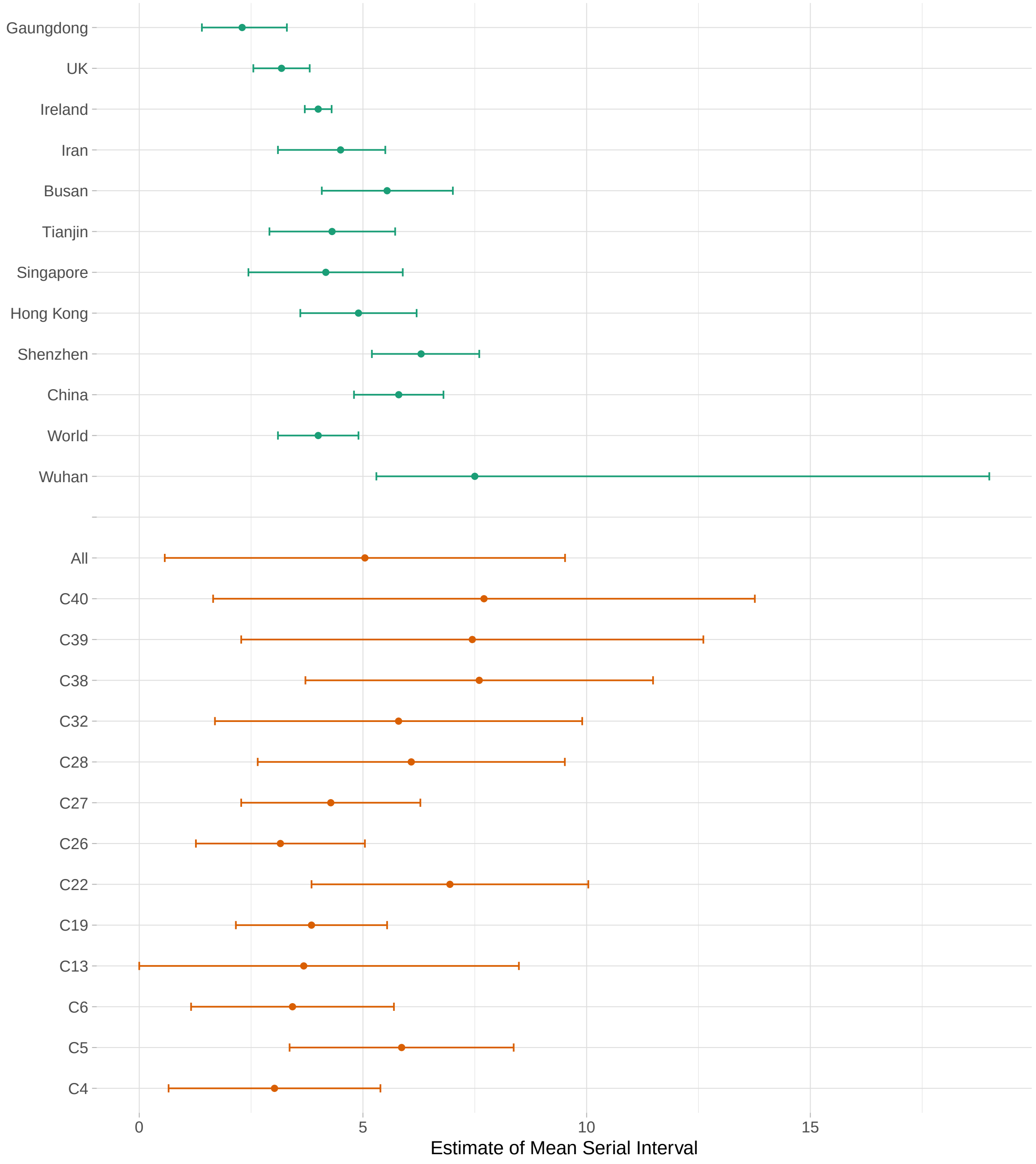


Figure S7: **Comparison of the mean serial interval for the epidemiologically defined clusters shown in Figure S4, with the published results listed in Table S2**. Each point indicates the estimate for the mean serial interval, with bars indicating the 95% confidence intervals. The first 12 rows (green) show the previously published estimates, while the remaining 14 rows (orange) show the results from this analysis.

[14] Meng Zhang, Jianpeng Xiao, Aiping Deng, Yingtao Zhang, Yali Zhuang, Ting Hu, Jiansen Li, Hongwei Tu, Bosheng Li, Yan Zhou, et al. Transmission Dynamics of an Outbreak of the

COVID-19 Delta Variant B.1.617.2 — Guangdong Province, China, May–June 2021. *China CDC Weekly*, 3(27):584–586, 2021.
