## Supplementary material for "Genomic epidemiology offers high resolution estimates of serial intervals for COVID-19": GISAID accession numbers and laboratory acknowledgements: gisaid_hcov-19_acknowledgement_table_Stockdaleetal.pdf

We gratefully acknowledge the following Authors from the Originating laboratories responsible for obtaining the specimens, as well as the Submitting laboratories where the genome data were generated and shared via GISAID, on which this research is based.

All Submitters of data may be contacted directly via www.gisaid.org

Authors are sorted alphabetically.

Acknowledgement EPI_SET Identifier: EPI_SET_20220223vy

| Accession ID | Originating Laboratory | Submitting Laboratory | Authors |
|---|---|---|---|
| EPI_ISL_594135, EPI_ISL_594136, EPI_ISL_594137, EPI_ISL_594138, EPI_ISL_594140, EPI_ISL_594141, EPI_ISL_594144, EPI_ISL_594145, EPI_ISL_594146, EPI_ISL_594147, EPI_ISL_603040, EPI_ISL_603041, EPI_ISL_603043, EPI_ISL_603044, EPI_ISL_603046, EPI_ISL_603047, EPI_ISL_603048 | | | |
| see above | MDU-PHL, The Peter Doherty Institute for Infection and Immunity | MDU-PHL, The Peter Doherty Institute for Infection and Immunity | Caly, L.; Druce, J.; M.B.; M.L.; N.L.; Sait; Sait, M.; Schultz; Seemann, T.; Sherry; Sherry, N. |

EPI_ISL_480558, EPI_ISL_480618, EPI_ISL_480747, EPI_ISL_480748, EPI_ISL_480749, EPI_ISL_480751, EPI_ISL_480752, EPI_ISL_480753, EPI_ISL_480754, EPI_ISL_480755, EPI_ISL_480756, EPI_ISL_480757, EPI_ISL_480758, EPI_ISL_480759, EPI_ISL_480760, EPI_ISL_480761, EPI_ISL_480762, EPI_ISL_480763, EPI_ISL_480767, EPI_ISL_480768, EPI_ISL_480769, EPI_ISL_480770, EPI_ISL_480771, EPI_ISL_480772,
EPI_ISL_480773, EPI_ISL_480774, EPI_ISL_480775, EPI_ISL_480777, EPI_ISL_518064, EPI_ISL_518065, EPI_ISL_518066, EPI_ISL_518067, EPI_ISL_518068, EPI_ISL_518069, EPI_ISL_518070, EPI_ISL_518071, EPI_ISL_518072, EPI_ISL_518073, EPI_ISL_518074, EPI_ISL_518075, EPI_ISL_518076, EPI_ISL_518077, EPI_ISL_518078, EPI_ISL_518079, EPI_ISL_518080, EPI_ISL_518093, EPI_ISL_518094, EPI_ISL_518096,
EPI_ISL_518097, EPI_ISL_518231, EPI_ISL_518232, EPI_ISL_518234, EPI_ISL_518235, EPI_ISL_518236, EPI_ISL_518241, EPI_ISL_518242, EPI_ISL_518243, EPI_ISL_518250, EPI_ISL_518251, EPI_ISL_518252, EPI_ISL_518254, EPI_ISL_518255, EPI_ISL_518256, EPI_ISL_518257, EPI_ISL_518258, EPI_ISL_518259, EPI_ISL_518261, EPI_ISL_518262, EPI_ISL_518264, EPI_ISL_518265, EPI_ISL_518266, EPI_ISL_518267,
EPI_ISL_518268, EPI_ISL_518269, EPI_ISL_518270, EPI_ISL_518271, EPI_ISL_518272, EPI_ISL_518273, EPI_ISL_518274, EPI_ISL_518275, EPI_ISL_518276, EPI_ISL_518277, EPI_ISL_518278, EPI_ISL_518279, EPI_ISL_518280, EPI_ISL_518282, EPI_ISL_518283, EPI_ISL_518284, EPI_ISL_518285, EPI_ISL_518286, EPI_ISL_518287, EPI_ISL_518288, EPI_ISL_518289, EPI_ISL_518290, EPI_ISL_518292, EPI_ISL_518293,
EPI_ISL_518294, EPI_ISL_518295, EPI_ISL_518296, EPI_ISL_518297, EPI_ISL_518298, EPI_ISL_518299, EPI_ISL_518300, EPI_ISL_518301, EPI_ISL_518302, EPI_ISL_518303, EPI_ISL_518305, EPI_ISL_518306, EPI_ISL_518307, EPI_ISL_518308, EPI_ISL_518309, EPI_ISL_518310, EPI_ISL_518311, EPI_ISL_518312, EPI_ISL_518313, EPI_ISL_518314, EPI_ISL_518315, EPI_ISL_518316, EPI_ISL_518319, EPI_ISL_518321,
EPI_ISL_518322, EPI_ISL_518323, EPI_ISL_518324, EPI_ISL_518325, EPI_ISL_518326, EPI_ISL_518327, EPI_ISL_518328, EPI_ISL_518329, EPI_ISL_518330, EPI_ISL_518331, EPI_ISL_518333, EPI_ISL_518334, EPI_ISL_518335, EPI_ISL_518336, EPI_ISL_518337, EPI_ISL_518339, EPI_ISL_518340, EPI_ISL_518342, EPI_ISL_518343, EPI_ISL_518344, EPI_ISL_518345, EPI_ISL_518346, EPI_ISL_518347, EPI_ISL_518349,
EPI_ISL_518350, EPI_ISL_518351, EPI_ISL_518353, EPI_ISL_518354, EPI_ISL_518355, EPI_ISL_518356, EPI_ISL_518357, EPI_ISL_518358, EPI_ISL_518359, EPI_ISL_518360, EPI_ISL_518361, EPI_ISL_518362, EPI_ISL_518363, EPI_ISL_518364, EPI_ISL_518365, EPI_ISL_518366, EPI_ISL_518367, EPI_ISL_518368, EPI_ISL_518369, EPI_ISL_518370, EPI_ISL_518371, EPI_ISL_518372, EPI_ISL_518373, EPI_ISL_518374,
EPI_ISL_518375, EPI_ISL_518376, EPI_ISL_518377, EPI_ISL_518378, EPI_ISL_518379, EPI_ISL_518380, EPI_ISL_518381, EPI_ISL_518382, EPI_ISL_518383, EPI_ISL_518384, EPI_ISL_518385, EPI_ISL_518386, EPI_ISL_518402, EPI_ISL_518403, EPI_ISL_518404, EPI_ISL_518405, EPI_ISL_518406, EPI_ISL_518407, EPI_ISL_518408, EPI_ISL_518410, EPI_ISL_518411, EPI_ISL_518412, EPI_ISL_518414, EPI_ISL_518415,
EPI_ISL_518416, EPI_ISL_518417, EPI_ISL_518418, EPI_ISL_518420, EPI_ISL_518421, EPI_ISL_518422, EPI_ISL_518473, EPI_ISL_518474, EPI_ISL_518475, EPI_ISL_518477, EPI_ISL_518479, EPI_ISL_518480, EPI_ISL_518481, EPI_ISL_518482, EPI_ISL_518483, EPI_ISL_518484, EPI_ISL_518486, EPI_ISL_518487, EPI_ISL_518488, EPI_ISL_518489, EPI_ISL_518490, EPI_ISL_518491, EPI_ISL_518492, EPI_ISL_518493,
EPI_ISL_518494, EPI_ISL_518495, EPI_ISL_518496, EPI_ISL_518497, EPI_ISL_518498, EPI_ISL_518499, EPI_ISL_518500, EPI_ISL_518501, EPI_ISL_518502, EPI_ISL_518503, EPI_ISL_518504, EPI_ISL_518505, EPI_ISL_518506, EPI_ISL_518507, EPI_ISL_518508, EPI_ISL_518509, EPI_ISL_518510, EPI_ISL_518511, EPI_ISL_518512, EPI_ISL_518513, EPI_ISL_518514, EPI_ISL_518515, EPI_ISL_518516, EPI_ISL_518517,
EPI_ISL_518518, EPI_ISL_518519, EPI_ISL_518520, EPI_ISL_518522, EPI_ISL_518523, EPI_ISL_518524, EPI_ISL_518525, EPI_ISL_518526, EPI_ISL_518527, EPI_ISL_518528, EPI_ISL_518529, EPI_ISL_518530, EPI_ISL_518531, EPI_ISL_518532, EPI_ISL_518533, EPI_ISL_518534, EPI_ISL_518535, EPI_ISL_518536, EPI_ISL_518537, EPI_ISL_518538, EPI_ISL_518539, EPI_ISL_518540, EPI_ISL_518541, EPI_ISL_518542,
EPI_ISL_518543, EPI_ISL_518545, EPI_ISL_518546, EPI_ISL_518547, EPI_ISL_518548, EPI_ISL_518549, EPI_ISL_518550, EPI_ISL_518551, EPI_ISL_518552, EPI_ISL_518553, EPI_ISL_518554, EPI_ISL_518555, EPI_ISL_518556, EPI_ISL_518557, EPI_ISL_518558, EPI_ISL_518559, EPI_ISL_518560, EPI_ISL_518561, EPI_ISL_518562, EPI_ISL_518563, EPI_ISL_518564, EPI_ISL_518565, EPI_ISL_518566, EPI_ISL_518567,
EPI_ISL_518568, EPI_ISL_518569, EPI_ISL_518571, EPI_ISL_518572, EPI_ISL_518573, EPI_ISL_518574, EPI_ISL_518576, EPI_ISL_518577, EPI_ISL_518578, EPI_ISL_518579, EPI_ISL_518580, EPI_ISL_518581, EPI_ISL_518582, EPI_ISL_518584, EPI_ISL_518585, EPI_ISL_518586, EPI_ISL_518587, EPI_ISL_518588, EPI_ISL_518589, EPI_ISL_518590, EPI_ISL_518591, EPI_ISL_518592, EPI_ISL_518594, EPI_ISL_518595,
EPI_ISL_518596, EPI_ISL_518597, EPI_ISL_518599, EPI_ISL_518600, EPI_ISL_518601, EPI_ISL_518602, EPI_ISL_518603, EPI_ISL_518604, EPI_ISL_518606, EPI_ISL_518609, EPI_ISL_518610, EPI_ISL_518611, EPI_ISL_518612, EPI_ISL_518613, EPI_ISL_518614, EPI_ISL_518615, EPI_ISL_518616, EPI_ISL_518617, EPI_ISL_518618, EPI_ISL_518619, EPI_ISL_518620, EPI_ISL_518621, EPI_ISL_518622, EPI_ISL_518623,
EPI_ISL_518624, EPI_ISL_518625, EPI_ISL_518626, EPI_ISL_518628, EPI_ISL_518629, EPI_ISL_518630, EPI_ISL_518631, EPI_ISL_518632, EPI_ISL_518633, EPI_ISL_518634, EPI_ISL_518635, EPI_ISL_518637, EPI_ISL_518638, EPI_ISL_518639, EPI_ISL_518640, EPI_ISL_518641, EPI_ISL_518642, EPI_ISL_518643, EPI_ISL_518644, EPI_ISL_518645, EPI_ISL_518646, EPI_ISL_518647, EPI_ISL_518648, EPI_ISL_518649,
EPI_ISL_518650, EPI_ISL_518652, EPI_ISL_518653, EPI_ISL_518654, EPI_ISL_518655, EPI_ISL_518656, EPI_ISL_518657, EPI_ISL_518658, EPI_ISL_518659, EPI_ISL_518662, EPI_ISL_518663, EPI_ISL_518664, EPI_ISL_518665, EPI_ISL_518666, EPI_ISL_518667, EPI_ISL_518668, EPI_ISL_518669, EPI_ISL_518670, EPI_ISL_518671, EPI_ISL_518672, EPI_ISL_518673, EPI_ISL_518674, EPI_ISL_518675, EPI_ISL_518676,
EPI_ISL_518677, EPI_ISL_518678, EPI_ISL_518680, EPI_ISL_518681, EPI_ISL_518682, EPI_ISL_518683, EPI_ISL_518684, EPI_ISL_518685, EPI_ISL_518686, EPI_ISL_518687, EPI_ISL_518688, EPI_ISL_518689, EPI_ISL_518690, EPI_ISL_518691, EPI_ISL_518692, EPI_ISL_518693, EPI_ISL_518694, EPI_ISL_518695, EPI_ISL_518696, EPI_ISL_518697, EPI_ISL_518698, EPI_ISL_518699, EPI_ISL_518701, EPI_ISL_518702,
EPI_ISL_518703, EPI_ISL_518704, EPI_ISL_518705, EPI_ISL_518706, EPI_ISL_518707, EPI_ISL_518708, EPI_ISL_518709, EPI_ISL_518710, EPI_ISL_518711, EPI_ISL_518712, EPI_ISL_518713, EPI_ISL_518714, EPI_ISL_518716, EPI_ISL_518717, EPI_ISL_518719, EPI_ISL_518720, EPI_ISL_518721, EPI_ISL_518722, EPI_ISL_518724, EPI_ISL_518725, EPI_ISL_518726, EPI_ISL_518727, EPI_ISL_518728,
EPI_ISL_518729, EPI_ISL_518730, EPI_ISL_518731, EPI_ISL_518732, EPI_ISL_518733, EPI_ISL_518734, EPI_ISL_518735, EPI_ISL_518736, EPI_ISL_518737, EPI_ISL_518738, EPI_ISL_518739, EPI_ISL_518740, EPI_ISL_518741, EPI_ISL_518742, EPI_ISL_518743, EPI_ISL_518744, EPI_ISL_518745, EPI_ISL_518746, EPI_ISL_518747, EPI_ISL_518748, EPI_ISL_518749, EPI_ISL_518750, EPI_ISL_518751, EPI_ISL_518752,
EPI_ISL_518753, EPI_ISL_518754, EPI_ISL_518755, EPI_ISL_518756, EPI_ISL_518757, EPI_ISL_518758, EPI_ISL_518759, EPI_ISL_518760, EPI_ISL_518761, EPI_ISL_518762, EPI_ISL_518763, EPI_ISL_518764, EPI_ISL_518765, EPI_ISL_518766, EPI_ISL_518767, EPI_ISL_518768, EPI_ISL_518769, EPI_ISL_518770, EPI_ISL_518771, EPI_ISL_518773, EPI_ISL_518774, EPI_ISL_518775, EPI_ISL_518776, EPI_ISL_518777,
EPI_ISL_518778, EPI_ISL_518779, EPI_ISL_518780, EPI_ISL_518781, EPI_ISL_518782, EPI_ISL_518783, EPI_ISL_518784, EPI_ISL_518785, EPI_ISL_518786, EPI_ISL_518787, EPI_ISL_518788, EPI_ISL_518789, EPI_ISL_518790, EPI_ISL_518791, EPI_ISL_518792, EPI_ISL_518794, EPI_ISL_518795, EPI_ISL_518796, EPI_ISL_518901, EPI_ISL_518902, EPI_ISL_518903, EPI_ISL_518906, EPI_ISL_518907, EPI_ISL_518908,
EPI_ISL_518909, EPI_ISL_518910, EPI_ISL_518911, EPI_ISL_518912, EPI_ISL_518913, EPI_ISL_518914, EPI_ISL_518917, EPI_ISL_518918, EPI_ISL_518919, EPI_ISL_518920, EPI_ISL_518921, EPI_ISL_518922, EPI_ISL_518923, EPI_ISL_518924, EPI_ISL_518925, EPI_ISL_518926, EPI_ISL_518927, EPI_ISL_518928, EPI_ISL_518929, EPI_ISL_518930, EPI_ISL_518931, EPI_ISL_518932, EPI_ISL_518933, EPI_ISL_518934,
EPI_ISL_518936, EPI_ISL_518937, EPI_ISL_518938, EPI_ISL_518939, EPI_ISL_518940, EPI_ISL_518941, EPI_ISL_518943, EPI_ISL_518944, EPI_ISL_518945, EPI_ISL_518946, EPI_ISL_518947, EPI_ISL_518948, EPI_ISL_518949, EPI_ISL_518950, EPI_ISL_518951, EPI_ISL_518952, EPI_ISL_518953, EPI_ISL_518955, EPI_ISL_518956, EPI_ISL_518957, EPI_ISL_518958, EPI_ISL_518959, EPI_ISL_518960, EPI_ISL_518961,
EPI_ISL_518962, EPI_ISL_518963, EPI_ISL_518964, EPI_ISL_518965, EPI_ISL_518967, EPI_ISL_518968, EPI_ISL_518969, EPI_ISL_518970, EPI_ISL_518971, EPI_ISL_518972, EPI_ISL_518973, EPI_ISL_518975, EPI_ISL_518977, EPI_ISL_518978, EPI_ISL_518979, EPI_ISL_518980, EPI_ISL_519052, EPI_ISL_519053, EPI_ISL_519054, EPI_ISL_519055, EPI_ISL_519056, EPI_ISL_519057, EPI_ISL_519058, EPI_ISL_519059,
EPI_ISL_519060, EPI_ISL_519061, EPI_ISL_519062, EPI_ISL_519063, EPI_ISL_519064, EPI_ISL_519065, EPI_ISL_519066, EPI_ISL_519067, EPI_ISL_519068, EPI_ISL_519069, EPI_ISL_519070, EPI_ISL_519071, EPI_ISL_519072, EPI_ISL_519073, EPI_ISL_519074, EPI_ISL_519075, EPI_ISL_519076, EPI_ISL_519077, EPI_ISL_519079, EPI_ISL_519082, EPI_ISL_519083, EPI_ISL_519084, EPI_ISL_519085, EPI_ISL_519086,
EPI_ISL_519087, EPI_ISL_519088, EPI_ISL_519089, EPI_ISL_519090, EPI_ISL_519092, EPI_ISL_519093, EPI_ISL_519094, EPI_ISL_519095, EPI_ISL_519096, EPI_ISL_519097, EPI_ISL_519098, EPI_ISL_519099, EPI_ISL_519100, EPI_ISL_519101, EPI_ISL_519102, EPI_ISL_519103, EPI_ISL_519104, EPI_ISL_519105, EPI_ISL_519106, EPI_ISL_519107, EPI_ISL_519108, EPI_ISL_519109, EPI_ISL_519110, EPI_ISL_519111,
EPI_ISL_519112, EPI_ISL_519113, EPI_ISL_519114, EPI_ISL_519115, EPI_ISL_519116, EPI_ISL_519117, EPI_ISL_519118, EPI_ISL_519119, EPI_ISL_519120, EPI_ISL_519121, EPI_ISL_519122, EPI_ISL_519123, EPI_ISL_519124, EPI_ISL_519125, EPI_ISL_519126, EPI_ISL_519127, EPI_ISL_519128, EPI_ISL_519129, EPI_ISL_519130, EPI_ISL_519131, EPI_ISL_519132, EPI_ISL_519133, EPI_ISL_519134, EPI_ISL_519135,
EPI_ISL_519136, EPI_ISL_519137, EPI_ISL_519138, EPI_ISL_519139, EPI_ISL_519140, EPI_ISL_519141, EPI_ISL_519142, EPI_ISL_519143, EPI_ISL_519144, EPI_ISL_519145, EPI_ISL_519146, EPI_ISL_519147, EPI_ISL_519148, EPI_ISL_519149, EPI_ISL_519150, EPI_ISL_519151, EPI_ISL_519152, EPI_ISL_519153, EPI_ISL_519154, EPI_ISL_519155, EPI_ISL_519156, EPI_ISL_519157, EPI_ISL_519158, EPI_ISL_519159,
EPI_ISL_519160, EPI_ISL_519161, EPI_ISL_519162, EPI_ISL_519163, EPI_ISL_519164, EPI_ISL_519165, EPI_ISL_519166, EPI_ISL_519167, EPI_ISL_519168, EPI_ISL_519169, EPI_ISL_519170, EPI_ISL_519171, EPI_ISL_519172, EPI_ISL_519173, EPI_ISL_519174, EPI_ISL_519175, EPI_ISL_519176, EPI_ISL_519177, EPI_ISL_519178, EPI_ISL_519179, EPI_ISL_519180, EPI_ISL_519181, EPI_ISL_519182, EPI_ISL_519183,
EPI_ISL_519184, EPI_ISL_519185, EPI_ISL_519187, EPI_ISL_519188, EPI_ISL_519189, EPI_ISL_519190, EPI_ISL_519191, EPI_ISL_519193, EPI_ISL_519194, EPI_ISL_519195, EPI_ISL_519196, EPI_ISL_519197, EPI_ISL_519198, EPI_ISL_519199, EPI_ISL_519200, EPI_ISL_519201, EPI_ISL_519202, EPI_ISL_519203, EPI_ISL_519204, EPI_ISL_519205, EPI_ISL_519206, EPI_ISL_519207, EPI_ISL_519208,
EPI_ISL_519209, EPI_ISL_519210, EPI_ISL_519211, EPI_ISL_519213, EPI_ISL_519214, EPI_ISL_519215, EPI_ISL_519216, EPI_ISL_519217, EPI_ISL_519218, EPI_ISL_519219, EPI_ISL_519220, EPI_ISL_519221, EPI_ISL_519222, EPI_ISL_519224, EPI_ISL_519225, EPI_ISL_519226, EPI_ISL_519228, EPI_ISL_519229, EPI_ISL_519231, EPI_ISL_519232, EPI_ISL_519233, EPI_ISL_519234, EPI_ISL_519235, EPI_ISL_519236,
EPI_ISL_519237, EPI_ISL_519238, EPI_ISL_519239, EPI_ISL_519240, EPI_ISL_519241, EPI_ISL_519243, EPI_ISL_519244, EPI_ISL_519245, EPI_ISL_519246, EPI_ISL_519247, EPI_ISL_519250, EPI_ISL_519251, EPI_ISL_519252, EPI_ISL_519253, EPI_ISL_519254, EPI_ISL_519255, EPI_ISL_519330, EPI_ISL_519333, EPI_ISL_519334, EPI_ISL_519335, EPI_ISL_519336, EPI_ISL_519337, EPI_ISL_519338,
EPI_ISL_519339, EPI_ISL_519340, EPI_ISL_519341, EPI_ISL_519342, EPI_ISL_519343, EPI_ISL_519344, EPI_ISL_519345, EPI_ISL_519346, EPI_ISL_519347, EPI_ISL_519348, EPI_ISL_519349, EPI_ISL_519350, EPI_ISL_519352, EPI_ISL_519353, EPI_ISL_519354, EPI_ISL_519355, EPI_ISL_519357, EPI_ISL_519358, EPI_ISL_519359, EPI_ISL_519360, EPI_ISL_519361, EPI_ISL_519362, EPI_ISL_519363, EPI_ISL_519364,
EPI_ISL_519365, EPI_ISL_519366, EPI_ISL_519367, EPI_ISL_519368, EPI_ISL_519369, EPI_ISL_519371, EPI_ISL_519372, EPI_ISL_519374, EPI_ISL_519375, EPI_ISL_519376, EPI_ISL_519377, EPI_ISL_519378, EPI_ISL_519379, EPI_ISL_519380, EPI_ISL_519381, EPI_ISL_519382, EPI_ISL_519383, EPI_ISL_519384, EPI_ISL_519386, EPI_ISL_519387, EPI_ISL_519389, EPI_ISL_519390, EPI_ISL_519392, EPI_ISL_519393,
EPI_ISL_519394, EPI_ISL_519395, EPI_ISL_519396, EPI_ISL_519397, EPI_ISL_519398, EPI_ISL_519399, EPI_ISL_519400, EPI_ISL_519401, EPI_ISL_519402, EPI_ISL_519403, EPI_ISL_519405, EPI_ISL_519406, EPI_ISL_519408, EPI_ISL_519409, EPI_ISL_519410, EPI_ISL_519416, EPI_ISL_519418, EPI_ISL_519422, EPI_ISL_519423, EPI_ISL_519424, EPI_ISL_519426, EPI_ISL_519428, EPI_ISL_519433, EPI_ISL_519436,
EPI_ISL_519438, EPI_ISL_519439, EPI_ISL_519440, EPI_ISL_519442, EPI_ISL_519443, EPI_ISL_519446, EPI_ISL_519447, EPI_ISL_519449, EPI_ISL_519452, EPI_ISL_519453, EPI_ISL_519455, EPI_ISL_519458, EPI_ISL_519459, EPI_ISL_519465, EPI_ISL_519468, EPI_ISL_519469, EPI_ISL_519470, EPI_ISL_519471, EPI_ISL_519472, EPI_ISL_519473, EPI_ISL_519474, EPI_ISL_519476, EPI_ISL_519478, EPI_ISL_519481,
EPI_ISL_519482, EPI_ISL_519484, EPI_ISL_519485, EPI_ISL_519503, EPI_ISL_519504, EPI_ISL_519505, EPI_ISL_519506, EPI_ISL_519507, EPI_ISL_519508, EPI_ISL_519522, EPI_ISL_519523, EPI_ISL_519524, EPI_ISL_519525, EPI_ISL_519526, EPI_ISL_519527, EPI_ISL_519528, EPI_ISL_519529, EPI_ISL_519530, EPI_ISL_519531, EPI_ISL_519532, EPI_ISL_519533, EPI_ISL_519534, EPI_ISL_519535, EPI_ISL_519536,
EPI_ISL_519537, EPI_ISL_519538, EPI_ISL_519539, EPI_ISL_519540, EPI_ISL_519541, EPI_ISL_519542, EPI_ISL_519543, EPI_ISL_519544, EPI_ISL_519545, EPI_ISL_519546, EPI_ISL_519547, EPI_ISL_519548, EPI_ISL_519549, EPI_ISL_519550, EPI_ISL_519551, EPI_ISL_519552, EPI_ISL_519553, EPI_ISL_519554, EPI_ISL_519555, EPI_ISL_519556, EPI_ISL_519557, EPI_ISL_519558, EPI_ISL_519559, EPI_ISL_519560,
EPI_ISL_519561, EPI_ISL_519562, EPI_ISL_519563, EPI_ISL_519564, EPI_ISL_519565, EPI_ISL_519566, EPI_ISL_519567, EPI_ISL_519568, EPI_ISL_519569, EPI_ISL_519570, EPI_ISL_519571, EPI_ISL_519572, EPI_ISL_519573, EPI_ISL_519574, EPI_ISL_519575, EPI_ISL_519576, EPI_ISL_519577, EPI_ISL_519578, EPI_ISL_519579, EPI_ISL_519580, EPI_ISL_519581, EPI_ISL_519582, EPI_ISL_519583, EPI_ISL_519584,
EPI_ISL_519585, EPI_ISL_519586, EPI_ISL_519587, EPI_ISL_519588, EPI_ISL_519589, EPI_ISL_519590, EPI_ISL_519591, EPI_ISL_519592, EPI_ISL_519593, EPI_ISL_519594, EPI_ISL_519595, EPI_ISL_519596, EPI_ISL_519598, EPI_ISL_519599, EPI_ISL_519600, EPI_ISL_519601, EPI_ISL_519602, EPI_ISL_519603, EPI_ISL_519604, EPI_ISL_519605, EPI_ISL_519606, EPI_ISL_519607, EPI_ISL_519609, EPI_ISL_519611,
EPI_ISL_519612, EPI_ISL_519613, EPI_ISL_519614, EPI_ISL_519615, EPI_ISL_519616, EPI_ISL_519617, EPI_ISL_519618, EPI_ISL_519619, EPI_ISL_519621, EPI_ISL_519622, EPI_ISL_519623, EPI_ISL_519624, EPI_ISL_519625, EPI_ISL_519626, EPI_ISL_519627, EPI_ISL_519628, EPI_ISL_519629, EPI_ISL_519631, EPI_ISL_519632, EPI_ISL_519633, EPI_ISL_519635, EPI_ISL_519637, EPI_ISL_519638, EPI_ISL_519639,
EPI_ISL_519641, EPI_ISL_519642, EPI_ISL_519643, EPI_ISL_519646, EPI_ISL_519648, EPI_ISL_519649, EPI_ISL_519650, EPI_ISL_519652, EPI_ISL_519653, EPI_ISL_519654, EPI_ISL_519655, EPI_ISL_519656, EPI_ISL_519657, EPI_ISL_519658, EPI_ISL_519660, EPI_ISL_519662, EPI_ISL_519663, EPI_ISL_519664, EPI_ISL_519665, EPI_ISL_519666, EPI_ISL_519667, EPI_ISL_519668, EPI_ISL_519669, EPI_ISL_519670,
EPI_ISL_519671, EPI_ISL_519672, EPI_ISL_519673, EPI_ISL_519674, EPI_ISL_519675, EPI_ISL_519676, EPI_ISL_519677, EPI_ISL_519678, EPI_ISL_519679, EPI_ISL_519680, EPI_ISL_519681, EPI_ISL_519682, EPI_ISL_519683, EPI_ISL_519684, EPI_ISL_519686, EPI_ISL_519687, EPI_ISL_519688, EPI_ISL_519689, EPI_ISL_519690, EPI_ISL_519691, EPI_ISL_519692, EPI_ISL_519694, EPI_ISL_519695, EPI_ISL_519696,
EPI_ISL_519697, EPI_ISL_519698, EPI_ISL_519699, EPI_ISL_519700, EPI_ISL_519701, EPI_ISL_519703, EPI_ISL_519704, EPI_ISL_519705, EPI_ISL_519706, EPI_ISL_519707, EPI_ISL_519708, EPI_ISL_519709, EPI_ISL_519710, EPI_ISL_519711, EPI_ISL_519712, EPI_ISL_519713, EPI_ISL_519714, EPI_ISL_519715, EPI_ISL_519716, EPI_ISL_519717, EPI_ISL_519718, EPI_ISL_519719, EPI_ISL_519720, EPI_ISL_519721,
EPI_ISL_519722, EPI_ISL_519723, EPI_ISL_519724, EPI_ISL_519726, EPI_ISL_519727, EPI_ISL_519728, EPI_ISL_519729, EPI_ISL_519730, EPI_ISL_519731, EPI_ISL_519732, EPI_ISL_519733, EPI_ISL_519734, EPI_ISL_519735, EPI_ISL_519736, EPI_ISL_519737, EPI_ISL_519738, EPI_ISL_519739, EPI_ISL_519740, EPI_ISL_519741, EPI_ISL_519742, EPI_ISL_519743, EPI_ISL_519744, EPI_ISL_519745, EPI_ISL_519746,
EPI_ISL_519747, EPI_ISL_519748, EPI_ISL_519749, EPI_ISL_519750, EPI_ISL_519751, EPI_ISL_519752, EPI_ISL_519753, EPI_ISL_519754, EPI_ISL_519755, EPI_ISL_519756, EPI_ISL_519757, EPI_ISL_519758, EPI_ISL_519759, EPI_ISL_519760, EPI_ISL_519761, EPI_ISL_519762, EPI_ISL_519763, EPI_ISL_519764, EPI_ISL_519765, EPI_ISL_519766, EPI_ISL_519767, EPI_ISL_519768, EPI_ISL_519769, EPI_ISL_519770,
EPI_ISL_519771, EPI_ISL_519772, EPI_ISL_519773, EPI_ISL_519774, EPI_ISL_519775, EPI_ISL_519776, EPI_ISL_519777, EPI_ISL_519778, EPI_ISL_519779, EPI_ISL_519780, EPI_ISL_519781, EPI_ISL_519782, EPI_ISL_519783, EPI_ISL_519784, EPI_ISL_519785, EPI_ISL_519786, EPI_ISL_519787, EPI_ISL_519788, EPI_ISL_519789, EPI_ISL_519790, EPI_ISL_519791, EPI_ISL_519792, EPI_ISL_519793, EPI_ISL_519794,
EPI_ISL_519795, EPI_ISL_519796, EPI_ISL_519797, EPI_ISL_519798, EPI_ISL_519799, EPI_ISL_519800, EPI_ISL_519801, EPI_ISL_519802, EPI_ISL_519803, EPI_ISL_519804, EPI_ISL_519805, EPI_ISL_519806, EPI_ISL_519807, EPI_ISL_519808, EPI_ISL_519810, EPI_ISL_519811, EPI_ISL_519812, EPI_ISL_519813, EPI_ISL_519814, EPI_ISL_519815, EPI_ISL_519816, EPI_ISL_519817, EPI_ISL_519818, EPI_ISL_519820,
EPI_ISL_519821, EPI_ISL_519822, EPI_ISL_519823, EPI_ISL_519824, EPI_ISL_519825, EPI_ISL_519826, EPI_ISL_519827, EPI_ISL_519828, EPI_ISL_519829, EPI_ISL_519830, EPI_ISL_519831, EPI_ISL_519832, EPI_ISL_519833, EPI_ISL_519834, EPI_ISL_519835, EPI_ISL_519836, EPI_ISL_519837, EPI_ISL_519838, EPI_ISL_519839, EPI_ISL_519840, EPI_ISL_519841, EPI_ISL_519842, EPI_ISL_519843, EPI_ISL_519844,
EPI_ISL_519845, EPI_ISL_519846, EPI_ISL_519847, EPI_ISL_519848, EPI_ISL_519849, EPI_ISL_519850, EPI_ISL_519851, EPI_ISL_519852, EPI_ISL_519853, EPI_ISL_519854, EPI_ISL_519855, EPI_ISL_519856, EPI_ISL_519857, EPI_ISL_519858, EPI_ISL_519859, EPI_ISL_519860, EPI_ISL_519861, EPI_ISL_519863, EPI_ISL_519864, EPI_ISL_519865, EPI_ISL_519866, EPI_ISL_519867, EPI_ISL_519868, EPI_ISL_519869,
EPI_ISL_519870, EPI_ISL_519871, EPI_ISL_519872, EPI_ISL_519873, EPI_ISL_519874, EPI_ISL_519875, EPI_ISL_519876, EPI_ISL_519877, EPI_ISL_519878, EPI_ISL_519880, EPI_ISL_519881, EPI_ISL_519882, EPI_ISL_519883, EPI_ISL_519884, EPI_ISL_519885, EPI_ISL_519886, EPI_ISL_519887, EPI_ISL_519888, EPI_ISL_519889, EPI_ISL_519890, EPI_ISL_519891, EPI_ISL_519892, EPI_ISL_519893, EPI_ISL_519894,
EPI_ISL_519896, EPI_ISL_519897, EPI_ISL_519898, EPI_ISL_519899, EPI_ISL_519900, EPI_ISL_519901, EPI_ISL_519902, EPI_ISL_519903, EPI_ISL_519904, EPI_ISL_519905, EPI_ISL_519906, EPI_ISL_519907, EPI_ISL_519908, EPI_ISL_519909, EPI_ISL_519910, EPI_ISL_519911, EPI_ISL_519912, EPI_ISL_519913, EPI_ISL_519914, EPI_ISL_519915, EPI_ISL_519916, EPI_ISL_519917, EPI_ISL_519918, EPI_ISL_519919,
EPI_ISL_519920, EPI_ISL_519922, EPI_ISL_519923, EPI_ISL_519924, EPI_ISL_519925, EPI_ISL_519926, EPI_ISL_519927, EPI_ISL_519928, EPI_ISL_519929, EPI_ISL_519930, EPI_ISL_519931, EPI_ISL_519932, EPI_ISL_519933, EPI_ISL_519934, EPI_ISL_519935, EPI_ISL_519936, EPI_ISL_519937, EPI_ISL_519938, EPI_ISL_519939, EPI_ISL_519940, EPI_ISL_519941, EPI_ISL_519942, EPI_ISL_519943,
EPI_ISL_519944, EPI_ISL_519945, EPI_ISL_519946, EPI_ISL_519951, EPI_ISL_519952, EPI_ISL_519953, EPI_ISL_519954, EPI_ISL_519955, EPI_ISL_519956, EPI_ISL_519957, EPI_ISL_519958, EPI_ISL_519959, EPI_ISL_519960, EPI_ISL_519961, EPI_ISL_519962, EPI_ISL_519964, EPI_ISL_519965, EPI_ISL_519966, EPI_ISL_519967, EPI_ISL_519968, EPI_ISL_519969, EPI_ISL_519970, EPI_ISL_519971, EPI_ISL_519972,
EPI_ISL_519973, EPI_ISL_519974, EPI_ISL_519975, EPI_ISL_519976, EPI_ISL_519977, EPI_ISL_519978, EPI_ISL_519979, EPI_ISL_519980, EPI_ISL_519982, EPI_ISL_519983, EPI_ISL_519984, EPI_ISL_519985, EPI_ISL_519986, EPI_ISL_519987, EPI_ISL_519988, EPI_ISL_519989, EPI_ISL_519990, EPI_ISL_519991, EPI_ISL_519992, EPI_ISL_519993, EPI_ISL_519994, EPI_ISL_519995, EPI_ISL_519996, EPI_ISL_519997,
EPI_ISL_519998, EPI_ISL_519999, EPI_ISL_520000, EPI_ISL_520001, EPI_ISL_520002, EPI_ISL_520003, EPI_ISL_520004, EPI_ISL_520005, EPI_ISL_520007, EPI_ISL_520008, EPI_ISL_520009, EPI_ISL_520010, EPI_ISL_520011, EPI_ISL_520012, EPI_ISL_520013, EPI_ISL_520014, EPI_ISL_520015, EPI_ISL_520016, EPI_ISL_520017, EPI_ISL_520018, EPI_ISL_520019, EPI_ISL_520020, EPI_ISL_520021, EPI_ISL_520022,
EPI_ISL_520023, EPI_ISL_520024, EPI_ISL_520025, EPI_ISL_520026, EPI_ISL_520027, EPI_ISL_520028, EPI_ISL_520029, EPI_ISL_520030, EPI_ISL_520031, EPI_ISL_520032, EPI_ISL_520033, EPI_ISL_520034, EPI_ISL_520035, EPI_ISL_520036, EPI_ISL_520037, EPI_ISL_520038, EPI_ISL_520039, EPI_ISL_520040, EPI_ISL_520041, EPI_ISL_520042, EPI_ISL_520043, EPI_ISL_520044, EPI_ISL_520045, EPI_ISL_520046,
EPI_ISL_520047, EPI_ISL_520048, EPI_ISL_520049, EPI_ISL_520050, EPI_ISL_520052, EPI_ISL_520053, EPI_ISL_520054, EPI_ISL_520055, EPI_ISL_520056, EPI_ISL_520057, EPI_ISL_520058, EPI_ISL_520059, EPI_ISL_520060, EPI_ISL_520061, EPI_ISL_520062, EPI_ISL_520063, EPI_ISL_520065, EPI_ISL_520066, EPI_ISL_520067, EPI_ISL_520068, EPI_ISL_520069, EPI_ISL_520070, EPI_ISL_520071, EPI_ISL_520073,
EPI_ISL_520074, EPI_ISL_520075, EPI_ISL_520076, EPI_ISL_520077, EPI_ISL_520078, EPI_ISL_520079, EPI_ISL_520080, EPI_ISL_520081, EPI_ISL_520082, EPI_ISL_520083, EPI_ISL_520084, EPI_ISL_520085, EPI_ISL_520086, EPI_ISL_520087, EPI_ISL_520088, EPI_ISL_520089, EPI_ISL_520090, EPI_ISL_520092, EPI_ISL_520093, EPI_ISL_520094, EPI_ISL_520095, EPI_ISL_520096, EPI_ISL_520097, EPI_ISL_520098,
EPI_ISL_520099, EPI_ISL_520100, EPI_ISL_520101, EPI_ISL_520102, EPI_ISL_520103, EPI_ISL_520104, EPI_ISL_520105, EPI_ISL_520106, EPI_ISL_520107, EPI_ISL_520108, EPI_ISL_520109, EPI_ISL_520110, EPI_ISL_520111, EPI_ISL_520112, EPI_ISL_520113, EPI_ISL_520114, EPI_ISL_520115, EPI_ISL_520116, EPI_ISL_520117, EPI_ISL_520118, EPI_ISL_520121, EPI_ISL_520122, EPI_ISL_520123, EPI_ISL_520124,
EPI_ISL_520125, EPI_ISL_520126, EPI_ISL_520127, EPI_ISL_520128, EPI_ISL_520129, EPI_ISL_520130, EPI_ISL_520131, EPI_ISL_520132, EPI_ISL_520133, EPI_ISL_520134, EPI_ISL_520135, EPI_ISL_520137, EPI_ISL_520138, EPI_ISL_520139, EPI_ISL_520140, EPI_ISL_520141, EPI_ISL_520142, EPI_ISL_520143, EPI_ISL_520144, EPI_ISL_520145, EPI_ISL_520146, EPI_ISL_520147, EPI_ISL_520148,
EPI_ISL_520149, EPI_ISL_520150, EPI_ISL_520151, EPI_ISL_520152, EPI_ISL_520153, EPI_ISL_520154, EPI_ISL_520155, EPI_ISL_520156, EPI_ISL_520157, EPI_ISL_520158, EPI_ISL_520159, EPI_ISL_520160, EPI_ISL_520161, EPI_ISL_520162, EPI_ISL_520163, EPI_ISL_520164, EPI_ISL_520165, EPI_ISL_520166, EPI_ISL_520167, EPI_ISL_520168, EPI_ISL_520169, EPI_ISL_520170, EPI_ISL_520171, EPI_ISL_520172,
EPI_ISL_520173, EPI_ISL_520174, EPI_ISL_520175, EPI_ISL_520176, EPI_ISL_520177, EPI_ISL_520178, EPI_ISL_520179, EPI_ISL_520180, EPI_ISL_520215, EPI_ISL_520216, EPI_ISL_520217, EPI_ISL_520218, EPI_ISL_520219, EPI_ISL_520220, EPI_ISL_520221, EPI_ISL_520222, EPI_ISL_520226, EPI_ISL_520227, EPI_ISL_520228, EPI_ISL_520229, EPI_ISL_520230, EPI_ISL_520231, EPI_ISL_520233,
EPI_ISL_520234, EPI_ISL_520235, EPI_ISL_520236, EPI_ISL_520237, EPI_ISL_520238, EPI_ISL_520239, EPI_ISL_520240, EPI_ISL_520241, EPI_ISL_520243, EPI_ISL_520244, EPI_ISL_520245, EPI_ISL_520246, EPI_ISL_520247, EPI_ISL_520248, EPI_ISL_520396, EPI_ISL_520397, EPI_ISL_520398, EPI_ISL_520399, EPI_ISL_520401, EPI_ISL_520402, EPI_ISL_520404, EPI_ISL_520405, EPI_ISL_520407, EPI_ISL_520408,
EPI_ISL_520410, EPI_ISL_520411, EPI_ISL_520412, EPI_ISL_520418, EPI_ISL_520422, EPI_ISL_520425, EPI_ISL_520426, EPI_ISL_520427, EPI_ISL_520430, EPI_ISL_520431, EPI_ISL_520433, EPI_ISL_520437, EPI_ISL_520439, EPI_ISL_520445, EPI_ISL_520446, EPI_ISL_520447, EPI_ISL_520449, EPI_ISL_520450, EPI_ISL_520452, EPI_ISL_520453, EPI_ISL_520454, EPI_ISL_520455, EPI_ISL_520456,
EPI_ISL_520458, EPI_ISL_520460, EPI_ISL_520465, EPI_ISL_520469, EPI_ISL_520473, EPI_ISL_520474, EPI_ISL_520475, EPI_ISL_520501, EPI_ISL_520503, EPI_ISL_520504, EPI_ISL_520505, EPI_ISL_520507, EPI_ISL_520508, EPI_ISL_520509, EPI_ISL_520510, EPI_ISL_520511, EPI_ISL_520512, EPI_ISL_520513, EPI_ISL_520515, EPI_ISL_520517, EPI_ISL_520518, EPI_ISL_520519, EPI_ISL_520520, EPI_ISL_520521,
EPI_ISL_520522, EPI_ISL_520523, EPI_ISL_520524, EPI_ISL_520525, EPI_ISL_520526, EPI_ISL_520527, EPI_ISL_520528, EPI_ISL_520529, EPI_ISL_520530, EPI_ISL_520532, EPI_ISL_520533, EPI_ISL_520534, EPI_ISL_520535, EPI_ISL_520536, EPI_ISL_520537, EPI_ISL_520538, EPI_ISL_520539, EPI_ISL_520540, EPI_ISL_520541, EPI_ISL_520542, EPI_ISL_520543, EPI_ISL_520544, EPI_ISL_520545, EPI_ISL_520546,
EPI_ISL_520548, EPI_ISL_520551, EPI_ISL_520552, EPI_ISL_520553, EPI_ISL_520554, EPI_ISL_520555, EPI_ISL_520556, EPI_ISL_520557, EPI_ISL_520558, EPI_ISL_520560, EPI_ISL_520561, EPI_ISL_520562, EPI_ISL_520563, EPI_ISL_520564, EPI_ISL_520566, EPI_ISL_520567, EPI_ISL_520568, EPI_ISL_520569, EPI_ISL_520570, EPI_ISL_520571, EPI_ISL_520572, EPI_ISL_520574, EPI_ISL_520575, EPI_ISL_520576,
EPI_ISL_520577, EPI_ISL_520578, EPI_ISL_520579, EPI_ISL_520580, EPI_ISL_520581, EPI_ISL_520582, EPI_ISL_520583, EPI_ISL_520584, EPI_ISL_520585, EPI_ISL_520586, EPI_ISL_520587, EPI_ISL_520588, EPI_ISL_520589, EPI_ISL_520590, EPI_ISL_520591, EPI_ISL_520592, EPI_ISL_520594, EPI_ISL_520595, EPI_ISL_520596, EPI_ISL_520597, EPI_ISL_520598, EPI_ISL_520599, EPI_ISL_520600,
EPI_ISL_520601, EPI_ISL_520602, EPI_ISL_520603, EPI_ISL_520606, EPI_ISL_520607, EPI_ISL_520608, EPI_ISL_520609, EPI_ISL_520610, EPI_ISL_520611, EPI_ISL_520612, EPI_ISL_520613, EPI_ISL_520614, EPI_ISL_520615, EPI_ISL_520616, EPI_ISL_520617, EPI_ISL_520620, EPI_ISL_520621, EPI_ISL_520622, EPI_ISL_520623, EPI_ISL_520624, EPI_ISL_520625, EPI_ISL_520626, EPI_ISL_520627, EPI_ISL_520628,
EPI_ISL_520629, EPI_ISL_520630, EPI_ISL_520633, EPI_ISL_520634, EPI_ISL_520635, EPI_ISL_520636, EPI_ISL_520637, EPI_ISL_520638, EPI_ISL_520640, EPI_ISL_520641, EPI_ISL_520642, EPI_ISL_520643, EPI_ISL_520645, EPI_ISL_520646, EPI_ISL_520647, EPI_ISL_520648, EPI_ISL_520649, EPI_ISL_520650, EPI_ISL_520651, EPI_ISL_520652, EPI_ISL_520654, EPI_ISL_520655, EPI_ISL_520656,
EPI_ISL_520657, EPI_ISL_520658, EPI_ISL_520659, EPI_ISL_520660, EPI_ISL_520661, EPI_ISL_520662, EPI_ISL_520765, EPI_ISL_520766, EPI_ISL_520767, EPI_ISL_520768, EPI_ISL_520769, EPI_ISL_520770, EPI_ISL_520771, EPI_ISL_520772, EPI_ISL_520773, EPI_ISL_520774, EPI_ISL_520776, EPI_ISL_520777, EPI_ISL_520778, EPI_ISL_520779, EPI_ISL_520780, EPI_ISL_520781, EPI_ISL_520782, EPI_ISL_520783,
EPI_ISL_520784, EPI_ISL_520785, EPI_ISL_520786, EPI_ISL_520787, EPI_ISL_520788, EPI_ISL_520789, EPI_ISL_520790, EPI_ISL_520844, EPI_ISL_520845, EPI_ISL_520846, EPI_ISL_520848, EPI_ISL_520849, EPI_ISL_520850, EPI_ISL_520851, EPI_ISL_520852, EPI_ISL_520853, EPI_ISL_520854, EPI_ISL_520855, EPI_ISL_520856, EPI_ISL_520857, EPI_ISL_520858, EPI_ISL_520859, EPI_ISL_520860, EPI_ISL_520861,
EPI_ISL_520862, EPI_ISL_520863, EPI_ISL_520929, EPI_ISL_520930, EPI_ISL_520931, EPI_ISL_520932, EPI_ISL_520933, EPI_ISL_520934, EPI_ISL_520935, EPI_ISL_520936, EPI_ISL_520937, EPI_ISL_520938, EPI_ISL_520939, EPI_ISL_520940, EPI_ISL_520941, EPI_ISL_520942, EPI_ISL_520943, EPI_ISL_520944, EPI_ISL_520945, EPI_ISL_520946, EPI_ISL_520947, EPI_ISL_520948, EPI_ISL_520949, EPI_ISL_520950,
EPI_ISL_520951, EPI_ISL_520952, EPI_ISL_520953, EPI_ISL_520954, EPI_ISL_520955, EPI_ISL_520956, EPI_ISL_520957, EPI_ISL_520958, EPI_ISL_520959, EPI_ISL_520960, EPI_ISL_520962, EPI_ISL_520963, EPI_ISL_520964, EPI_ISL_520965, EPI_ISL_520966, EPI_ISL_520967, EPI_ISL_520968, EPI_ISL_520969, EPI_ISL_520970, EPI_ISL_520971, EPI_ISL_520972, EPI_ISL_520973, EPI_ISL_520974, EPI_ISL_520975,
EPI_ISL_520976, EPI_ISL_520977, EPI_ISL_520978, EPI_ISL_520979, EPI_ISL_520980, EPI_ISL_520981, EPI_ISL_520982, EPI_ISL_520983, EPI_ISL_520984, EPI_ISL_520986, EPI_ISL_520987, EPI_ISL_520988, EPI_ISL_520989, EPI_ISL_520991, EPI_ISL_520992, EPI_ISL_520993, EPI_ISL_520994, EPI_ISL_520995, EPI_ISL_520996, EPI_ISL_520997, EPI_ISL_520998, EPI_ISL_520999, EPI_ISL_521000, EPI_ISL_521001,
EPI_ISL_521002, EPI_ISL_521003, EPI_ISL_521004, EPI_ISL_521005, EPI_ISL_521006, EPI_ISL_521007, EPI_ISL_521008, EPI_ISL_521009, EPI_ISL_521010, EPI_ISL_521011, EPI_ISL_521012, EPI_ISL_521013, EPI_ISL_521014, EPI_ISL_521015, EPI_ISL_521016, EPI_ISL_521017, EPI_ISL_521018, EPI_ISL_521019, EPI_ISL_521020, EPI_ISL_521021, EPI_ISL_521022, EPI_ISL_521023, EPI_ISL_521024, EPI_ISL_521025,
EPI_ISL_521026, EPI_ISL_521027, EPI_ISL_521028, EPI_ISL_521029, EPI_ISL_521030, EPI_ISL_521031, EPI_ISL_521032, EPI_ISL_521033, EPI_ISL_521034, EPI_ISL_521035, EPI_ISL_521036, EPI_ISL_521037, EPI_ISL_521038, EPI_ISL_521039, EPI_ISL_521040, EPI_ISL_521041, EPI_ISL_521042, EPI_ISL_521043, EPI_ISL_521044, EPI_ISL_521045, EPI_ISL_521046, EPI_ISL_521047, EPI_ISL_521048, EPI_ISL_521049,
EPI_ISL_521050, EPI_ISL_521051, EPI_ISL_521052, EPI_ISL_521053, EPI_ISL_521054, EPI_ISL_521055, EPI_ISL_521056, EPI_ISL_521057, EPI_ISL_521058, EPI_ISL_521059, EPI_ISL_521060, EPI_ISL_521061, EPI_ISL_521062, EPI_ISL_521063, EPI_ISL_521064, EPI_ISL_521065, EPI_ISL_521066, EPI_ISL_521067, EPI_ISL_521068, EPI_ISL_521069, EPI_ISL_521070, EPI_ISL_521071, EPI_ISL_521072, EPI_ISL_521073,
EPI_ISL_521074, EPI_ISL_521075, EPI_ISL_521076, EPI_ISL_521077, EPI_ISL_521078, EPI_ISL_521079, EPI_ISL_521080, EPI_ISL_521081, EPI_ISL_521082, EPI_ISL_521083, EPI_ISL_521084, EPI_ISL_521085, EPI_ISL_521086, EPI_ISL_521087, EPI_ISL_521089, EPI_ISL_521090, EPI_ISL_521091, EPI_ISL_521092, EPI_ISL_521093, EPI_ISL_521094, EPI_ISL_521095, EPI_ISL_521096, EPI_ISL_521097, EPI_ISL_521098,
EPI_ISL_521099, EPI_ISL_521100, EPI_ISL_521101, EPI_ISL_521154, EPI_ISL_521156, EPI_ISL_521157, EPI_ISL_521158, EPI_ISL_521159, EPI_ISL_521160, EPI_ISL_521161, EPI_ISL_521162, EPI_ISL_521163, EPI_ISL_521164, EPI_ISL_521165, EPI_ISL_521166, EPI_ISL_521167, EPI_ISL_521168, EPI_ISL_521170, EPI_ISL_521171, EPI_ISL_521172, EPI_ISL_521173, EPI_ISL_521174, EPI_ISL_521175, EPI_ISL_521176,
EPI_ISL_521177, EPI_ISL_521178, EPI_ISL_521179, EPI_ISL_521180, EPI_ISL_521181, EPI_ISL_521182, EPI_ISL_521183, EPI_ISL_521184, EPI_ISL_521185, EPI_ISL_521186, EPI_ISL_521187, EPI_ISL_521188, EPI_ISL_521189, EPI_ISL_521190, EPI_ISL_521191, EPI_ISL_521192, EPI_ISL_521194, EPI_ISL_521195, EPI_ISL_521196, EPI_ISL_521197, EPI_ISL_521198, EPI_ISL_521199, EPI_ISL_521200, EPI_ISL_521201,
EPI_ISL_521202, EPI_ISL_521203, EPI_ISL_521204, EPI_ISL_521205, EPI_ISL_521206, EPI_ISL_521207, EPI_ISL_521208, EPI_ISL_521210, EPI_ISL_521212, EPI_ISL_521213, EPI_ISL_521214, EPI_ISL_521215, EPI_ISL_521217, EPI_ISL_521218, EPI_ISL_521219, EPI_ISL_521220, EPI_ISL_521221, EPI_ISL_521227, EPI_ISL_521228, EPI_ISL_521229, EPI_ISL_521230, EPI_ISL_521231, EPI_ISL_521232, EPI_ISL_521234,
EPI_ISL_521235, EPI_ISL_521237, EPI_ISL_521238, EPI_ISL_521239, EPI_ISL_521240, EPI_ISL_521241, EPI_ISL_521243, EPI_ISL_521244, EPI_ISL_521245, EPI_ISL_521246, EPI_ISL_521247, EPI_ISL_521248, EPI_ISL_521249, EPI_ISL_521250, EPI_ISL_521251, EPI_ISL_521252, EPI_ISL_521253, EPI_ISL_521254, EPI_ISL_521255, EPI_ISL_521256, EPI_ISL_521257, EPI_ISL_521258, EPI_ISL_521259, EPI_ISL_521260,
EPI_ISL_521261, EPI_ISL_521262, EPI_ISL_521263, EPI_ISL_521264, EPI_ISL_521265, EPI_ISL_521266, EPI_ISL_521267, EPI_ISL_521268, EPI_ISL_521278, EPI_ISL_521279, EPI_ISL_521280, EPI_ISL_521281, EPI_ISL_521282, EPI_ISL_521283, EPI_ISL_521286, EPI_ISL_521288, EPI_ISL_521289, EPI_ISL_521290, EPI_ISL_521291, EPI_ISL_521292, EPI_ISL_521293, EPI_ISL_521294, EPI_ISL_521295, EPI_ISL_521296,
EPI_ISL_521297, EPI_ISL_521298, EPI_ISL_521299, EPI_ISL_521300, EPI_ISL_521301, EPI_ISL_521302, EPI_ISL_521303, EPI_ISL_521304, EPI_ISL_521305, EPI_ISL_521306, EPI_ISL_521307, EPI_ISL_521308, EPI_ISL_521310, EPI_ISL_521311, EPI_ISL_521312, EPI_ISL_521313, EPI_ISL_521314, EPI_ISL_521452, EPI_ISL_521453, EPI_ISL_521454, EPI_ISL_521455, EPI_ISL_521456, EPI_ISL_521457, EPI_ISL_521458,
EPI_ISL_521459, EPI_ISL_521460, EPI_ISL_521461, EPI_ISL_521562, EPI_ISL_521563, EPI_ISL_521564, EPI_ISL_521565, EPI_ISL_521566, EPI_ISL_521567, EPI_ISL_521568, EPI_ISL_521569, EPI_ISL_521570, EPI_ISL_521571, EPI_ISL_521572, EPI_ISL_521573, EPI_ISL_521574, EPI_ISL_521575, EPI_ISL_521576, EPI_ISL_521577, EPI_ISL_521578, EPI_ISL_521579, EPI_ISL_521580, EPI_ISL_521581, EPI_ISL_521582,
EPI_ISL_521583, EPI_ISL_521584, EPI_ISL_521585, EPI_ISL_521586, EPI_ISL_521587, EPI_ISL_521588, EPI_ISL_521589, EPI_ISL_521590, EPI_ISL_521591, EPI_ISL_521592, EPI_ISL_521593, EPI_ISL_521594, EPI_ISL_521595, EPI_ISL_521596, EPI_ISL_521597, EPI_ISL_521598, EPI_ISL_521599, EPI_ISL_521600, EPI_ISL_521601, EPI_ISL_521602, EPI_ISL_521603, EPI_ISL_521604, EPI_ISL_521605, EPI_ISL_521606,
EPI_ISL_521607, EPI_ISL_521608, EPI_ISL_521609, EPI_ISL_521610, EPI_ISL_521611, EPI_ISL_521612, EPI_ISL_521613, EPI_ISL_521614, EPI_ISL_521615, EPI_ISL_521616, EPI_ISL_521617, EPI_ISL_521618, EPI_ISL_521619, EPI_ISL_521620, EPI_ISL_521621, EPI_ISL_521622, EPI_ISL_521623, EPI_ISL_521624, EPI_ISL_521626, EPI_ISL_521627, EPI_ISL_521628, EPI_ISL_521629, EPI_ISL_521630, EPI_ISL_521631,
EPI_ISL_521632, EPI_ISL_521633, EPI_ISL_521634, EPI_ISL_521635, EPI_ISL_521636, EPI_ISL_521637, EPI_ISL_521638, EPI_ISL_521639, EPI_ISL_521640, EPI_ISL_521641, EPI_ISL_521642, EPI_ISL_521643, EPI_ISL_521645, EPI_ISL_521646, EPI_ISL_521647, EPI_ISL_521648, EPI_ISL_521649, EPI_ISL_521650, EPI_ISL_521651, EPI_ISL_521652, EPI_ISL_521653, EPI_ISL_521654, EPI_ISL_521655,
EPI_ISL_521656, EPI_ISL_521657, EPI_ISL_521658, EPI_ISL_521659, EPI_ISL_521660, EPI_ISL_521661, EPI_ISL_521662, EPI_ISL_521665, EPI_ISL_521666, EPI_ISL_521667, EPI_ISL_521668, EPI_ISL_521669, EPI_ISL_521670, EPI_ISL_521671, EPI_ISL_521672, EPI_ISL_521673, EPI_ISL_521674, EPI_ISL_521675, EPI_ISL_521676, EPI_ISL_521677, EPI_ISL_521678, EPI_ISL_521679, EPI_ISL_521680,
EPI_ISL_521681, EPI_ISL_521682, EPI_ISL_521683, EPI_ISL_521684, EPI_ISL_521685, EPI_ISL_521686, EPI_ISL_521687, EPI_ISL_521688, EPI_ISL_521689, EPI_ISL_521690, EPI_ISL_521691, EPI_ISL_521692, EPI_ISL_521693, EPI_ISL_521694, EPI_ISL_521695, EPI_ISL_521696, EPI_ISL_521697, EPI_ISL_521698, EPI_ISL_521699, EPI_ISL_521700, EPI_ISL_521701, EPI_ISL_521702, EPI_ISL_521703, EPI_ISL_521704,
EPI_ISL_521705, EPI_ISL_521706, EPI_ISL_521707, EPI_ISL_521708, EPI_ISL_521709, EPI_ISL_521710, EPI_ISL_521711, EPI_ISL_521712, EPI_ISL_521713, EPI_ISL_521714, EPI_ISL_521715, EPI_ISL_521716, EPI_ISL_521717, EPI_ISL_521718, EPI_ISL_521719, EPI_ISL_521720, EPI_ISL_521721, EPI_ISL_521722, EPI_ISL_521723, EPI_ISL_521724, EPI_ISL_521725, EPI_ISL_521726, EPI_ISL_521727, EPI_ISL_521728,

EPI_ISL_521729, EPI_ISL_521730, EPI_ISL_521732, EPI_ISL_521733, EPI_ISL_521734, EPI_ISL_521735, EPI_ISL_521736, EPI_ISL_521737, EPI_ISL_521738, EPI_ISL_521739, EPI_ISL_521740, EPI_ISL_521741, EPI_ISL_521742, EPI_ISL_521743, EPI_ISL_521744, EPI_ISL_521745, EPI_ISL_521746, EPI_ISL_521747, EPI_ISL_521748, EPI_ISL_521749, EPI_ISL_521750, EPI_ISL_521751, EPI_ISL_521752, EPI_ISL_521753,
EPI_ISL_521754, EPI_ISL_521755, EPI_ISL_521756, EPI_ISL_521757, EPI_ISL_521759, EPI_ISL_521760, EPI_ISL_521761, EPI_ISL_521762, EPI_ISL_521763, EPI_ISL_521764, EPI_ISL_521765, EPI_ISL_521766, EPI_ISL_521767, EPI_ISL_521768, EPI_ISL_521769, EPI_ISL_521770, EPI_ISL_521771, EPI_ISL_521772, EPI_ISL_521773, EPI_ISL_521774, EPI_ISL_521775, EPI_ISL_521776, EPI_ISL_521777, EPI_ISL_521778,
EPI_ISL_521779, EPI_ISL_521780, EPI_ISL_521781, EPI_ISL_521782, EPI_ISL_521783, EPI_ISL_521784, EPI_ISL_521785, EPI_ISL_521786, EPI_ISL_521787, EPI_ISL_521789, EPI_ISL_521790, EPI_ISL_521791, EPI_ISL_521792, EPI_ISL_521793, EPI_ISL_521794, EPI_ISL_521795, EPI_ISL_521796, EPI_ISL_521797, EPI_ISL_521798, EPI_ISL_521799, EPI_ISL_521800, EPI_ISL_521801, EPI_ISL_521802, EPI_ISL_521803,
EPI_ISL_521804, EPI_ISL_521806, EPI_ISL_521807, EPI_ISL_521808, EPI_ISL_521809, EPI_ISL_521811, EPI_ISL_521813, EPI_ISL_521814, EPI_ISL_521815, EPI_ISL_521816, EPI_ISL_521817, EPI_ISL_521818, EPI_ISL_521819, EPI_ISL_521820, EPI_ISL_521821, EPI_ISL_521823, EPI_ISL_521824, EPI_ISL_521825, EPI_ISL_521826, EPI_ISL_521827, EPI_ISL_521828, EPI_ISL_521829, EPI_ISL_521830, EPI_ISL_521831,
EPI_ISL_521832, EPI_ISL_521833, EPI_ISL_521834, EPI_ISL_521835, EPI_ISL_521836, EPI_ISL_521837, EPI_ISL_521838, EPI_ISL_521839, EPI_ISL_521927, EPI_ISL_521928, EPI_ISL_521929, EPI_ISL_521930, EPI_ISL_521931, EPI_ISL_521932, EPI_ISL_521933, EPI_ISL_521934, EPI_ISL_521935, EPI_ISL_521936, EPI_ISL_521937, EPI_ISL_521938, EPI_ISL_521939, EPI_ISL_521940, EPI_ISL_521941, EPI_ISL_521942,
EPI_ISL_521943, EPI_ISL_521944, EPI_ISL_521973, EPI_ISL_521974, EPI_ISL_521989, EPI_ISL_521990, EPI_ISL_521991, EPI_ISL_521992, EPI_ISL_521993, EPI_ISL_521994, EPI_ISL_521995, EPI_ISL_521996, EPI_ISL_521997, EPI_ISL_521998, EPI_ISL_521999, EPI_ISL_522000, EPI_ISL_522001, EPI_ISL_522002, EPI_ISL_522003, EPI_ISL_522004, EPI_ISL_522005, EPI_ISL_522006, EPI_ISL_522007, EPI_ISL_522008,
EPI_ISL_522009, EPI_ISL_522010, EPI_ISL_522011, EPI_ISL_522012, EPI_ISL_522013, EPI_ISL_522015, EPI_ISL_522016, EPI_ISL_522018, EPI_ISL_522019, EPI_ISL_522020, EPI_ISL_522021, EPI_ISL_522022, EPI_ISL_522023, EPI_ISL_522024, EPI_ISL_522025, EPI_ISL_522027, EPI_ISL_522029, EPI_ISL_522030, EPI_ISL_522031, EPI_ISL_522032, EPI_ISL_522033, EPI_ISL_522034, EPI_ISL_522036, EPI_ISL_522037,
EPI_ISL_522038, EPI_ISL_522039, EPI_ISL_522040, EPI_ISL_522041, EPI_ISL_522042, EPI_ISL_522043, EPI_ISL_522044, EPI_ISL_522045, EPI_ISL_522046, EPI_ISL_522047, EPI_ISL_522048, EPI_ISL_522049, EPI_ISL_522050, EPI_ISL_522051, EPI_ISL_522052, EPI_ISL_522053, EPI_ISL_522054, EPI_ISL_522112, EPI_ISL_522113, EPI_ISL_522114, EPI_ISL_522115, EPI_ISL_522116, EPI_ISL_522117, EPI_ISL_522118,
EPI_ISL_522188, EPI_ISL_522189, EPI_ISL_522190, EPI_ISL_522191, EPI_ISL_522192, EPI_ISL_522193, EPI_ISL_522194, EPI_ISL_522195, EPI_ISL_522196, EPI_ISL_522197, EPI_ISL_522198, EPI_ISL_522199, EPI_ISL_522201, EPI_ISL_522202, EPI_ISL_522203, EPI_ISL_522204, EPI_ISL_522205, EPI_ISL_522206, EPI_ISL_522207, EPI_ISL_522208, EPI_ISL_522209, EPI_ISL_522210, EPI_ISL_522211, EPI_ISL_522212,
EPI_ISL_522213, EPI_ISL_522214, EPI_ISL_522215, EPI_ISL_522216, EPI_ISL_522217, EPI_ISL_522218, EPI_ISL_522219, EPI_ISL_522220, EPI_ISL_522221, EPI_ISL_522222, EPI_ISL_522223, EPI_ISL_522224, EPI_ISL_522225, EPI_ISL_522226, EPI_ISL_522227, EPI_ISL_522228, EPI_ISL_522229, EPI_ISL_522230, EPI_ISL_522231, EPI_ISL_522243, EPI_ISL_522245, EPI_ISL_522247, EPI_ISL_522248, EPI_ISL_522249,
EPI_ISL_522250, EPI_ISL_522252, EPI_ISL_522253, EPI_ISL_561411, EPI_ISL_561415, EPI_ISL_561417, EPI_ISL_561422, EPI_ISL_561423, EPI_ISL_561424, EPI_ISL_561427, EPI_ISL_561430, EPI_ISL_561431, EPI_ISL_561432, EPI_ISL_561435, EPI_ISL_561436, EPI_ISL_561439, EPI_ISL_561442, EPI_ISL_561443, EPI_ISL_561444, EPI_ISL_561445, EPI_ISL_561446, EPI_ISL_561447, EPI_ISL_561448, EPI_ISL_561452,
EPI_ISL_561460, EPI_ISL_561461, EPI_ISL_561463, EPI_ISL_561464, EPI_ISL_561466, EPI_ISL_561467, EPI_ISL_561469, EPI_ISL_561472, EPI_ISL_561474, EPI_ISL_561475, EPI_ISL_561476, EPI_ISL_561478, EPI_ISL_561481, EPI_ISL_561482, EPI_ISL_561483, EPI_ISL_561484, EPI_ISL_561485, EPI_ISL_561486, EPI_ISL_561487, EPI_ISL_561488, EPI_ISL_561489, EPI_ISL_561490, EPI_ISL_561492, EPI_ISL_561493,
EPI_ISL_561494, EPI_ISL_561495, EPI_ISL_561496, EPI_ISL_561497, EPI_ISL_561499, EPI_ISL_561500, EPI_ISL_561502, EPI_ISL_561503, EPI_ISL_561504, EPI_ISL_561505, EPI_ISL_561506, EPI_ISL_561507, EPI_ISL_561509, EPI_ISL_561511, EPI_ISL_561512, EPI_ISL_561513, EPI_ISL_561516, EPI_ISL_561520, EPI_ISL_561521, EPI_ISL_561522, EPI_ISL_561524, EPI_ISL_561525, EPI_ISL_561526, EPI_ISL_561527,
EPI_ISL_561528, EPI_ISL_561529, EPI_ISL_561530, EPI_ISL_561534, EPI_ISL_561536, EPI_ISL_561537, EPI_ISL_561538, EPI_ISL_561539, EPI_ISL_561540, EPI_ISL_561542, EPI_ISL_561543, EPI_ISL_561545, EPI_ISL_561546, EPI_ISL_561547, EPI_ISL_561548, EPI_ISL_561549, EPI_ISL_561551, EPI_ISL_561552, EPI_ISL_561553, EPI_ISL_561554, EPI_ISL_561555, EPI_ISL_561556, EPI_ISL_561557, EPI_ISL_561558,
EPI_ISL_561560, EPI_ISL_561561, EPI_ISL_561563, EPI_ISL_561564, EPI_ISL_561566, EPI_ISL_561568, EPI_ISL_561569, EPI_ISL_561570, EPI_ISL_561571, EPI_ISL_561572, EPI_ISL_561573, EPI_ISL_561575, EPI_ISL_561576, EPI_ISL_561579, EPI_ISL_561580, EPI_ISL_561581, EPI_ISL_561585, EPI_ISL_561589, EPI_ISL_561592, EPI_ISL_561593, EPI_ISL_561594, EPI_ISL_561596, EPI_ISL_561598, EPI_ISL_561599,
EPI_ISL_561600, EPI_ISL_561601, EPI_ISL_561603, EPI_ISL_561604, EPI_ISL_561605, EPI_ISL_561606, EPI_ISL_561607, EPI_ISL_561608, EPI_ISL_561609, EPI_ISL_561610, EPI_ISL_561611, EPI_ISL_561615, EPI_ISL_561616, EPI_ISL_561617, EPI_ISL_561618, EPI_ISL_561619, EPI_ISL_561620, EPI_ISL_561624, EPI_ISL_561627, EPI_ISL_561628, EPI_ISL_561631, EPI_ISL_561632, EPI_ISL_561634, EPI_ISL_561635,
EPI_ISL_561636, EPI_ISL_561637, EPI_ISL_561638, EPI_ISL_561639, EPI_ISL_561640, EPI_ISL_561641, EPI_ISL_561642, EPI_ISL_561643, EPI_ISL_561644, EPI_ISL_561645, EPI_ISL_561646, EPI_ISL_561648, EPI_ISL_561649, EPI_ISL_561650, EPI_ISL_561652, EPI_ISL_561654, EPI_ISL_561658, EPI_ISL_561659, EPI_ISL_561660, EPI_ISL_561662, EPI_ISL_561663, EPI_ISL_561664, EPI_ISL_561665, EPI_ISL_561667,
EPI_ISL_561670, EPI_ISL_561671, EPI_ISL_561672, EPI_ISL_561675, EPI_ISL_561676, EPI_ISL_561677, EPI_ISL_561678, EPI_ISL_561680, EPI_ISL_561681, EPI_ISL_561683, EPI_ISL_561684, EPI_ISL_561685, EPI_ISL_561687, EPI_ISL_561688, EPI_ISL_561689, EPI_ISL_561691, EPI_ISL_561692, EPI_ISL_561693, EPI_ISL_561694, EPI_ISL_561695, EPI_ISL_561696, EPI_ISL_561699, EPI_ISL_561700, EPI_ISL_561701,
EPI_ISL_561702, EPI_ISL_561703, EPI_ISL_561704, EPI_ISL_561705, EPI_ISL_561706, EPI_ISL_561708, EPI_ISL_561709, EPI_ISL_561710, EPI_ISL_561712, EPI_ISL_561713, EPI_ISL_561716, EPI_ISL_561717, EPI_ISL_561718, EPI_ISL_561719, EPI_ISL_561720, EPI_ISL_561722, EPI_ISL_561723, EPI_ISL_561724, EPI_ISL_561725, EPI_ISL_561726, EPI_ISL_561729, EPI_ISL_561730, EPI_ISL_561731, EPI_ISL_561732,
EPI_ISL_561733, EPI_ISL_561734, EPI_ISL_561735, EPI_ISL_561736, EPI_ISL_561737, EPI_ISL_561738, EPI_ISL_561739, EPI_ISL_561740, EPI_ISL_561742, EPI_ISL_561743, EPI_ISL_561744, EPI_ISL_561745, EPI_ISL_561746, EPI_ISL_561747, EPI_ISL_561748, EPI_ISL_561749, EPI_ISL_561751, EPI_ISL_561753, EPI_ISL_561754, EPI_ISL_561755, EPI_ISL_561756, EPI_ISL_561757, EPI_ISL_561758, EPI_ISL_561759,
EPI_ISL_561760, EPI_ISL_561761, EPI_ISL_561762, EPI_ISL_561763, EPI_ISL_561767, EPI_ISL_561768, EPI_ISL_561770, EPI_ISL_561771, EPI_ISL_561772, EPI_ISL_561773, EPI_ISL_561774, EPI_ISL_561775, EPI_ISL_561779, EPI_ISL_561780, EPI_ISL_561781, EPI_ISL_561784, EPI_ISL_561786, EPI_ISL_561790, EPI_ISL_561792, EPI_ISL_561794, EPI_ISL_561795, EPI_ISL_561797, EPI_ISL_561800, EPI_ISL_561801,
EPI_ISL_561802, EPI_ISL_561804, EPI_ISL_561806, EPI_ISL_561807, EPI_ISL_561809, EPI_ISL_561810, EPI_ISL_561811, EPI_ISL_561812, EPI_ISL_561813, EPI_ISL_561814, EPI_ISL_561815, EPI_ISL_561817, EPI_ISL_561818, EPI_ISL_561819, EPI_ISL_561820, EPI_ISL_561821, EPI_ISL_561823, EPI_ISL_561824, EPI_ISL_561826, EPI_ISL_561827, EPI_ISL_561828, EPI_ISL_561829, EPI_ISL_561831, EPI_ISL_561832,
EPI_ISL_561833, EPI_ISL_561834, EPI_ISL_561835, EPI_ISL_561836, EPI_ISL_561837, EPI_ISL_561838, EPI_ISL_561839, EPI_ISL_561841, EPI_ISL_561842, EPI_ISL_561845, EPI_ISL_561846, EPI_ISL_561847, EPI_ISL_561848, EPI_ISL_561849, EPI_ISL_561850, EPI_ISL_561851, EPI_ISL_561853, EPI_ISL_561854, EPI_ISL_561857, EPI_ISL_561859, EPI_ISL_561860, EPI_ISL_561862, EPI_ISL_561865, EPI_ISL_561867,
EPI_ISL_561868, EPI_ISL_561869, EPI_ISL_561870, EPI_ISL_561871, EPI_ISL_561872, EPI_ISL_561873, EPI_ISL_561874, EPI_ISL_561875, EPI_ISL_561876, EPI_ISL_561878, EPI_ISL_561880, EPI_ISL_561881, EPI_ISL_561882, EPI_ISL_561884, EPI_ISL_561885, EPI_ISL_561886, EPI_ISL_561887, EPI_ISL_561889, EPI_ISL_561890, EPI_ISL_561891, EPI_ISL_561892, EPI_ISL_561895, EPI_ISL_561896, EPI_ISL_561897,
EPI_ISL_561898, EPI_ISL_561899, EPI_ISL_561900, EPI_ISL_561901, EPI_ISL_561902, EPI_ISL_561905, EPI_ISL_561906, EPI_ISL_561907, EPI_ISL_561908, EPI_ISL_561909, EPI_ISL_561910, EPI_ISL_561912, EPI_ISL_561913, EPI_ISL_561914, EPI_ISL_561915, EPI_ISL_561916, EPI_ISL_561917, EPI_ISL_561918, EPI_ISL_561919, EPI_ISL_561922, EPI_ISL_561923, EPI_ISL_561924, EPI_ISL_561925, EPI_ISL_561926,
EPI_ISL_561927, EPI_ISL_561928, EPI_ISL_561929, EPI_ISL_561930, EPI_ISL_561931, EPI_ISL_561932, EPI_ISL_561933, EPI_ISL_561934, EPI_ISL_561937, EPI_ISL_561938, EPI_ISL_561939, EPI_ISL_561941, EPI_ISL_561943, EPI_ISL_561947, EPI_ISL_561948, EPI_ISL_561949, EPI_ISL_561950, EPI_ISL_561951, EPI_ISL_561952, EPI_ISL_561953, EPI_ISL_561954, EPI_ISL_561956, EPI_ISL_561960, EPI_ISL_561961,
EPI_ISL_561962, EPI_ISL_561963, EPI_ISL_561965, EPI_ISL_561966, EPI_ISL_561968, EPI_ISL_561969, EPI_ISL_561970, EPI_ISL_561971, EPI_ISL_561973, EPI_ISL_561974, EPI_ISL_561976, EPI_ISL_561977, EPI_ISL_561978, EPI_ISL_561979, EPI_ISL_561980, EPI_ISL_561981, EPI_ISL_561982, EPI_ISL_561983, EPI_ISL_561984, EPI_ISL_561985, EPI_ISL_561989, EPI_ISL_561990, EPI_ISL_561992, EPI_ISL_561993,
EPI_ISL_561995, EPI_ISL_561996, EPI_ISL_561997, EPI_ISL_561998, EPI_ISL_561999, EPI_ISL_562000, EPI_ISL_562004, EPI_ISL_562006, EPI_ISL_562008, EPI_ISL_562009, EPI_ISL_562010, EPI_ISL_562014, EPI_ISL_562017, EPI_ISL_562019, EPI_ISL_562020, EPI_ISL_562022, EPI_ISL_562024, EPI_ISL_562025, EPI_ISL_562026, EPI_ISL_562027, EPI_ISL_562028, EPI_ISL_562029, EPI_ISL_562030, EPI_ISL_562031,
EPI_ISL_562032, EPI_ISL_562034, EPI_ISL_562035, EPI_ISL_562036, EPI_ISL_562037, EPI_ISL_562038, EPI_ISL_562039, EPI_ISL_562040, EPI_ISL_562041, EPI_ISL_562042, EPI_ISL_562043, EPI_ISL_562044, EPI_ISL_562045, EPI_ISL_562046, EPI_ISL_562047, EPI_ISL_562049, EPI_ISL_562050, EPI_ISL_562051, EPI_ISL_562052, EPI_ISL_562053, EPI_ISL_562054, EPI_ISL_562058, EPI_ISL_562059, EPI_ISL_562060,
EPI_ISL_562061, EPI_ISL_562062, EPI_ISL_562063, EPI_ISL_562064, EPI_ISL_562065, EPI_ISL_562068, EPI_ISL_562070, EPI_ISL_562071, EPI_ISL_562073, EPI_ISL_562075, EPI_ISL_562077, EPI_ISL_562078, EPI_ISL_562079, EPI_ISL_562082, EPI_ISL_562083, EPI_ISL_562084, EPI_ISL_562086, EPI_ISL_562087, EPI_ISL_562088, EPI_ISL_562089, EPI_ISL_562090, EPI_ISL_562091, EPI_ISL_562092, EPI_ISL_562093,
EPI_ISL_562094, EPI_ISL_562095, EPI_ISL_562096, EPI_ISL_562097, EPI_ISL_562098, EPI_ISL_562099, EPI_ISL_562100, EPI_ISL_562101, EPI_ISL_562102, EPI_ISL_562103, EPI_ISL_562107, EPI_ISL_562108, EPI_ISL_562111, EPI_ISL_562112, EPI_ISL_562113, EPI_ISL_562114, EPI_ISL_562116, EPI_ISL_562117, EPI_ISL_562118, EPI_ISL_562119, EPI_ISL_562120, EPI_ISL_562121, EPI_ISL_562125, EPI_ISL_562126,
EPI_ISL_562127, EPI_ISL_562128, EPI_ISL_562130, EPI_ISL_562131, EPI_ISL_562132, EPI_ISL_562133, EPI_ISL_562134, EPI_ISL_562135, EPI_ISL_562139, EPI_ISL_562140, EPI_ISL_562142, EPI_ISL_562143, EPI_ISL_562144, EPI_ISL_562145, EPI_ISL_562146, EPI_ISL_562147, EPI_ISL_562148, EPI_ISL_562149, EPI_ISL_562150, EPI_ISL_562151, EPI_ISL_562153, EPI_ISL_562154, EPI_ISL_562156, EPI_ISL_562159,
EPI_ISL_562160, EPI_ISL_562161, EPI_ISL_562168, EPI_ISL_562169, EPI_ISL_562170, EPI_ISL_562171, EPI_ISL_562172, EPI_ISL_562173, EPI_ISL_562174, EPI_ISL_562579, EPI_ISL_562591, EPI_ISL_562592, EPI_ISL_562594, EPI_ISL_562596, EPI_ISL_562597, EPI_ISL_562598, EPI_ISL_562599, EPI_ISL_562600, EPI_ISL_562601, EPI_ISL_562602, EPI_ISL_562603, EPI_ISL_562604, EPI_ISL_562605, EPI_ISL_562606,
EPI_ISL_562607, EPI_ISL_562608, EPI_ISL_562609, EPI_ISL_562610, EPI_ISL_562611, EPI_ISL_562612, EPI_ISL_562613, EPI_ISL_562614, EPI_ISL_562615, EPI_ISL_562616, EPI_ISL_562617, EPI_ISL_562618, EPI_ISL_562619, EPI_ISL_562621, EPI_ISL_562622, EPI_ISL_562623, EPI_ISL_562624, EPI_ISL_562625, EPI_ISL_562626, EPI_ISL_562627, EPI_ISL_562629, EPI_ISL_562630, EPI_ISL_562631, EPI_ISL_562632,
EPI_ISL_562633, EPI_ISL_562634, EPI_ISL_562636, EPI_ISL_562637, EPI_ISL_562638, EPI_ISL_562639, EPI_ISL_562641, EPI_ISL_562642, EPI_ISL_562643, EPI_ISL_562644, EPI_ISL_562647, EPI_ISL_562648, EPI_ISL_562649, EPI_ISL_562650, EPI_ISL_562651, EPI_ISL_562652, EPI_ISL_562653, EPI_ISL_562654, EPI_ISL_562655, EPI_ISL_562656, EPI_ISL_562657, EPI_ISL_562660, EPI_ISL_562661, EPI_ISL_562662,
EPI_ISL_562663, EPI_ISL_562664, EPI_ISL_562665, EPI_ISL_562666, EPI_ISL_562667, EPI_ISL_562668, EPI_ISL_562670, EPI_ISL_562672, EPI_ISL_562673, EPI_ISL_562674, EPI_ISL_562675, EPI_ISL_562676, EPI_ISL_562677, EPI_ISL_562678, EPI_ISL_562679, EPI_ISL_562680, EPI_ISL_562681, EPI_ISL_562682, EPI_ISL_562683, EPI_ISL_562684, EPI_ISL_562685, EPI_ISL_562686, EPI_ISL_562687, EPI_ISL_562688,
EPI_ISL_562689, EPI_ISL_562690, EPI_ISL_562691, EPI_ISL_562692, EPI_ISL_562693, EPI_ISL_562694, EPI_ISL_562695, EPI_ISL_562698, EPI_ISL_562699, EPI_ISL_562701, EPI_ISL_562702, EPI_ISL_562703, EPI_ISL_562704, EPI_ISL_562706, EPI_ISL_562707, EPI_ISL_562708, EPI_ISL_562710, EPI_ISL_562711, EPI_ISL_562712, EPI_ISL_562713, EPI_ISL_562714, EPI_ISL_562715, EPI_ISL_562716, EPI_ISL_562717,
EPI_ISL_562718, EPI_ISL_562719, EPI_ISL_562720, EPI_ISL_562721, EPI_ISL_562722, EPI_ISL_562723, EPI_ISL_562724, EPI_ISL_562725, EPI_ISL_562726, EPI_ISL_562727, EPI_ISL_562728, EPI_ISL_562730, EPI_ISL_562731, EPI_ISL_562733, EPI_ISL_562734, EPI_ISL_562735, EPI_ISL_562736, EPI_ISL_562737, EPI_ISL_562738, EPI_ISL_562739, EPI_ISL_562740, EPI_ISL_562741, EPI_ISL_562742, EPI_ISL_562743,
EPI_ISL_562744, EPI_ISL_562745, EPI_ISL_562746, EPI_ISL_562747, EPI_ISL_562748, EPI_ISL_562749, EPI_ISL_562750, EPI_ISL_562751, EPI_ISL_562752, EPI_ISL_562753, EPI_ISL_562755, EPI_ISL_562756, EPI_ISL_562757, EPI_ISL_562758, EPI_ISL_562759, EPI_ISL_562760, EPI_ISL_562761, EPI_ISL_562762, EPI_ISL_562763, EPI_ISL_562764, EPI_ISL_562765, EPI_ISL_562766, EPI_ISL_562767, EPI_ISL_562768,
EPI_ISL_562769, EPI_ISL_562770, EPI_ISL_562771, EPI_ISL_562772, EPI_ISL_562773, EPI_ISL_562774, EPI_ISL_562775, EPI_ISL_562776, EPI_ISL_562777, EPI_ISL_562778, EPI_ISL_562779, EPI_ISL_562780, EPI_ISL_562781, EPI_ISL_562782, EPI_ISL_562783, EPI_ISL_562784, EPI_ISL_562785, EPI_ISL_562786, EPI_ISL_562787, EPI_ISL_562788, EPI_ISL_562789, EPI_ISL_562790, EPI_ISL_562793, EPI_ISL_562794,
EPI_ISL_562795, EPI_ISL_562796, EPI_ISL_562797, EPI_ISL_562798, EPI_ISL_562799, EPI_ISL_562800, EPI_ISL_562801, EPI_ISL_562802, EPI_ISL_562810, EPI_ISL_562812, EPI_ISL_562813, EPI_ISL_562814, EPI_ISL_562815, EPI_ISL_562816, EPI_ISL_562817, EPI_ISL_562818, EPI_ISL_562819, EPI_ISL_562820, EPI_ISL_562821, EPI_ISL_562822, EPI_ISL_562823, EPI_ISL_562824, EPI_ISL_562825, EPI_ISL_562826,
EPI_ISL_562827, EPI_ISL_562828, EPI_ISL_562829, EPI_ISL_562830, EPI_ISL_562831, EPI_ISL_562832, EPI_ISL_562833, EPI_ISL_562834, EPI_ISL_562835, EPI_ISL_562836, EPI_ISL_562837, EPI_ISL_562846, EPI_ISL_562848, EPI_ISL_562849, EPI_ISL_562850, EPI_ISL_562851, EPI_ISL_562852, EPI_ISL_562853, EPI_ISL_562854, EPI_ISL_562855, EPI_ISL_562856, EPI_ISL_562857, EPI_ISL_562858, EPI_ISL_562859,
EPI_ISL_562860, EPI_ISL_562861, EPI_ISL_562862, EPI_ISL_562863, EPI_ISL_562864, EPI_ISL_562865, EPI_ISL_562866, EPI_ISL_562867, EPI_ISL_562868, EPI_ISL_562869, EPI_ISL_562870, EPI_ISL_562871, EPI_ISL_562872, EPI_ISL_562873, EPI_ISL_562874, EPI_ISL_562875, EPI_ISL_562876, EPI_ISL_562877, EPI_ISL_562878, EPI_ISL_562879, EPI_ISL_562881, EPI_ISL_562883, EPI_ISL_562884, EPI_ISL_562885,
EPI_ISL_562886, EPI_ISL_562887, EPI_ISL_562888, EPI_ISL_562889, EPI_ISL_562890, EPI_ISL_562891, EPI_ISL_562892, EPI_ISL_562893, EPI_ISL_562894, EPI_ISL_562895, EPI_ISL_562896, EPI_ISL_562897, EPI_ISL_562898, EPI_ISL_562899, EPI_ISL_562900, EPI_ISL_562901, EPI_ISL_562902, EPI_ISL_562903, EPI_ISL_562904, EPI_ISL_562906, EPI_ISL_562907, EPI_ISL_562908, EPI_ISL_562909, EPI_ISL_562914,
EPI_ISL_562915, EPI_ISL_562916, EPI_ISL_562917, EPI_ISL_562918, EPI_ISL_562919, EPI_ISL_562920, EPI_ISL_562921, EPI_ISL_562922, EPI_ISL_562923, EPI_ISL_562925, EPI_ISL_562926, EPI_ISL_562927, EPI_ISL_562928, EPI_ISL_562930, EPI_ISL_562931, EPI_ISL_562932, EPI_ISL_562933, EPI_ISL_562934, EPI_ISL_562935, EPI_ISL_562938, EPI_ISL_562939, EPI_ISL_562940, EPI_ISL_562941, EPI_ISL_562942,
EPI_ISL_562943, EPI_ISL_562944, EPI_ISL_562945, EPI_ISL_562947, EPI_ISL_562948, EPI_ISL_562949, EPI_ISL_562950, EPI_ISL_562952, EPI_ISL_562953, EPI_ISL_562954, EPI_ISL_562955, EPI_ISL_562956, EPI_ISL_562957, EPI_ISL_562958, EPI_ISL_562959, EPI_ISL_562960, EPI_ISL_562962, EPI_ISL_562963, EPI_ISL_562964, EPI_ISL_562965, EPI_ISL_562966, EPI_ISL_562967, EPI_ISL_562968, EPI_ISL_562969,
EPI_ISL_562970, EPI_ISL_562971, EPI_ISL_562973, EPI_ISL_562974, EPI_ISL_562975, EPI_ISL_562976, EPI_ISL_562977, EPI_ISL_562978, EPI_ISL_562979, EPI_ISL_562980, EPI_ISL_562981, EPI_ISL_562982, EPI_ISL_562983, EPI_ISL_562984, EPI_ISL_562985, EPI_ISL_562986, EPI_ISL_562987, EPI_ISL_562988, EPI_ISL_562989, EPI_ISL_562990, EPI_ISL_562991, EPI_ISL_562992, EPI_ISL_562993, EPI_ISL_562994,
EPI_ISL_562995, EPI_ISL_562996, EPI_ISL_562997, EPI_ISL_562998, EPI_ISL_562999, EPI_ISL_563001, EPI_ISL_563002, EPI_ISL_563003, EPI_ISL_563004, EPI_ISL_563005, EPI_ISL_563006, EPI_ISL_563007, EPI_ISL_563008, EPI_ISL_563009, EPI_ISL_563010, EPI_ISL_563011, EPI_ISL_563012, EPI_ISL_563013, EPI_ISL_563014, EPI_ISL_563015, EPI_ISL_563016, EPI_ISL_563018, EPI_ISL_563019, EPI_ISL_563020,
EPI_ISL_563021, EPI_ISL_563022, EPI_ISL_563023, EPI_ISL_563024, EPI_ISL_563025, EPI_ISL_563026, EPI_ISL_563027, EPI_ISL_563028, EPI_ISL_563029, EPI_ISL_563030, EPI_ISL_563031, EPI_ISL_563032, EPI_ISL_563033, EPI_ISL_563034, EPI_ISL_563035, EPI_ISL_563036, EPI_ISL_563037, EPI_ISL_563038, EPI_ISL_563039, EPI_ISL_563040, EPI_ISL_563041, EPI_ISL_563042, EPI_ISL_563043, EPI_ISL_563044,
EPI_ISL_563045, EPI_ISL_563046, EPI_ISL_563047, EPI_ISL_563048, EPI_ISL_563049, EPI_ISL_563050, EPI_ISL_563051, EPI_ISL_563052, EPI_ISL_563053, EPI_ISL_563054, EPI_ISL_563055, EPI_ISL_563056, EPI_ISL_563057, EPI_ISL_563058, EPI_ISL_563059, EPI_ISL_563060, EPI_ISL_563061, EPI_ISL_563062, EPI_ISL_563063, EPI_ISL_563064, EPI_ISL_563065, EPI_ISL_563066, EPI_ISL_563067, EPI_ISL_563068,
EPI_ISL_563069, EPI_ISL_563070, EPI_ISL_563071, EPI_ISL_563072, EPI_ISL_563073, EPI_ISL_563074, EPI_ISL_563075, EPI_ISL_563076, EPI_ISL_563077, EPI_ISL_563078, EPI_ISL_563079, EPI_ISL_563080, EPI_ISL_563081, EPI_ISL_563082, EPI_ISL_563083, EPI_ISL_563084, EPI_ISL_563085, EPI_ISL_563086, EPI_ISL_563087, EPI_ISL_563088, EPI_ISL_563089, EPI_ISL_563090, EPI_ISL_563091, EPI_ISL_563092,
EPI_ISL_563093, EPI_ISL_563094, EPI_ISL_563095, EPI_ISL_563096, EPI_ISL_563097, EPI_ISL_563098, EPI_ISL_563099, EPI_ISL_563101, EPI_ISL_563102, EPI_ISL_563103, EPI_ISL_563104, EPI_ISL_563105, EPI_ISL_563106, EPI_ISL_563107, EPI_ISL_563108, EPI_ISL_563109, EPI_ISL_563110, EPI_ISL_563111, EPI_ISL_563112, EPI_ISL_563113, EPI_ISL_563114, EPI_ISL_563115, EPI_ISL_563116, EPI_ISL_563117,
EPI_ISL_563118, EPI_ISL_563119, EPI_ISL_563120, EPI_ISL_563121, EPI_ISL_563122, EPI_ISL_563123, EPI_ISL_563124, EPI_ISL_563125, EPI_ISL_563126, EPI_ISL_563127, EPI_ISL_563128, EPI_ISL_563129, EPI_ISL_563130, EPI_ISL_563134, EPI_ISL_563135, EPI_ISL_563145, EPI_ISL_563146, EPI_ISL_563147, EPI_ISL_563148, EPI_ISL_563149, EPI_ISL_563150, EPI_ISL_563151, EPI_ISL_563152, EPI_ISL_563153,
EPI_ISL_563154, EPI_ISL_563155, EPI_ISL_563156, EPI_ISL_563157, EPI_ISL_563158, EPI_ISL_563159, EPI_ISL_563160, EPI_ISL_563161, EPI_ISL_563162, EPI_ISL_563163, EPI_ISL_563164, EPI_ISL_563165, EPI_ISL_563166, EPI_ISL_563167, EPI_ISL_563168, EPI_ISL_563169, EPI_ISL_563170, EPI_ISL_563171, EPI_ISL_563172, EPI_ISL_563173, EPI_ISL_563174, EPI_ISL_563175, EPI_ISL_563176, EPI_ISL_563177,
EPI_ISL_563178, EPI_ISL_563179, EPI_ISL_563180, EPI_ISL_563181, EPI_ISL_563182, EPI_ISL_563183, EPI_ISL_563184, EPI_ISL_563185, EPI_ISL_563186, EPI_ISL_563187, EPI_ISL_563188, EPI_ISL_563189, EPI_ISL_563190, EPI_ISL_563191, EPI_ISL_563192, EPI_ISL_563193, EPI_ISL_563194, EPI_ISL_563195, EPI_ISL_563196, EPI_ISL_563197, EPI_ISL_563198, EPI_ISL_563199, EPI_ISL_563200, EPI_ISL_563201,
EPI_ISL_563202, EPI_ISL_563204, EPI_ISL_563206, EPI_ISL_563207, EPI_ISL_563208, EPI_ISL_563209, EPI_ISL_563210, EPI_ISL_563211, EPI_ISL_563212, EPI_ISL_563213, EPI_ISL_563214, EPI_ISL_563215, EPI_ISL_563216, EPI_ISL_563217, EPI_ISL_563218, EPI_ISL_563219, EPI_ISL_563220, EPI_ISL_563222, EPI_ISL_563223, EPI_ISL_563224, EPI_ISL_563225, EPI_ISL_563226, EPI_ISL_563227, EPI_ISL_563228,
EPI_ISL_563229, EPI_ISL_563230, EPI_ISL_563231, EPI_ISL_563232, EPI_ISL_563233, EPI_ISL_563234, EPI_ISL_563235, EPI_ISL_563236, EPI_ISL_563237, EPI_ISL_563238, EPI_ISL_563239, EPI_ISL_563240, EPI_ISL_563241, EPI_ISL_563242, EPI_ISL_563243, EPI_ISL_563244, EPI_ISL_563245, EPI_ISL_563246, EPI_ISL_563247, EPI_ISL_563248, EPI_ISL_563249, EPI_ISL_563250, EPI_ISL_563251, EPI_ISL_563252,
EPI_ISL_563253, EPI_ISL_563254, EPI_ISL_563255, EPI_ISL_563256, EPI_ISL_563257, EPI_ISL_563258, EPI_ISL_563259, EPI_ISL_563260, EPI_ISL_563261, EPI_ISL_563262, EPI_ISL_563263, EPI_ISL_563264, EPI_ISL_563265, EPI_ISL_563266, EPI_ISL_563267, EPI_ISL_563268, EPI_ISL_563269, EPI_ISL_563270, EPI_ISL_563271, EPI_ISL_563272, EPI_ISL_563273, EPI_ISL_563274, EPI_ISL_563275, EPI_ISL_563276,
EPI_ISL_563277, EPI_ISL_563278, EPI_ISL_563279, EPI_ISL_563280, EPI_ISL_563281, EPI_ISL_563282, EPI_ISL_563283, EPI_ISL_563284, EPI_ISL_563285, EPI_ISL_563286, EPI_ISL_563287, EPI_ISL_563288, EPI_ISL_563289, EPI_ISL_563291, EPI_ISL_563292, EPI_ISL_563293, EPI_ISL_563294, EPI_ISL_563295, EPI_ISL_563296, EPI_ISL_563297, EPI_ISL_563298, EPI_ISL_563299, EPI_ISL_563300, EPI_ISL_563301,
EPI_ISL_563302, EPI_ISL_563303, EPI_ISL_563304, EPI_ISL_563305, EPI_ISL_563306, EPI_ISL_563307, EPI_ISL_563308, EPI_ISL_563309, EPI_ISL_563310, EPI_ISL_563311, EPI_ISL_563312, EPI_ISL_563313, EPI_ISL_563314, EPI_ISL_563315, EPI_ISL_563316, EPI_ISL_563317, EPI_ISL_563318, EPI_ISL_563319, EPI_ISL_563320, EPI_ISL_563321, EPI_ISL_563322, EPI_ISL_563323, EPI_ISL_563324, EPI_ISL_563325,
EPI_ISL_563326, EPI_ISL_563327, EPI_ISL_563328, EPI_ISL_563329, EPI_ISL_563331, EPI_ISL_563332, EPI_ISL_563333, EPI_ISL_563334, EPI_ISL_563335, EPI_ISL_563336, EPI_ISL_563337, EPI_ISL_563338, EPI_ISL_563339, EPI_ISL_563340, EPI_ISL_563341, EPI_ISL_563342, EPI_ISL_563343, EPI_ISL_563344, EPI_ISL_563345, EPI_ISL_563346, EPI_ISL_563347, EPI_ISL_563348, EPI_ISL_563349, EPI_ISL_563350,
EPI_ISL_563351, EPI_ISL_563352, EPI_ISL_563353, EPI_ISL_563354, EPI_ISL_563355, EPI_ISL_563356, EPI_ISL_563357, EPI_ISL_563358, EPI_ISL_563359, EPI_ISL_563360, EPI_ISL_563361, EPI_ISL_563362, EPI_ISL_563363, EPI_ISL_563364, EPI_ISL_563365, EPI_ISL_563366, EPI_ISL_563367, EPI_ISL_563368, EPI_ISL_563369, EPI_ISL_563370, EPI_ISL_563371, EPI_ISL_563372, EPI_ISL_563373, EPI_ISL_563374,
EPI_ISL_563375, EPI_ISL_563376, EPI_ISL_563377, EPI_ISL_563378, EPI_ISL_563379, EPI_ISL_563380, EPI_ISL_563381, EPI_ISL_563382, EPI_ISL_563384, EPI_ISL_563385, EPI_ISL_563386, EPI_ISL_563387, EPI_ISL_563388, EPI_ISL_563389, EPI_ISL_563390, EPI_ISL_563391, EPI_ISL_563392, EPI_ISL_563393, EPI_ISL_563394, EPI_ISL_563395, EPI_ISL_563396, EPI_ISL_563397, EPI_ISL_563398, EPI_ISL_563399,
EPI_ISL_563400, EPI_ISL_563401, EPI_ISL_563402, EPI_ISL_563403, EPI_ISL_563404, EPI_ISL_563405, EPI_ISL_563406, EPI_ISL_563424, EPI_ISL_563425, EPI_ISL_563426, EPI_ISL_563427, EPI_ISL_563428, EPI_ISL_563429, EPI_ISL_563430, EPI_ISL_563431, EPI_ISL_563432, EPI_ISL_563433, EPI_ISL_563434, EPI_ISL_563435, EPI_ISL_563436, EPI_ISL_563437, EPI_ISL_563438, EPI_ISL_563439, EPI_ISL_563440,
EPI_ISL_563441, EPI_ISL_563442, EPI_ISL_563443, EPI_ISL_563444, EPI_ISL_563445, EPI_ISL_563446, EPI_ISL_563447, EPI_ISL_563448, EPI_ISL_563449, EPI_ISL_563450, EPI_ISL_563451, EPI_ISL_563452, EPI_ISL_563453, EPI_ISL_563454, EPI_ISL_563455, EPI_ISL_563456, EPI_ISL_563457, EPI_ISL_563458, EPI_ISL_563459, EPI_ISL_563460, EPI_ISL_563461, EPI_ISL_563462, EPI_ISL_563463, EPI_ISL_563464,
EPI_ISL_563465, EPI_ISL_563466, EPI_ISL_563467, EPI_ISL_563468, EPI_ISL_563469, EPI_ISL_563470, EPI_ISL_563471, EPI_ISL_563473, EPI_ISL_563474, EPI_ISL_563475, EPI_ISL_563476, EPI_ISL_563477, EPI_ISL_563478, EPI_ISL_563479, EPI_ISL_563480, EPI_ISL_563481, EPI_ISL_563482, EPI_ISL_563483, EPI_ISL_563484, EPI_ISL_563485, EPI_ISL_563486, EPI_ISL_563487, EPI_ISL_563488, EPI_ISL_563489,
EPI_ISL_563490, EPI_ISL_563491, EPI_ISL_563492, EPI_ISL_563493, EPI_ISL_563494, EPI_ISL_563495, EPI_ISL_563496, EPI_ISL_563500, EPI_ISL_563501, EPI_ISL_563502, EPI_ISL_563503, EPI_ISL_563504, EPI_ISL_563505, EPI_ISL_563506, EPI_ISL_563507, EPI_ISL_563508, EPI_ISL_563509, EPI_ISL_563510, EPI_ISL_563511, EPI_ISL_563513, EPI_ISL_563514, EPI_ISL_563515, EPI_ISL_563516, EPI_ISL_563517,
EPI_ISL_563518, EPI_ISL_563519, EPI_ISL_563520, EPI_ISL_563521, EPI_ISL_563522, EPI_ISL_563524, EPI_ISL_563525, EPI_ISL_563526, EPI_ISL_563527, EPI_ISL_563528, EPI_ISL_563529, EPI_ISL_563530, EPI_ISL_563531, EPI_ISL_563532, EPI_ISL_563533, EPI_ISL_563534, EPI_ISL_563535, EPI_ISL_563536, EPI_ISL_563537, EPI_ISL_563538, EPI_ISL_563539, EPI_ISL_563540, EPI_ISL_563541, EPI_ISL_563542,
EPI_ISL_563543, EPI_ISL_563544, EPI_ISL_563546, EPI_ISL_563547, EPI_ISL_563548, EPI_ISL_563550, EPI_ISL_563552, EPI_ISL_563553, EPI_ISL_563554, EPI_ISL_563555, EPI_ISL_563556, EPI_ISL_563557, EPI_ISL_563558, EPI_ISL_563559, EPI_ISL_563560, EPI_ISL_563561, EPI_ISL_563562, EPI_ISL_563563, EPI_ISL_563564, EPI_ISL_563565, EPI_ISL_563566, EPI_ISL_563567, EPI_ISL_563568, EPI_ISL_563569,
EPI_ISL_563570, EPI_ISL_563571, EPI_ISL_563572, EPI_ISL_563573, EPI_ISL_563574, EPI_ISL_563575, EPI_ISL_563576, EPI_ISL_563577, EPI_ISL_563578, EPI_ISL_563579, EPI_ISL_563581, EPI_ISL_563582, EPI_ISL_563583, EPI_ISL_563584, EPI_ISL_563585, EPI_ISL_563586, EPI_ISL_563587, EPI_ISL_563588, EPI_ISL_563589, EPI_ISL_563590, EPI_ISL_563591, EPI_ISL_563592, EPI_ISL_563593, EPI_ISL_563594,
EPI_ISL_563595, EPI_ISL_563596, EPI_ISL_563597, EPI_ISL_563598, EPI_ISL_563599, EPI_ISL_563600, EPI_ISL_563601, EPI_ISL_563602, EPI_ISL_563603, EPI_ISL_563604, EPI_ISL_563605, EPI_ISL_563606, EPI_ISL_563607, EPI_ISL_563608, EPI_ISL_563610, EPI_ISL_563611, EPI_ISL_563612, EPI_ISL_563613, EPI_ISL_563614, EPI_ISL_563615, EPI_ISL_563616, EPI_ISL_563617, EPI_ISL_563619, EPI_ISL_563621,
EPI_ISL_563622, EPI_ISL_563623, EPI_ISL_563624, EPI_ISL_563625, EPI_ISL_563626, EPI_ISL_563627, EPI_ISL_563628, EPI_ISL_563629, EPI_ISL_563630, EPI_ISL_563631, EPI_ISL_563632, EPI_ISL_563633, EPI_ISL_563634, EPI_ISL_563635, EPI_ISL_563636, EPI_ISL_563637, EPI_ISL_563639, EPI_ISL_563640, EPI_ISL_563641, EPI_ISL_563642, EPI_ISL_563643, EPI_ISL_563644, EPI_ISL_563645, EPI_ISL_563646,
EPI_ISL_563647, EPI_ISL_563648, EPI_ISL_563649, EPI_ISL_563650, EPI_ISL_563651, EPI_ISL_563652, EPI_ISL_563653, EPI_ISL_563654, EPI_ISL_563655, EPI_ISL_563656, EPI_ISL_563657, EPI_ISL_563659, EPI_ISL_563660, EPI_ISL_563661, EPI_ISL_563662, EPI_ISL_563663, EPI_ISL_563664, EPI_ISL_563665, EPI_ISL_563666, EPI_ISL_563667, EPI_ISL_563668, EPI_ISL_563669, EPI_ISL_563670, EPI_ISL_563671,
EPI_ISL_563672, EPI_ISL_563673, EPI_ISL_563674, EPI_ISL_563675, EPI_ISL_563676, EPI_ISL_563677, EPI_ISL_563678, EPI_ISL_563679, EPI_ISL_563680, EPI_ISL_563681, EPI_ISL_563682, EPI_ISL_563683, EPI_ISL_563684, EPI_ISL_563685, EPI_ISL_563686, EPI_ISL_563687, EPI_ISL_563688, EPI_ISL_563689, EPI_ISL_563691, EPI_ISL_563692, EPI_ISL_563693, EPI_ISL_563694, EPI_ISL_563695, EPI_ISL_563696,
EPI_ISL_563697, EPI_ISL_563698, EPI_ISL_563699, EPI_ISL_563700, EPI_ISL_563701, EPI_ISL_563702, EPI_ISL_563703, EPI_ISL_563704, EPI_ISL_563705, EPI_ISL_563706, EPI_ISL_563707, EPI_ISL_563708, EPI_ISL_563709, EPI_ISL_563710, EPI_ISL_563711, EPI_ISL_563712, EPI_ISL_563713, EPI_ISL_563714, EPI_ISL_563715, EPI_ISL_563716, EPI_ISL_563717, EPI_ISL_563718, EPI_ISL_563719, EPI_ISL_563720,
EPI_ISL_563724, EPI_ISL_563725, EPI_ISL_563726, EPI_ISL_563727, EPI_ISL_563728, EPI_ISL_563729, EPI_ISL_563730, EPI_ISL_563731, EPI_ISL_563732, EPI_ISL_563733, EPI_ISL_563734, EPI_ISL_563735, EPI_ISL_563737, EPI_ISL_563739, EPI_ISL_563740, EPI_ISL_563741, EPI_ISL_563742, EPI_ISL_563743, EPI_ISL_563744, EPI_ISL_563745, EPI_ISL_563746, EPI_ISL_563747, EPI_ISL_563748, EPI_ISL_563749,
EPI_ISL_563750, EPI_ISL_563751, EPI_ISL_563752, EPI_ISL_563753, EPI_ISL_563754, EPI_ISL_563755, EPI_ISL_563756, EPI_ISL_563757, EPI_ISL_563758, EPI_ISL_563759, EPI_ISL_563760, EPI_ISL_563761, EPI_ISL_563762, EPI_ISL_563763, EPI_ISL_563764, EPI_ISL_563765, EPI_ISL_563766, EPI_ISL_563767, EPI_ISL_563768, EPI_ISL_563769, EPI_ISL_563770, EPI_ISL_563771, EPI_ISL_563772, EPI_ISL_563773,
EPI_ISL_563774, EPI_ISL_563775, EPI_ISL_563776, EPI_ISL_563777, EPI_ISL_563778, EPI_ISL_563779, EPI_ISL_563780, EPI_ISL_563781, EPI_ISL_563782, EPI_ISL_563783, EPI_ISL_563784, EPI_ISL_563785, EPI_ISL_563786, EPI_ISL_563787, EPI_ISL_563788, EPI_ISL_563789, EPI_ISL_563791, EPI_ISL_563793, EPI_ISL_563794, EPI_ISL_563795, EPI_ISL_563796, EPI_ISL_563797, EPI_ISL_563798, EPI_ISL_563799,
EPI_ISL_563800, EPI_ISL_563801, EPI_ISL_563802, EPI_ISL_563803, EPI_ISL_563804, EPI_ISL_563805, EPI_ISL_563806, EPI_ISL_563807, EPI_ISL_563809, EPI_ISL_563810, EPI_ISL_563812, EPI_ISL_563813, EPI_ISL_563814, EPI_ISL_563815, EPI_ISL_563816, EPI_ISL_563818, EPI_ISL_563819, EPI_ISL_563821, EPI_ISL_563822, EPI_ISL_563823, EPI_ISL_563824, EPI_ISL_563825, EPI_ISL_563826, EPI_ISL_563827,
EPI_ISL_563828, EPI_ISL_563829, EPI_ISL_563830, EPI_ISL_563831, EPI_ISL_563833, EPI_ISL_563834, EPI_ISL_563835, EPI_ISL_563836, EPI_ISL_563837, EPI_ISL_563838, EPI_ISL_563839, EPI_ISL_563842, EPI_ISL_563843, EPI_ISL_563844, EPI_ISL_563845, EPI_ISL_563846, EPI_ISL_563847, EPI_ISL_563848, EPI_ISL_563849, EPI_ISL_563850, EPI_ISL_563851, EPI_ISL_563852, EPI_ISL_563853, EPI_ISL_563854,
EPI_ISL_563855, EPI_ISL_563856, EPI_ISL_563857, EPI_ISL_563858, EPI_ISL_563859, EPI_ISL_563860, EPI_ISL_563861, EPI_ISL_563862, EPI_ISL_563863, EPI_ISL_563864, EPI_ISL_563865, EPI_ISL_563867, EPI_ISL_563868, EPI_ISL_563869, EPI_ISL_563870, EPI_ISL_563871, EPI_ISL_563872, EPI_ISL_563873, EPI_ISL_563874, EPI_ISL_563875, EPI_ISL_563876, EPI_ISL_563877, EPI_ISL_563878, EPI_ISL_563879,
EPI_ISL_563881, EPI_ISL_563882, EPI_ISL_563884, EPI_ISL_563885, EPI_ISL_563886, EPI_ISL_563887, EPI_ISL_563888, EPI_ISL_563889, EPI_ISL_563890, EPI_ISL_563891, EPI_ISL_563892, EPI_ISL_563895, EPI_ISL_563896, EPI_ISL_563897, EPI_ISL_563898, EPI_ISL_563899, EPI_ISL_563900, EPI_ISL_563901, EPI_ISL_563902, EPI_ISL_563903, EPI_ISL_563904, EPI_ISL_563905, EPI_ISL_563906, EPI_ISL_563908,
EPI_ISL_563909, EPI_ISL_563910, EPI_ISL_563911, EPI_ISL_563912, EPI_ISL_563913, EPI_ISL_563914, EPI_ISL_563915, EPI_ISL_563916, EPI_ISL_563917, EPI_ISL_563918, EPI_ISL_563919, EPI_ISL_563920, EPI_ISL_563921, EPI_ISL_563922, EPI_ISL_563923, EPI_ISL_563924, EPI_ISL_563926, EPI_ISL_563927, EPI_ISL_563928, EPI_ISL_563930, EPI_ISL_563932, EPI_ISL_563933, EPI_ISL_563935, EPI_ISL_563936,
EPI_ISL_563937, EPI_ISL_563939, EPI_ISL_563940, EPI_ISL_563941, EPI_ISL_563942, EPI_ISL_563943, EPI_ISL_563944, EPI_ISL_563945, EPI_ISL_563946, EPI_ISL_563947, EPI_ISL_563948, EPI_ISL_563949, EPI_ISL_563950, EPI_ISL_563951, EPI_ISL_563952, EPI_ISL_563953, EPI_ISL_563956, EPI_ISL_563957, EPI_ISL_563958, EPI_ISL_563959, EPI_ISL_563960, EPI_ISL_563961, EPI_ISL_563962, EPI_ISL_563963,
EPI_ISL_563964, EPI_ISL_563965, EPI_ISL_563966, EPI_ISL_563967, EPI_ISL_563968, EPI_ISL_563969, EPI_ISL_563970, EPI_ISL_563971, EPI_ISL_563972, EPI_ISL_563976, EPI_ISL_563984, EPI_ISL_563991, EPI_ISL_563993, EPI_ISL_563994, EPI_ISL_564050, EPI_ISL_564051, EPI_ISL_564052, EPI_ISL_564053, EPI_ISL_564054, EPI_ISL_564111, EPI_ISL_564112, EPI_ISL_564113, EPI_ISL_564114, EPI_ISL_564115,
EPI_ISL_564116, EPI_ISL_564117, EPI_ISL_564118, EPI_ISL_564120, EPI_ISL_564121, EPI_ISL_564122, EPI_ISL_564123, EPI_ISL_564124, EPI_ISL_564126, EPI_ISL_564127, EPI_ISL_564128, EPI_ISL_564129, EPI_ISL_564130, EPI_ISL_564131, EPI_ISL_564132, EPI_ISL_564133, EPI_ISL_564134, EPI_ISL_564135, EPI_ISL_564136, EPI_ISL_564137, EPI_ISL_564138, EPI_ISL_564139, EPI_ISL_564140, EPI_ISL_564141,
EPI_ISL_564142, EPI_ISL_564143, EPI_ISL_564144, EPI_ISL_564145, EPI_ISL_564146, EPI_ISL_564147, EPI_ISL_564148, EPI_ISL_564149, EPI_ISL_564150, EPI_ISL_564151, EPI_ISL_564152, EPI_ISL_564154, EPI_ISL_564155, EPI_ISL_564156, EPI_ISL_564157, EPI_ISL_564158, EPI_ISL_564159, EPI_ISL_564160, EPI_ISL_564161, EPI_ISL_564162, EPI_ISL_564163, EPI_ISL_564164, EPI_ISL_564165, EPI_ISL_564166,
EPI_ISL_564167, EPI_ISL_564168, EPI_ISL_564169, EPI_ISL_564170, EPI_ISL_564171, EPI_ISL_564172, EPI_ISL_564173, EPI_ISL_564174, EPI_ISL_564175, EPI_ISL_564176, EPI_ISL_564177, EPI_ISL_564179, EPI_ISL_564255, EPI_ISL_564256, EPI_ISL_564257, EPI_ISL_564258, EPI_ISL_564259, EPI_ISL_564260, EPI_ISL_564261, EPI_ISL_564262, EPI_ISL_564263, EPI_ISL_564264, EPI_ISL_564265,
EPI_ISL_564266, EPI_ISL_564267, EPI_ISL_564268, EPI_ISL_564271, EPI_ISL_564272, EPI_ISL_564273, EPI_ISL_564274, EPI_ISL_564275, EPI_ISL_564276, EPI_ISL_564277, EPI_ISL_564278, EPI_ISL_564279, EPI_ISL_564280, EPI_ISL_564281, EPI_ISL_564282, EPI_ISL_564283, EPI_ISL_564284, EPI_ISL_564285, EPI_ISL_564286, EPI_ISL_564287, EPI_ISL_564288, EPI_ISL_564289, EPI_ISL_564290, EPI_ISL_564291,
EPI_ISL_564292, EPI_ISL_564293, EPI_ISL_564294, EPI_ISL_564295, EPI_ISL_564296, EPI_ISL_564297, EPI_ISL_564299, EPI_ISL_564300, EPI_ISL_564301, EPI_ISL_564302, EPI_ISL_564303, EPI_ISL_564304, EPI_ISL_564305, EPI_ISL_564306, EPI_ISL_564307, EPI_ISL_564308, EPI_ISL_564309, EPI_ISL_564310, EPI_ISL_564311, EPI_ISL_564312, EPI_ISL_564313, EPI_ISL_564314, EPI_ISL_564315, EPI_ISL_564316,
EPI_ISL_564317, EPI_ISL_564351, EPI_ISL_564398, EPI_ISL_564399, EPI_ISL_564400, EPI_ISL_564401, EPI_ISL_564402, EPI_ISL_564403, EPI_ISL_564405, EPI_ISL_564406, EPI_ISL_564407, EPI_ISL_564408, EPI_ISL_564409, EPI_ISL_564410, EPI_ISL_564411, EPI_ISL_564412, EPI_ISL_564413, EPI_ISL_564414, EPI_ISL_564415, EPI_ISL_564416, EPI_ISL_564417, EPI_ISL_564418, EPI_ISL_564420, EPI_ISL_564421,
EPI_ISL_564422, EPI_ISL_564423, EPI_ISL_564424, EPI_ISL_564425, EPI_ISL_564426, EPI_ISL_564428, EPI_ISL_564429, EPI_ISL_564430, EPI_ISL_564431, EPI_ISL_564432, EPI_ISL_564433, EPI_ISL_564434, EPI_ISL_564435, EPI_ISL_564436, EPI_ISL_564437, EPI_ISL_564438, EPI_ISL_564439, EPI_ISL_564440, EPI_ISL_564441, EPI_ISL_564442, EPI_ISL_564443, EPI_ISL_564444, EPI_ISL_564445, EPI_ISL_564446,
EPI_ISL_564447, EPI_ISL_564448, EPI_ISL_564449, EPI_ISL_564450, EPI_ISL_564451, EPI_ISL_564452, EPI_ISL_564453, EPI_ISL_564454, EPI_ISL_564455, EPI_ISL_564456, EPI_ISL_564457, EPI_ISL_564459, EPI_ISL_564460, EPI_ISL_564461, EPI_ISL_564463, EPI_ISL_564464, EPI_ISL_564466, EPI_ISL_564467, EPI_ISL_564468, EPI_ISL_564469, EPI_ISL_564470, EPI_ISL_564471, EPI_ISL_564472, EPI_ISL_564473,
EPI_ISL_564474, EPI_ISL_564475, EPI_ISL_564476, EPI_ISL_564477, EPI_ISL_564478, EPI_ISL_564479, EPI_ISL_564480, EPI_ISL_564481, EPI_ISL_564482, EPI_ISL_564483, EPI_ISL_564484, EPI_ISL_564485, EPI_ISL_564486, EPI_ISL_564487, EPI_ISL_564488, EPI_ISL_564489, EPI_ISL_564490, EPI_ISL_564491, EPI_ISL_564492, EPI_ISL_564493, EPI_ISL_564494, EPI_ISL_564495, EPI_ISL_564496, EPI_ISL_564497,
EPI_ISL_564498, EPI_ISL_564499, EPI_ISL_564500, EPI_ISL_564501, EPI_ISL_564502, EPI_ISL_564503, EPI_ISL_564505, EPI_ISL_564506, EPI_ISL_564507, EPI_ISL_564508, EPI_ISL_564509, EPI_ISL_564511, EPI_ISL_564512, EPI_ISL_564513, EPI_ISL_564514, EPI_ISL_564515, EPI_ISL_564516, EPI_ISL_564517, EPI_ISL_564518, EPI_ISL_564519, EPI_ISL_564520, EPI_ISL_564521, EPI_ISL_564522, EPI_ISL_564523,
EPI_ISL_564524, EPI_ISL_564525, EPI_ISL_564526, EPI_ISL_564527, EPI_ISL_564528, EPI_ISL_564529, EPI_ISL_564530, EPI_ISL_564531, EPI_ISL_564532, EPI_ISL_564533, EPI_ISL_564534, EPI_ISL_564535, EPI_ISL_564536, EPI_ISL_564537, EPI_ISL_564538, EPI_ISL_564539, EPI_ISL_564540, EPI_ISL_564541, EPI_ISL_564542, EPI_ISL_564543, EPI_ISL_564545, EPI_ISL_564565, EPI_ISL_564566, EPI_ISL_564567,
EPI_ISL_564568, EPI_ISL_564569, EPI_ISL_564570, EPI_ISL_564571, EPI_ISL_564572, EPI_ISL_564573, EPI_ISL_564574, EPI_ISL_564575, EPI_ISL_564576, EPI_ISL_564577, EPI_ISL_564578, EPI_ISL_564579, EPI_ISL_564580, EPI_ISL_564581, EPI_ISL_564582, EPI_ISL_564583, EPI_ISL_564584, EPI_ISL_564585, EPI_ISL_564586, EPI_ISL_564587, EPI_ISL_564588, EPI_ISL_564589, EPI_ISL_564590, EPI_ISL_564591,
EPI_ISL_564592, EPI_ISL_564593, EPI_ISL_564594, EPI_ISL_564595, EPI_ISL_564596, EPI_ISL_564597, EPI_ISL_564598, EPI_ISL_564599, EPI_ISL_564600, EPI_ISL_564601, EPI_ISL_564602, EPI_ISL_564603, EPI_ISL_564604, EPI_ISL_564605, EPI_ISL_564606, EPI_ISL_564607, EPI_ISL_564608, EPI_ISL_564609, EPI_ISL_564610, EPI_ISL_564611, EPI_ISL_564613, EPI_ISL_564614, EPI_ISL_564615, EPI_ISL_564616,
EPI_ISL_564617, EPI_ISL_564618, EPI_ISL_564619, EPI_ISL_564620, EPI_ISL_564621, EPI_ISL_564622, EPI_ISL_564623, EPI_ISL_564624, EPI_ISL_564626, EPI_ISL_564627, EPI_ISL_564628, EPI_ISL_564629, EPI_ISL_564630, EPI_ISL_564631, EPI_ISL_564632, EPI_ISL_564634, EPI_ISL_564635, EPI_ISL_564636, EPI_ISL_564637, EPI_ISL_564638, EPI_ISL_564639, EPI_ISL_564640, EPI_ISL_564641, EPI_ISL_564642,
EPI_ISL_564643, EPI_ISL_564644, EPI_ISL_564646, EPI_ISL_564647, EPI_ISL_564648, EPI_ISL_564649, EPI_ISL_564650, EPI_ISL_564651, EPI_ISL_564652, EPI_ISL_564653, EPI_ISL_564655, EPI_ISL_564656, EPI_ISL_564657, EPI_ISL_564658, EPI_ISL_564659, EPI_ISL_564660, EPI_ISL_564661, EPI_ISL_564662, EPI_ISL_564663, EPI_ISL_564664, EPI_ISL_564665, EPI_ISL_564666, EPI_ISL_564667, EPI_ISL_564668,
EPI_ISL_564669, EPI_ISL_564670, EPI_ISL_564671, EPI_ISL_564672, EPI_ISL_564673, EPI_ISL_564674, EPI_ISL_564675, EPI_ISL_564676, EPI_ISL_564677, EPI_ISL_564678, EPI_ISL_564679, EPI_ISL_564680, EPI_ISL_564681, EPI_ISL_564682, EPI_ISL_564683, EPI_ISL_564684, EPI_ISL_564685, EPI_ISL_564686, EPI_ISL_564687, EPI_ISL_564688, EPI_ISL_564689, EPI_ISL_564690, EPI_ISL_564691, EPI_ISL_564692,
EPI_ISL_564693, EPI_ISL_564694, EPI_ISL_564695, EPI_ISL_564696, EPI_ISL_564697, EPI_ISL_564698, EPI_ISL_564699, EPI_ISL_564700, EPI_ISL_564701, EPI_ISL_564702, EPI_ISL_564703, EPI_ISL_564704, EPI_ISL_564705, EPI_ISL_564706, EPI_ISL_564707, EPI_ISL_564708, EPI_ISL_564709, EPI_ISL_564710, EPI_ISL_564711, EPI_ISL_564712, EPI_ISL_564713, EPI_ISL_564714, EPI_ISL_564715, EPI_ISL_564716,
EPI_ISL_564717, EPI_ISL_564718, EPI_ISL_564719, EPI_ISL_564720, EPI_ISL_564721, EPI_ISL_564722, EPI_ISL_564723, EPI_ISL_564724, EPI_ISL_564725, EPI_ISL_564726, EPI_ISL_564727, EPI_ISL_564728, EPI_ISL_564729, EPI_ISL_564730, EPI_ISL_564732, EPI_ISL_564733, EPI_ISL_564735, EPI_ISL_564736, EPI_ISL_564737, EPI_ISL_564738, EPI_ISL_564739, EPI_ISL_564740, EPI_ISL_564741, EPI_ISL_564742,
EPI_ISL_564743, EPI_ISL_564744, EPI_ISL_564745, EPI_ISL_564746, EPI_ISL_564748, EPI_ISL_564749, EPI_ISL_564750, EPI_ISL_564751, EPI_ISL_564752, EPI_ISL_564753, EPI_ISL_564754, EPI_ISL_564755, EPI_ISL_564756, EPI_ISL_564757, EPI_ISL_564758, EPI_ISL_564759, EPI_ISL_564760, EPI_ISL_564761, EPI_ISL_564762, EPI_ISL_564763, EPI_ISL_564764, EPI_ISL_564765, EPI_ISL_564766, EPI_ISL_564767,
EPI_ISL_564768, EPI_ISL_564769, EPI_ISL_564770, EPI_ISL_564771, EPI_ISL_564772, EPI_ISL_564773, EPI_ISL_564774, EPI_ISL_564775, EPI_ISL_564776, EPI_ISL_564777, EPI_ISL_564778, EPI_ISL_564779, EPI_ISL_564780, EPI_ISL_564781, EPI_ISL_564782, EPI_ISL_564783, EPI_ISL_564784, EPI_ISL_564785, EPI_ISL_564786, EPI_ISL_564787, EPI_ISL_564788, EPI_ISL_564789, EPI_ISL_564790, EPI_ISL_564791,
EPI_ISL_564792, EPI_ISL_564793, EPI_ISL_564794, EPI_ISL_564795, EPI_ISL_564797, EPI_ISL_564798, EPI_ISL_564799, EPI_ISL_564800, EPI_ISL_564801, EPI_ISL_564802, EPI_ISL_564804, EPI_ISL_564805, EPI_ISL_564806, EPI_ISL_564807, EPI_ISL_564808, EPI_ISL_564809, EPI_ISL_564810, EPI_ISL_564811, EPI_ISL_564812, EPI_ISL_564813, EPI_ISL_564814, EPI_ISL_564815, EPI_ISL_564816, EPI_ISL_564817,
EPI_ISL_564818, EPI_ISL_564819, EPI_ISL_564821, EPI_ISL_564822, EPI_ISL_564823, EPI_ISL_564824, EPI_ISL_564825, EPI_ISL_564826, EPI_ISL_564827, EPI_ISL_564830, EPI_ISL_564832, EPI_ISL_564833, EPI_ISL_564835, EPI_ISL_564836, EPI_ISL_564837, EPI_ISL_564838, EPI_ISL_564839, EPI_ISL_564840, EPI_ISL_564841, EPI_ISL_564842, EPI_ISL_564843, EPI_ISL_564844, EPI_ISL_564845, EPI_ISL_564846,
EPI_ISL_564848, EPI_ISL_564849, EPI_ISL_564850, EPI_ISL_564851, EPI_ISL_564852, EPI_ISL_564853, EPI_ISL_564854, EPI_ISL_564855, EPI_ISL_564856, EPI_ISL_564857, EPI_ISL_564858, EPI_ISL_564859, EPI_ISL_564860, EPI_ISL_564861, EPI_ISL_564862, EPI_ISL_564863, EPI_ISL_564864, EPI_ISL_564865, EPI_ISL_564866, EPI_ISL_564867, EPI_ISL_564868, EPI_ISL_564869, EPI_ISL_564870, EPI_ISL_564871,
EPI_ISL_564872, EPI_ISL_564873, EPI_ISL_564874, EPI_ISL_564875, EPI_ISL_564876, EPI_ISL_564877, EPI_ISL_564878, EPI_ISL_564879, EPI_ISL_564880, EPI_ISL_564881, EPI_ISL_564882, EPI_ISL_564883, EPI_ISL_564885, EPI_ISL_564886, EPI_ISL_564887, EPI_ISL_564888, EPI_ISL_564889, EPI_ISL_564890, EPI_ISL_564891, EPI_ISL_564892, EPI_ISL_564893, EPI_ISL_564894, EPI_ISL_564895, EPI_ISL_564896,
EPI_ISL_564897, EPI_ISL_564898, EPI_ISL_564899, EPI_ISL_564901, EPI_ISL_564902, EPI_ISL_564903, EPI_ISL_564904, EPI_ISL_564905, EPI_ISL_564906, EPI_ISL_564907, EPI_ISL_564908, EPI_ISL_564909, EPI_ISL_564910, EPI_ISL_564911, EPI_ISL_564912, EPI_ISL_564913, EPI_ISL_564915, EPI_ISL_564916, EPI_ISL_564917, EPI_ISL_564918, EPI_ISL_564919, EPI_ISL_564920, EPI_ISL_564921, EPI_ISL_564922,

EPI_ISL_564923, EPI_ISL_564925, EPI_ISL_564927, EPI_ISL_564928, EPI_ISL_564929, EPI_ISL_564930, EPI_ISL_564931, EPI_ISL_564932, EPI_ISL_564933, EPI_ISL_564934, EPI_ISL_564935, EPI_ISL_564936, EPI_ISL_564937, EPI_ISL_564938, EPI_ISL_564939, EPI_ISL_564940, EPI_ISL_564942, EPI_ISL_564943, EPI_ISL_564944, EPI_ISL_564945, EPI_ISL_564946, EPI_ISL_564947, EPI_ISL_564948, EPI_ISL_564949,
EPI_ISL_564950, EPI_ISL_564951, EPI_ISL_564952, EPI_ISL_564953, EPI_ISL_564954, EPI_ISL_564955, EPI_ISL_564956, EPI_ISL_564957, EPI_ISL_564958, EPI_ISL_564959, EPI_ISL_564960, EPI_ISL_564961, EPI_ISL_564962, EPI_ISL_564963, EPI_ISL_564964, EPI_ISL_564965, EPI_ISL_564966, EPI_ISL_564967, EPI_ISL_564968, EPI_ISL_564969, EPI_ISL_564970, EPI_ISL_564971, EPI_ISL_564972, EPI_ISL_564973,
EPI_ISL_564974, EPI_ISL_564975, EPI_ISL_564976, EPI_ISL_564977, EPI_ISL_564978, EPI_ISL_564979, EPI_ISL_564980, EPI_ISL_564981, EPI_ISL_564982, EPI_ISL_564983, EPI_ISL_564984, EPI_ISL_564985, EPI_ISL_564986, EPI_ISL_564987, EPI_ISL_564988, EPI_ISL_564989, EPI_ISL_564990, EPI_ISL_564991, EPI_ISL_564992, EPI_ISL_564993, EPI_ISL_564994, EPI_ISL_564995, EPI_ISL_564996, EPI_ISL_564997,
EPI_ISL_564998, EPI_ISL_564999, EPI_ISL_565000, EPI_ISL_565001, EPI_ISL_565002, EPI_ISL_565003, EPI_ISL_565004, EPI_ISL_565005, EPI_ISL_565006, EPI_ISL_565007, EPI_ISL_565008, EPI_ISL_565009, EPI_ISL_565010, EPI_ISL_565011, EPI_ISL_565012, EPI_ISL_565013, EPI_ISL_565014, EPI_ISL_565015, EPI_ISL_565016, EPI_ISL_565017, EPI_ISL_565018, EPI_ISL_565019, EPI_ISL_565020, EPI_ISL_565021,
EPI_ISL_565022, EPI_ISL_565023, EPI_ISL_565024, EPI_ISL_565025, EPI_ISL_565026, EPI_ISL_565027, EPI_ISL_565028, EPI_ISL_565029, EPI_ISL_565030, EPI_ISL_565031, EPI_ISL_565032, EPI_ISL_565033, EPI_ISL_565034, EPI_ISL_565035, EPI_ISL_565036, EPI_ISL_565037, EPI_ISL_565038, EPI_ISL_565039, EPI_ISL_565040, EPI_ISL_565041, EPI_ISL_565042, EPI_ISL_565043, EPI_ISL_565044, EPI_ISL_565045,
EPI_ISL_565046, EPI_ISL_565048, EPI_ISL_565049, EPI_ISL_565051, EPI_ISL_565052, EPI_ISL_565053, EPI_ISL_565054, EPI_ISL_565055, EPI_ISL_565056, EPI_ISL_565057, EPI_ISL_565058, EPI_ISL_565059, EPI_ISL_565060, EPI_ISL_565061, EPI_ISL_565062, EPI_ISL_565063, EPI_ISL_565064, EPI_ISL_565065, EPI_ISL_565066, EPI_ISL_565067, EPI_ISL_565068, EPI_ISL_565069, EPI_ISL_565070, EPI_ISL_565071,
EPI_ISL_565072, EPI_ISL_565073, EPI_ISL_565074, EPI_ISL_565075, EPI_ISL_565076, EPI_ISL_565077, EPI_ISL_565078, EPI_ISL_565079, EPI_ISL_565080, EPI_ISL_565081, EPI_ISL_565082, EPI_ISL_565083, EPI_ISL_565084, EPI_ISL_565085, EPI_ISL_565086, EPI_ISL_565087, EPI_ISL_565088, EPI_ISL_565089, EPI_ISL_565090, EPI_ISL_565091, EPI_ISL_565092, EPI_ISL_565093, EPI_ISL_565094, EPI_ISL_565096,
EPI_ISL_565097, EPI_ISL_565099, EPI_ISL_565100, EPI_ISL_565101, EPI_ISL_565102, EPI_ISL_565103, EPI_ISL_565104, EPI_ISL_565105, EPI_ISL_565106, EPI_ISL_565107, EPI_ISL_565108, EPI_ISL_565109, EPI_ISL_565110, EPI_ISL_565111, EPI_ISL_565112, EPI_ISL_565113, EPI_ISL_565114, EPI_ISL_565115, EPI_ISL_565116, EPI_ISL_565117, EPI_ISL_565118, EPI_ISL_565119, EPI_ISL_565120, EPI_ISL_565122,
EPI_ISL_565123, EPI_ISL_565124, EPI_ISL_565125, EPI_ISL_565126, EPI_ISL_565127, EPI_ISL_565128, EPI_ISL_565129, EPI_ISL_565130, EPI_ISL_565131, EPI_ISL_565132, EPI_ISL_565133, EPI_ISL_565134, EPI_ISL_565135, EPI_ISL_565223, EPI_ISL_565224, EPI_ISL_565225, EPI_ISL_565226, EPI_ISL_565227, EPI_ISL_565228, EPI_ISL_565229, EPI_ISL_565230, EPI_ISL_565231, EPI_ISL_565232, EPI_ISL_565233,
EPI_ISL_565234, EPI_ISL_565235, EPI_ISL_565236, EPI_ISL_565237, EPI_ISL_565238, EPI_ISL_565239, EPI_ISL_565240, EPI_ISL_565241, EPI_ISL_565242, EPI_ISL_565243, EPI_ISL_565245, EPI_ISL_565246, EPI_ISL_565248, EPI_ISL_565249, EPI_ISL_565250, EPI_ISL_565251, EPI_ISL_565252, EPI_ISL_565253, EPI_ISL_565254, EPI_ISL_565255, EPI_ISL_565256, EPI_ISL_565257, EPI_ISL_565258, EPI_ISL_565259,
EPI_ISL_565260, EPI_ISL_565261, EPI_ISL_565262, EPI_ISL_565263, EPI_ISL_565264, EPI_ISL_565266, EPI_ISL_565267, EPI_ISL_565268, EPI_ISL_565269, EPI_ISL_565270, EPI_ISL_565271, EPI_ISL_565272, EPI_ISL_565273, EPI_ISL_565274, EPI_ISL_565275, EPI_ISL_565276, EPI_ISL_565277, EPI_ISL_565278, EPI_ISL_565279, EPI_ISL_565280, EPI_ISL_565281, EPI_ISL_565282, EPI_ISL_565283, EPI_ISL_565284,
EPI_ISL_565285, EPI_ISL_565286, EPI_ISL_565287, EPI_ISL_565288, EPI_ISL_565290, EPI_ISL_565291, EPI_ISL_565292, EPI_ISL_565293, EPI_ISL_565294, EPI_ISL_565295, EPI_ISL_565296, EPI_ISL_565297, EPI_ISL_565298, EPI_ISL_565299, EPI_ISL_565300, EPI_ISL_565301, EPI_ISL_565302, EPI_ISL_565303, EPI_ISL_565304, EPI_ISL_565305, EPI_ISL_565306, EPI_ISL_565307, EPI_ISL_565308, EPI_ISL_565309,
EPI_ISL_565310, EPI_ISL_565312, EPI_ISL_565313, EPI_ISL_565314, EPI_ISL_565315, EPI_ISL_565316, EPI_ISL_565317, EPI_ISL_565318, EPI_ISL_565319, EPI_ISL_565320, EPI_ISL_565321, EPI_ISL_565323, EPI_ISL_565324, EPI_ISL_565325, EPI_ISL_565327, EPI_ISL_565328, EPI_ISL_565329, EPI_ISL_565330, EPI_ISL_565331, EPI_ISL_565332, EPI_ISL_565333, EPI_ISL_565334, EPI_ISL_565335, EPI_ISL_565336,
EPI_ISL_565337, EPI_ISL_565338, EPI_ISL_565339, EPI_ISL_565340, EPI_ISL_565341, EPI_ISL_565342, EPI_ISL_565343, EPI_ISL_565344, EPI_ISL_565345, EPI_ISL_565346, EPI_ISL_565347, EPI_ISL_565348, EPI_ISL_565349, EPI_ISL_565350, EPI_ISL_565351, EPI_ISL_565355, EPI_ISL_565356, EPI_ISL_565357, EPI_ISL_565358, EPI_ISL_565359, EPI_ISL_565360, EPI_ISL_565361, EPI_ISL_565362, EPI_ISL_565363,
EPI_ISL_565365, EPI_ISL_565366, EPI_ISL_565367, EPI_ISL_565368, EPI_ISL_565369, EPI_ISL_565370, EPI_ISL_565371, EPI_ISL_565372, EPI_ISL_565373, EPI_ISL_565374, EPI_ISL_565375, EPI_ISL_565376, EPI_ISL_565377, EPI_ISL_565378, EPI_ISL_565379, EPI_ISL_565380, EPI_ISL_565381, EPI_ISL_565383, EPI_ISL_565384, EPI_ISL_565386, EPI_ISL_565387, EPI_ISL_565389, EPI_ISL_565390, EPI_ISL_565393,
EPI_ISL_565397, EPI_ISL_565398, EPI_ISL_565400, EPI_ISL_565405, EPI_ISL_565406, EPI_ISL_565407, EPI_ISL_565408, EPI_ISL_565410, EPI_ISL_565411, EPI_ISL_565412, EPI_ISL_565413, EPI_ISL_565414, EPI_ISL_565417, EPI_ISL_565418, EPI_ISL_565419, EPI_ISL_565420, EPI_ISL_565421, EPI_ISL_565422, EPI_ISL_565423, EPI_ISL_565424, EPI_ISL_565425, EPI_ISL_565426, EPI_ISL_565427, EPI_ISL_565428,
EPI_ISL_565429, EPI_ISL_565430, EPI_ISL_565431, EPI_ISL_565432, EPI_ISL_565433, EPI_ISL_565434, EPI_ISL_565435, EPI_ISL_565436, EPI_ISL_565437, EPI_ISL_565438, EPI_ISL_565439, EPI_ISL_565440, EPI_ISL_565441, EPI_ISL_565442, EPI_ISL_565443, EPI_ISL_565444, EPI_ISL_565445, EPI_ISL_565446, EPI_ISL_565447, EPI_ISL_565448, EPI_ISL_565449, EPI_ISL_565450, EPI_ISL_565452, EPI_ISL_565453,
EPI_ISL_565454, EPI_ISL_565455, EPI_ISL_565456, EPI_ISL_565457, EPI_ISL_565458, EPI_ISL_565460, EPI_ISL_565462, EPI_ISL_565468, EPI_ISL_565470, EPI_ISL_565471, EPI_ISL_565472, EPI_ISL_565473, EPI_ISL_565474, EPI_ISL_565475, EPI_ISL_565476, EPI_ISL_565477, EPI_ISL_565478, EPI_ISL_565479, EPI_ISL_565480, EPI_ISL_565481, EPI_ISL_565482, EPI_ISL_565483, EPI_ISL_565484, EPI_ISL_565485,
EPI_ISL_565486, EPI_ISL_565487, EPI_ISL_565488, EPI_ISL_565489, EPI_ISL_565490, EPI_ISL_565491, EPI_ISL_565492, EPI_ISL_565493, EPI_ISL_565494, EPI_ISL_565495, EPI_ISL_565496, EPI_ISL_565497, EPI_ISL_565498, EPI_ISL_565499, EPI_ISL_565500, EPI_ISL_565501, EPI_ISL_565502, EPI_ISL_565503, EPI_ISL_565504, EPI_ISL_565505, EPI_ISL_565506, EPI_ISL_565507, EPI_ISL_565508, EPI_ISL_565510,
EPI_ISL_565512, EPI_ISL_565513, EPI_ISL_565514, EPI_ISL_565515, EPI_ISL_565516, EPI_ISL_565517, EPI_ISL_565518, EPI_ISL_565519, EPI_ISL_565520, EPI_ISL_565521, EPI_ISL_565522, EPI_ISL_565523, EPI_ISL_565524, EPI_ISL_565525, EPI_ISL_565526, EPI_ISL_565527, EPI_ISL_565528, EPI_ISL_565529, EPI_ISL_565530, EPI_ISL_565532, EPI_ISL_565533, EPI_ISL_565534, EPI_ISL_565535, EPI_ISL_565536,
EPI_ISL_565537, EPI_ISL_565538, EPI_ISL_565539, EPI_ISL_565540, EPI_ISL_565541, EPI_ISL_565542, EPI_ISL_565543, EPI_ISL_565545, EPI_ISL_565546, EPI_ISL_565547, EPI_ISL_565548, EPI_ISL_565549, EPI_ISL_565550, EPI_ISL_565551, EPI_ISL_565552, EPI_ISL_565553, EPI_ISL_565554, EPI_ISL_565555, EPI_ISL_565556, EPI_ISL_565557, EPI_ISL_565558, EPI_ISL_565559, EPI_ISL_565560, EPI_ISL_565561,
EPI_ISL_565562, EPI_ISL_565563, EPI_ISL_565564, EPI_ISL_565566, EPI_ISL_565567, EPI_ISL_565568, EPI_ISL_565569, EPI_ISL_565570, EPI_ISL_565571, EPI_ISL_565572, EPI_ISL_565573, EPI_ISL_565574, EPI_ISL_565575, EPI_ISL_565576, EPI_ISL_565577, EPI_ISL_565578, EPI_ISL_565579, EPI_ISL_565580, EPI_ISL_565581, EPI_ISL_565582, EPI_ISL_565583, EPI_ISL_565584, EPI_ISL_565585, EPI_ISL_565586,
EPI_ISL_565587, EPI_ISL_565588, EPI_ISL_565589, EPI_ISL_565590, EPI_ISL_565591, EPI_ISL_565592, EPI_ISL_565593, EPI_ISL_565594, EPI_ISL_565595, EPI_ISL_565596, EPI_ISL_565597, EPI_ISL_565598, EPI_ISL_565599, EPI_ISL_565600, EPI_ISL_565601, EPI_ISL_565602, EPI_ISL_565603, EPI_ISL_565604, EPI_ISL_565605, EPI_ISL_565606, EPI_ISL_565607, EPI_ISL_565608, EPI_ISL_565609, EPI_ISL_565610,
EPI_ISL_565611, EPI_ISL_565612, EPI_ISL_565613, EPI_ISL_565614, EPI_ISL_565615, EPI_ISL_565616, EPI_ISL_565618, EPI_ISL_565619, EPI_ISL_565620, EPI_ISL_565621, EPI_ISL_565622, EPI_ISL_565624, EPI_ISL_565625, EPI_ISL_565626, EPI_ISL_565627, EPI_ISL_565628, EPI_ISL_565629, EPI_ISL_565630, EPI_ISL_565631, EPI_ISL_565633, EPI_ISL_565634, EPI_ISL_565635, EPI_ISL_565636, EPI_ISL_565637,
EPI_ISL_565638, EPI_ISL_565639, EPI_ISL_565640, EPI_ISL_565641, EPI_ISL_565642, EPI_ISL_565643, EPI_ISL_565644, EPI_ISL_565645, EPI_ISL_565646, EPI_ISL_565647, EPI_ISL_565648, EPI_ISL_565649, EPI_ISL_565650, EPI_ISL_565651, EPI_ISL_565652, EPI_ISL_565653, EPI_ISL_565654, EPI_ISL_565655, EPI_ISL_565656, EPI_ISL_565657, EPI_ISL_565658, EPI_ISL_565659, EPI_ISL_565660, EPI_ISL_565661,
EPI_ISL_565662, EPI_ISL_565663, EPI_ISL_565664, EPI_ISL_565665, EPI_ISL_565666, EPI_ISL_565667, EPI_ISL_565668, EPI_ISL_565669, EPI_ISL_565671, EPI_ISL_565672, EPI_ISL_565673, EPI_ISL_565674, EPI_ISL_565675, EPI_ISL_565676, EPI_ISL_565677, EPI_ISL_565678, EPI_ISL_565679, EPI_ISL_565682, EPI_ISL_565683, EPI_ISL_565684, EPI_ISL_565685, EPI_ISL_565686, EPI_ISL_565687, EPI_ISL_565688,
EPI_ISL_565689, EPI_ISL_565690, EPI_ISL_565691, EPI_ISL_565692, EPI_ISL_565693, EPI_ISL_565694, EPI_ISL_565695, EPI_ISL_565696, EPI_ISL_565697, EPI_ISL_565698, EPI_ISL_565699, EPI_ISL_565700, EPI_ISL_565701, EPI_ISL_565702, EPI_ISL_565703, EPI_ISL_565704, EPI_ISL_565705, EPI_ISL_565706, EPI_ISL_565707, EPI_ISL_565708, EPI_ISL_565709, EPI_ISL_565710, EPI_ISL_565711, EPI_ISL_565712,
EPI_ISL_565713, EPI_ISL_565714, EPI_ISL_565715, EPI_ISL_565716, EPI_ISL_565717, EPI_ISL_565718, EPI_ISL_565719, EPI_ISL_565720, EPI_ISL_565721, EPI_ISL_565722, EPI_ISL_565723, EPI_ISL_565724, EPI_ISL_565727, EPI_ISL_565729, EPI_ISL_565743, EPI_ISL_565744, EPI_ISL_565745, EPI_ISL_565746, EPI_ISL_565747, EPI_ISL_565748, EPI_ISL_565749, EPI_ISL_565750, EPI_ISL_565751, EPI_ISL_565752,
EPI_ISL_565753, EPI_ISL_565754, EPI_ISL_565755, EPI_ISL_565756, EPI_ISL_565757, EPI_ISL_565758, EPI_ISL_565759, EPI_ISL_565760, EPI_ISL_565761, EPI_ISL_565762, EPI_ISL_565763, EPI_ISL_565764, EPI_ISL_565765, EPI_ISL_565766, EPI_ISL_565767, EPI_ISL_565768, EPI_ISL_565769, EPI_ISL_565770, EPI_ISL_565771, EPI_ISL_565772, EPI_ISL_565773, EPI_ISL_565774, EPI_ISL_565775, EPI_ISL_565776,
EPI_ISL_565777, EPI_ISL_565779, EPI_ISL_565780, EPI_ISL_565781, EPI_ISL_565782, EPI_ISL_565783, EPI_ISL_565784, EPI_ISL_565785, EPI_ISL_565786, EPI_ISL_565787, EPI_ISL_565788, EPI_ISL_565789, EPI_ISL_565790, EPI_ISL_565791, EPI_ISL_565792, EPI_ISL_565793, EPI_ISL_565794, EPI_ISL_565795, EPI_ISL_565796, EPI_ISL_565797, EPI_ISL_565798, EPI_ISL_565799, EPI_ISL_565800, EPI_ISL_565801,
EPI_ISL_565802, EPI_ISL_565803, EPI_ISL_565804, EPI_ISL_565805, EPI_ISL_565806, EPI_ISL_565807, EPI_ISL_565808, EPI_ISL_565809, EPI_ISL_565810, EPI_ISL_565811, EPI_ISL_565812, EPI_ISL_565813, EPI_ISL_565814, EPI_ISL_565815, EPI_ISL_565816, EPI_ISL_565817, EPI_ISL_565818, EPI_ISL_565819, EPI_ISL_565820, EPI_ISL_565821, EPI_ISL_565822, EPI_ISL_565823, EPI_ISL_565824, EPI_ISL_565825,
EPI_ISL_565826, EPI_ISL_565827, EPI_ISL_565828, EPI_ISL_565829, EPI_ISL_591563, EPI_ISL_591564, EPI_ISL_591567, EPI_ISL_591569, EPI_ISL_591572, EPI_ISL_591575, EPI_ISL_591576, EPI_ISL_591579, EPI_ISL_591582, EPI_ISL_591586, EPI_ISL_591588, EPI_ISL_591589, EPI_ISL_591593, EPI_ISL_591598, EPI_ISL_591599, EPI_ISL_591600, EPI_ISL_591601, EPI_ISL_591602, EPI_ISL_591604, EPI_ISL_591605,
EPI_ISL_591606, EPI_ISL_591607, EPI_ISL_591608, EPI_ISL_591610, EPI_ISL_591611, EPI_ISL_591612, EPI_ISL_591614, EPI_ISL_591616, EPI_ISL_591617, EPI_ISL_591618, EPI_ISL_591620, EPI_ISL_591622, EPI_ISL_591624, EPI_ISL_591625, EPI_ISL_591627, EPI_ISL_591628, EPI_ISL_591630, EPI_ISL_591631, EPI_ISL_591632, EPI_ISL_591633, EPI_ISL_591634, EPI_ISL_591635, EPI_ISL_591636, EPI_ISL_591637,
EPI_ISL_591639, EPI_ISL_591641, EPI_ISL_591642, EPI_ISL_591644, EPI_ISL_591646, EPI_ISL_591648, EPI_ISL_591649, EPI_ISL_591650, EPI_ISL_591651, EPI_ISL_591655, EPI_ISL_591658, EPI_ISL_591660, EPI_ISL_591661, EPI_ISL_591663, EPI_ISL_591664, EPI_ISL_591665, EPI_ISL_591668, EPI_ISL_591673, EPI_ISL_591674, EPI_ISL_591675, EPI_ISL_591676, EPI_ISL_591677, EPI_ISL_591678, EPI_ISL_591680,
EPI_ISL_591682, EPI_ISL_591683, EPI_ISL_591685, EPI_ISL_591686, EPI_ISL_591687, EPI_ISL_591689, EPI_ISL_591690, EPI_ISL_591691, EPI_ISL_591692, EPI_ISL_591693, EPI_ISL_591694, EPI_ISL_591695, EPI_ISL_591696, EPI_ISL_591697, EPI_ISL_591699, EPI_ISL_591701, EPI_ISL_591702, EPI_ISL_591705, EPI_ISL_591709, EPI_ISL_591710, EPI_ISL_591711, EPI_ISL_591712, EPI_ISL_591713, EPI_ISL_591714,
EPI_ISL_591716, EPI_ISL_591717, EPI_ISL_591718, EPI_ISL_591793, EPI_ISL_591794, EPI_ISL_591795, EPI_ISL_591796, EPI_ISL_591797, EPI_ISL_591798, EPI_ISL_591801, EPI_ISL_591802, EPI_ISL_591803, EPI_ISL_591804, EPI_ISL_591806, EPI_ISL_591808, EPI_ISL_591809, EPI_ISL_591810, EPI_ISL_591811, EPI_ISL_591812, EPI_ISL_591813, EPI_ISL_591814, EPI_ISL_591815, EPI_ISL_591816, EPI_ISL_591817,
EPI_ISL_591818, EPI_ISL_591819, EPI_ISL_591820, EPI_ISL_591821, EPI_ISL_591823, EPI_ISL_591826, EPI_ISL_591827, EPI_ISL_591828, EPI_ISL_591829, EPI_ISL_591830, EPI_ISL_591831, EPI_ISL_591832, EPI_ISL_591833, EPI_ISL_591834, EPI_ISL_591835, EPI_ISL_591836, EPI_ISL_591837, EPI_ISL_591838, EPI_ISL_591839, EPI_ISL_591840, EPI_ISL_591841, EPI_ISL_591843, EPI_ISL_591844, EPI_ISL_591845,
EPI_ISL_591846, EPI_ISL_591847, EPI_ISL_591848, EPI_ISL_591853, EPI_ISL_591905, EPI_ISL_591906, EPI_ISL_591910, EPI_ISL_591911, EPI_ISL_591913, EPI_ISL_591914, EPI_ISL_591915, EPI_ISL_591916, EPI_ISL_591918, EPI_ISL_591920, EPI_ISL_591921, EPI_ISL_591922, EPI_ISL_591923, EPI_ISL_591924, EPI_ISL_591926, EPI_ISL_591927, EPI_ISL_591929, EPI_ISL_591930, EPI_ISL_591931, EPI_ISL_591933,
EPI_ISL_591934, EPI_ISL_591935, EPI_ISL_591936, EPI_ISL_591937, EPI_ISL_591938, EPI_ISL_591939, EPI_ISL_591961, EPI_ISL_591962, EPI_ISL_591963, EPI_ISL_591964, EPI_ISL_591965, EPI_ISL_591966, EPI_ISL_591967, EPI_ISL_591969, EPI_ISL_591970, EPI_ISL_591971, EPI_ISL_591972, EPI_ISL_591973, EPI_ISL_591974, EPI_ISL_591975, EPI_ISL_591976, EPI_ISL_591977, EPI_ISL_591978, EPI_ISL_591979,
EPI_ISL_591980, EPI_ISL_591981, EPI_ISL_591982, EPI_ISL_591983, EPI_ISL_591984, EPI_ISL_591985, EPI_ISL_591986, EPI_ISL_591987, EPI_ISL_591988, EPI_ISL_591989, EPI_ISL_591990, EPI_ISL_591991, EPI_ISL_591992, EPI_ISL_591993, EPI_ISL_591994, EPI_ISL_591995, EPI_ISL_591996, EPI_ISL_591997, EPI_ISL_591998, EPI_ISL_591999, EPI_ISL_592000, EPI_ISL_592001, EPI_ISL_592002, EPI_ISL_592003,
EPI_ISL_592004, EPI_ISL_592005, EPI_ISL_592006, EPI_ISL_592007, EPI_ISL_592008, EPI_ISL_592009, EPI_ISL_592010, EPI_ISL_592011, EPI_ISL_592012, EPI_ISL_592013, EPI_ISL_592015, EPI_ISL_592016, EPI_ISL_592017, EPI_ISL_592018, EPI_ISL_592019, EPI_ISL_592020, EPI_ISL_592021, EPI_ISL_592022, EPI_ISL_592023, EPI_ISL_592024, EPI_ISL_592025, EPI_ISL_592026, EPI_ISL_592027, EPI_ISL_592028,
EPI_ISL_592029, EPI_ISL_592030, EPI_ISL_592031, EPI_ISL_592032, EPI_ISL_592033, EPI_ISL_592034, EPI_ISL_592035, EPI_ISL_592036, EPI_ISL_592037, EPI_ISL_592038, EPI_ISL_592039, EPI_ISL_592040, EPI_ISL_592041, EPI_ISL_592042, EPI_ISL_592043, EPI_ISL_592045, EPI_ISL_592046, EPI_ISL_592047, EPI_ISL_592048, EPI_ISL_592049, EPI_ISL_592050, EPI_ISL_592051, EPI_ISL_592052, EPI_ISL_592053,
EPI_ISL_592054, EPI_ISL_592055, EPI_ISL_592056, EPI_ISL_592057, EPI_ISL_592058, EPI_ISL_592059, EPI_ISL_592060, EPI_ISL_592061, EPI_ISL_592062, EPI_ISL_592063, EPI_ISL_592065, EPI_ISL_592066, EPI_ISL_592067, EPI_ISL_592068, EPI_ISL_592069, EPI_ISL_592070, EPI_ISL_592071, EPI_ISL_592072, EPI_ISL_592073, EPI_ISL_592074, EPI_ISL_592075, EPI_ISL_592076, EPI_ISL_592077, EPI_ISL_592078,
EPI_ISL_592079, EPI_ISL_592080, EPI_ISL_592081, EPI_ISL_592082, EPI_ISL_592083, EPI_ISL_592084, EPI_ISL_592085, EPI_ISL_592086, EPI_ISL_592087, EPI_ISL_592088, EPI_ISL_592089, EPI_ISL_592090, EPI_ISL_592091, EPI_ISL_592092, EPI_ISL_592093, EPI_ISL_592094, EPI_ISL_592095, EPI_ISL_592096, EPI_ISL_592097, EPI_ISL_592098, EPI_ISL_592099, EPI_ISL_592100, EPI_ISL_592101, EPI_ISL_592102,
EPI_ISL_592103, EPI_ISL_592104, EPI_ISL_592105, EPI_ISL_592106, EPI_ISL_592107, EPI_ISL_592108, EPI_ISL_592109, EPI_ISL_592110, EPI_ISL_592111, EPI_ISL_592112, EPI_ISL_592113, EPI_ISL_592114, EPI_ISL_592115, EPI_ISL_592116, EPI_ISL_592117, EPI_ISL_592118, EPI_ISL_592119, EPI_ISL_592120, EPI_ISL_592121, EPI_ISL_592122, EPI_ISL_592123, EPI_ISL_592124, EPI_ISL_592125, EPI_ISL_592126,
EPI_ISL_592127, EPI_ISL_592128, EPI_ISL_592129, EPI_ISL_592130, EPI_ISL_592131, EPI_ISL_592132, EPI_ISL_592133, EPI_ISL_592134, EPI_ISL_592135, EPI_ISL_592196, EPI_ISL_592197, EPI_ISL_592198, EPI_ISL_592199, EPI_ISL_592200, EPI_ISL_592201, EPI_ISL_592202, EPI_ISL_592203, EPI_ISL_592204, EPI_ISL_592205, EPI_ISL_592206, EPI_ISL_592207, EPI_ISL_592208, EPI_ISL_592209, EPI_ISL_592210,
EPI_ISL_592211, EPI_ISL_592212, EPI_ISL_592213, EPI_ISL_592214, EPI_ISL_592215, EPI_ISL_592216, EPI_ISL_592217, EPI_ISL_592218, EPI_ISL_592219, EPI_ISL_592220, EPI_ISL_592221, EPI_ISL_592222, EPI_ISL_592223, EPI_ISL_592224, EPI_ISL_592225, EPI_ISL_592226, EPI_ISL_592227, EPI_ISL_592228, EPI_ISL_592229, EPI_ISL_592230, EPI_ISL_592231, EPI_ISL_592232, EPI_ISL_592233, EPI_ISL_592234,
EPI_ISL_592236, EPI_ISL_592237, EPI_ISL_592238, EPI_ISL_592239, EPI_ISL_592240, EPI_ISL_592241, EPI_ISL_592242, EPI_ISL_592243, EPI_ISL_592244, EPI_ISL_592245, EPI_ISL_592246, EPI_ISL_592247, EPI_ISL_592248, EPI_ISL_592249, EPI_ISL_592250, EPI_ISL_592251, EPI_ISL_592252, EPI_ISL_592253, EPI_ISL_592254, EPI_ISL_592255, EPI_ISL_592256, EPI_ISL_592257, EPI_ISL_592258, EPI_ISL_592259,
EPI_ISL_592260, EPI_ISL_592261, EPI_ISL_592262, EPI_ISL_592263, EPI_ISL_592264, EPI_ISL_592265, EPI_ISL_592266, EPI_ISL_592267, EPI_ISL_592268, EPI_ISL_592269, EPI_ISL_592270, EPI_ISL_592271, EPI_ISL_592272, EPI_ISL_592273, EPI_ISL_592274, EPI_ISL_592275, EPI_ISL_592276, EPI_ISL_592277, EPI_ISL_592278, EPI_ISL_592279, EPI_ISL_592280, EPI_ISL_592281, EPI_ISL_592282, EPI_ISL_592283,
EPI_ISL_592285, EPI_ISL_592286, EPI_ISL_592287, EPI_ISL_592288, EPI_ISL_592289, EPI_ISL_592290, EPI_ISL_592291, EPI_ISL_592292, EPI_ISL_592293, EPI_ISL_592294, EPI_ISL_592295, EPI_ISL_592296, EPI_ISL_592297, EPI_ISL_592299, EPI_ISL_592300, EPI_ISL_592301, EPI_ISL_592302, EPI_ISL_592303, EPI_ISL_592304, EPI_ISL_592305, EPI_ISL_592306, EPI_ISL_592308, EPI_ISL_592309, EPI_ISL_592310,
EPI_ISL_592311, EPI_ISL_592312, EPI_ISL_592313, EPI_ISL_592314, EPI_ISL_592315, EPI_ISL_592316, EPI_ISL_592317, EPI_ISL_592318, EPI_ISL_592319, EPI_ISL_592320, EPI_ISL_592321, EPI_ISL_592322, EPI_ISL_592323, EPI_ISL_592324, EPI_ISL_592325, EPI_ISL_592326, EPI_ISL_592327, EPI_ISL_592328, EPI_ISL_592329, EPI_ISL_592330, EPI_ISL_592331, EPI_ISL_592332, EPI_ISL_592333, EPI_ISL_592334,
EPI_ISL_592335, EPI_ISL_592336, EPI_ISL_592337, EPI_ISL_592338, EPI_ISL_592339, EPI_ISL_592340, EPI_ISL_592342, EPI_ISL_592343, EPI_ISL_592344, EPI_ISL_592345, EPI_ISL_592346, EPI_ISL_592347, EPI_ISL_592348, EPI_ISL_592349, EPI_ISL_592350, EPI_ISL_592351, EPI_ISL_592352, EPI_ISL_592353, EPI_ISL_592354, EPI_ISL_592355, EPI_ISL_592356, EPI_ISL_592357, EPI_ISL_592358, EPI_ISL_592359,
EPI_ISL_592360, EPI_ISL_592361, EPI_ISL_592362, EPI_ISL_592363, EPI_ISL_592364, EPI_ISL_592365, EPI_ISL_592366, EPI_ISL_592367, EPI_ISL_592368, EPI_ISL_592369, EPI_ISL_592370, EPI_ISL_592371, EPI_ISL_592372, EPI_ISL_592373, EPI_ISL_592374, EPI_ISL_592375, EPI_ISL_592376, EPI_ISL_592377, EPI_ISL_592378, EPI_ISL_592379, EPI_ISL_592380, EPI_ISL_592381, EPI_ISL_592382, EPI_ISL_592383,
EPI_ISL_592384, EPI_ISL_592385, EPI_ISL_592386, EPI_ISL_592387, EPI_ISL_592388, EPI_ISL_592389, EPI_ISL_592390, EPI_ISL_592391, EPI_ISL_592392, EPI_ISL_592393, EPI_ISL_592394, EPI_ISL_592395, EPI_ISL_592396, EPI_ISL_592397, EPI_ISL_592398, EPI_ISL_592399, EPI_ISL_592400, EPI_ISL_592401, EPI_ISL_592402, EPI_ISL_592403, EPI_ISL_592404, EPI_ISL_592405, EPI_ISL_592406, EPI_ISL_592407,
EPI_ISL_592408, EPI_ISL_592409, EPI_ISL_592410, EPI_ISL_592411, EPI_ISL_592412, EPI_ISL_592413, EPI_ISL_592414, EPI_ISL_592415, EPI_ISL_592416, EPI_ISL_592417, EPI_ISL_592418, EPI_ISL_592419, EPI_ISL_592420, EPI_ISL_592421, EPI_ISL_592422, EPI_ISL_592423, EPI_ISL_592424, EPI_ISL_592425, EPI_ISL_592426, EPI_ISL_592427, EPI_ISL_592428, EPI_ISL_592429, EPI_ISL_592430, EPI_ISL_592431,
EPI_ISL_592432, EPI_ISL_592433, EPI_ISL_592434, EPI_ISL_592435, EPI_ISL_592436, EPI_ISL_592437, EPI_ISL_592438, EPI_ISL_592439, EPI_ISL_592440, EPI_ISL_592441, EPI_ISL_592442, EPI_ISL_592443, EPI_ISL_592444, EPI_ISL_592445, EPI_ISL_592446, EPI_ISL_592447, EPI_ISL_592448, EPI_ISL_592449, EPI_ISL_592450, EPI_ISL_592451, EPI_ISL_592452, EPI_ISL_592453, EPI_ISL_592454, EPI_ISL_592455,
EPI_ISL_592456, EPI_ISL_592457, EPI_ISL_592458, EPI_ISL_592459, EPI_ISL_592460, EPI_ISL_592461, EPI_ISL_592462, EPI_ISL_592463, EPI_ISL_592464, EPI_ISL_592465, EPI_ISL_592466, EPI_ISL_592467, EPI_ISL_592468, EPI_ISL_592469, EPI_ISL_592470, EPI_ISL_592471, EPI_ISL_592472, EPI_ISL_592473, EPI_ISL_592474, EPI_ISL_592475, EPI_ISL_592476, EPI_ISL_592477, EPI_ISL_592478, EPI_ISL_592479,
EPI_ISL_592480, EPI_ISL_592481, EPI_ISL_592482, EPI_ISL_592483, EPI_ISL_592484, EPI_ISL_592487, EPI_ISL_592510, EPI_ISL_592517, EPI_ISL_592518, EPI_ISL_592519, EPI_ISL_592520, EPI_ISL_592521, EPI_ISL_592522, EPI_ISL_592523, EPI_ISL_592524, EPI_ISL_592525, EPI_ISL_592526, EPI_ISL_592527, EPI_ISL_592528, EPI_ISL_592529, EPI_ISL_592530, EPI_ISL_592552, EPI_ISL_592556, EPI_ISL_592557,
EPI_ISL_592560, EPI_ISL_592561, EPI_ISL_592565, EPI_ISL_592566, EPI_ISL_592567, EPI_ISL_592569, EPI_ISL_592570, EPI_ISL_592571, EPI_ISL_592572, EPI_ISL_592573, EPI_ISL_592574, EPI_ISL_592575, EPI_ISL_592576, EPI_ISL_592578, EPI_ISL_592579, EPI_ISL_592580, EPI_ISL_592581, EPI_ISL_592582, EPI_ISL_592583, EPI_ISL_592584, EPI_ISL_592585, EPI_ISL_592586, EPI_ISL_592589, EPI_ISL_592591,
EPI_ISL_592592, EPI_ISL_592593, EPI_ISL_592594, EPI_ISL_592595, EPI_ISL_592596, EPI_ISL_592598, EPI_ISL_592599, EPI_ISL_592602, EPI_ISL_592604, EPI_ISL_592606, EPI_ISL_592607, EPI_ISL_592608, EPI_ISL_592609, EPI_ISL_592610, EPI_ISL_592611, EPI_ISL_592613, EPI_ISL_592614, EPI_ISL_592617, EPI_ISL_592618, EPI_ISL_592619, EPI_ISL_592620, EPI_ISL_592621, EPI_ISL_592622, EPI_ISL_592623,
EPI_ISL_592624, EPI_ISL_592625, EPI_ISL_592626, EPI_ISL_592627, EPI_ISL_592628, EPI_ISL_592629, EPI_ISL_592630, EPI_ISL_592631, EPI_ISL_592632, EPI_ISL_592633, EPI_ISL_592634, EPI_ISL_592635, EPI_ISL_592637, EPI_ISL_592638, EPI_ISL_592639, EPI_ISL_592640, EPI_ISL_592641, EPI_ISL_592642, EPI_ISL_592643, EPI_ISL_592644, EPI_ISL_592645, EPI_ISL_592646, EPI_ISL_592647, EPI_ISL_592648,
EPI_ISL_592649, EPI_ISL_592650, EPI_ISL_592651, EPI_ISL_592652, EPI_ISL_592653, EPI_ISL_592654, EPI_ISL_592655, EPI_ISL_592656, EPI_ISL_592657, EPI_ISL_592658, EPI_ISL_592659, EPI_ISL_592662, EPI_ISL_592663, EPI_ISL_592664, EPI_ISL_592665, EPI_ISL_592666, EPI_ISL_592667, EPI_ISL_592669, EPI_ISL_592670, EPI_ISL_592671, EPI_ISL_592672, EPI_ISL_592673, EPI_ISL_592674, EPI_ISL_592675,
EPI_ISL_592676, EPI_ISL_592677, EPI_ISL_592678, EPI_ISL_592679, EPI_ISL_592680, EPI_ISL_592681, EPI_ISL_592682, EPI_ISL_592683, EPI_ISL_592684, EPI_ISL_592685, EPI_ISL_592686, EPI_ISL_592687, EPI_ISL_592688, EPI_ISL_592689, EPI_ISL_592690, EPI_ISL_592691, EPI_ISL_592692, EPI_ISL_592693, EPI_ISL_592694, EPI_ISL_592695, EPI_ISL_592696, EPI_ISL_592697, EPI_ISL_592698, EPI_ISL_592699,
EPI_ISL_592700, EPI_ISL_592701, EPI_ISL_592702, EPI_ISL_592703, EPI_ISL_592704, EPI_ISL_592819, EPI_ISL_592822, EPI_ISL_592823, EPI_ISL_592825, EPI_ISL_592826, EPI_ISL_592829, EPI_ISL_592830, EPI_ISL_592831, EPI_ISL_592833, EPI_ISL_592834, EPI_ISL_592835, EPI_ISL_592838, EPI_ISL_592839, EPI_ISL_592840, EPI_ISL_592841, EPI_ISL_592842, EPI_ISL_592843, EPI_ISL_592844, EPI_ISL_592845,
EPI_ISL_592846, EPI_ISL_592847, EPI_ISL_592848, EPI_ISL_592849, EPI_ISL_592850, EPI_ISL_592851, EPI_ISL_592852, EPI_ISL_592853, EPI_ISL_592854, EPI_ISL_592855, EPI_ISL_592856, EPI_ISL_592857, EPI_ISL_592861, EPI_ISL_592866, EPI_ISL_592867, EPI_ISL_592868, EPI_ISL_592869, EPI_ISL_592870, EPI_ISL_592871, EPI_ISL_592872, EPI_ISL_592873, EPI_ISL_592874, EPI_ISL_592875, EPI_ISL_592876,
EPI_ISL_592877, EPI_ISL_592882, EPI_ISL_592883, EPI_ISL_592884, EPI_ISL_592885, EPI_ISL_592886, EPI_ISL_592887, EPI_ISL_592888, EPI_ISL_592889, EPI_ISL_592890, EPI_ISL_592891, EPI_ISL_592892, EPI_ISL_592893, EPI_ISL_592894, EPI_ISL_592895, EPI_ISL_592896, EPI_ISL_592897, EPI_ISL_592898, EPI_ISL_592899, EPI_ISL_592900, EPI_ISL_592901, EPI_ISL_592902, EPI_ISL_592903, EPI_ISL_592904,
EPI_ISL_592905, EPI_ISL_592906, EPI_ISL_592907, EPI_ISL_592908, EPI_ISL_592909, EPI_ISL_592910, EPI_ISL_592911, EPI_ISL_592912, EPI_ISL_592913, EPI_ISL_592914, EPI_ISL_592915, EPI_ISL_592916, EPI_ISL_592917, EPI_ISL_592918, EPI_ISL_592919, EPI_ISL_592920, EPI_ISL_592921, EPI_ISL_592922, EPI_ISL_592923, EPI_ISL_592924, EPI_ISL_592925, EPI_ISL_592926, EPI_ISL_592927, EPI_ISL_592928,
EPI_ISL_592929, EPI_ISL_592930, EPI_ISL_592931, EPI_ISL_592932, EPI_ISL_592933, EPI_ISL_592934, EPI_ISL_592935, EPI_ISL_592936, EPI_ISL_592937, EPI_ISL_592938, EPI_ISL_592939, EPI_ISL_592940, EPI_ISL_592941, EPI_ISL_592942, EPI_ISL_592943, EPI_ISL_592944, EPI_ISL_592945, EPI_ISL_592946, EPI_ISL_592947, EPI_ISL_592948, EPI_ISL_592949, EPI_ISL_592950, EPI_ISL_592951, EPI_ISL_592952,
EPI_ISL_592953, EPI_ISL_592954, EPI_ISL_592955, EPI_ISL_592956, EPI_ISL_592957, EPI_ISL_592959, EPI_ISL_592960, EPI_ISL_592961, EPI_ISL_592962, EPI_ISL_592963, EPI_ISL_592964, EPI_ISL_592965, EPI_ISL_592966, EPI_ISL_592967, EPI_ISL_592968, EPI_ISL_592986, EPI_ISL_592987, EPI_ISL_592988, EPI_ISL_592989, EPI_ISL_592990, EPI_ISL_592991, EPI_ISL_592992, EPI_ISL_592993, EPI_ISL_592994,
EPI_ISL_592995, EPI_ISL_592996, EPI_ISL_592997, EPI_ISL_592998, EPI_ISL_592999, EPI_ISL_593000, EPI_ISL_593001, EPI_ISL_593002, EPI_ISL_593008, EPI_ISL_593009, EPI_ISL_593010, EPI_ISL_593011, EPI_ISL_593012, EPI_ISL_593014, EPI_ISL_593016, EPI_ISL_593018, EPI_ISL_593019, EPI_ISL_593020, EPI_ISL_593021, EPI_ISL_593022, EPI_ISL_593023, EPI_ISL_593024, EPI_ISL_593025, EPI_ISL_593026,
EPI_ISL_593027, EPI_ISL_593028, EPI_ISL_593029, EPI_ISL_593030, EPI_ISL_593031, EPI_ISL_593034, EPI_ISL_593036, EPI_ISL_593037, EPI_ISL_593040, EPI_ISL_593041, EPI_ISL_593042, EPI_ISL_593043, EPI_ISL_593045, EPI_ISL_593046, EPI_ISL_593047, EPI_ISL_593049, EPI_ISL_593050, EPI_ISL_593051, EPI_ISL_593052, EPI_ISL_593053, EPI_ISL_593054, EPI_ISL_593055, EPI_ISL_593056, EPI_ISL_593057,
EPI_ISL_593058, EPI_ISL_593059, EPI_ISL_593060, EPI_ISL_593061, EPI_ISL_593062, EPI_ISL_593063, EPI_ISL_593064, EPI_ISL_593065, EPI_ISL_593066, EPI_ISL_593067, EPI_ISL_593068, EPI_ISL_593069, EPI_ISL_593070, EPI_ISL_593071, EPI_ISL_593072, EPI_ISL_593073, EPI_ISL_593074, EPI_ISL_593075, EPI_ISL_593076, EPI_ISL_593077, EPI_ISL_593078, EPI_ISL_593079, EPI_ISL_593080, EPI_ISL_593081,
EPI_ISL_593082, EPI_ISL_593083, EPI_ISL_593084, EPI_ISL_593085, EPI_ISL_593086, EPI_ISL_593087, EPI_ISL_593088, EPI_ISL_593089, EPI_ISL_593090, EPI_ISL_593091, EPI_ISL_593092, EPI_ISL_593093, EPI_ISL_593094, EPI_ISL_593095, EPI_ISL_593096, EPI_ISL_593097, EPI_ISL_593098, EPI_ISL_593099, EPI_ISL_593100, EPI_ISL_593101, EPI_ISL_593105, EPI_ISL_593108, EPI_ISL_593110, EPI_ISL_593112,
EPI_ISL_593113, EPI_ISL_593118, EPI_ISL_593124, EPI_ISL_593129, EPI_ISL_593130, EPI_ISL_593131, EPI_ISL_593132, EPI_ISL_593133, EPI_ISL_593134, EPI_ISL_593135, EPI_ISL_593136, EPI_ISL_593137, EPI_ISL_593138, EPI_ISL_593139, EPI_ISL_593140, EPI_ISL_593141, EPI_ISL_593142, EPI_ISL_593143, EPI_ISL_593144, EPI_ISL_593145, EPI_ISL_593146, EPI_ISL_593147, EPI_ISL_593148, EPI_ISL_593149,
EPI_ISL_593150, EPI_ISL_593151, EPI_ISL_593152, EPI_ISL_593153, EPI_ISL_593154, EPI_ISL_593155, EPI_ISL_593156, EPI_ISL_593157, EPI_ISL_593158, EPI_ISL_593159, EPI_ISL_593160, EPI_ISL_593162, EPI_ISL_593163, EPI_ISL_593164, EPI_ISL_593165, EPI_ISL_593166, EPI_ISL_593167, EPI_ISL_593168, EPI_ISL_593169, EPI_ISL_593170, EPI_ISL_593171, EPI_ISL_593172, EPI_ISL_593173, EPI_ISL_593174,
EPI_ISL_593175, EPI_ISL_593176, EPI_ISL_593177, EPI_ISL_593178, EPI_ISL_593179, EPI_ISL_593180, EPI_ISL_593181, EPI_ISL_593182, EPI_ISL_593183, EPI_ISL_593184, EPI_ISL_593185, EPI_ISL_593186, EPI_ISL_593187, EPI_ISL_593190, EPI_ISL_593198, EPI_ISL_593213, EPI_ISL_593218, EPI_ISL_593220, EPI_ISL_593221, EPI_ISL_593222, EPI_ISL_593223, EPI_ISL_593226, EPI_ISL_593227, EPI_ISL_593229,
EPI_ISL_593230, EPI_ISL_593231, EPI_ISL_593234, EPI_ISL_593237, EPI_ISL_593238, EPI_ISL_593239, EPI_ISL_593241, EPI_ISL_593242, EPI_ISL_593244, EPI_ISL_593245, EPI_ISL_593246, EPI_ISL_593247, EPI_ISL_593248, EPI_ISL_593250, EPI_ISL_593251, EPI_ISL_593252, EPI_ISL_593253, EPI_ISL_593254, EPI_ISL_593257, EPI_ISL_593258, EPI_ISL_593259, EPI_ISL_593260, EPI_ISL_593261, EPI_ISL_593262,
EPI_ISL_593263, EPI_ISL_593264, EPI_ISL_593265, EPI_ISL_593266, EPI_ISL_593267, EPI_ISL_593268, EPI_ISL_593269, EPI_ISL_593270, EPI_ISL_593271, EPI_ISL_593272, EPI_ISL_593273, EPI_ISL_593274, EPI_ISL_593275, EPI_ISL_593276, EPI_ISL_593277, EPI_ISL_593278, EPI_ISL_593279, EPI_ISL_593280, EPI_ISL_593281, EPI_ISL_593282, EPI_ISL_593283, EPI_ISL_593284, EPI_ISL_593285, EPI_ISL_593286,
EPI_ISL_593287, EPI_ISL_593288, EPI_ISL_593289, EPI_ISL_593290, EPI_ISL_593291, EPI_ISL_593292, EPI_ISL_593293, EPI_ISL_593295, EPI_ISL_593296, EPI_ISL_593297, EPI_ISL_593298, EPI_ISL_593299, EPI_ISL_593300, EPI_ISL_593301, EPI_ISL_593302, EPI_ISL_593303, EPI_ISL_593304, EPI_ISL_593305, EPI_ISL_593306, EPI_ISL_593307, EPI_ISL_593308, EPI_ISL_593309, EPI_ISL_593310, EPI_ISL_593311,
EPI_ISL_593312, EPI_ISL_593313, EPI_ISL_593314, EPI_ISL_593315, EPI_ISL_593316, EPI_ISL_593317, EPI_ISL_593318, EPI_ISL_593319, EPI_ISL_593320, EPI_ISL_593321, EPI_ISL_593322, EPI_ISL_593323, EPI_ISL_593324, EPI_ISL_593325, EPI_ISL_593326, EPI_ISL_593327, EPI_ISL_593328, EPI_ISL_593329, EPI_ISL_593330, EPI_ISL_593331, EPI_ISL_593332, EPI_ISL_593333, EPI_ISL_593334, EPI_ISL_593335,
EPI_ISL_593336, EPI_ISL_593337, EPI_ISL_593338, EPI_ISL_593339, EPI_ISL_593340, EPI_ISL_593341, EPI_ISL_593342, EPI_ISL_593343, EPI_ISL_593344, EPI_ISL_593345, EPI_ISL_593346, EPI_ISL_593347, EPI_ISL_593348, EPI_ISL_593349, EPI_ISL_593350, EPI_ISL_593351, EPI_ISL_593352, EPI_ISL_593353, EPI_ISL_593354, EPI_ISL_593355, EPI_ISL_593356, EPI_ISL_593357, EPI_ISL_593358, EPI_ISL_593359,
EPI_ISL_593360, EPI_ISL_593361, EPI_ISL_593362, EPI_ISL_593363, EPI_ISL_593368, EPI_ISL_593369, EPI_ISL_593370, EPI_ISL_593371, EPI_ISL_593372, EPI_ISL_593373, EPI_ISL_593374, EPI_ISL_593375, EPI_ISL_593376, EPI_ISL_593377, EPI_ISL_593378, EPI_ISL_593379, EPI_ISL_593380, EPI_ISL_593381, EPI_ISL_593382, EPI_ISL_593383, EPI_ISL_593384, EPI_ISL_593385, EPI_ISL_593386, EPI_ISL_593387,
EPI_ISL_593388, EPI_ISL_593389, EPI_ISL_593390, EPI_ISL_593391, EPI_ISL_593392, EPI_ISL_593393, EPI_ISL_593394, EPI_ISL_593395, EPI_ISL_593396, EPI_ISL_593397, EPI_ISL_593398, EPI_ISL_593399, EPI_ISL_593400, EPI_ISL_593401, EPI_ISL_593402, EPI_ISL_593403, EPI_ISL_593404, EPI_ISL_593405, EPI_ISL_593406, EPI_ISL_593407, EPI_ISL_593408, EPI_ISL_593409, EPI_ISL_593410, EPI_ISL_593413,
EPI_ISL_593414, EPI_ISL_593415, EPI_ISL_593416, EPI_ISL_593417, EPI_ISL_593418, EPI_ISL_593419, EPI_ISL_593420, EPI_ISL_593421, EPI_ISL_593422, EPI_ISL_593423, EPI_ISL_593424, EPI_ISL_593425, EPI_ISL_593426, EPI_ISL_593427, EPI_ISL_593428, EPI_ISL_593429, EPI_ISL_593430, EPI_ISL_593431, EPI_ISL_593432, EPI_ISL_593433, EPI_ISL_593434, EPI_ISL_593435, EPI_ISL_593436, EPI_ISL_593437,
EPI_ISL_593438, EPI_ISL_593440, EPI_ISL_593441, EPI_ISL_593442, EPI_ISL_593443, EPI_ISL_593444, EPI_ISL_593445, EPI_ISL_593446, EPI_ISL_593447, EPI_ISL_593448, EPI_ISL_593449, EPI_ISL_593450, EPI_ISL_593451, EPI_ISL_593452, EPI_ISL_593453, EPI_ISL_593454, EPI_ISL_593456, EPI_ISL_593457, EPI_ISL_593458, EPI_ISL_593459, EPI_ISL_593460, EPI_ISL_593461, EPI_ISL_593462, EPI_ISL_593463,
EPI_ISL_593464, EPI_ISL_593465, EPI_ISL_593467, EPI_ISL_593468, EPI_ISL_593469, EPI_ISL_593470, EPI_ISL_593471, EPI_ISL_593472, EPI_ISL_593473, EPI_ISL_593474, EPI_ISL_593475, EPI_ISL_640301, EPI_ISL_640303, EPI_ISL_640304, EPI_ISL_640305, EPI_ISL_640306, EPI_ISL_640307, EPI_ISL_640308, EPI_ISL_640309, EPI_ISL_640310, EPI_ISL_640312, EPI_ISL_640313, EPI_ISL_640314, EPI_ISL_640315,
EPI_ISL_640316, EPI_ISL_640317, EPI_ISL_640318, EPI_ISL_640320, EPI_ISL_640321, EPI_ISL_640322, EPI_ISL_640323, EPI_ISL_640324, EPI_ISL_640325, EPI_ISL_640327, EPI_ISL_640328, EPI_ISL_640329, EPI_ISL_640331, EPI_ISL_640332, EPI_ISL_640333, EPI_ISL_640334, EPI_ISL_640335, EPI_ISL_640336, EPI_ISL_640338, EPI_ISL_640340, EPI_ISL_640341, EPI_ISL_640343, EPI_ISL_640344, EPI_ISL_640345,
EPI_ISL_640346, EPI_ISL_640347, EPI_ISL_640348, EPI_ISL_640350, EPI_ISL_640351, EPI_ISL_640352, EPI_ISL_640353, EPI_ISL_640354, EPI_ISL_640355, EPI_ISL_640356, EPI_ISL_640357, EPI_ISL_640358, EPI_ISL_640359, EPI_ISL_640360, EPI_ISL_640361, EPI_ISL_640362, EPI_ISL_640363, EPI_ISL_640364, EPI_ISL_640365, EPI_ISL_640367, EPI_ISL_640368, EPI_ISL_640369, EPI_ISL_640370, EPI_ISL_640371,
EPI_ISL_640372, EPI_ISL_640373, EPI_ISL_640374, EPI_ISL_640376, EPI_ISL_640377, EPI_ISL_640378, EPI_ISL_640379, EPI_ISL_640380, EPI_ISL_640381, EPI_ISL_640382, EPI_ISL_640383, EPI_ISL_640384, EPI_ISL_640385, EPI_ISL_640387, EPI_ISL_640388, EPI_ISL_640390, EPI_ISL_640392, EPI_ISL_640394, EPI_ISL_640395, EPI_ISL_640396, EPI_ISL_640397, EPI_ISL_640399, EPI_ISL_640400, EPI_ISL_640401,
EPI_ISL_640402, EPI_ISL_640403, EPI_ISL_640404, EPI_ISL_640405, EPI_ISL_640406, EPI_ISL_640407, EPI_ISL_640408, EPI_ISL_640409, EPI_ISL_640410, EPI_ISL_640411, EPI_ISL_640412, EPI_ISL_640413, EPI_ISL_640414, EPI_ISL_640415, EPI_ISL_640416, EPI_ISL_640417, EPI_ISL_640418, EPI_ISL_640419, EPI_ISL_640420, EPI_ISL_640421, EPI_ISL_640422, EPI_ISL_640424, EPI_ISL_640425, EPI_ISL_640426,
EPI_ISL_640427, EPI_ISL_640428, EPI_ISL_640430, EPI_ISL_640432, EPI_ISL_640433, EPI_ISL_640434, EPI_ISL_640435, EPI_ISL_640436, EPI_ISL_640437, EPI_ISL_640439, EPI_ISL_640440, EPI_ISL_640441, EPI_ISL_640442, EPI_ISL_640443, EPI_ISL_640444, EPI_ISL_640445, EPI_ISL_640446, EPI_ISL_640448, EPI_ISL_640449, EPI_ISL_640450, EPI_ISL_640451, EPI_ISL_640452, EPI_ISL_640454, EPI_ISL_640455,
EPI_ISL_640456, EPI_ISL_640457, EPI_ISL_640458, EPI_ISL_640459, EPI_ISL_640460, EPI_ISL_640461, EPI_ISL_640462, EPI_ISL_640463, EPI_ISL_640464, EPI_ISL_640465, EPI_ISL_640466, EPI_ISL_640467, EPI_ISL_640468, EPI_ISL_640469, EPI_ISL_640470, EPI_ISL_640472, EPI_ISL_640473, EPI_ISL_640474, EPI_ISL_640475, EPI_ISL_640478, EPI_ISL_640479, EPI_ISL_640480, EPI_ISL_640481, EPI_ISL_640482,
EPI_ISL_640484, EPI_ISL_640485, EPI_ISL_640486, EPI_ISL_640487, EPI_ISL_640490, EPI_ISL_640491, EPI_ISL_640494, EPI_ISL_640495, EPI_ISL_640496, EPI_ISL_640497, EPI_ISL_640498, EPI_ISL_640499, EPI_ISL_640500, EPI_ISL_640501, EPI_ISL_640502, EPI_ISL_640503, EPI_ISL_640504, EPI_ISL_640505, EPI_ISL_640506, EPI_ISL_640507, EPI_ISL_640508, EPI_ISL_640510, EPI_ISL_640513, EPI_ISL_640514,
EPI_ISL_640515, EPI_ISL_640516, EPI_ISL_640517, EPI_ISL_640518, EPI_ISL_640520, EPI_ISL_640521, EPI_ISL_640523, EPI_ISL_640524, EPI_ISL_640525, EPI_ISL_640528, EPI_ISL_640529, EPI_ISL_640530, EPI_ISL_640531, EPI_ISL_640532, EPI_ISL_640533, EPI_ISL_640535, EPI_ISL_640536, EPI_ISL_640538, EPI_ISL_640539, EPI_ISL_640540, EPI_ISL_640541, EPI_ISL_640542, EPI_ISL_640544, EPI_ISL_640545,
EPI_ISL_640546, EPI_ISL_640547, EPI_ISL_640548, EPI_ISL_640549, EPI_ISL_640550, EPI_ISL_640551, EPI_ISL_640552, EPI_ISL_640554, EPI_ISL_640555, EPI_ISL_640556, EPI_ISL_640557, EPI_ISL_640558, EPI_ISL_640559, EPI_ISL_640560, EPI_ISL_640561, EPI_ISL_640564, EPI_ISL_640565, EPI_ISL_640566, EPI_ISL_640567, EPI_ISL_640569, EPI_ISL_640570, EPI_ISL_640571, EPI_ISL_640572, EPI_ISL_640573,
EPI_ISL_640574, EPI_ISL_640575, EPI_ISL_640576, EPI_ISL_640577, EPI_ISL_640578, EPI_ISL_640579, EPI_ISL_640580, EPI_ISL_640581, EPI_ISL_640582, EPI_ISL_640584, EPI_ISL_640586, EPI_ISL_640587, EPI_ISL_640589, EPI_ISL_640591, EPI_ISL_640592, EPI_ISL_640593, EPI_ISL_640594, EPI_ISL_640595, EPI_ISL_640596, EPI_ISL_640597, EPI_ISL_640598, EPI_ISL_640599, EPI_ISL_640600, EPI_ISL_640601,
EPI_ISL_640602, EPI_ISL_640603, EPI_ISL_640604, EPI_ISL_640609, EPI_ISL_640610, EPI_ISL_640612, EPI_ISL_640613, EPI_ISL_640614, EPI_ISL_640615, EPI_ISL_640616, EPI_ISL_640617, EPI_ISL_640618, EPI_ISL_640619, EPI_ISL_640620, EPI_ISL_640621, EPI_ISL_640622, EPI_ISL_640624, EPI_ISL_640627, EPI_ISL_640628, EPI_ISL_640629, EPI_ISL_640631, EPI_ISL_640632, EPI_ISL_640633, EPI_ISL_640634,
EPI_ISL_640635, EPI_ISL_640636, EPI_ISL_640637, EPI_ISL_640638, EPI_ISL_640639, EPI_ISL_640640, EPI_ISL_640642, EPI_ISL_640643, EPI_ISL_640644, EPI_ISL_640645, EPI_ISL_640646, EPI_ISL_640647, EPI_ISL_640648, EPI_ISL_640649, EPI_ISL_640650, EPI_ISL_640651, EPI_ISL_640652, EPI_ISL_640653, EPI_ISL_640654, EPI_ISL_640655, EPI_ISL_640656, EPI_ISL_640657, EPI_ISL_640659, EPI_ISL_640660,
EPI_ISL_640661, EPI_ISL_640662, EPI_ISL_640663, EPI_ISL_640664, EPI_ISL_640665, EPI_ISL_640667, EPI_ISL_640668, EPI_ISL_640669, EPI_ISL_640670, EPI_ISL_640671, EPI_ISL_640672, EPI_ISL_640673, EPI_ISL_640675, EPI_ISL_640676, EPI_ISL_640677, EPI_ISL_640678, EPI_ISL_640679, EPI_ISL_640680, EPI_ISL_640683, EPI_ISL_640684, EPI_ISL_640685, EPI_ISL_640686, EPI_ISL_640687, EPI_ISL_640688,
EPI_ISL_640689, EPI_ISL_640691, EPI_ISL_640692, EPI_ISL_640693, EPI_ISL_640694, EPI_ISL_640695, EPI_ISL_640696, EPI_ISL_640697, EPI_ISL_640698, EPI_ISL_640699, EPI_ISL_640702, EPI_ISL_640703, EPI_ISL_640704, EPI_ISL_640705, EPI_ISL_640706, EPI_ISL_640707, EPI_ISL_640708, EPI_ISL_640709, EPI_ISL_640710, EPI_ISL_640711, EPI_ISL_640713, EPI_ISL_640714, EPI_ISL_640715, EPI_ISL_640716,
EPI_ISL_640717, EPI_ISL_640718, EPI_ISL_640719, EPI_ISL_640721, EPI_ISL_640724, EPI_ISL_640725, EPI_ISL_640726, EPI_ISL_640727, EPI_ISL_640728, EPI_ISL_640729, EPI_ISL_640730, EPI_ISL_640731, EPI_ISL_640733, EPI_ISL_640734, EPI_ISL_640735, EPI_ISL_640736, EPI_ISL_640737, EPI_ISL_640738, EPI_ISL_640741, EPI_ISL_640742, EPI_ISL_640743, EPI_ISL_640744, EPI_ISL_640745, EPI_ISL_640746,
EPI_ISL_640747, EPI_ISL_640748, EPI_ISL_640749, EPI_ISL_640750, EPI_ISL_640752, EPI_ISL_640753, EPI_ISL_640754, EPI_ISL_640755, EPI_ISL_640756, EPI_ISL_640757, EPI_ISL_640758, EPI_ISL_640759, EPI_ISL_640760, EPI_ISL_640761, EPI_ISL_640762, EPI_ISL_640764, EPI_ISL_640765, EPI_ISL_640766, EPI_ISL_640767, EPI_ISL_640768, EPI_ISL_640770, EPI_ISL_640771, EPI_ISL_640773, EPI_ISL_640774,
EPI_ISL_640777, EPI_ISL_640778, EPI_ISL_640779, EPI_ISL_640780, EPI_ISL_640781, EPI_ISL_640782, EPI_ISL_640783, EPI_ISL_640785, EPI_ISL_640786, EPI_ISL_640787, EPI_ISL_640788, EPI_ISL_640789, EPI_ISL_640790, EPI_ISL_640791, EPI_ISL_640792, EPI_ISL_640793, EPI_ISL_640794, EPI_ISL_640796, EPI_ISL_640798, EPI_ISL_640799, EPI_ISL_640800, EPI_ISL_640801, EPI_ISL_640802, EPI_ISL_640804,
EPI_ISL_640805, EPI_ISL_640806, EPI_ISL_640807, EPI_ISL_640808, EPI_ISL_640809, EPI_ISL_640810, EPI_ISL_640811, EPI_ISL_640812, EPI_ISL_640813, EPI_ISL_640814, EPI_ISL_640816, EPI_ISL_640817, EPI_ISL_640818, EPI_ISL_640819, EPI_ISL_640820, EPI_ISL_640821, EPI_ISL_640822, EPI_ISL_640823, EPI_ISL_640824, EPI_ISL_640825, EPI_ISL_640826, EPI_ISL_640828, EPI_ISL_640829, EPI_ISL_640830,
EPI_ISL_640831, EPI_ISL_640832, EPI_ISL_640833, EPI_ISL_640834, EPI_ISL_640835, EPI_ISL_640837, EPI_ISL_640838, EPI_ISL_640839, EPI_ISL_640840, EPI_ISL_640841, EPI_ISL_640842, EPI_ISL_640843, EPI_ISL_640844, EPI_ISL_640846, EPI_ISL_640847, EPI_ISL_640849, EPI_ISL_640850, EPI_ISL_640851, EPI_ISL_640852, EPI_ISL_640853, EPI_ISL_640854, EPI_ISL_640855, EPI_ISL_640856, EPI_ISL_640857,
EPI_ISL_640858, EPI_ISL_640859, EPI_ISL_640860, EPI_ISL_640861, EPI_ISL_640862, EPI_ISL_640864, EPI_ISL_640865, EPI_ISL_640866, EPI_ISL_640867, EPI_ISL_640868, EPI_ISL_640869, EPI_ISL_640870, EPI_ISL_640871, EPI_ISL_640872, EPI_ISL_640873, EPI_ISL_640875, EPI_ISL_640876, EPI_ISL_640877, EPI_ISL_640878, EPI_ISL_640879, EPI_ISL_640880, EPI_ISL_640881, EPI_ISL_640882, EPI_ISL_640883,
EPI_ISL_640884, EPI_ISL_640885, EPI_ISL_640886, EPI_ISL_640887, EPI_ISL_640888, EPI_ISL_640889, EPI_ISL_640890, EPI_ISL_640891, EPI_ISL_640892, EPI_ISL_640893, EPI_ISL_640894, EPI_ISL_640895, EPI_ISL_640896, EPI_ISL_640897, EPI_ISL_640899, EPI_ISL_640900, EPI_ISL_640901, EPI_ISL_640902, EPI_ISL_640903, EPI_ISL_640904, EPI_ISL_640905, EPI_ISL_640906, EPI_ISL_640907, EPI_ISL_640908,
EPI_ISL_640910, EPI_ISL_640911, EPI_ISL_640913, EPI_ISL_640914, EPI_ISL_640915, EPI_ISL_640916, EPI_ISL_640918, EPI_ISL_640919, EPI_ISL_640920, EPI_ISL_640921, EPI_ISL_640922, EPI_ISL_640923, EPI_ISL_640924, EPI_ISL_640925, EPI_ISL_640926, EPI_ISL_640927, EPI_ISL_640928, EPI_ISL_640929, EPI_ISL_640930, EPI_ISL_640931, EPI_ISL_640932, EPI_ISL_640933, EPI_ISL_640934, EPI_ISL_640935,
EPI_ISL_640936, EPI_ISL_640937, EPI_ISL_640938, EPI_ISL_640940, EPI_ISL_640941, EPI_ISL_640943, EPI_ISL_640944, EPI_ISL_640945, EPI_ISL_640946, EPI_ISL_640947, EPI_ISL_640948, EPI_ISL_640949, EPI_ISL_640950, EPI_ISL_640951, EPI_ISL_640952, EPI_ISL_640953, EPI_ISL_640954, EPI_ISL_640955, EPI_ISL_640956, EPI_ISL_640957, EPI_ISL_640959, EPI_ISL_640960, EPI_ISL_640961, EPI_ISL_640962,
EPI_ISL_640963, EPI_ISL_640964, EPI_ISL_640965, EPI_ISL_640966, EPI_ISL_640968, EPI_ISL_640969, EPI_ISL_640970, EPI_ISL_640971, EPI_ISL_640972, EPI_ISL_640973, EPI_ISL_640974, EPI_ISL_640975, EPI_ISL_640976, EPI_ISL_640977, EPI_ISL_640978, EPI_ISL_640979, EPI_ISL_640980, EPI_ISL_640983, EPI_ISL_640984, EPI_ISL_640985, EPI_ISL_640987, EPI_ISL_640988, EPI_ISL_640989, EPI_ISL_640990,

| | | | |
|---|---|---|---|
| EPI_ISL_640991, EPI_ISL_640992, EPI_ISL_640994, EPI_ISL_640995, EPI_ISL_640996, EPI_ISL_640997, EPI_ISL_640999, EPI_ISL_641001, EPI_ISL_641002, EPI_ISL_641003, EPI_ISL_641004, EPI_ISL_641005, EPI_ISL_641006, EPI_ISL_641008, EPI_ISL_641009, EPI_ISL_641010, EPI_ISL_641011, EPI_ISL_641013, EPI_ISL_641014, EPI_ISL_641015, EPI_ISL_641016, EPI_ISL_641018, EPI_ISL_641019, EPI_ISL_641020, EPI_ISL_641021, EPI_ISL_641022, EPI_ISL_641023, EPI_ISL_641024, EPI_ISL_641025, EPI_ISL_641026, EPI_ISL_641027, EPI_ISL_641028, EPI_ISL_641029, EPI_ISL_641030, EPI_ISL_641031, EPI_ISL_641032, EPI_ISL_641034, EPI_ISL_641035, EPI_ISL_641036, EPI_ISL_641037, EPI_ISL_641038, EPI_ISL_641040, EPI_ISL_641041, EPI_ISL_641042, EPI_ISL_641043, EPI_ISL_641045, EPI_ISL_641046, EPI_ISL_641047, EPI_ISL_641048, EPI_ISL_641050, EPI_ISL_641051, EPI_ISL_641052, EPI_ISL_641053, EPI_ISL_641054, EPI_ISL_641055, EPI_ISL_641056, EPI_ISL_641057, EPI_ISL_641058, EPI_ISL_641060, EPI_ISL_641061, EPI_ISL_641062, EPI_ISL_641063, EPI_ISL_641064, EPI_ISL_641065, EPI_ISL_641066, EPI_ISL_641068, EPI_ISL_641070, EPI_ISL_641071, EPI_ISL_641072, EPI_ISL_641073, EPI_ISL_641074, EPI_ISL_641077, EPI_ISL_641078, EPI_ISL_641079, EPI_ISL_641080, EPI_ISL_641081, EPI_ISL_641082, EPI_ISL_641083, EPI_ISL_641084, EPI_ISL_641085, EPI_ISL_641087, EPI_ISL_641089, EPI_ISL_641090, EPI_ISL_641091, EPI_ISL_641092, EPI_ISL_641093, EPI_ISL_641094, EPI_ISL_641095, EPI_ISL_641096, EPI_ISL_641098, EPI_ISL_641099, EPI_ISL_641100, EPI_ISL_641101, EPI_ISL_641103, EPI_ISL_641104, EPI_ISL_641105, EPI_ISL_641107, EPI_ISL_641108, EPI_ISL_641109, EPI_ISL_641111, EPI_ISL_641112, EPI_ISL_641113, EPI_ISL_641114, EPI_ISL_641115, EPI_ISL_641117, EPI_ISL_641118, EPI_ISL_641120, EPI_ISL_641122, EPI_ISL_641123, EPI_ISL_641124, EPI_ISL_641126, EPI_ISL_641127, EPI_ISL_641128, EPI_ISL_641129, EPI_ISL_641130, EPI_ISL_641131, EPI_ISL_641132, EPI_ISL_641133, EPI_ISL_641134, EPI_ISL_641135, EPI_ISL_641136, EPI_ISL_641137, EPI_ISL_641138, EPI_ISL_641139, EPI_ISL_641140, EPI_ISL_641141, EPI_ISL_641142, EPI_ISL_641145, EPI_ISL_641146, EPI_ISL_641147, EPI_ISL_641148, EPI_ISL_641149, EPI_ISL_641150, EPI_ISL_641151, EPI_ISL_641152, EPI_ISL_641153, EPI_ISL_641154, EPI_ISL_641155, EPI_ISL_641156, EPI_ISL_641157, EPI_ISL_641158, EPI_ISL_641159, EPI_ISL_641160, EPI_ISL_641161, EPI_ISL_641162, EPI_ISL_641163, EPI_ISL_641164, EPI_ISL_641165, EPI_ISL_641167, EPI_ISL_641168, EPI_ISL_641169, EPI_ISL_641170, EPI_ISL_641171, EPI_ISL_641172, EPI_ISL_641173, EPI_ISL_641174, EPI_ISL_641176, EPI_ISL_641177, EPI_ISL_641178, EPI_ISL_641179, EPI_ISL_641181, EPI_ISL_641183, EPI_ISL_641185, EPI_ISL_641186, EPI_ISL_641187, EPI_ISL_641189, EPI_ISL_641192, EPI_ISL_641193, EPI_ISL_641194, EPI_ISL_641195, EPI_ISL_641196, EPI_ISL_641197, EPI_ISL_641199, EPI_ISL_641200, EPI_ISL_641201, EPI_ISL_641203, EPI_ISL_641204, EPI_ISL_641205, EPI_ISL_641206, EPI_ISL_641207, EPI_ISL_641208, EPI_ISL_641209, EPI_ISL_641211, EPI_ISL_641212, EPI_ISL_641213, EPI_ISL_641214, EPI_ISL_641216, EPI_ISL_641217, EPI_ISL_641219, EPI_ISL_641221, EPI_ISL_641222, EPI_ISL_641223, EPI_ISL_641224, EPI_ISL_641225, EPI_ISL_641226, EPI_ISL_641227, EPI_ISL_641228, EPI_ISL_641229, EPI_ISL_641230, EPI_ISL_641233, EPI_ISL_641234, EPI_ISL_641235, EPI_ISL_641238, EPI_ISL_641239, EPI_ISL_641240, EPI_ISL_641241, EPI_ISL_641242, EPI_ISL_641243, EPI_ISL_641244, EPI_ISL_641245, EPI_ISL_641246, EPI_ISL_641247, EPI_ISL_641248, EPI_ISL_641249, EPI_ISL_641250, EPI_ISL_641251, EPI_ISL_641252, EPI_ISL_641253, EPI_ISL_641254, EPI_ISL_641256, EPI_ISL_641257, EPI_ISL_641258, EPI_ISL_641259, EPI_ISL_641260, EPI_ISL_641261, EPI_ISL_641262, EPI_ISL_641263, EPI_ISL_641264, EPI_ISL_641266, EPI_ISL_641267, EPI_ISL_641268, EPI_ISL_641270, EPI_ISL_641271, EPI_ISL_641272, EPI_ISL_641274, EPI_ISL_641275, EPI_ISL_641278, EPI_ISL_641281, EPI_ISL_641282, EPI_ISL_641283, EPI_ISL_641284, EPI_ISL_641285, EPI_ISL_641286, EPI_ISL_641287, EPI_ISL_641288, EPI_ISL_641289, EPI_ISL_641290, EPI_ISL_641291, EPI_ISL_641293, EPI_ISL_641294, EPI_ISL_641295, EPI_ISL_641296, EPI_ISL_641297, EPI_ISL_641298, EPI_ISL_641299, EPI_ISL_641300, EPI_ISL_641301, EPI_ISL_641302, EPI_ISL_641303, EPI_ISL_641304, EPI_ISL_641305, EPI_ISL_641307, EPI_ISL_641308, EPI_ISL_663298, EPI_ISL_663299, EPI_ISL_663300, EPI_ISL_663301, EPI_ISL_663302, EPI_ISL_663303, EPI_ISL_663304, EPI_ISL_663305, EPI_ISL_663306, EPI_ISL_663307, EPI_ISL_663308, EPI_ISL_663309, EPI_ISL_663310, EPI_ISL_663311, EPI_ISL_663312, EPI_ISL_663313, EPI_ISL_663314, EPI_ISL_663315, EPI_ISL_663316, EPI_ISL_663318, EPI_ISL_663319, EPI_ISL_663320, EPI_ISL_663321, EPI_ISL_663322, EPI_ISL_663323, EPI_ISL_663324, EPI_ISL_663325, EPI_ISL_663327, EPI_ISL_663328, EPI_ISL_663329, EPI_ISL_663330, EPI_ISL_663331, EPI_ISL_663332, EPI_ISL_663334, EPI_ISL_663335, EPI_ISL_663337, EPI_ISL_663338, EPI_ISL_663339, EPI_ISL_663340, EPI_ISL_663341, EPI_ISL_663342, EPI_ISL_663343, EPI_ISL_663344, EPI_ISL_663345, EPI_ISL_663346, EPI_ISL_663347, EPI_ISL_663348, EPI_ISL_663349, EPI_ISL_663350, EPI_ISL_663351, EPI_ISL_663352, EPI_ISL_663353, EPI_ISL_663354, EPI_ISL_663355, EPI_ISL_663356, EPI_ISL_663357, EPI_ISL_663358, EPI_ISL_663359, EPI_ISL_663360, EPI_ISL_663361, EPI_ISL_663362, EPI_ISL_663363, EPI_ISL_663364, EPI_ISL_663365, EPI_ISL_663366, EPI_ISL_663367, EPI_ISL_663368, EPI_ISL_663369, EPI_ISL_663370, EPI_ISL_663371, EPI_ISL_663372, EPI_ISL_663373, EPI_ISL_663374, EPI_ISL_663375, EPI_ISL_663377, EPI_ISL_663378, EPI_ISL_663379, EPI_ISL_663380, EPI_ISL_663381, EPI_ISL_663382, EPI_ISL_663383, EPI_ISL_663384, EPI_ISL_663385, EPI_ISL_663386, EPI_ISL_663388, EPI_ISL_663389, EPI_ISL_663390, EPI_ISL_663391, EPI_ISL_663392, EPI_ISL_663393, EPI_ISL_663394, EPI_ISL_663395, EPI_ISL_663396, EPI_ISL_663398, EPI_ISL_663399, EPI_ISL_663400, EPI_ISL_663401, EPI_ISL_663402, EPI_ISL_663403, EPI_ISL_663404, EPI_ISL_663405, EPI_ISL_663406, EPI_ISL_663407, EPI_ISL_663408, EPI_ISL_663409, EPI_ISL_663410, EPI_ISL_663411, EPI_ISL_663412, EPI_ISL_663413, EPI_ISL_663415, EPI_ISL_663416, EPI_ISL_663417, EPI_ISL_663418, EPI_ISL_663419, EPI_ISL_663420, EPI_ISL_663421, EPI_ISL_663422, EPI_ISL_663423, EPI_ISL_663424, EPI_ISL_663425, EPI_ISL_663426, EPI_ISL_663427, EPI_ISL_663428, EPI_ISL_663429, EPI_ISL_663430, EPI_ISL_663431, EPI_ISL_663432, EPI_ISL_663433, EPI_ISL_663434, EPI_ISL_663435, EPI_ISL_663436, EPI_ISL_663438, EPI_ISL_663439, EPI_ISL_663440, EPI_ISL_663529, EPI_ISL_663530, EPI_ISL_663531, EPI_ISL_663532, EPI_ISL_663533, EPI_ISL_663534, EPI_ISL_663535, EPI_ISL_663536, EPI_ISL_663537, EPI_ISL_663538, EPI_ISL_663539, EPI_ISL_663540, EPI_ISL_663541, EPI_ISL_663542, EPI_ISL_663543, EPI_ISL_663544, EPI_ISL_663545, EPI_ISL_663546, EPI_ISL_663547, EPI_ISL_663548, EPI_ISL_663549, EPI_ISL_663550, EPI_ISL_663551, EPI_ISL_663552, EPI_ISL_663553, EPI_ISL_663554, EPI_ISL_663555, EPI_ISL_663556, EPI_ISL_663557, EPI_ISL_663558, EPI_ISL_663561, EPI_ISL_663562, EPI_ISL_663563, EPI_ISL_663564, EPI_ISL_663565, EPI_ISL_663566, EPI_ISL_663567, EPI_ISL_663568, EPI_ISL_663569, EPI_ISL_663570, EPI_ISL_663571, EPI_ISL_663572, EPI_ISL_663573, EPI_ISL_663574, EPI_ISL_663575, EPI_ISL_663576, EPI_ISL_663577, EPI_ISL_663578, EPI_ISL_663579, EPI_ISL_663580, EPI_ISL_663581, EPI_ISL_663582, EPI_ISL_663583, EPI_ISL_663584, EPI_ISL_663585, EPI_ISL_663586, EPI_ISL_663587, EPI_ISL_663588, EPI_ISL_663589, EPI_ISL_663590, EPI_ISL_663591, EPI_ISL_663592, EPI_ISL_663593, EPI_ISL_663594, EPI_ISL_663595, EPI_ISL_663596, EPI_ISL_663597, EPI_ISL_663598, EPI_ISL_663599, EPI_ISL_663600, EPI_ISL_663601, EPI_ISL_663602, EPI_ISL_663603, EPI_ISL_663604, EPI_ISL_663605, EPI_ISL_663606, EPI_ISL_663607, EPI_ISL_663608, EPI_ISL_663609, EPI_ISL_663610, EPI_ISL_663611, EPI_ISL_663612, EPI_ISL_663613, EPI_ISL_663614, EPI_ISL_663615, EPI_ISL_663616, EPI_ISL_663617, EPI_ISL_663618, EPI_ISL_663619, EPI_ISL_663620, EPI_ISL_663621, EPI_ISL_663622, EPI_ISL_663623, EPI_ISL_663624, EPI_ISL_663625, EPI_ISL_663626, EPI_ISL_663627, EPI_ISL_663628, EPI_ISL_663629, EPI_ISL_663630, EPI_ISL_663631, EPI_ISL_663632, EPI_ISL_663633, EPI_ISL_663634, EPI_ISL_663635, EPI_ISL_663636, EPI_ISL_663637, EPI_ISL_663638, EPI_ISL_663639, EPI_ISL_663640, EPI_ISL_663641, EPI_ISL_663642, EPI_ISL_663643, EPI_ISL_663644, EPI_ISL_663645, EPI_ISL_663646, EPI_ISL_663647, EPI_ISL_663648, EPI_ISL_663649, EPI_ISL_663650, EPI_ISL_663651, EPI_ISL_663652, EPI_ISL_663654, EPI_ISL_663655, EPI_ISL_663656, EPI_ISL_663657, EPI_ISL_663658, EPI_ISL_663659, EPI_ISL_663660, EPI_ISL_663661, EPI_ISL_663662, EPI_ISL_663663, EPI_ISL_663664, EPI_ISL_663665, EPI_ISL_663666, EPI_ISL_663667, EPI_ISL_663668, EPI_ISL_663669, EPI_ISL_663670, EPI_ISL_663671, EPI_ISL_663672, EPI_ISL_663673, EPI_ISL_663674, EPI_ISL_663675, EPI_ISL_663676, EPI_ISL_663677, EPI_ISL_663678, EPI_ISL_663679, EPI_ISL_663680, EPI_ISL_663681, EPI_ISL_663682, EPI_ISL_663684, EPI_ISL_663685, EPI_ISL_663686, EPI_ISL_663687, EPI_ISL_663688, EPI_ISL_663689, EPI_ISL_663690, EPI_ISL_663691, EPI_ISL_663692, EPI_ISL_663693, EPI_ISL_663694, EPI_ISL_663695, EPI_ISL_663696, EPI_ISL_663697, EPI_ISL_663698, EPI_ISL_663699, EPI_ISL_663700, EPI_ISL_663701, EPI_ISL_663702, EPI_ISL_663703, EPI_ISL_663704, EPI_ISL_663705, EPI_ISL_663707, EPI_ISL_663708, EPI_ISL_663709, EPI_ISL_663710, EPI_ISL_663711, EPI_ISL_663712, EPI_ISL_663713, EPI_ISL_663714, EPI_ISL_663715, EPI_ISL_663716, EPI_ISL_663717, EPI_ISL_663718, EPI_ISL_663719, EPI_ISL_663720, EPI_ISL_663721, EPI_ISL_663722, EPI_ISL_663723, EPI_ISL_663724, EPI_ISL_663725, EPI_ISL_663726, EPI_ISL_663727, EPI_ISL_663728, EPI_ISL_663729, EPI_ISL_663730, EPI_ISL_663731, EPI_ISL_663732, EPI_ISL_663733, EPI_ISL_663734, EPI_ISL_663735, EPI_ISL_663736, EPI_ISL_663737, EPI_ISL_663738, EPI_ISL_663739, EPI_ISL_663740, EPI_ISL_663741, EPI_ISL_663742, EPI_ISL_663743, EPI_ISL_663744, EPI_ISL_663745, EPI_ISL_663746, EPI_ISL_663747, EPI_ISL_663748, EPI_ISL_663749, EPI_ISL_663750, EPI_ISL_663751, EPI_ISL_663752, EPI_ISL_663753, EPI_ISL_663754, EPI_ISL_663755, EPI_ISL_663756, EPI_ISL_663757, EPI_ISL_663758, EPI_ISL_663759, EPI_ISL_663760, EPI_ISL_663761, EPI_ISL_663762, EPI_ISL_663763, EPI_ISL_663764, EPI_ISL_663765, EPI_ISL_663766, EPI_ISL_663767, EPI_ISL_663768, EPI_ISL_663769, EPI_ISL_663770, EPI_ISL_663771, EPI_ISL_663772, EPI_ISL_663773, EPI_ISL_663774, EPI_ISL_663775, EPI_ISL_663776, EPI_ISL_663777, EPI_ISL_663778, EPI_ISL_663779, EPI_ISL_663780, EPI_ISL_663781, EPI_ISL_663782, EPI_ISL_663784, EPI_ISL_663785, EPI_ISL_663786, EPI_ISL_663787, EPI_ISL_663788, EPI_ISL_663789, EPI_ISL_663790, EPI_ISL_663791, EPI_ISL_663792, EPI_ISL_663793, EPI_ISL_663794, EPI_ISL_663795, EPI_ISL_663796, EPI_ISL_663797, EPI_ISL_663798, EPI_ISL_663799, EPI_ISL_663800, EPI_ISL_663801, EPI_ISL_663802, EPI_ISL_663803, EPI_ISL_663804, EPI_ISL_663805, EPI_ISL_663807, EPI_ISL_663808, EPI_ISL_663809, EPI_ISL_663810, EPI_ISL_663811, EPI_ISL_663812, EPI_ISL_663813, EPI_ISL_663814, EPI_ISL_663815, EPI_ISL_663816, EPI_ISL_663817, EPI_ISL_663818, EPI_ISL_663819, EPI_ISL_663820, EPI_ISL_663821, EPI_ISL_663822, EPI_ISL_663823, EPI_ISL_663824, EPI_ISL_663825, EPI_ISL_663826, EPI_ISL_663827, EPI_ISL_663828, EPI_ISL_663829, EPI_ISL_663830, EPI_ISL_663831, EPI_ISL_663832, EPI_ISL_663834, EPI_ISL_663835, EPI_ISL_663836, EPI_ISL_663837, EPI_ISL_663838, EPI_ISL_663839, EPI_ISL_663840, EPI_ISL_663841, EPI_ISL_663842, EPI_ISL_663843, EPI_ISL_663844, EPI_ISL_663845, EPI_ISL_663846, EPI_ISL_663847, EPI_ISL_663848, EPI_ISL_663849, EPI_ISL_663850, EPI_ISL_663851, EPI_ISL_663852, EPI_ISL_663853, EPI_ISL_663854, EPI_ISL_663855, EPI_ISL_663856, EPI_ISL_663857, EPI_ISL_663858, EPI_ISL_663859, EPI_ISL_663860, EPI_ISL_663861, EPI_ISL_663862, EPI_ISL_663863, EPI_ISL_663864, EPI_ISL_663865, EPI_ISL_663866, EPI_ISL_663867, EPI_ISL_663868, EPI_ISL_663869, EPI_ISL_663870, EPI_ISL_663871, EPI_ISL_663872, EPI_ISL_663873, EPI_ISL_663874, EPI_ISL_663875, EPI_ISL_663876, EPI_ISL_663877, EPI_ISL_663878, EPI_ISL_663879, EPI_ISL_663880, EPI_ISL_663881, EPI_ISL_663882, EPI_ISL_663883, EPI_ISL_663884, EPI_ISL_663885, EPI_ISL_663886, EPI_ISL_663887, EPI_ISL_663888, EPI_ISL_663889, EPI_ISL_663890, EPI_ISL_663891, EPI_ISL_663892, EPI_ISL_663893, EPI_ISL_663894, EPI_ISL_663895, EPI_ISL_663896, EPI_ISL_663897, EPI_ISL_663898, EPI_ISL_663899, EPI_ISL_663900, EPI_ISL_663901, EPI_ISL_663902, EPI_ISL_663903, EPI_ISL_663904, EPI_ISL_663905, EPI_ISL_663906, EPI_ISL_663907, EPI_ISL_663908, EPI_ISL_663909, EPI_ISL_663910, EPI_ISL_663911, EPI_ISL_663912, EPI_ISL_663913, EPI_ISL_663914, EPI_ISL_663915, EPI_ISL_663916, EPI_ISL_663917, EPI_ISL_663918, EPI_ISL_663919, EPI_ISL_663920, EPI_ISL_663921, EPI_ISL_663922, EPI_ISL_663923, EPI_ISL_663924, EPI_ISL_663925, EPI_ISL_663926, EPI_ISL_663927, EPI_ISL_663928, EPI_ISL_663929, EPI_ISL_663930, EPI_ISL_663931, EPI_ISL_663932, EPI_ISL_663933, EPI_ISL_663934, EPI_ISL_663935, EPI_ISL_663936, EPI_ISL_663937, EPI_ISL_663938, EPI_ISL_663939, EPI_ISL_663941, EPI_ISL_663942, EPI_ISL_663944, EPI_ISL_663945, EPI_ISL_663946, EPI_ISL_663948, EPI_ISL_663949, EPI_ISL_663951, EPI_ISL_663952, EPI_ISL_663953, EPI_ISL_663954, EPI_ISL_663955, EPI_ISL_663956, EPI_ISL_663957, EPI_ISL_663958, EPI_ISL_663959, EPI_ISL_663960, EPI_ISL_663961, EPI_ISL_663962, EPI_ISL_663963, EPI_ISL_663964, EPI_ISL_663965, EPI_ISL_663966, EPI_ISL_663967, EPI_ISL_663968, EPI_ISL_663969, EPI_ISL_663970, EPI_ISL_663971, EPI_ISL_663972, EPI_ISL_663973, EPI_ISL_663974, EPI_ISL_663975, EPI_ISL_663976, EPI_ISL_663977, EPI_ISL_663978, EPI_ISL_663979, EPI_ISL_663980, EPI_ISL_663981, EPI_ISL_663982, EPI_ISL_663983, EPI_ISL_663984, EPI_ISL_663985, EPI_ISL_663986, EPI_ISL_663987, EPI_ISL_663988, EPI_ISL_663989, EPI_ISL_663990, EPI_ISL_663991, EPI_ISL_663992, EPI_ISL_663993, EPI_ISL_663994, EPI_ISL_663995, EPI_ISL_663996, EPI_ISL_663997, EPI_ISL_663998, EPI_ISL_663999, EPI_ISL_664000, EPI_ISL_664001, EPI_ISL_664002, EPI_ISL_664003, EPI_ISL_664004, EPI_ISL_664005, EPI_ISL_664007, EPI_ISL_664008, EPI_ISL_664009, EPI_ISL_664010, EPI_ISL_664011, EPI_ISL_664012, EPI_ISL_664013, EPI_ISL_664014, EPI_ISL_664015, EPI_ISL_664016, EPI_ISL_664017, EPI_ISL_664018, EPI_ISL_664019, EPI_ISL_779414, EPI_ISL_779415, EPI_ISL_779416, EPI_ISL_779417, EPI_ISL_779418, EPI_ISL_779419, EPI_ISL_779420, EPI_ISL_779421, EPI_ISL_779422, EPI_ISL_779423, EPI_ISL_779425, EPI_ISL_779426, EPI_ISL_779427, EPI_ISL_779428, EPI_ISL_779429, EPI_ISL_779430, EPI_ISL_779431, EPI_ISL_779432, EPI_ISL_779433, EPI_ISL_779434, EPI_ISL_779437, EPI_ISL_779438, EPI_ISL_779439, EPI_ISL_779440, EPI_ISL_779441, EPI_ISL_779444, EPI_ISL_779445, EPI_ISL_779448, EPI_ISL_779449, EPI_ISL_779450, EPI_ISL_779451, EPI_ISL_779452, EPI_ISL_779453, EPI_ISL_779455, EPI_ISL_779456, EPI_ISL_779457, EPI_ISL_779458, EPI_ISL_779460, EPI_ISL_779461, EPI_ISL_779463, EPI_ISL_779467, EPI_ISL_779468, EPI_ISL_779469, EPI_ISL_779470, EPI_ISL_779471, EPI_ISL_779472, EPI_ISL_779473, EPI_ISL_779474, EPI_ISL_779475, EPI_ISL_779476, EPI_ISL_779477, EPI_ISL_779478, EPI_ISL_779480, EPI_ISL_779481, EPI_ISL_779482, EPI_ISL_779483, EPI_ISL_779484, EPI_ISL_779486, EPI_ISL_779487, EPI_ISL_779488, EPI_ISL_779489, EPI_ISL_779490, EPI_ISL_779491, EPI_ISL_779492, EPI_ISL_779493, EPI_ISL_779494, EPI_ISL_779495, EPI_ISL_779496, EPI_ISL_779497, EPI_ISL_779498, EPI_ISL_779499, EPI_ISL_779500, EPI_ISL_779501, EPI_ISL_779502, EPI_ISL_779503, EPI_ISL_779504, EPI_ISL_779505, EPI_ISL_779506, EPI_ISL_779509, EPI_ISL_779510, EPI_ISL_779511, EPI_ISL_779512, EPI_ISL_779513, EPI_ISL_779514, EPI_ISL_779515, EPI_ISL_779517, EPI_ISL_779518, EPI_ISL_779519, EPI_ISL_779520, EPI_ISL_779521, EPI_ISL_779522, EPI_ISL_779523, EPI_ISL_779524, EPI_ISL_779525, EPI_ISL_779526, EPI_ISL_779527, EPI_ISL_779528, EPI_ISL_779529, EPI_ISL_779530, EPI_ISL_779531, EPI_ISL_779532, EPI_ISL_779534, EPI_ISL_779536, EPI_ISL_779538, EPI_ISL_779539, EPI_ISL_779540, EPI_ISL_779541, EPI_ISL_779542, EPI_ISL_779543, EPI_ISL_779544, EPI_ISL_779546, EPI_ISL_779547, EPI_ISL_779548, EPI_ISL_779549, EPI_ISL_779550, EPI_ISL_779551, EPI_ISL_779552, EPI_ISL_779553, EPI_ISL_779555, EPI_ISL_779556, EPI_ISL_779557, EPI_ISL_779558, EPI_ISL_779559, EPI_ISL_779561, EPI_ISL_779562, EPI_ISL_779563, EPI_ISL_779564, EPI_ISL_779565, EPI_ISL_779566, EPI_ISL_779567, EPI_ISL_779568, EPI_ISL_779571, EPI_ISL_779573, EPI_ISL_779574, EPI_ISL_779575, EPI_ISL_779576, EPI_ISL_779577, EPI_ISL_779578, EPI_ISL_779579, EPI_ISL_779581, EPI_ISL_779589, EPI_ISL_877579, EPI_ISL_962816, EPI_ISL_962817, EPI_ISL_1033125, EPI_ISL_1033126, EPI_ISL_1033127, EPI_ISL_1033128, EPI_ISL_1033129, EPI_ISL_1033132, EPI_ISL_1033135, EPI_ISL_1033136, EPI_ISL_1033137, EPI_ISL_1033139, EPI_ISL_1055265, EPI_ISL_1055266, EPI_ISL_1055267, EPI_ISL_1055268, EPI_ISL_1055269, EPI_ISL_1055270, EPI_ISL_1055271, EPI_ISL_1055272, EPI_ISL_1055273, EPI_ISL_1055274, EPI_ISL_1055275, EPI_ISL_1055276, EPI_ISL_1055277, EPI_ISL_1055279, EPI_ISL_1055280, EPI_ISL_1055281, EPI_ISL_1055282, EPI_ISL_1055283, EPI_ISL_1055284, EPI_ISL_1055285, EPI_ISL_1055286, EPI_ISL_1055287, EPI_ISL_1055288, EPI_ISL_1055289, EPI_ISL_1055290, EPI_ISL_1055291, EPI_ISL_1055292, EPI_ISL_1055293, EPI_ISL_1055294, EPI_ISL_1055295, EPI_ISL_1055296, EPI_ISL_1055297, EPI_ISL_1055298, EPI_ISL_1055299, EPI_ISL_1055300, EPI_ISL_1055303, EPI_ISL_1055304, EPI_ISL_1055305, EPI_ISL_1055306, EPI_ISL_1055307, EPI_ISL_1055308, EPI_ISL_1055309, EPI_ISL_1055310, EPI_ISL_1055311, EPI_ISL_1055312, EPI_ISL_1055313, EPI_ISL_1055314, EPI_ISL_1055315, EPI_ISL_1055316, EPI_ISL_1055317, EPI_ISL_1055318, EPI_ISL_1055319, EPI_ISL_1055320, EPI_ISL_1055321, EPI_ISL_1055322, EPI_ISL_1055323, EPI_ISL_1055324, EPI_ISL_1055325, EPI_ISL_1055326, EPI_ISL_1055327, EPI_ISL_1055328, EPI_ISL_1055329, EPI_ISL_1055330, EPI_ISL_1055331, EPI_ISL_1055332, EPI_ISL_1055333, EPI_ISL_1055334, EPI_ISL_1055335, EPI_ISL_1055336, EPI_ISL_1055337, EPI_ISL_1055338, EPI_ISL_1055339, EPI_ISL_1055342, EPI_ISL_1055343, EPI_ISL_1055344, EPI_ISL_1055345, EPI_ISL_1055346, EPI_ISL_1055347, EPI_ISL_1055349, EPI_ISL_1055350, EPI_ISL_1055351, EPI_ISL_1055353, EPI_ISL_1055354, EPI_ISL_1055355, EPI_ISL_1055356, EPI_ISL_1055357, EPI_ISL_1055359, EPI_ISL_1055360, EPI_ISL_1061399, EPI_ISL_1061400, EPI_ISL_1061401 | | | |
| see above | Microbiological Diagnostic Unit - Public Health Laboratory (MDU-PHL) | MDU-PHL | M. B.; M.L.; N.L.; Sait; Sait, M.; Schultz; Schultz M.; Schultz M. B.; Schultz M.B.; Seemann T.; Seemann, T.; Sherry; Sherry, N. |
| EPI_ISL_518651, EPI_ISL_518935, EPI_ISL_519895, EPI_ISL_520604, EPI_ISL_520605, EPI_ISL_520644 | Microbiological Diagnostic Unit - Public Health Laboratory (MDU-PHL) | Microbiological Diagnostic Unit - Public Health Laboratory (MDU-PHL) | Sait, M.; Schultz M.; Seemann T.; Sherry, N. |
| EPI_ISL_649149, EPI_ISL_649150, EPI_ISL_649151 | Microbiological Diagnostic Unit - Public Health Laboratory (MDU-PHL), The Peter Doherty Institute for Infection and Immunity | Microbiological Diagnostic Unit - Public Health Laboratory (MDU-PHL), The Peter Doherty Institute for Infection and Immunity | Caly, L.; Druce, J.; M.B.; M.L.; N.L.; Sait; Schultz; Seemann, T.; Sherry |
| EPI_ISL_419714, EPI_ISL_419715, EPI_ISL_419716, EPI_ISL_419717, EPI_ISL_419718, EPI_ISL_419719, EPI_ISL_419720, EPI_ISL_419721, EPI_ISL_419722, EPI_ISL_419723, EPI_ISL_419724, EPI_ISL_419725, EPI_ISL_419726, EPI_ISL_419727, EPI_ISL_419728, EPI_ISL_419729, EPI_ISL_419730, EPI_ISL_419731, EPI_ISL_419823, EPI_ISL_419824, EPI_ISL_419827, EPI_ISL_419828, EPI_ISL_419829, EPI_ISL_419830, EPI_ISL_419980, EPI_ISL_419999, EPI_ISL_420000, EPI_ISL_420001, EPI_ISL_420002, EPI_ISL_420003, EPI_ISL_420006, EPI_ISL_420007, EPI_ISL_420008, EPI_ISL_420009, EPI_ISL_420010, EPI_ISL_420011, EPI_ISL_420012, EPI_ISL_420013, EPI_ISL_420014, EPI_ISL_420015, EPI_ISL_420016, EPI_ISL_420017, EPI_ISL_426905, EPI_ISL_426906, EPI_ISL_426907, EPI_ISL_426908, EPI_ISL_426909, EPI_ISL_426910, EPI_ISL_426911, EPI_ISL_426912, EPI_ISL_426914, EPI_ISL_426915, EPI_ISL_426916, EPI_ISL_426917, EPI_ISL_426918, EPI_ISL_426919, EPI_ISL_426920, EPI_ISL_426921, EPI_ISL_426922, EPI_ISL_427055, EPI_ISL_427056, EPI_ISL_427057, EPI_ISL_427058, EPI_ISL_427059, EPI_ISL_427060, EPI_ISL_427061, EPI_ISL_427062, EPI_ISL_427063, EPI_ISL_427064, EPI_ISL_427065, EPI_ISL_427066, EPI_ISL_427067, EPI_ISL_427068, EPI_ISL_427069, EPI_ISL_427070, EPI_ISL_427071, EPI_ISL_427072, EPI_ISL_427073, EPI_ISL_427074, EPI_ISL_427076, EPI_ISL_427077, EPI_ISL_427078, EPI_ISL_427133, EPI_ISL_427148, EPI_ISL_430470, EPI_ISL_430471, EPI_ISL_430472, EPI_ISL_430639, EPI_ISL_430640, EPI_ISL_430641, EPI_ISL_430643, EPI_ISL_430644, EPI_ISL_430645, EPI_ISL_430646, EPI_ISL_430647, EPI_ISL_430648, EPI_ISL_430649, EPI_ISL_430650, EPI_ISL_430651, EPI_ISL_430653, EPI_ISL_430654, EPI_ISL_430655, EPI_ISL_430656, EPI_ISL_430657, EPI_ISL_430658, EPI_ISL_430659, EPI_ISL_430660, EPI_ISL_430661, EPI_ISL_430662, EPI_ISL_430663, EPI_ISL_430664, EPI_ISL_430665, EPI_ISL_430666, EPI_ISL_430667, EPI_ISL_430668, EPI_ISL_430669, EPI_ISL_430670, EPI_ISL_430671, EPI_ISL_430672, EPI_ISL_430673, EPI_ISL_430674, EPI_ISL_430675, EPI_ISL_430676, EPI_ISL_430677, EPI_ISL_430678, EPI_ISL_430680, EPI_ISL_430681, EPI_ISL_430682, EPI_ISL_430683, EPI_ISL_430684, EPI_ISL_430685, EPI_ISL_430716, EPI_ISL_430717 | | | |
| see above | Microbiological Diagnostic Unit Public Health Laboratory | Microbiological Diagnostic Unit Public Health Laboratory | Sait, M.; Schultz M.; Seemann T.; Sherry, N. |
| EPI_ISL_426637, EPI_ISL_426638, EPI_ISL_426639, EPI_ISL_426640, EPI_ISL_426641, EPI_ISL_426642, EPI_ISL_426643, EPI_ISL_426644, EPI_ISL_426645, EPI_ISL_426646, EPI_ISL_426647, EPI_ISL_426648, EPI_ISL_426649, EPI_ISL_426650, EPI_ISL_426651, EPI_ISL_426652, EPI_ISL_426653, EPI_ISL_426654, EPI_ISL_426655, EPI_ISL_426656, EPI_ISL_426657, EPI_ISL_426658, EPI_ISL_426659, EPI_ISL_426660, EPI_ISL_426661, EPI_ISL_426662, EPI_ISL_426663, EPI_ISL_426664, EPI_ISL_426665, EPI_ISL_426666, EPI_ISL_426667, EPI_ISL_426668, EPI_ISL_426669, EPI_ISL_426670, EPI_ISL_426671, EPI_ISL_426672, EPI_ISL_426673, EPI_ISL_426674, EPI_ISL_426675, EPI_ISL_426676, EPI_ISL_426679, EPI_ISL_426680, EPI_ISL_426681, EPI_ISL_426682, EPI_ISL_426683, EPI_ISL_426684, EPI_ISL_426686, EPI_ISL_426687, EPI_ISL_426688, EPI_ISL_426689, EPI_ISL_426690, EPI_ISL_426691, EPI_ISL_426692, EPI_ISL_426693, EPI_ISL_426694, EPI_ISL_426695, EPI_ISL_426696, EPI_ISL_426697, EPI_ISL_426698, EPI_ISL_426700, EPI_ISL_426701, EPI_ISL_426702, EPI_ISL_426705, EPI_ISL_426707, EPI_ISL_426708, EPI_ISL_426709, EPI_ISL_426710, EPI_ISL_426711, EPI_ISL_426713, EPI_ISL_426714, EPI_ISL_426715, EPI_ISL_426716, EPI_ISL_426717, EPI_ISL_426719, EPI_ISL_426721, EPI_ISL_426722, EPI_ISL_426723, EPI_ISL_426724, EPI_ISL_426725, EPI_ISL_426726, EPI_ISL_426727, EPI_ISL_426728, EPI_ISL_426729, EPI_ISL_426730, EPI_ISL_426731, EPI_ISL_426732, EPI_ISL_426733, EPI_ISL_426734, EPI_ISL_426735, EPI_ISL_426736, EPI_ISL_426737, EPI_ISL_426738, EPI_ISL_426739, EPI_ISL_426740, EPI_ISL_426743, EPI_ISL_426744, EPI_ISL_426745, EPI_ISL_426746, EPI_ISL_426747, EPI_ISL_426749, EPI_ISL_426750, EPI_ISL_426751, EPI_ISL_426753, EPI_ISL_426754, EPI_ISL_426755, EPI_ISL_426756, EPI_ISL_426757, EPI_ISL_426758, EPI_ISL_426759, EPI_ISL_426760, EPI_ISL_426762, EPI_ISL_426765, EPI_ISL_426766, EPI_ISL_426767, EPI_ISL_426768, EPI_ISL_426769, EPI_ISL_426771, EPI_ISL_426772, EPI_ISL_426775, EPI_ISL_426778, EPI_ISL_426789, EPI_ISL_426791, EPI_ISL_426792, EPI_ISL_426793, EPI_ISL_426794, EPI_ISL_426795, EPI_ISL_426796, EPI_ISL_426797, EPI_ISL_426798, EPI_ISL_426799, EPI_ISL_426800, EPI_ISL_426802, EPI_ISL_426804, EPI_ISL_426806, EPI_ISL_426807, EPI_ISL_426808, EPI_ISL_426809, EPI_ISL_426810, EPI_ISL_426811, EPI_ISL_426812, EPI_ISL_426813, EPI_ISL_426814, EPI_ISL_426816, EPI_ISL_426817, EPI_ISL_426819, EPI_ISL_426820, EPI_ISL_426821, EPI_ISL_426822, EPI_ISL_426823, EPI_ISL_426824, EPI_ISL_426825, EPI_ISL_426826, EPI_ISL_426831, EPI_ISL_426832, EPI_ISL_426833, EPI_ISL_426835, EPI_ISL_426836, EPI_ISL_426837, EPI_ISL_426838, EPI_ISL_426839, EPI_ISL_426840, EPI_ISL_426841, EPI_ISL_426842, EPI_ISL_426843, EPI_ISL_426844, EPI_ISL_426846, EPI_ISL_426847, EPI_ISL_426848, EPI_ISL_426849, EPI_ISL_426850, EPI_ISL_426851, EPI_ISL_426854, EPI_ISL_426855, EPI_ISL_426856, EPI_ISL_426859, EPI_ISL_426860, EPI_ISL_426861, EPI_ISL_426862, EPI_ISL_426863, EPI_ISL_426864, EPI_ISL_426865, EPI_ISL_426866, EPI_ISL_426867, EPI_ISL_426869, EPI_ISL_426870, EPI_ISL_426871, EPI_ISL_426872, EPI_ISL_426873, EPI_ISL_426874, EPI_ISL_426875, EPI_ISL_426876, EPI_ISL_426877, EPI_ISL_426878, EPI_ISL_426881, EPI_ISL_426923, EPI_ISL_426924, EPI_ISL_426925, EPI_ISL_426926, EPI_ISL_426927, EPI_ISL_426928, EPI_ISL_426929, EPI_ISL_426931, EPI_ISL_426932, EPI_ISL_426933, EPI_ISL_426934, EPI_ISL_426935, EPI_ISL_426936, EPI_ISL_426937, EPI_ISL_426938, EPI_ISL_426939, EPI_ISL_426940, EPI_ISL_426941, EPI_ISL_426942, EPI_ISL_426943, EPI_ISL_426945, EPI_ISL_426947, EPI_ISL_426948, EPI_ISL_426949, EPI_ISL_426950, EPI_ISL_426951, EPI_ISL_426952, EPI_ISL_426953, EPI_ISL_426954, EPI_ISL_426955, EPI_ISL_426956, EPI_ISL_426957, EPI_ISL_426958, EPI_ISL_426960, EPI_ISL_426961, EPI_ISL_426962, EPI_ISL_426963, EPI_ISL_426964, EPI_ISL_426965, EPI_ISL_426966, EPI_ISL_426968, EPI_ISL_426969, EPI_ISL_426970, EPI_ISL_426971, EPI_ISL_426972, EPI_ISL_426973, EPI_ISL_426975, EPI_ISL_426976, EPI_ISL_426977, EPI_ISL_426978, EPI_ISL_426979, EPI_ISL_426980, EPI_ISL_426981, EPI_ISL_426982, EPI_ISL_426985, EPI_ISL_426986, EPI_ISL_426987, EPI_ISL_426988, EPI_ISL_426989, EPI_ISL_426990, EPI_ISL_426991, EPI_ISL_426992, EPI_ISL_426993, EPI_ISL_426994, EPI_ISL_426995, EPI_ISL_426998, EPI_ISL_426999, EPI_ISL_427000, EPI_ISL_427001, EPI_ISL_427002, EPI_ISL_427003, EPI_ISL_427004, EPI_ISL_427005, EPI_ISL_427007, EPI_ISL_427008, EPI_ISL_427009, EPI_ISL_427010, EPI_ISL_427011, EPI_ISL_427012, EPI_ISL_427013, EPI_ISL_427014, EPI_ISL_427015, EPI_ISL_427016, EPI_ISL_427017, EPI_ISL_427018, EPI_ISL_427019, EPI_ISL_427020, EPI_ISL_427022, EPI_ISL_427023, EPI_ISL_427024, EPI_ISL_427025, EPI_ISL_427026, EPI_ISL_427028, EPI_ISL_427029, EPI_ISL_427030, EPI_ISL_427031, EPI_ISL_427032, EPI_ISL_427034, EPI_ISL_427036, EPI_ISL_427038, EPI_ISL_427039, EPI_ISL_427040, EPI_ISL_427041, EPI_ISL_427042, EPI_ISL_427044, EPI_ISL_427045, EPI_ISL_427046, EPI_ISL_427047, EPI_ISL_427048, EPI_ISL_427049, EPI_ISL_427050, EPI_ISL_427051, EPI_ISL_427053, EPI_ISL_427079, EPI_ISL_427080, EPI_ISL_427081, EPI_ISL_427082, EPI_ISL_427083, EPI_ISL_427084, EPI_ISL_427087, EPI_ISL_427088, EPI_ISL_427089, EPI_ISL_427090, EPI_ISL_427091, EPI_ISL_427092, EPI_ISL_427093, EPI_ISL_427094, EPI_ISL_427095, EPI_ISL_427096, EPI_ISL_427097, EPI_ISL_427100, EPI_ISL_427101, EPI_ISL_427102, EPI_ISL_427103, EPI_ISL_427105, EPI_ISL_427106, EPI_ISL_427107, EPI_ISL_427108, EPI_ISL_427109, EPI_ISL_427110, EPI_ISL_427111, EPI_ISL_427112, EPI_ISL_427113, EPI_ISL_427114, EPI_ISL_427115, EPI_ISL_427116, EPI_ISL_427117, EPI_ISL_427118, EPI_ISL_427119, EPI_ISL_427120, EPI_ISL_427121, EPI_ISL_427122, EPI_ISL_427123, EPI_ISL_427124, EPI_ISL_427125, EPI_ISL_427126, EPI_ISL_427128, EPI_ISL_427129, EPI_ISL_427130, EPI_ISL_427131, EPI_ISL_427134, EPI_ISL_427135, EPI_ISL_427137, EPI_ISL_427138, EPI_ISL_427139, EPI_ISL_427140, EPI_ISL_427141, EPI_ISL_427142, EPI_ISL_427143, EPI_ISL_427144, EPI_ISL_427145, EPI_ISL_427146, EPI_ISL_427147, EPI_ISL_427153, EPI_ISL_427155, EPI_ISL_427156, EPI_ISL_427157, EPI_ISL_427158, EPI_ISL_427159, EPI_ISL_427160, EPI_ISL_436111, EPI_ISL_436112, EPI_ISL_436113, EPI_ISL_436114, EPI_ISL_436115, EPI_ISL_436116, EPI_ISL_436117, EPI_ISL_436118, EPI_ISL_436119, EPI_ISL_436120, EPI_ISL_436121, EPI_ISL_436122, EPI_ISL_436123, EPI_ISL_436124, EPI_ISL_436125, EPI_ISL_436126, EPI_ISL_436128, EPI_ISL_436129, EPI_ISL_436130, EPI_ISL_436131, EPI_ISL_436132, EPI_ISL_456432, EPI_ISL_456433, EPI_ISL_456444, EPI_ISL_456445, EPI_ISL_456449, EPI_ISL_456451, EPI_ISL_456465, EPI_ISL_456474, EPI_ISL_456491, EPI_ISL_456516, EPI_ISL_456532, EPI_ISL_456629, EPI_ISL_456630 | | | |
| see above | Victorian Infectious Diseases Reference Laboratory (VIDRL) | Microbiological Diagnostic Unit Public Health Laboratory and Victorian Infectious Diseases Reference Laboratory, Doherty Institute | Caly L.; Druce J.; Sait, M.; Schultz M.; Seemann T.; Sherry, N. |
| EPI_ISL_436688, EPI_ISL_436689 | Victorian Infectious Diseases Reference Laboratory (VIDRL) | Microbiological Diagnostic Unit Public Health Laboratory and Victorian Infectious Diseases Reference Laboratory, The Peter Doherty Institute for Infection & Immunity | Caly L.; Druce J.; Sait, M.; Schultz M.; Seemann T.; Sherry, N. |
| EPI_ISL_430473, EPI_ISL_430474, EPI_ISL_430475, EPI_ISL_430476, EPI_ISL_430477, EPI_ISL_430478, EPI_ISL_430479, EPI_ISL_430480, EPI_ISL_430481, EPI_ISL_430482, EPI_ISL_430484, EPI_ISL_430485, EPI_ISL_430486, EPI_ISL_430487, EPI_ISL_430488, EPI_ISL_430489, EPI_ISL_430490, EPI_ISL_430491, EPI_ISL_430492, EPI_ISL_430493, EPI_ISL_430498, EPI_ISL_430499, EPI_ISL_430500, EPI_ISL_430501, EPI_ISL_430502, EPI_ISL_430503, EPI_ISL_430505, EPI_ISL_430506, EPI_ISL_430507, EPI_ISL_430508, EPI_ISL_430509, EPI_ISL_430510, EPI_ISL_430511, EPI_ISL_430512, EPI_ISL_430513, EPI_ISL_430515, EPI_ISL_430516, EPI_ISL_430517, EPI_ISL_430518, EPI_ISL_430519, EPI_ISL_430520, EPI_ISL_430521, EPI_ISL_430522, EPI_ISL_430523, EPI_ISL_430524, EPI_ISL_430525, EPI_ISL_430526, EPI_ISL_430527, EPI_ISL_430528, EPI_ISL_430529, EPI_ISL_430530, EPI_ISL_430531, EPI_ISL_430533, EPI_ISL_430534, EPI_ISL_430535, EPI_ISL_430536, EPI_ISL_430537, EPI_ISL_430538, EPI_ISL_430539, EPI_ISL_430540, EPI_ISL_430542, EPI_ISL_430543, EPI_ISL_430544, EPI_ISL_430545, EPI_ISL_430546, EPI_ISL_430547, EPI_ISL_430548, EPI_ISL_430549, EPI_ISL_430550, EPI_ISL_430551, EPI_ISL_430552, EPI_ISL_430553, EPI_ISL_430554, EPI_ISL_430555, EPI_ISL_430556, EPI_ISL_430557, EPI_ISL_430558, EPI_ISL_430559, EPI_ISL_430560, EPI_ISL_430561, EPI_ISL_430562, EPI_ISL_430563, EPI_ISL_430564, EPI_ISL_430565, EPI_ISL_430567, EPI_ISL_430569, EPI_ISL_430570, EPI_ISL_430573, EPI_ISL_430574, EPI_ISL_430575, EPI_ISL_430576, EPI_ISL_430577, EPI_ISL_430578, EPI_ISL_430579, EPI_ISL_430580, EPI_ISL_430581, EPI_ISL_430582, EPI_ISL_430583, EPI_ISL_430584, EPI_ISL_430585, EPI_ISL_430586, EPI_ISL_430587, EPI_ISL_430588, EPI_ISL_430589, EPI_ISL_430590, EPI_ISL_430591, EPI_ISL_430592, EPI_ISL_430594, EPI_ISL_430595, EPI_ISL_430596, EPI_ISL_430597, EPI_ISL_430598, EPI_ISL_430599, EPI_ISL_430600, EPI_ISL_430601, EPI_ISL_430602, EPI_ISL_430603, EPI_ISL_430604, EPI_ISL_430605, EPI_ISL_430606, EPI_ISL_430607, EPI_ISL_430608, EPI_ISL_430609, EPI_ISL_430610, EPI_ISL_430611, EPI_ISL_430612, EPI_ISL_430613, EPI_ISL_430614, EPI_ISL_430615, EPI_ISL_430616, EPI_ISL_430617, EPI_ISL_430618, EPI_ISL_430619, EPI_ISL_430620, EPI_ISL_430622, EPI_ISL_430623, EPI_ISL_430624, EPI_ISL_430625, EPI_ISL_430626, EPI_ISL_430627, EPI_ISL_430628, EPI_ISL_430629, EPI_ISL_430630, EPI_ISL_430638, EPI_ISL_430688, EPI_ISL_430689, EPI_ISL_430690, EPI_ISL_430691, EPI_ISL_430692, EPI_ISL_430693, EPI_ISL_430694, EPI_ISL_430695, EPI_ISL_430696, EPI_ISL_430697, EPI_ISL_430698, EPI_ISL_430700, EPI_ISL_430701, EPI_ISL_430702, EPI_ISL_430704, EPI_ISL_430705, EPI_ISL_430706, EPI_ISL_430707, EPI_ISL_430708, EPI_ISL_430714 | | | |
| see above | Victorian Infectious Diseases Reference Laboratory (VIDRL) | Microbiological Diagnostic Unit Public Health Laboratory and Victorian Infectious Diseases Reference Laboratory, The Peter Doherty Institute for Infection and Immunity | Caly L.; Druce J.; Sait, M.; Schultz M.; Seemann T.; Sherry, N. |
| EPI_ISL_480641, EPI_ISL_480642, EPI_ISL_480643, EPI_ISL_480644, EPI_ISL_480645, EPI_ISL_480646, EPI_ISL_480650, EPI_ISL_480651, EPI_ISL_480652, EPI_ISL_480654, EPI_ISL_480655, EPI_ISL_480656, EPI_ISL_480662, EPI_ISL_480663, EPI_ISL_480668, EPI_ISL_480669, EPI_ISL_480672, EPI_ISL_480673, EPI_ISL_480674, EPI_ISL_480675, EPI_ISL_480676, EPI_ISL_480677, EPI_ISL_480678, EPI_ISL_480679, EPI_ISL_480680, EPI_ISL_480681, EPI_ISL_480682, EPI_ISL_480688, EPI_ISL_480689, EPI_ISL_480698, EPI_ISL_480699, EPI_ISL_480700, EPI_ISL_480701, EPI_ISL_480702, EPI_ISL_480704, EPI_ISL_480707, EPI_ISL_480708, EPI_ISL_480709, EPI_ISL_480710, EPI_ISL_480715, EPI_ISL_480716, EPI_ISL_480717, EPI_ISL_480718, EPI_ISL_480722, EPI_ISL_480723, EPI_ISL_480725, EPI_ISL_480726, EPI_ISL_480727, EPI_ISL_480728, EPI_ISL_480729, EPI_ISL_480730, EPI_ISL_480731, EPI_ISL_480732, EPI_ISL_480733, EPI_ISL_480735, EPI_ISL_480736, EPI_ISL_480737, EPI_ISL_480738, EPI_ISL_480740, EPI_ISL_480741, EPI_ISL_480742, EPI_ISL_480744, EPI_ISL_480765, EPI_ISL_480766, EPI_ISL_518081, EPI_ISL_518082, EPI_ISL_518083, EPI_ISL_518084, EPI_ISL_518085, EPI_ISL_518086, EPI_ISL_518088, EPI_ISL_518089, EPI_ISL_518090, EPI_ISL_518091, EPI_ISL_518092, EPI_ISL_518099, EPI_ISL_518100, EPI_ISL_518101, EPI_ISL_518103, EPI_ISL_518104, EPI_ISL_518105, EPI_ISL_518106, EPI_ISL_518109, EPI_ISL_518112, EPI_ISL_518113, EPI_ISL_518114, EPI_ISL_518117, EPI_ISL_518122, EPI_ISL_518123, EPI_ISL_518126, EPI_ISL_518127, EPI_ISL_518128, EPI_ISL_518131, EPI_ISL_518132, EPI_ISL_518133, EPI_ISL_518134, EPI_ISL_518135, EPI_ISL_518138, EPI_ISL_518139, EPI_ISL_518140, EPI_ISL_518141, EPI_ISL_518142, EPI_ISL_518147, EPI_ISL_518149, EPI_ISL_518150, EPI_ISL_518151, EPI_ISL_518153, EPI_ISL_518154, EPI_ISL_518155, EPI_ISL_518156, EPI_ISL_518157, EPI_ISL_518158, EPI_ISL_518160, EPI_ISL_518161, EPI_ISL_518162, EPI_ISL_518163, EPI_ISL_518165, EPI_ISL_518166, EPI_ISL_518167, EPI_ISL_518168, EPI_ISL_518169, EPI_ISL_518170, EPI_ISL_518171, EPI_ISL_518172, EPI_ISL_518174, EPI_ISL_518175, EPI_ISL_518179, EPI_ISL_518180, EPI_ISL_518181, EPI_ISL_518182, EPI_ISL_518185, EPI_ISL_518186, EPI_ISL_518193, EPI_ISL_518194, EPI_ISL_518195, EPI_ISL_518196, EPI_ISL_518198, EPI_ISL_518199, EPI_ISL_518200, EPI_ISL_518201, EPI_ISL_518202, EPI_ISL_518204, EPI_ISL_518206, EPI_ISL_518218, EPI_ISL_518219, EPI_ISL_518220, EPI_ISL_518224, EPI_ISL_518229, EPI_ISL_518230, EPI_ISL_518239, EPI_ISL_518240, EPI_ISL_518246, EPI_ISL_518247, EPI_ISL_518248, EPI_ISL_518249, EPI_ISL_518388, EPI_ISL_518389, EPI_ISL_518390, EPI_ISL_518391, EPI_ISL_518393, EPI_ISL_518394, EPI_ISL_518395, EPI_ISL_518396, EPI_ISL_518397, EPI_ISL_518398, EPI_ISL_518399, EPI_ISL_518400, EPI_ISL_518401, EPI_ISL_518424, EPI_ISL_518425, EPI_ISL_518426, EPI_ISL_518428, EPI_ISL_518429, EPI_ISL_518430, EPI_ISL_518433, EPI_ISL_518434, EPI_ISL_518435, EPI_ISL_518436, EPI_ISL_518437, EPI_ISL_518438, EPI_ISL_518439, EPI_ISL_518440, EPI_ISL_518442, EPI_ISL_518443, EPI_ISL_518444, EPI_ISL_518445, EPI_ISL_518446, EPI_ISL_518448, EPI_ISL_518449, EPI_ISL_518450, EPI_ISL_518451, EPI_ISL_518452, EPI_ISL_518453, EPI_ISL_518455, EPI_ISL_518456, EPI_ISL_518457, EPI_ISL_518458, EPI_ISL_518459, EPI_ISL_518460, EPI_ISL_518461, EPI_ISL_518462, EPI_ISL_518463, EPI_ISL_518465, EPI_ISL_518466, EPI_ISL_518467, EPI_ISL_518468, EPI_ISL_518469, EPI_ISL_518470, EPI_ISL_518471, EPI_ISL_518723, EPI_ISL_518981, EPI_ISL_518982, EPI_ISL_518983, EPI_ISL_518984, EPI_ISL_518985, EPI_ISL_518986, EPI_ISL_518987, EPI_ISL_518988, EPI_ISL_518989, EPI_ISL_518990, EPI_ISL_518991, EPI_ISL_518992, EPI_ISL_518994, EPI_ISL_518995, EPI_ISL_518996, EPI_ISL_518997, EPI_ISL_518998, EPI_ISL_518999, EPI_ISL_519000, EPI_ISL_519001, EPI_ISL_519002, EPI_ISL_519003, EPI_ISL_519004, EPI_ISL_519005, EPI_ISL_519006, EPI_ISL_519008, EPI_ISL_519009, EPI_ISL_519010, EPI_ISL_519011, EPI_ISL_519013, EPI_ISL_519014, EPI_ISL_519015, EPI_ISL_519016, EPI_ISL_519017, EPI_ISL_519018, EPI_ISL_519019, EPI_ISL_519020, EPI_ISL_519022, EPI_ISL_519023, EPI_ISL_519025, EPI_ISL_519026, EPI_ISL_519027, EPI_ISL_519028, EPI_ISL_519029, EPI_ISL_519030, EPI_ISL_519031, EPI_ISL_519032, EPI_ISL_519033, EPI_ISL_519034, EPI_ISL_519035, EPI_ISL_519036, EPI_ISL_519037, EPI_ISL_519038, EPI_ISL_519039, EPI_ISL_519040, EPI_ISL_519041, EPI_ISL_519042, EPI_ISL_519043, EPI_ISL_519044, EPI_ISL_519045, EPI_ISL_519046, EPI_ISL_519047, EPI_ISL_519048, EPI_ISL_519049, EPI_ISL_519050, EPI_ISL_519051, EPI_ISL_519256, EPI_ISL_519257, EPI_ISL_519258, EPI_ISL_519259, EPI_ISL_519260, EPI_ISL_519261, EPI_ISL_519262, EPI_ISL_519263, EPI_ISL_519264, EPI_ISL_519265, EPI_ISL_519266, EPI_ISL_519267, EPI_ISL_519269, EPI_ISL_519270, EPI_ISL_519272, EPI_ISL_519274, EPI_ISL_519275, EPI_ISL_519277, EPI_ISL_519278, EPI_ISL_519279, EPI_ISL_519280, EPI_ISL_519281, EPI_ISL_519282, EPI_ISL_519283, EPI_ISL_519284, EPI_ISL_519285, EPI_ISL_519286, EPI_ISL_519287, EPI_ISL_519288, EPI_ISL_519289, EPI_ISL_519290, EPI_ISL_519291, EPI_ISL_519292, EPI_ISL_519293, EPI_ISL_519294, EPI_ISL_519295, EPI_ISL_519296, EPI_ISL_519297, EPI_ISL_519298, EPI_ISL_519299, EPI_ISL_519300, EPI_ISL_519301, EPI_ISL_519305, EPI_ISL_519306, EPI_ISL_519307, EPI_ISL_519308, EPI_ISL_519309, EPI_ISL_519310, EPI_ISL_519311, EPI_ISL_519312, EPI_ISL_519313, EPI_ISL_519314, EPI_ISL_519315, EPI_ISL_519317, EPI_ISL_519318, EPI_ISL_519319, EPI_ISL_519320, EPI_ISL_519321, EPI_ISL_519322, EPI_ISL_519324, | | | |

EPI_ISL_519325, EPI_ISL_519326, EPI_ISL_519385, EPI_ISL_519948, EPI_ISL_519949, EPI_ISL_519950, EPI_ISL_520183, EPI_ISL_520184, EPI_ISL_520185, EPI_ISL_520186, EPI_ISL_520188, EPI_ISL_520192, EPI_ISL_520193, EPI_ISL_520194, EPI_ISL_520195, EPI_ISL_520196, EPI_ISL_520197, EPI_ISL_520198, EPI_ISL_520199, EPI_ISL_520200, EPI_ISL_520202, EPI_ISL_520203, EPI_ISL_520204, EPI_ISL_520205, EPI_ISL_520206, EPI_ISL_520207, EPI_ISL_520208, EPI_ISL_520209, EPI_ISL_520210, EPI_ISL_520211, EPI_ISL_520212, EPI_ISL_520213, EPI_ISL_520214, EPI_ISL_520249, EPI_ISL_520250, EPI_ISL_520251, EPI_ISL_520252, EPI_ISL_520254, EPI_ISL_520255, EPI_ISL_520256, EPI_ISL_520257, EPI_ISL_520259, EPI_ISL_520260, EPI_ISL_520261, EPI_ISL_520262, EPI_ISL_520263, EPI_ISL_520264, EPI_ISL_520265, EPI_ISL_520267, EPI_ISL_520268, EPI_ISL_520271, EPI_ISL_520272, EPI_ISL_520273, EPI_ISL_520274, EPI_ISL_520275, EPI_ISL_520277, EPI_ISL_520278, EPI_ISL_520279, EPI_ISL_520280, EPI_ISL_520281, EPI_ISL_520282, EPI_ISL_520283, EPI_ISL_520284, EPI_ISL_520287, EPI_ISL_520289, EPI_ISL_520290, EPI_ISL_520291, EPI_ISL_520292, EPI_ISL_520293, EPI_ISL_520294, EPI_ISL_520295, EPI_ISL_520296, EPI_ISL_520297, EPI_ISL_520300, EPI_ISL_520302, EPI_ISL_520303, EPI_ISL_520304, EPI_ISL_520305, EPI_ISL_520306, EPI_ISL_520308, EPI_ISL_520309, EPI_ISL_520311, EPI_ISL_520312, EPI_ISL_520313, EPI_ISL_520314, EPI_ISL_520315, EPI_ISL_520316, EPI_ISL_520317, EPI_ISL_520319, EPI_ISL_520320, EPI_ISL_520322, EPI_ISL_520323, EPI_ISL_520325, EPI_ISL_520326, EPI_ISL_520327, EPI_ISL_520328, EPI_ISL_520329, EPI_ISL_520331, EPI_ISL_520332, EPI_ISL_520333, EPI_ISL_520334, EPI_ISL_520335, EPI_ISL_520336, EPI_ISL_520338, EPI_ISL_520339, EPI_ISL_520340, EPI_ISL_520341, EPI_ISL_520342, EPI_ISL_520343, EPI_ISL_520344, EPI_ISL_520345, EPI_ISL_520346, EPI_ISL_520347, EPI_ISL_520349, EPI_ISL_520350, EPI_ISL_520351, EPI_ISL_520352, EPI_ISL_520353, EPI_ISL_520354, EPI_ISL_520356, EPI_ISL_520357, EPI_ISL_520358, EPI_ISL_520360, EPI_ISL_520361, EPI_ISL_520362, EPI_ISL_520363, EPI_ISL_520364, EPI_ISL_520365, EPI_ISL_520366, EPI_ISL_520367, EPI_ISL_520368, EPI_ISL_520369, EPI_ISL_520370, EPI_ISL_520371, EPI_ISL_520372, EPI_ISL_520374, EPI_ISL_520375, EPI_ISL_520377, EPI_ISL_520379, EPI_ISL_520381, EPI_ISL_520383, EPI_ISL_520385, EPI_ISL_520386, EPI_ISL_520387, EPI_ISL_520388, EPI_ISL_520389, EPI_ISL_520390, EPI_ISL_520745, EPI_ISL_520746, EPI_ISL_520747, EPI_ISL_520748, EPI_ISL_520749, EPI_ISL_520750, EPI_ISL_520751, EPI_ISL_520752, EPI_ISL_520753, EPI_ISL_520754, EPI_ISL_520755, EPI_ISL_520756, EPI_ISL_520757, EPI_ISL_520758, EPI_ISL_520759, EPI_ISL_520760, EPI_ISL_520761, EPI_ISL_520762, EPI_ISL_520764, EPI_ISL_520791, EPI_ISL_520792, EPI_ISL_520793, EPI_ISL_520794, EPI_ISL_520795, EPI_ISL_520796, EPI_ISL_520797, EPI_ISL_520798, EPI_ISL_520799, EPI_ISL_520800, EPI_ISL_520801, EPI_ISL_520802, EPI_ISL_520803, EPI_ISL_520804, EPI_ISL_520805, EPI_ISL_520806, EPI_ISL_520807, EPI_ISL_520808, EPI_ISL_520809, EPI_ISL_520810, EPI_ISL_520811, EPI_ISL_520812, EPI_ISL_520813, EPI_ISL_520814, EPI_ISL_520815, EPI_ISL_520816, EPI_ISL_520818, EPI_ISL_520819, EPI_ISL_520820, EPI_ISL_520821, EPI_ISL_520822, EPI_ISL_520823, EPI_ISL_520824, EPI_ISL_520825, EPI_ISL_520826, EPI_ISL_520827, EPI_ISL_520828, EPI_ISL_520829, EPI_ISL_520830, EPI_ISL_520831, EPI_ISL_520832, EPI_ISL_520833, EPI_ISL_520834, EPI_ISL_520835, EPI_ISL_520836, EPI_ISL_520837, EPI_ISL_520838, EPI_ISL_520840, EPI_ISL_520841, EPI_ISL_520843, EPI_ISL_520864, EPI_ISL_520865, EPI_ISL_520866, EPI_ISL_520867, EPI_ISL_520868, EPI_ISL_520869, EPI_ISL_520870, EPI_ISL_520871, EPI_ISL_520872, EPI_ISL_520873, EPI_ISL_520874, EPI_ISL_520875, EPI_ISL_520876, EPI_ISL_520877, EPI_ISL_520878, EPI_ISL_520879, EPI_ISL_520880, EPI_ISL_520881, EPI_ISL_520882, EPI_ISL_520887, EPI_ISL_520888, EPI_ISL_520889, EPI_ISL_520890, EPI_ISL_520891, EPI_ISL_520892, EPI_ISL_520893, EPI_ISL_520894, EPI_ISL_520895, EPI_ISL_520896, EPI_ISL_520897, EPI_ISL_520898, EPI_ISL_520899, EPI_ISL_520900, EPI_ISL_520901, EPI_ISL_520902, EPI_ISL_520903, EPI_ISL_520904, EPI_ISL_520905, EPI_ISL_520906, EPI_ISL_520907, EPI_ISL_520908, EPI_ISL_520909, EPI_ISL_520910, EPI_ISL_520911, EPI_ISL_520912, EPI_ISL_520913, EPI_ISL_520914, EPI_ISL_520915, EPI_ISL_520921, EPI_ISL_520922, EPI_ISL_520923, EPI_ISL_520924, EPI_ISL_521102, EPI_ISL_521104, EPI_ISL_521105, EPI_ISL_521106, EPI_ISL_521107, EPI_ISL_521108, EPI_ISL_521110, EPI_ISL_521111, EPI_ISL_521112, EPI_ISL_521114, EPI_ISL_521115, EPI_ISL_521116, EPI_ISL_521117, EPI_ISL_521118, EPI_ISL_521119, EPI_ISL_521121, EPI_ISL_521123, EPI_ISL_521125, EPI_ISL_521128, EPI_ISL_521131, EPI_ISL_521133, EPI_ISL_521134, EPI_ISL_521136, EPI_ISL_521137, EPI_ISL_521138, EPI_ISL_521139, EPI_ISL_521140, EPI_ISL_521141, EPI_ISL_521142, EPI_ISL_521143, EPI_ISL_521144, EPI_ISL_521145, EPI_ISL_521146, EPI_ISL_521147, EPI_ISL_521148, EPI_ISL_521150, EPI_ISL_521151, EPI_ISL_521152, EPI_ISL_521270, EPI_ISL_521271, EPI_ISL_521272, EPI_ISL_521274, EPI_ISL_521275, EPI_ISL_521276, EPI_ISL_521277, EPI_ISL_521315, EPI_ISL_521316, EPI_ISL_521317, EPI_ISL_521318, EPI_ISL_521320, EPI_ISL_521321, EPI_ISL_521323, EPI_ISL_521324, EPI_ISL_521325, EPI_ISL_521326, EPI_ISL_521327, EPI_ISL_521328, EPI_ISL_521329, EPI_ISL_521330, EPI_ISL_521332, EPI_ISL_521334, EPI_ISL_521335, EPI_ISL_521338, EPI_ISL_521339, EPI_ISL_521340, EPI_ISL_521341, EPI_ISL_521342, EPI_ISL_521343, EPI_ISL_521345, EPI_ISL_521346, EPI_ISL_521347, EPI_ISL_521348, EPI_ISL_521349, EPI_ISL_521350, EPI_ISL_521352, EPI_ISL_521353, EPI_ISL_521354, EPI_ISL_521355, EPI_ISL_521356, EPI_ISL_521357, EPI_ISL_521358, EPI_ISL_521359, EPI_ISL_521361, EPI_ISL_521363, EPI_ISL_521364, EPI_ISL_521365, EPI_ISL_521366, EPI_ISL_521367, EPI_ISL_521368, EPI_ISL_521369, EPI_ISL_521370, EPI_ISL_521371, EPI_ISL_521372, EPI_ISL_521375, EPI_ISL_521376, EPI_ISL_521377, EPI_ISL_521379, EPI_ISL_521380, EPI_ISL_521381, EPI_ISL_521382, EPI_ISL_521383, EPI_ISL_521384, EPI_ISL_521385, EPI_ISL_521386, EPI_ISL_521387, EPI_ISL_521388, EPI_ISL_521389, EPI_ISL_521393, EPI_ISL_521394, EPI_ISL_521395, EPI_ISL_521396, EPI_ISL_521397, EPI_ISL_521398, EPI_ISL_521399, EPI_ISL_521400, EPI_ISL_521402, EPI_ISL_521403, EPI_ISL_521404, EPI_ISL_521405, EPI_ISL_521406, EPI_ISL_521407, EPI_ISL_521408, EPI_ISL_521409, EPI_ISL_521410, EPI_ISL_521411, EPI_ISL_521412, EPI_ISL_521414, EPI_ISL_521415, EPI_ISL_521416, EPI_ISL_521418, EPI_ISL_521419, EPI_ISL_521420, EPI_ISL_521421, EPI_ISL_521422, EPI_ISL_521423, EPI_ISL_521424, EPI_ISL_521425, EPI_ISL_521426, EPI_ISL_521427, EPI_ISL_521428, EPI_ISL_521429, EPI_ISL_521430, EPI_ISL_521431, EPI_ISL_521432, EPI_ISL_521433, EPI_ISL_521434, EPI_ISL_521435, EPI_ISL_521436, EPI_ISL_521438, EPI_ISL_521440, EPI_ISL_521441, EPI_ISL_521442, EPI_ISL_521443, EPI_ISL_521444, EPI_ISL_521445, EPI_ISL_521447, EPI_ISL_521448, EPI_ISL_521449, EPI_ISL_521463, EPI_ISL_521464, EPI_ISL_521465, EPI_ISL_521466, EPI_ISL_521467, EPI_ISL_521468, EPI_ISL_521469, EPI_ISL_521470, EPI_ISL_521471, EPI_ISL_521472, EPI_ISL_521473, EPI_ISL_521474, EPI_ISL_521475, EPI_ISL_521476, EPI_ISL_521477, EPI_ISL_521478, EPI_ISL_521479, EPI_ISL_521480, EPI_ISL_521481, EPI_ISL_521482, EPI_ISL_521483, EPI_ISL_521484, EPI_ISL_521485, EPI_ISL_521486, EPI_ISL_521487, EPI_ISL_521488, EPI_ISL_521489, EPI_ISL_521490, EPI_ISL_521491, EPI_ISL_521492, EPI_ISL_521493, EPI_ISL_521494, EPI_ISL_521495, EPI_ISL_521496, EPI_ISL_521497, EPI_ISL_521498, EPI_ISL_521499, EPI_ISL_521500, EPI_ISL_521502, EPI_ISL_521503, EPI_ISL_521504, EPI_ISL_521505, EPI_ISL_521506, EPI_ISL_521507, EPI_ISL_521508, EPI_ISL_521509, EPI_ISL_521510, EPI_ISL_521511, EPI_ISL_521512, EPI_ISL_521514, EPI_ISL_521515, EPI_ISL_521517, EPI_ISL_521518, EPI_ISL_521519, EPI_ISL_521520, EPI_ISL_521521, EPI_ISL_521522, EPI_ISL_521523, EPI_ISL_521524, EPI_ISL_521525, EPI_ISL_521527, EPI_ISL_521528, EPI_ISL_521533, EPI_ISL_521534, EPI_ISL_521535, EPI_ISL_521536, EPI_ISL_521537, EPI_ISL_521538, EPI_ISL_521539, EPI_ISL_521540, EPI_ISL_521541, EPI_ISL_521542, EPI_ISL_521543, EPI_ISL_521545, EPI_ISL_521547, EPI_ISL_521548, EPI_ISL_521549, EPI_ISL_521550, EPI_ISL_521551, EPI_ISL_521552, EPI_ISL_521553, EPI_ISL_521554, EPI_ISL_521555, EPI_ISL_521556, EPI_ISL_521557, EPI_ISL_521558, EPI_ISL_521559, EPI_ISL_521560, EPI_ISL_521561, EPI_ISL_521841, EPI_ISL_521842, EPI_ISL_521843, EPI_ISL_521844, EPI_ISL_521845, EPI_ISL_521846, EPI_ISL_521847, EPI_ISL_521849, EPI_ISL_521850, EPI_ISL_521851, EPI_ISL_521852, EPI_ISL_521853, EPI_ISL_521854, EPI_ISL_521855, EPI_ISL_521856, EPI_ISL_521857, EPI_ISL_521858, EPI_ISL_521859, EPI_ISL_521870, EPI_ISL_521871, EPI_ISL_521895, EPI_ISL_521905, EPI_ISL_521913, EPI_ISL_521914, EPI_ISL_521915, EPI_ISL_521918, EPI_ISL_521919, EPI_ISL_521922, EPI_ISL_521924, EPI_ISL_521925, EPI_ISL_521945, EPI_ISL_521946, EPI_ISL_521947, EPI_ISL_521948, EPI_ISL_521949, EPI_ISL_521950, EPI_ISL_521951, EPI_ISL_521952, EPI_ISL_521953, EPI_ISL_521954, EPI_ISL_521956, EPI_ISL_521957, EPI_ISL_521959, EPI_ISL_521961, EPI_ISL_521963, EPI_ISL_521964, EPI_ISL_521966, EPI_ISL_521967, EPI_ISL_521968, EPI_ISL_521969, EPI_ISL_521975, EPI_ISL_521976, EPI_ISL_521977, EPI_ISL_521978, EPI_ISL_521981, EPI_ISL_521982, EPI_ISL_521984, EPI_ISL_521985, EPI_ISL_521986, EPI_ISL_521987, EPI_ISL_522055, EPI_ISL_522056, EPI_ISL_522057, EPI_ISL_522058, EPI_ISL_522059, EPI_ISL_522060, EPI_ISL_522061, EPI_ISL_522062, EPI_ISL_522063, EPI_ISL_522064, EPI_ISL_522065, EPI_ISL_522066, EPI_ISL_522068, EPI_ISL_522069, EPI_ISL_522070, EPI_ISL_522071, EPI_ISL_522073, EPI_ISL_522074, EPI_ISL_522075, EPI_ISL_522076, EPI_ISL_522077, EPI_ISL_522078, EPI_ISL_522079, EPI_ISL_522080, EPI_ISL_522081, EPI_ISL_522082, EPI_ISL_522083, EPI_ISL_522084, EPI_ISL_522085, EPI_ISL_522086, EPI_ISL_522087, EPI_ISL_522088, EPI_ISL_522089, EPI_ISL_522090, EPI_ISL_522091, EPI_ISL_522093, EPI_ISL_522094, EPI_ISL_522096, EPI_ISL_522097, EPI_ISL_522098, EPI_ISL_522099, EPI_ISL_522100, EPI_ISL_522102, EPI_ISL_522103, EPI_ISL_522104, EPI_ISL_522105, EPI_ISL_522106, EPI_ISL_522107, EPI_ISL_522108, EPI_ISL_522109, EPI_ISL_522120, EPI_ISL_522121, EPI_ISL_522122, EPI_ISL_522123, EPI_ISL_522124, EPI_ISL_522125, EPI_ISL_522126, EPI_ISL_522127, EPI_ISL_522128, EPI_ISL_522129, EPI_ISL_522130, EPI_ISL_522132, EPI_ISL_522133, EPI_ISL_522134, EPI_ISL_522135, EPI_ISL_522136, EPI_ISL_522137, EPI_ISL_522138, EPI_ISL_522139, EPI_ISL_522140, EPI_ISL_522141, EPI_ISL_522142, EPI_ISL_522143, EPI_ISL_522145, EPI_ISL_522146, EPI_ISL_522147, EPI_ISL_522148, EPI_ISL_522149, EPI_ISL_522150, EPI_ISL_522151, EPI_ISL_522152, EPI_ISL_522153, EPI_ISL_522154, EPI_ISL_522155, EPI_ISL_522157, EPI_ISL_522158, EPI_ISL_522159, EPI_ISL_522160, EPI_ISL_522161, EPI_ISL_522162, EPI_ISL_522163, EPI_ISL_522164, EPI_ISL_522165, EPI_ISL_522166, EPI_ISL_522168, EPI_ISL_522169, EPI_ISL_522170, EPI_ISL_522171, EPI_ISL_522172, EPI_ISL_522173, EPI_ISL_522174, EPI_ISL_522175, EPI_ISL_522176, EPI_ISL_522177, EPI_ISL_522178, EPI_ISL_522179, EPI_ISL_522180, EPI_ISL_522181, EPI_ISL_522182, EPI_ISL_522183, EPI_ISL_522184, EPI_ISL_522185, EPI_ISL_522186, EPI_ISL_522187, EPI_ISL_522233, EPI_ISL_522234, EPI_ISL_522235, EPI_ISL_522236, EPI_ISL_522237, EPI_ISL_522238, EPI_ISL_522239, EPI_ISL_522240, EPI_ISL_522241, EPI_ISL_561403, EPI_ISL_561409, EPI_ISL_561414, EPI_ISL_561420, EPI_ISL_561425, EPI_ISL_561426, EPI_ISL_561428, EPI_ISL_561440, EPI_ISL_561453, EPI_ISL_561458, EPI_ISL_561462, EPI_ISL_561465, EPI_ISL_561468, EPI_ISL_561470, EPI_ISL_561491, EPI_ISL_561498, EPI_ISL_561508, EPI_ISL_561514, EPI_ISL_561517, EPI_ISL_561523, EPI_ISL_561533, EPI_ISL_561541, EPI_ISL_561544, EPI_ISL_561559, EPI_ISL_561565, EPI_ISL_561582, EPI_ISL_561584, EPI_ISL_561586, EPI_ISL_561590, EPI_ISL_561591, EPI_ISL_561612, EPI_ISL_561621, EPI_ISL_561629, EPI_ISL_561651, EPI_ISL_561653, EPI_ISL_561655, EPI_ISL_561656, EPI_ISL_561657, EPI_ISL_561661, EPI_ISL_561673, EPI_ISL_561682, EPI_ISL_561686, EPI_ISL_561690, EPI_ISL_561697, EPI_ISL_561707, EPI_ISL_561711, EPI_ISL_561715, EPI_ISL_561728, EPI_ISL_561741, EPI_ISL_561750, EPI_ISL_561752, EPI_ISL_561765, EPI_ISL_561778, EPI_ISL_561787, EPI_ISL_561788, EPI_ISL_561791, EPI_ISL_561803, EPI_ISL_561816, EPI_ISL_561825, EPI_ISL_561840, EPI_ISL_561852, EPI_ISL_561858, EPI_ISL_561863, EPI_ISL_561893, EPI_ISL_561894, EPI_ISL_561935, EPI_ISL_561936, EPI_ISL_561946, EPI_ISL_561957, EPI_ISL_561975, EPI_ISL_561986, EPI_ISL_561987, EPI_ISL_561988, EPI_ISL_561991, EPI_ISL_561994, EPI_ISL_562011, EPI_ISL_562012, EPI_ISL_562016, EPI_ISL_562023, EPI_ISL_562033, EPI_ISL_562048, EPI_ISL_562055, EPI_ISL_562057, EPI_ISL_562067, EPI_ISL_562069, EPI_ISL_562076, EPI_ISL_562080, EPI_ISL_562081, EPI_ISL_562085, EPI_ISL_562106, EPI_ISL_562109, EPI_ISL_562110, EPI_ISL_562136, EPI_ISL_562138, EPI_ISL_562152, EPI_ISL_562155, EPI_ISL_562157, EPI_ISL_562163, EPI_ISL_562164, EPI_ISL_562216, EPI_ISL_562348, EPI_ISL_562482, EPI_ISL_562518, EPI_ISL_562539, EPI_ISL_562540, EPI_ISL_562593, EPI_ISL_562595, EPI_ISL_562628, EPI_ISL_562640, EPI_ISL_562645, EPI_ISL_562646, EPI_ISL_562658, EPI_ISL_562659, EPI_ISL_562696, EPI_ISL_562697, EPI_ISL_562700, EPI_ISL_562754, EPI_ISL_562808, EPI_ISL_562838, EPI_ISL_562839, EPI_ISL_562842, EPI_ISL_562843, EPI_ISL_562844, EPI_ISL_562910, EPI_ISL_562911, EPI_ISL_562912, EPI_ISL_562913, EPI_ISL_562924, EPI_ISL_562929, EPI_ISL_562936, EPI_ISL_562946, EPI_ISL_562951, EPI_ISL_562961, EPI_ISL_563000, EPI_ISL_563131, EPI_ISL_563132, EPI_ISL_563133, EPI_ISL_563137, EPI_ISL_563138, EPI_ISL_563139, EPI_ISL_563140, EPI_ISL_563141, EPI_ISL_563142, EPI_ISL_563144, EPI_ISL_563409, EPI_ISL_563410, EPI_ISL_563411, EPI_ISL_563412, EPI_ISL_563413, EPI_ISL_563414, EPI_ISL_563415, EPI_ISL_563416, EPI_ISL_563417, EPI_ISL_563418, EPI_ISL_563419, EPI_ISL_563420, EPI_ISL_563421, EPI_ISL_563422, EPI_ISL_563423, EPI_ISL_563497, EPI_ISL_563498, EPI_ISL_563499, EPI_ISL_563551, EPI_ISL_563721, EPI_ISL_563723, EPI_ISL_563790, EPI_ISL_563792, EPI_ISL_563808, EPI_ISL_563811, EPI_ISL_563866, EPI_ISL_563925, EPI_ISL_563973, EPI_ISL_563974, EPI_ISL_563975, EPI_ISL_563998, EPI_ISL_563999, EPI_ISL_564000, EPI_ISL_564001, EPI_ISL_564003, EPI_ISL_564004, EPI_ISL_564005, EPI_ISL_564007, EPI_ISL_564008, EPI_ISL_564009, EPI_ISL_564010, EPI_ISL_564011, EPI_ISL_564012, EPI_ISL_564013, EPI_ISL_564014, EPI_ISL_564015, EPI_ISL_564016, EPI_ISL_564017, EPI_ISL_564018, EPI_ISL_564019, EPI_ISL_564020, EPI_ISL_564021, EPI_ISL_564022, EPI_ISL_564023, EPI_ISL_564024, EPI_ISL_564025, EPI_ISL_564026, EPI_ISL_564027, EPI_ISL_564028, EPI_ISL_564029, EPI_ISL_564030, EPI_ISL_564031, EPI_ISL_564032, EPI_ISL_564033, EPI_ISL_564034, EPI_ISL_564035, EPI_ISL_564036, EPI_ISL_564037, EPI_ISL_564038, EPI_ISL_564039, EPI_ISL_564040, EPI_ISL_564041, EPI_ISL_564042, EPI_ISL_564043, EPI_ISL_564044, EPI_ISL_564045, EPI_ISL_564046, EPI_ISL_564048, EPI_ISL_564049, EPI_ISL_564055, EPI_ISL_564057, EPI_ISL_564058, EPI_ISL_564059, EPI_ISL_564060, EPI_ISL_564061, EPI_ISL_564062, EPI_ISL_564063, EPI_ISL_564064, EPI_ISL_564065, EPI_ISL_564066, EPI_ISL_564067, EPI_ISL_564068, EPI_ISL_564069, EPI_ISL_564070, EPI_ISL_564071, EPI_ISL_564074, EPI_ISL_564075, EPI_ISL_564076, EPI_ISL_564077, EPI_ISL_564078, EPI_ISL_564079, EPI_ISL_564080, EPI_ISL_564081, EPI_ISL_564082, EPI_ISL_564083, EPI_ISL_564084, EPI_ISL_564085, EPI_ISL_564086, EPI_ISL_564088, EPI_ISL_564089, EPI_ISL_564090, EPI_ISL_564091, EPI_ISL_564092, EPI_ISL_564093, EPI_ISL_564094, EPI_ISL_564095, EPI_ISL_564096, EPI_ISL_564097, EPI_ISL_564098, EPI_ISL_564099, EPI_ISL_564100, EPI_ISL_564101, EPI_ISL_564103, EPI_ISL_564104, EPI_ISL_564105, EPI_ISL_564106, EPI_ISL_564107, EPI_ISL_564108, EPI_ISL_564109, EPI_ISL_564110, EPI_ISL_564180, EPI_ISL_564181, EPI_ISL_564182, EPI_ISL_564184, EPI_ISL_564185, EPI_ISL_564186, EPI_ISL_564187, EPI_ISL_564188, EPI_ISL_564189, EPI_ISL_564190, EPI_ISL_564191, EPI_ISL_564192, EPI_ISL_564193, EPI_ISL_564194, EPI_ISL_564195, EPI_ISL_564196, EPI_ISL_564197, EPI_ISL_564198, EPI_ISL_564199, EPI_ISL_564200, EPI_ISL_564203, EPI_ISL_564204, EPI_ISL_564205, EPI_ISL_564206, EPI_ISL_564207, EPI_ISL_564208, EPI_ISL_564209, EPI_ISL_564210, EPI_ISL_564212, EPI_ISL_564213, EPI_ISL_564214, EPI_ISL_564215, EPI_ISL_564216, EPI_ISL_564217, EPI_ISL_564218, EPI_ISL_564219, EPI_ISL_564220, EPI_ISL_564221, EPI_ISL_564222, EPI_ISL_564223, EPI_ISL_564224, EPI_ISL_564225, EPI_ISL_564226, EPI_ISL_564227, EPI_ISL_564228, EPI_ISL_564229, EPI_ISL_564230, EPI_ISL_564231, EPI_ISL_564232, EPI_ISL_564233, EPI_ISL_564234, EPI_ISL_564235, EPI_ISL_564236, EPI_ISL_564237, EPI_ISL_564238, EPI_ISL_564239, EPI_ISL_564240, EPI_ISL_564241, EPI_ISL_564242, EPI_ISL_564243, EPI_ISL_564244, EPI_ISL_564245, EPI_ISL_564246, EPI_ISL_564247, EPI_ISL_564248, EPI_ISL_564249, EPI_ISL_564250, EPI_ISL_564251, EPI_ISL_564254, EPI_ISL_564270, EPI_ISL_564319, EPI_ISL_564320, EPI_ISL_564321, EPI_ISL_564322, EPI_ISL_564323, EPI_ISL_564324, EPI_ISL_564325, EPI_ISL_564327, EPI_ISL_564328, EPI_ISL_564329, EPI_ISL_564330, EPI_ISL_564331, EPI_ISL_564332, EPI_ISL_564333, EPI_ISL_564334, EPI_ISL_564335, EPI_ISL_564336, EPI_ISL_564337, EPI_ISL_564338, EPI_ISL_564339, EPI_ISL_564340, EPI_ISL_564341, EPI_ISL_564342, EPI_ISL_564343, EPI_ISL_564344, EPI_ISL_564345, EPI_ISL_564346, EPI_ISL_564348, EPI_ISL_564349, EPI_ISL_564350, EPI_ISL_564353, EPI_ISL_564354, EPI_ISL_564355, EPI_ISL_564356, EPI_ISL_564358, EPI_ISL_564359, EPI_ISL_564360, EPI_ISL_564361, EPI_ISL_564362, EPI_ISL_564363, EPI_ISL_564364, EPI_ISL_564365, EPI_ISL_564366, EPI_ISL_564367, EPI_ISL_564368, EPI_ISL_564369, EPI_ISL_564370, EPI_ISL_564371, EPI_ISL_564372, EPI_ISL_564373, EPI_ISL_564374, EPI_ISL_564375, EPI_ISL_564376, EPI_ISL_564377, EPI_ISL_564378, EPI_ISL_564379, EPI_ISL_564380, EPI_ISL_564381, EPI_ISL_564382, EPI_ISL_564383, EPI_ISL_564384, EPI_ISL_564385, EPI_ISL_564386, EPI_ISL_564387, EPI_ISL_564388, EPI_ISL_564389, EPI_ISL_564390, EPI_ISL_564391, EPI_ISL_564392, EPI_ISL_564393, EPI_ISL_564395, EPI_ISL_564396, EPI_ISL_564397, EPI_ISL_564404, EPI_ISL_564544, EPI_ISL_564551, EPI_ISL_564556, EPI_ISL_564557, EPI_ISL_564558, EPI_ISL_564559, EPI_ISL_564560, EPI_ISL_564561, EPI_ISL_564563, EPI_ISL_565137, EPI_ISL_565138, EPI_ISL_565139, EPI_ISL_565141, EPI_ISL_565144, EPI_ISL_565145, EPI_ISL_565146, EPI_ISL_565147, EPI_ISL_565148, EPI_ISL_565149, EPI_ISL_565150, EPI_ISL_565151, EPI_ISL_565152, EPI_ISL_565153, EPI_ISL_565154, EPI_ISL_565156, EPI_ISL_565157, EPI_ISL_565158, EPI_ISL_565159, EPI_ISL_565160, EPI_ISL_565161, EPI_ISL_565162, EPI_ISL_565163, EPI_ISL_565164, EPI_ISL_565165, EPI_ISL_565166, EPI_ISL_565167, EPI_ISL_565168, EPI_ISL_565169, EPI_ISL_565170, EPI_ISL_565171, EPI_ISL_565172, EPI_ISL_565173, EPI_ISL_565174, EPI_ISL_565175, EPI_ISL_565176, EPI_ISL_565177, EPI_ISL_565178, EPI_ISL_565179, EPI_ISL_565180, EPI_ISL_565181, EPI_ISL_565182, EPI_ISL_565183, EPI_ISL_565184, EPI_ISL_565185, EPI_ISL_565186, EPI_ISL_565187, EPI_ISL_565188, EPI_ISL_565189, EPI_ISL_565190, EPI_ISL_565191, EPI_ISL_565192, EPI_ISL_565193, EPI_ISL_565194, EPI_ISL_565195, EPI_ISL_565196, EPI_ISL_565197, EPI_ISL_565198, EPI_ISL_565199, EPI_ISL_565200, EPI_ISL_565201, EPI_ISL_565202, EPI_ISL_565203, EPI_ISL_565204, EPI_ISL_565205, EPI_ISL_565206, EPI_ISL_565207, EPI_ISL_565208, EPI_ISL_565209, EPI_ISL_565210, EPI_ISL_565211, EPI_ISL_565212, EPI_ISL_565213, EPI_ISL_565214, EPI_ISL_565215, EPI_ISL_565216, EPI_ISL_565217, EPI_ISL_565218, EPI_ISL_565219, EPI_ISL_565220, EPI_ISL_565221, EPI_ISL_565222, EPI_ISL_565382, EPI_ISL_565385, EPI_ISL_565394, EPI_ISL_565395, EPI_ISL_565403, EPI_ISL_565404, EPI_ISL_565409, EPI_ISL_565415, EPI_ISL_565459, EPI_ISL_565461, EPI_ISL_565464, EPI_ISL_565465, EPI_ISL_565466, EPI_ISL_565467, EPI_ISL_565469, EPI_ISL_565733, EPI_ISL_565734, EPI_ISL_565736, EPI_ISL_565737, EPI_ISL_565738, EPI_ISL_565739, EPI_ISL_565740, EPI_ISL_565741, EPI_ISL_565742, EPI_ISL_591558, EPI_ISL_591562, EPI_ISL_591571, EPI_ISL_591573, EPI_ISL_591580, EPI_ISL_591585, EPI_ISL_591587, EPI_ISL_591592, EPI_ISL_591619, EPI_ISL_591621, EPI_ISL_591623, EPI_ISL_591638, EPI_ISL_591643, EPI_ISL_591645, EPI_ISL_591654, EPI_ISL_591656, EPI_ISL_591657, EPI_ISL_591666, EPI_ISL_591669, EPI_ISL_591684, EPI_ISL_591688, EPI_ISL_591706, EPI_ISL_591707, EPI_ISL_591708, EPI_ISL_591792, EPI_ISL_591807, EPI_ISL_591824, EPI_ISL_591825, EPI_ISL_591849, EPI_ISL_591850, EPI_ISL_591852, EPI_ISL_591854, EPI_ISL_591856, EPI_ISL_591858, EPI_ISL_591860, EPI_ISL_591861, EPI_ISL_591862, EPI_ISL_591863, EPI_ISL_591864, EPI_ISL_591866, EPI_ISL_591870, EPI_ISL_591871, EPI_ISL_591872, EPI_ISL_591873, EPI_ISL_591878, EPI_ISL_591879, EPI_ISL_591881, EPI_ISL_591883, EPI_ISL_591884, EPI_ISL_591887, EPI_ISL_591889, EPI_ISL_591890, EPI_ISL_591891, EPI_ISL_591894, EPI_ISL_591895, EPI_ISL_591896, EPI_ISL_591897, EPI_ISL_591899, EPI_ISL_591900, EPI_ISL_591901, EPI_ISL_591904, EPI_ISL_591907, EPI_ISL_591908, EPI_ISL_591909, EPI_ISL_591912, EPI_ISL_591917, EPI_ISL_591919, EPI_ISL_591925, EPI_ISL_591928, EPI_ISL_591932, EPI_ISL_591940, EPI_ISL_591943, EPI_ISL_591947, EPI_ISL_591948, EPI_ISL_591952, EPI_ISL_591954, EPI_ISL_591955, EPI_ISL_591956, EPI_ISL_592137, EPI_ISL_592138, EPI_ISL_592139, EPI_ISL_592142, EPI_ISL_592143, EPI_ISL_592144, EPI_ISL_592145, EPI_ISL_592146, EPI_ISL_592147, EPI_ISL_592148, EPI_ISL_592149, EPI_ISL_592150, EPI_ISL_592151, EPI_ISL_592152, EPI_ISL_592154, EPI_ISL_592156, EPI_ISL_592157, EPI_ISL_592159, EPI_ISL_592160, EPI_ISL_592161, EPI_ISL_592162, EPI_ISL_592163, EPI_ISL_592164, EPI_ISL_592165, EPI_ISL_592166, EPI_ISL_592168, EPI_ISL_592169, EPI_ISL_592170, EPI_ISL_592171, EPI_ISL_592175, EPI_ISL_592180, EPI_ISL_592184, EPI_ISL_592188, EPI_ISL_592189, EPI_ISL_592191, EPI_ISL_592194, EPI_ISL_592195, EPI_ISL_592486, EPI_ISL_592488, EPI_ISL_592489, EPI_ISL_592491, EPI_ISL_592493, EPI_ISL_592494, EPI_ISL_592495, EPI_ISL_592496, EPI_ISL_592497, EPI_ISL_592498, EPI_ISL_592499, EPI_ISL_592500, EPI_ISL_592503, EPI_ISL_592504, EPI_ISL_592505, EPI_ISL_592506, EPI_ISL_592511, EPI_ISL_592514, EPI_ISL_592516, EPI_ISL_592568, EPI_ISL_592577, EPI_ISL_592590, EPI_ISL_592597, EPI_ISL_592600, EPI_ISL_592616, EPI_ISL_592660, EPI_ISL_592661, EPI_ISL_592668, EPI_ISL_592820, EPI_ISL_592821, EPI_ISL_592824, EPI_ISL_592836, EPI_ISL_592837, EPI_ISL_592858, EPI_ISL_592859, EPI_ISL_592860, EPI_ISL_592862, EPI_ISL_592863, EPI_ISL_592864, EPI_ISL_592865, EPI_ISL_592878, EPI_ISL_592879, EPI_ISL_592969, EPI_ISL_592973, EPI_ISL_592974, EPI_ISL_592976, EPI_ISL_592977, EPI_ISL_592978, EPI_ISL_592979, EPI_ISL_592980, EPI_ISL_592981, EPI_ISL_592982, EPI_ISL_592983, EPI_ISL_592984, EPI_ISL_592985, EPI_ISL_593003, EPI_ISL_593005, EPI_ISL_593006, EPI_ISL_593007, EPI_ISL_593033, EPI_ISL_593035, EPI_ISL_593048, EPI_ISL_593126, EPI_ISL_593127, EPI_ISL_593128, EPI_ISL_593215, EPI_ISL_593216, EPI_ISL_593256, EPI_ISL_593476, EPI_ISL_593477, EPI_ISL_640519, EPI_ISL_640583, EPI_ISL_640588, EPI_ISL_640763, EPI_ISL_640784, EPI_ISL_641121, EPI_ISL_1055278, EPI_ISL_1055301, EPI_ISL_1055340, EPI_ISL_1055341, EPI_ISL_1055348, EPI_ISL_1055358

| | | | |
|---|---|---|---|
| see above | Victorian Infectious Diseases Reference Laboratory (VIDRL) | VIDRL and MDU-PHL | Caly L.; Caly, L.; Druce J.; M. B.; M.L.; N.L.; Sait; Sait, M.; Schultz; Schultz M.; Schultz M.B.; Seemann T.; Seemann, T.; Sherry; Sherry, N. |

EPI_ISL_416410, EPI_ISL_416411, EPI_ISL_416412, EPI_ISL_416413, EPI_ISL_416414, EPI_ISL_416415, EPI_ISL_416514, EPI_ISL_416515, EPI_ISL_416516, EPI_ISL_416517, EPI_ISL_416518, EPI_ISL_419733, EPI_ISL_419734, EPI_ISL_419736, EPI_ISL_419737, EPI_ISL_419738, EPI_ISL_419739, EPI_ISL_419740, EPI_ISL_419741, EPI_ISL_419742, EPI_ISL_419743, EPI_ISL_419744, EPI_ISL_419745, EPI_ISL_419746, EPI_ISL_419747, EPI_ISL_419748, EPI_ISL_419749, EPI_ISL_419750, EPI_ISL_419751, EPI_ISL_419752, EPI_ISL_419753, EPI_ISL_419754, EPI_ISL_419755, EPI_ISL_419756, EPI_ISL_419757, EPI_ISL_419758, EPI_ISL_419759, EPI_ISL_419760, EPI_ISL_419761, EPI_ISL_419762, EPI_ISL_419763, EPI_ISL_419764, EPI_ISL_419765, EPI_ISL_419766, EPI_ISL_419767, EPI_ISL_419768, EPI_ISL_419769, EPI_ISL_419770, EPI_ISL_419771, EPI_ISL_419772, EPI_ISL_419773, EPI_ISL_419774, EPI_ISL_419775, EPI_ISL_419776, EPI_ISL_419777, EPI_ISL_419778, EPI_ISL_419779, EPI_ISL_419780, EPI_ISL_419781, EPI_ISL_419782, EPI_ISL_419783, EPI_ISL_419784, EPI_ISL_419785, EPI_ISL_419786, EPI_ISL_419787, EPI_ISL_419788, EPI_ISL_419789, EPI_ISL_419791, EPI_ISL_419792, EPI_ISL_419793, EPI_ISL_419794, EPI_ISL_419795, EPI_ISL_419796, EPI_ISL_419797, EPI_ISL_419798, EPI_ISL_419799, EPI_ISL_419800, EPI_ISL_419801, EPI_ISL_419802, EPI_ISL_419803, EPI_ISL_419804, EPI_ISL_419805, EPI_ISL_419806, EPI_ISL_419807, EPI_ISL_419808, EPI_ISL_419809, EPI_ISL_419810, EPI_ISL_419811, EPI_ISL_419812, EPI_ISL_419813, EPI_ISL_419814, EPI_ISL_419815, EPI_ISL_419816, EPI_ISL_419817, EPI_ISL_419818, EPI_ISL_419819, EPI_ISL_419821, EPI_ISL_419822, EPI_ISL_419826, EPI_ISL_419834, EPI_ISL_419836, EPI_ISL_419837, EPI_ISL_419838, EPI_ISL_419839, EPI_ISL_419840, EPI_ISL_419841, EPI_ISL_419842, EPI_ISL_419843, EPI_ISL_419844, EPI_ISL_419845, EPI_ISL_419846, EPI_ISL_419847, EPI_ISL_419848, EPI_ISL_419849, EPI_ISL_419850, EPI_ISL_419851, EPI_ISL_419852, EPI_ISL_419853, EPI_ISL_419854, EPI_ISL_419855, EPI_ISL_419856, EPI_ISL_419857, EPI_ISL_419859, EPI_ISL_419860, EPI_ISL_419861, EPI_ISL_419862, EPI_ISL_419863, EPI_ISL_419864, EPI_ISL_419865, EPI_ISL_419866, EPI_ISL_419867, EPI_ISL_419868, EPI_ISL_419869, EPI_ISL_419870, EPI_ISL_419871, EPI_ISL_419872, EPI_ISL_419873, EPI_ISL_419874, EPI_ISL_419875, EPI_ISL_419876, EPI_ISL_419878, EPI_ISL_419879, EPI_ISL_419881, EPI_ISL_419882, EPI_ISL_419883, EPI_ISL_419884, EPI_ISL_419885, EPI_ISL_419886, EPI_ISL_419887, EPI_ISL_419888, EPI_ISL_419889, EPI_ISL_419890, EPI_ISL_419891, EPI_ISL_419892, EPI_ISL_419893, EPI_ISL_419894, EPI_ISL_419895, EPI_ISL_419896, EPI_ISL_419897, EPI_ISL_419898, EPI_ISL_419899, EPI_ISL_419900, EPI_ISL_419901, EPI_ISL_419902, EPI_ISL_419903, EPI_ISL_419904, EPI_ISL_419905, EPI_ISL_419906, EPI_ISL_419907, EPI_ISL_419908, EPI_ISL_419909, EPI_ISL_419910, EPI_ISL_419911, EPI_ISL_419912, EPI_ISL_419913, EPI_ISL_419914, EPI_ISL_419915, EPI_ISL_419917, EPI_ISL_419918, EPI_ISL_419919, EPI_ISL_419920, EPI_ISL_419921, EPI_ISL_419922, EPI_ISL_419923, EPI_ISL_419924, EPI_ISL_419926, EPI_ISL_419927, EPI_ISL_419928, EPI_ISL_419929, EPI_ISL_419931, EPI_ISL_419932, EPI_ISL_419933, EPI_ISL_419934, EPI_ISL_419935, EPI_ISL_419936, EPI_ISL_419938, EPI_ISL_419939, EPI_ISL_419940, EPI_ISL_419941, EPI_ISL_419942, EPI_ISL_419943, EPI_ISL_419944, EPI_ISL_419945, EPI_ISL_419946, EPI_ISL_419947, EPI_ISL_419948, EPI_ISL_419949, EPI_ISL_419950, EPI_ISL_419951, EPI_ISL_419952, EPI_ISL_419953, EPI_ISL_419954, EPI_ISL_419955, EPI_ISL_419956, EPI_ISL_419957, EPI_ISL_419958, EPI_ISL_419959, EPI_ISL_419960, EPI_ISL_419961, EPI_ISL_419962, EPI_ISL_419963, EPI_ISL_419964, EPI_ISL_419965, EPI_ISL_419966, EPI_ISL_419967, EPI_ISL_419968, EPI_ISL_419969, EPI_ISL_419970, EPI_ISL_419971, EPI_ISL_419972, EPI_ISL_419973, EPI_ISL_419974, EPI_ISL_419975, EPI_ISL_419976, EPI_ISL_419977, EPI_ISL_419978, EPI_ISL_419979, EPI_ISL_419983, EPI_ISL_419984, EPI_ISL_419985, EPI_ISL_419986, EPI_ISL_419987, EPI_ISL_419988, EPI_ISL_419989, EPI_ISL_419990, EPI_ISL_419991, EPI_ISL_419992, EPI_ISL_419993, EPI_ISL_419994, EPI_ISL_419995, EPI_ISL_419996, EPI_ISL_419997, EPI_ISL_419998, EPI_ISL_420004, EPI_ISL_420005, EPI_ISL_420036

| | | | |
|---|---|---|---|
| see above | Victorian Infectious Diseases Reference Laboratory (VIDRL) | Victorian Infectious Diseases Reference Laboratory and Microbiological Diagnostic Unit Public Health Laboratory, Doherty Institute | Caly L.; Druce J.; Sait, M.; Schultz M.; Seemann T.; Sherry, N.; Taiaroa, G. |
| EPI_ISL_487271 | unknown | MDU-PHL, The Peter Doherty Institute for Infection and Immunity | Antonia da Costa, E.; Baird, R.; Baretto, I.; Bastian, I.; C.K.; Caly, L.; Canisia, D.; Dakh, F.; Dolores de Jesus da Costa, M.; Douglas, N.; Francis, J.; Freeman, K.; Higgins, G.; Jayanti Pereira Tilman, A.; Leong, L.; Lim; M.B.; Marr, I.; Meumann, E.; Sait, M.; Salles de Sousa, A.; Schultz; Seemann, T.; Sherry, N.; Soares da Silva, E.; Turra, M.; Wapling, J.; Ximenes, J. |
| EPI_ISL_450212, EPI_ISL_450213, EPI_ISL_450214, EPI_ISL_450215, EPI_ISL_450216 | unknown | Microbiological Diagnostic Unit Public Health Laboratory (MDU-PHL) and Victorian Infectious Disease Reference Laboratory (VIDRL) | Alpren, C.; B.P.; Ballard; C.R.; Caly, L.; Catton, M.; D.A.; Dougal, S.; Druce, J.; Duchene, S.; Easton, M.; Goncalves da Silva, A.; Hoang, T.; Horan, K.; Howden; Lane; M.B.; N.L.; S.A.; Sait, M.; Schultz; Seemann, T.; Sherry; Stinear; Sutton, B.; T.P.; Williamson; van Diemen, A. |
